# International Expert-Based Consensus Definition, Staging Criteria, and Minimum Data Elements for Osteoradionecrosis of the Jaw: An Inter-Disciplinary Modified Delphi Study

**DOI:** 10.1101/2024.04.07.24305400

**Authors:** The International ORAL Consortium, Amy C. Moreno, Erin E. Watson, Laia Humbert-Vidan, Douglas E. Peterson, Lisanne V van Dijk, Teresa Guerrero Urbano, Lisa Van den Bosch, Andrew J. Hope, Matthew S. Katz, Frank J.P. Hoebers, Ruth A. Aponte Wesson, James E. Bates, Paolo Bossi, Adeyinka F. Dayo, Mélanie Doré, Eduardo Rodrigues Fregnani, Thomas J. Galloway, Daphna Y. Gelblum, Issa A. Hanna, Christina E. Henson, Sudarat Kiat-amnuay, Anke Korfage, Nancy Y. Lee, Carol M. Lewis, Charlotte Duch Lynggaard, Antti A. Mäkitie, Marco Magalhaes, Yvonne M. Mowery, Carles Muñoz-Montplet, Jeffrey N. Myers, Ester Orlandi, Jaymit Patel, Jillian M. Rigert, Deborah Saunders, Jonathan D. Schoenfeld, Ugur Selek, Efsun Somay, Vinita Takiar, Juliette Thariat, Gerda M. Verduijn, Alessandro Villa, Nick West, Max J.H. Witjes, Alex Won, Mark E. Wong, Christopher M.K.L. Yao, Simon W. Young, Kamal Al-eryani, Carly E.A. Barbon, Doke J.M. Buurman, François J. Dieleman, Theresa M. Hofstede, Abdul Ahad Khan, Adegbenga O. Otun, John C. Robinson, Lauren Hum, Jorgen Johansen, Rajesh Lalla, Alexander Lin, Vinod Patel, Richard J. Shaw, Mark S. Chambers, Daniel Ma, Mabi Singh, Noam Yarom, Abdallah Sherif Radwan Mohamed, Katherine A. Hutcheson, Stephen Y. Lai, Clifton David Fuller

## Abstract

**Purpose:** Osteoradionecrosis of the jaw (ORNJ) is a severe iatrogenic disease characterized by bone death after radiation therapy (RT) to the head and neck. With over 9 published definitions and at least 16 diagnostic/staging systems, the true incidence and severity of ORNJ are obscured by lack of a standard for disease definition and severity assessment, leading to inaccurate estimation of incidence, reporting ambiguity, and likely under-diagnosis worldwide. This study aimed to achieve consensus on an explicit definition and phenotype of ORNJ and related precursor states through data standardization to facilitate effective diagnosis, monitoring, and multidisciplinary management of ORNJ.

**Methods:** The ORAL Consortium comprised 69 international experts, including representatives from medical, surgical, radiation oncology, and oral/dental disciplines. Using a web-based modified Delphi technique, panelists classified descriptive cases using existing staging systems, reviewed systems for feature extraction and specification, and iteratively classified cases based on clinical/imaging feature combinations.

**Results:** The Consortium ORNJ definition was developed in alignment with SNOMED-CT terminology and recent ISOO-MASCC-ASCO guideline recommendations. Case review using existing ORNJ staging systems showed high rates of inability to classify (up to 76%). Ten consensus statements and nine minimum data elements (MDEs) were outlined for prospective collection and classification of precursor/ORNJ stages.

**Conclusion:** This study provides an international, consensus-based definition and MDE foundation for standardized ORNJ reporting in cancer survivors treated with RT. Head and neck surgeons, radiation, surgical, medical oncologists, and dental specialists should adopt MDEs to enable scalable health information exchange and analytics. Work is underway to develop both a human- and machine-readable knowledge representation for ORNJ (i.e., ontology) and multidisciplinary resources for dissemination to improve ORNJ reporting in academic and community practice settings.

## INTRODUCTION

Osteoradionecrosis of the jaw (ORNJ) is a morbid iatrogenic disease experienced by cancer patients treated with radiation therapy (RT) to the head and neck region. ORNJ incidence in head and neck cancer (HNC) survivors is estimated to range from 5 to 15% with higher rates associated with risk factors such as poor oral hygiene, pre- and post-RT dental extractions, and high maxillary and/or mandibular radiation dose/volumes.^1–6^ ORNJ can manifest as early as 6 months post-RT, and if not diagnosed and successfully managed at its initial stage, can progress to morbid states of symptom burden and poor quality of life (QOL) via tooth loss, compromised orofacial function, and pain.^7–9^ Financial repercussions of progressive ORNJ are substantial as major medical and surgical interventions for ORNJ can cost up to $170,000 per patient.^10,11^

Until 2023, there was no International Classification of Diseases (ICD) diagnostic code specific to ORNJ, resulting in an inability to formally report and assess ORNJ incidence.^12^ Moreover, there has been no consensus on specific diagnostic criteria that represent different ORNJ disease states, as well as scant agreement regarding severity or grading of ORNJ. This lack of consensus is discernable within existing literature which includes at least 16 ORNJ staging/grading systems published over a span of 4 decades, some of which only include data elements on treatment required or response to therapy in their classification schema.^1,13–27^ This ambiguity in what constitutes minimum data elements (MDEs) to diagnose and stage ORNJ translates to gross variability in estimating the true event rate and poor intelligibility of cross-scale reporting. Ultimately, the state of classification hinders reusability of ORNJ data for comparing outcomes, building data-driven models of disease risk, and facilitating large-scale, multi-institutional interventional trials to mitigate ORNJ.

To address this unmet need, the Orodental Radiotherapy-Associated Late-Effects (ORAL) Consortium was formed, comprised of 69 internationally recognized multidisciplinary experts for consensus formation through a modified Delphi study. The specific aims of this study include:

1. Assessment of contextual overlap between existing ORNJ definitions and disease severity criteria;
2. Characterization of item-specific inter-rater and inter-specialty conceptual agreement across definitional systems for ORNJ;
3. Determination of consensus-derived components of extant and proposed consensus ORNJ grading systems using best informatics principles; and
4. Generation of standardized, clinically relevant criteria (i.e., MDEs) for clinical and radiographic diagnosis, assessment, and reporting of ORNJ across interdisciplinary care.

## METHODS

### The Consensus Process

The consensus process was achieved via Delphi method, and is not an update to pre-existing guidelines.^28–30^ A process flow chart for this study is shown in Figure 1, and an Accurate Consensus Reporting Document (ACCORD) guideline checklist for reporting consensus methods in biomedicine can be found in S1_ACCORD.^31^ A group of international multidisciplinary oncology and oral/dental specialists were invited electronically to participate (n=75). Members of the ORAL Consortium participated in at least one Delphi survey (n=69). Surveys were developed in REDCap® and Qualtrics (see S2 series).^32,33^ Each questionnaire included an introduction, primary objectives for the round, and aggregated group feedback for consensus building. After each round, items meeting consensus were reported in the following rounds as “consensus reports,” and no further questions were asked on those items. Details on Delphi methodology and statistical analysis can be found in S3_Methods. No patients were involved, and this study was approved by our Institutional Review Board (MDA PA 2020-1096).

**Figure 1.**
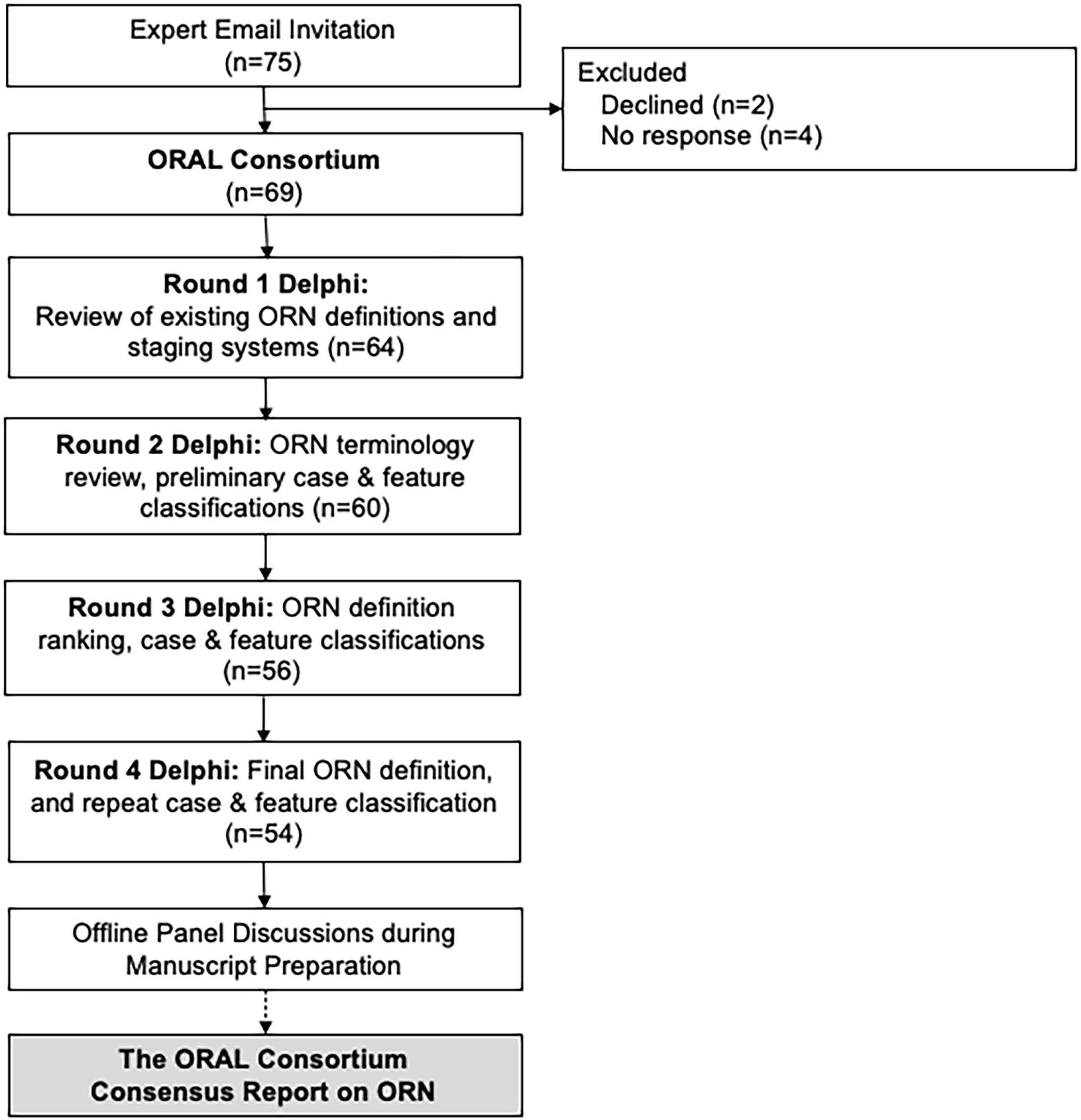
Delphi Consensus Process Flow Chart. Abbreviations: ORN: Osteoradionecrosis.

## RESULTS

### The International ORAL Consortium

Characteristics of the ORAL Consortium are shown in Table 1 with representatives from head and neck surgery, radiation oncology, medical oncology, oral and maxillofacial surgery (OMFS), oral oncology/oral medicine, and other specialties. Nearly half were women (43%), and the average age and time in practice were 47 years and 15 years (range, 0-38), respectively. Experts estimated a 7% annual incidence of ORNJ in their practices and treated a median of 4 cases of ORNJ per year. Participation throughout the study remained high, with 64 (93%), 60 (87%), 56 (81%), and 54 (78%) experts responding to rounds 1 through 4, respectively, and 64% of the Consortium participating in all four rounds.

**Table 1.**
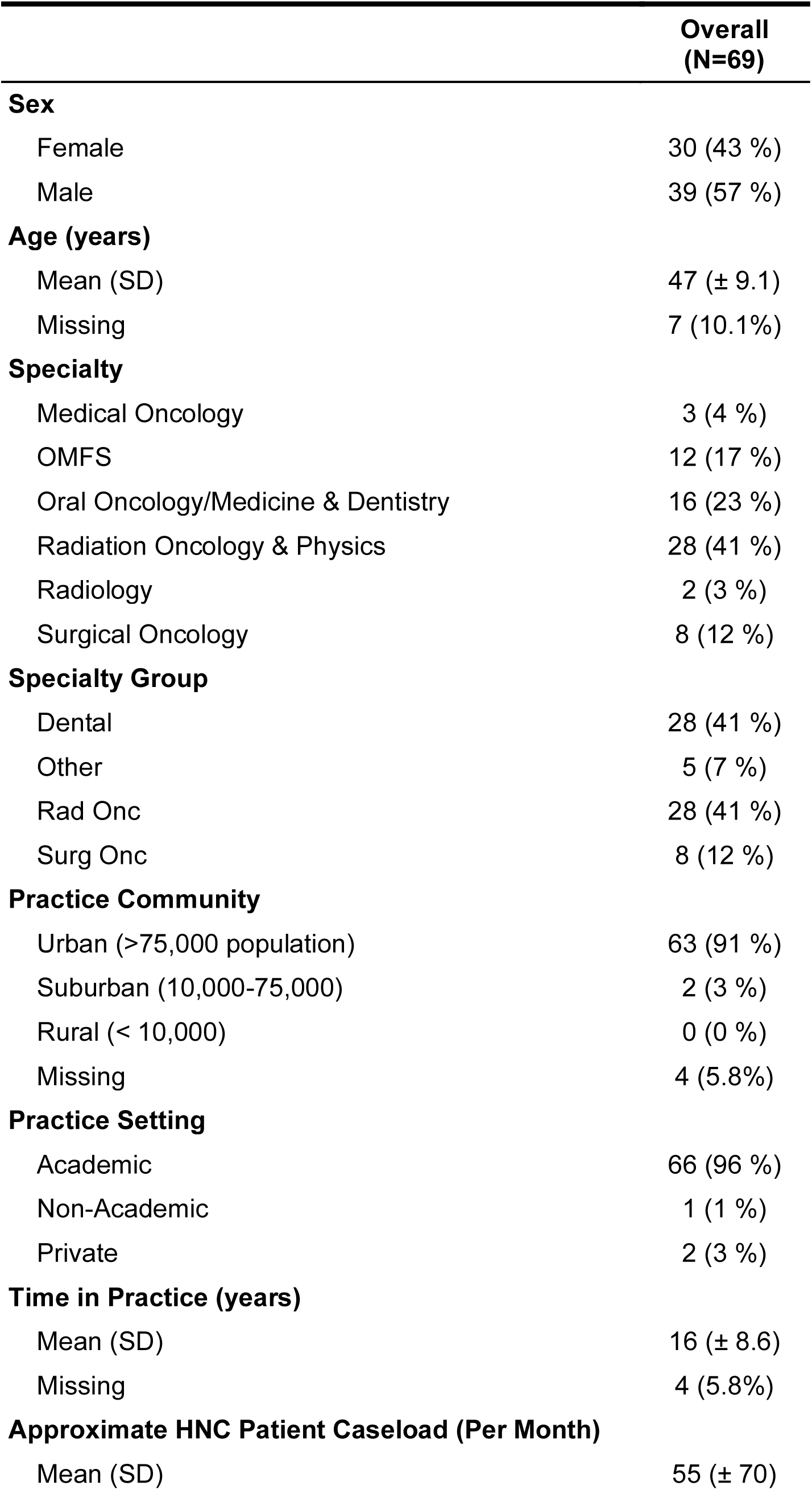
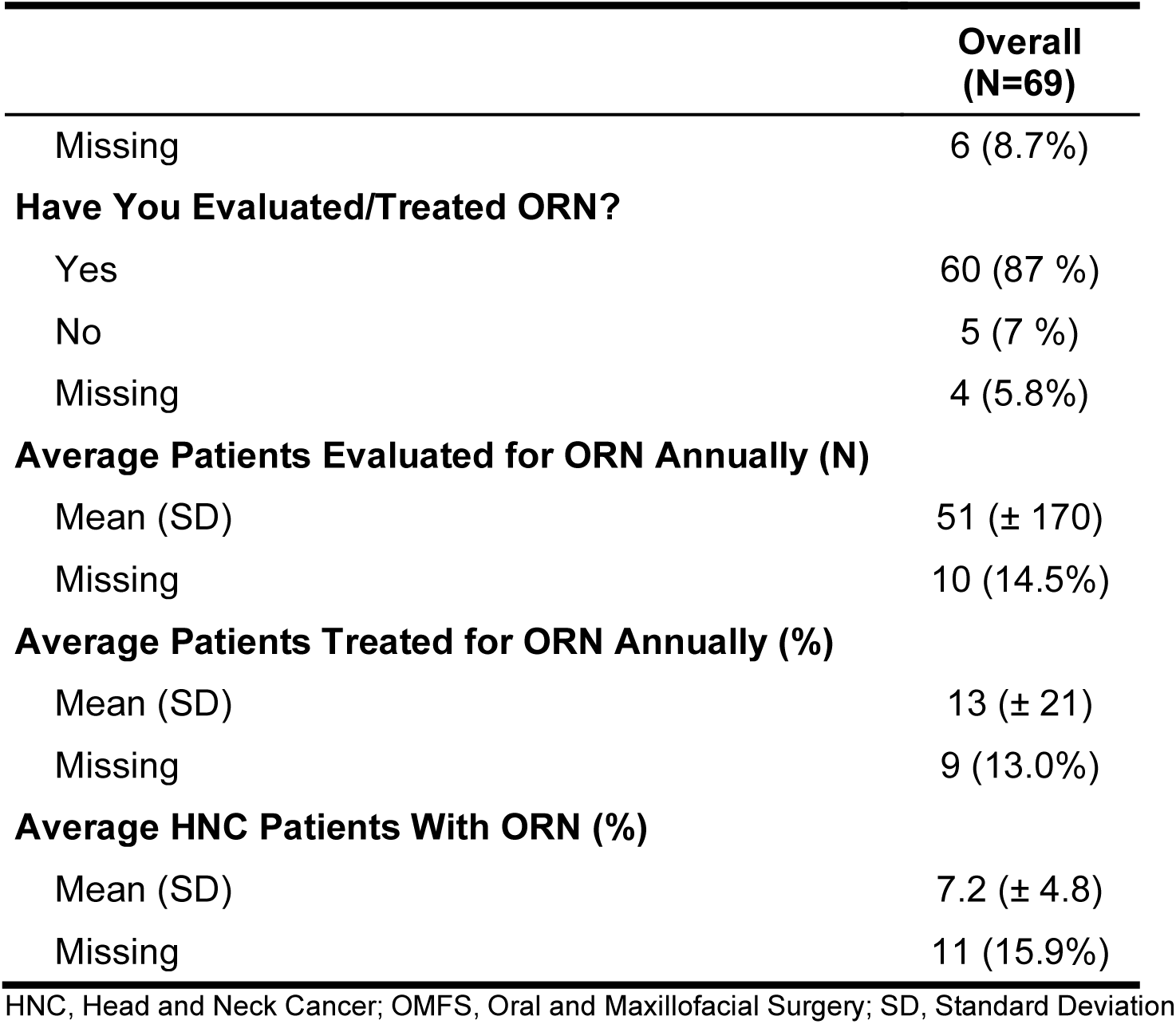
The ORAL Consortium Expert Characteristics.

|  | Overall<br>(N=69) |
| --- | --- |
| **Sex** |  |
| Female | 30 (43 %) |
| Male | 39 (57 %) |
| **Age (years)** |  |
| Mean (SD) | 47 (± 9.1) |
| Missing | 7 (10.1%) |
| **Specialty** |  |
| Medical Oncology | 3 (4 %) |
| OMFS | 12 (17 %) |
| Oral Oncology/Medicine & Dentistry | 16 (23 %) |
| Radiation Oncology & Physics | 28 (41 %) |
| Radiology | 2 (3 %) |
| Surgical Oncology | 8 (12 %) |
| **Specialty Group** |  |
| Dental | 28 (41 %) |
| Other | 5 (7 %) |
| Rad Onc | 28 (41 %) |
| Surg Onc | 8 (12 %) |
| **Practice Community** |  |
| Urban (>75,000 population) | 63 (91 %) |
| Suburban (10,000-75,000) | 2 (3 %) |
| Rural (< 10,000) | 0 (0 %) |
| Missing | 4 (5.8%) |
| **Practice Setting** |  |
| Academic | 66 (96 %) |
| Non-Academic | 1 (1 %) |
| Private | 2 (3 %) |
| **Time in Practice (years)** |  |
| Mean (SD) | 16 (± 8.6) |
| Missing | 4 (5.8%) |
| **Approximate HNC Patient Caseload (Per Month)** |  |
| Mean (SD) | 55 (± 70) |

|  | **Overall**<br>**(N=69)** |
| --- | --- |
| Missing | 6 (8.7%) |
| **Have You Evaluated/Treated ORN?** |  |
| Yes | 60 (87 %) |
| No | 5 (7 %) |
| Missing | 4 (5.8%) |
| **Average Patients Evaluated for ORN Annually (N)** |  |
| Mean (SD) | 51 (± 170) |
| Missing | 10 (14.5%) |
| **Average Patients Treated for ORN Annually (%)** |  |
| Mean (SD) | 13 (± 21) |
| Missing | 9 (13.0%) |
| **Average HNC Patients With ORN (%)** |  |
| Mean (SD) | 7.2 (± 4.8) |
| Missing | 11 (15.9%) |

HNC, Head and Neck Cancer; OMFS, Oral and Maxillofacial Surgery; SD, Standard Deviation

### Consensus-based Definition of ORNJ (high consensus, 86%)

ORNJ is defined *pathognomonically* by the Consortium as “a condition in which there is a loss of blood flow to bone tissue, which causes the bone to die. Findings of bone death may be clinical (i.e., exposed bone) and/or radiographic (i.e., sclerosis, pathologic fracture). It is caused by exposure to ionizing radiation and may occur at some point in time after radiation and in the absence of active disease (i.e., cancer) in the site of bone death.” In contrast to several existing scales, the consensus definition does not require an explicit time duration of exposed bone nor explicit exclusion of concurrent local inflammation or infection (i.e., osteomyelitis). It also incorporates the capacity to formally diagnose ORNJ using imaging criteria; 61/63 (97%) of experts felt it was very/somewhat important to include “radiographic findings” in a formal definition.

Achieving this consensus-based definition required substantial iterative questioning of minimum data elements (MDEs) and derivation of expert- and specialty-specific implicit conceptual frameworks through case-based questioning (see S4_Results). In round 1, six distinct elements were identified; only 3 conceptually included across highly rated definitions/scales: 1) *exposed or necrotic bone*, 2) *RT-induced disorder*, and 3) *absence of tumor* (i.e., primary or recurrence). Round 2 capitalized on MDE identification, asking experts to review MedDRA and SNOMED-CT terminologies/nomenclatures for ORNJ from which 83% (43/52) and 85% (45/53) of panelists agreed that the Consortium’s definition should align with these existing terminologies, respectively. There was also high consensus (80%) that 1) a (then-current) ICD-10 diagnostic code for ORNJ was needed and 2) the National Cancer Institute’s definition for osteonecrosis included a relevant term of ‘vascular insufficiency’ in defining the pathophysiology of bone death.^34^ A total of 87% (52/60) experts agreed this vascular term or ‘devascularization’ should be included in any definition of necrosis. This resulted in the Consortium’s first consensus statement (CS):

**CS 1: The Consortium’s definition for ORNJ will reflect features in existing terminologies, including:**

1. **Bone Disorder**
2. **Radiation injury and/or caused by ionizing radiation**
3. **Loss of blow flow or vascular insufficiency AND findings of bone death/necrosis**

### The Time Feature for Diagnosing ORNJ Conundrum

During round 1, a time feature was identified in 6 of 9 published definitions, and 54 (92%) of experts favored including a time component (i.e., duration of exposure) in the Consortium’s definition for ORNJ. However, the minimum duration of exposed bone required for diagnosing ORNJ varies significantly in the literature from 1-6 months.^15,35–37^ Adding a time feature posed a significant challenge when considering different provider surveillance schedules after RT (time bias). Therefore, a time-feature case scenario (Figure 2) was presented during round 2 to further refine the nuances of time and its relevance as a diagnostic feature for ORNJ. After reviewing 3 existing terminologies, which do not include a diagnostic time feature for ORNJ, 70% (41/59) of experts strongly/somewhat agreed that a diagnosis of ORNJ could be met without a time element while 84% (49/58) agreed a time element is useful for assessing response to therapy, distinct from staging. This analysis resulted in the designation of an MDE for **date_of_assessment** (i.e., date of clinical or radiographic evaluation post-RT) that should be included in ORNJ-related databases for iterative surveillance assessments.

**Figure 2.**
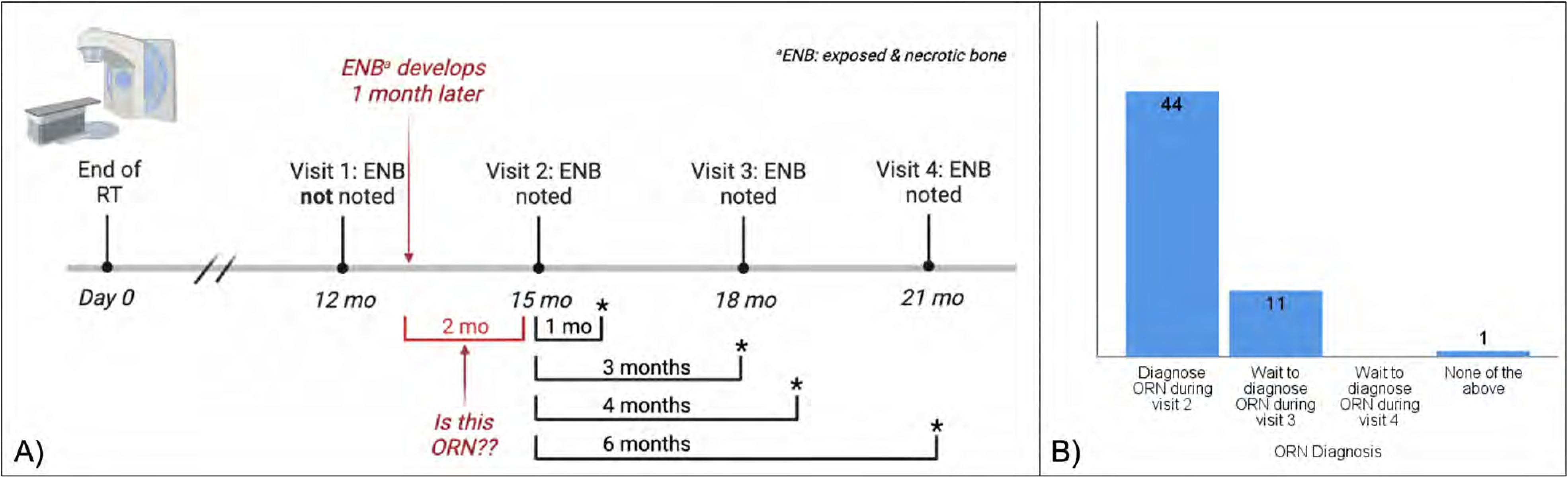
Time Feature and ORNJ Diagnosis Scenario (A) and Expert Panel Consensus (B).

**CS 2: While valuable to report and reflect the duration of non-healing changes observed in irradiated bone, the time or duration of exposed bone is not a mandatory diagnostic feature for ORNJ.**

### Does Exposed Bone Equal Necrotic Bone?

This topic was reviewed to identify differences in conceptualization of exposed bone which may result from other causative agents like trauma after a dental extraction. When asked if all cases of exposed bone automatically equate to necrotic bone, 87% (52/60) disagreed.

**CS 3: Not all cases of exposed bone are necessarily considered to be exposed necrotic bone.**

### Intact Mucosa and Diagnosing ORNJ

Fifty-six (93%) of experts agreed that ORNJ can be diagnosed in cases with intact mucosa. This is significant from a clinical perspective as most published ORNJ definitions include the presence of clinically exposed bone whereas the Consortium’s definition allows for a clinical or radiographic manifestation of features associated with ORNJ. Possible examples include intact mucosa with new lytic and/or mixed sclerotic changes seen on panoramic radiograph or cortical deconstruction of previously irradiated bone noted on surveillance CT scan.

**CS 4: ORNJ can be diagnosed in a patient treated with RT presenting with intact mucosa (i.e., no clinical bone exposure) if there is supporting radiographic evidence of bone death/necrosis.**

### Identification of Elements to Stage ORNJ and Precursor Conditions

#### Review of Existing Staging and Grading Systems

Round 1 provided experts with a comprehensive overview of 15 staging/grading systems. After presenting each classification system, experts were asked 1) if they had ever used the staging/grading system before, 2) to rate the effectiveness of each system for classifying ORNJ, and 3) to classify three clinical scenarios with each system.

Expert utilization and expert-deemed effectiveness/utility ratings of staging/grading systems can be found in Figure S1. The most commonly used system was the Common Terminology Criteria for Adverse Events (CTCAE)^26^ (n=41, 70%) followed by Notani^20^ (n=18, 32%), and Marx^14^ (n=18, 31%). The inability to consistently classify the text-based cases using existing systems was evident during round 1 (Figure 3). The inability to classify ranged from 16-76% (Case 1), 12-70% (Case 2), and 0-62% (Case 3). Interestingly, the CTCAE resulted in higher completion rates of staging cases 1 and 2 with only 16% and 12% of the Consortium reporting an inability to classify the case. Four additional systems were requested by experts and reviewed during round 2.^38–40^ Of these, MRONJ, a staging system for medication-related osteonecrosis, was considered the most effective for diagnosing ORNJ (57%) but its use should be cautioned as it references a distinct causative agent (i.e., medications, not RT). None of the reviewed classification methods met the consensus threshold for being highly effective at staging ORNJ, reflecting a critical need for developing and adopting a comprehensive and updated classification system with explicit clinical and imaging features specified to clearly differentiate between stages. The Watson et al. risk-based model for ORNJ (ClinRad), which was recently published and shown to outperform existing systems for classifying ORNJ severity,^1^ was presented to the Consortium during round 4 and was evaluated favorably with 92% (48/52) of experts agreeing with the clinical-radiographic system.

**Figure 3.**
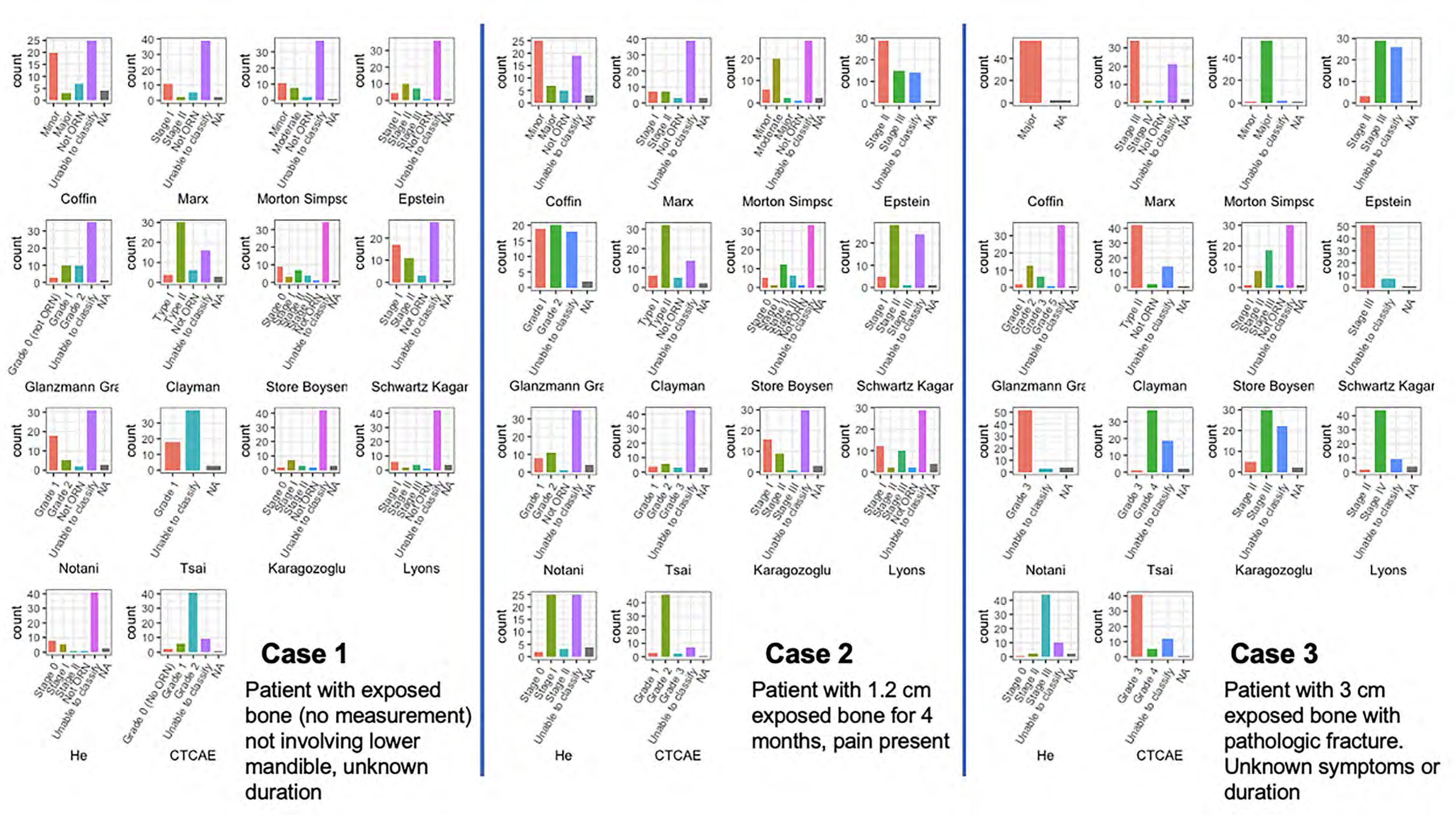
Three Case Classification Using Existing Staging Systems. Magenta bars represent ‘unable to classify’ across all staging systems.

#### Guidelines Regarding Application of the CTCAE Grading System for ORNJ

The potential recommendation to use the CTCAE system to stage ORNJ did not meet consensus (52%). Overall, 90% of experts agreed that the CTCAE system is still valuable for toxicity reporting but not comprehensive enough for classifying ORNJ, and 95% agreed it could be used in parallel with another ORNJ classification method.

**CS 5: CTCAE is a valuable toxicity grading system that should be used in parallel with, but not replace, an ORNJ classification system inclusive of explicit and stage-specific clinical and radiographic features.**

#### Time Feature and ORNJ Staging

Similar time feature-related questions were asked for *staging* disease severity. During round 2, 68% (40/59) agreed that a staging system for ORNJ should be developed without a mandatory inclusion of a time feature. When rephrased to state that a time feature, which is not a clinical exam or radiographic finding, could be an optional modifier but not a necessary feature for staging ORNJ, 83% (47/57) of the Consortium agreed (Figure S2). The Consortium also demonstrated high agreement (85%, 49/58) in considering time features relevant for assessing response to therapy but that therapy response should be separated from a staging system characterizing ORNJ.

**CS 6: A time feature is not necessary for staging ORNJ. However, reporting time features may be complementary for monitoring the duration of observed ORNJ alone or in response to any therapy.**

#### Symptoms and ORNJ Staging

Specific disease-induced symptoms (i.e., pain) or non-explicit symptom presence has been used for upstaging ORNJ.^15,23,24^ When reviewing symptoms, 72% (41/57) of experts agreed that symptoms are ambiguous and therefore should not be stage-defining. However, documenting clear descriptions of specific symptoms including their onset, temporal profiles, and resolution, if any, is highly encouraged in parallel to explicit clinical/radiographic findings so that a **functional** classification system can be developed in the future. For symptom surveillance, the Consortium recommends using standardized assessment tools for patient-reported outcomes (PROs).^41–44^

**CS 7: Symptoms associated with ORNJ should not be used as stage-defining features. However, longitudinal reporting of the presence or absence of concurrent symptoms using validated patient-reported outcomes (PRO) question items is strongly encouraged.**

#### Staging Data Element Extraction and Classification

A data element tracker flowsheet was developed inclusive of all clinical, radiographic, therapy, and treatment response elements identified in reviewed staging/grading systems (Figure S3). Experts were then asked to rate the importance of each feature (not important, somewhat important, very important). Figure S4 shows the distribution of expert responses with only three data elements being considered somewhat/very important by the entire group: pathologic fracture, extent of bone involvement, and exposed bone. A complete description of the MDE development process can be found in S4_Results. Figure 4 shows the heterogeneity in stage and extent of bone involvement classification for ten image-based case studies. A final list of Consortium-approved MDEs for staging ORNJ and precursor stages is shown in Table 2 along with recommended coding standards for building artificial intelligence/machine learning (AI/ML) ready datasets.

**Figure 4.**
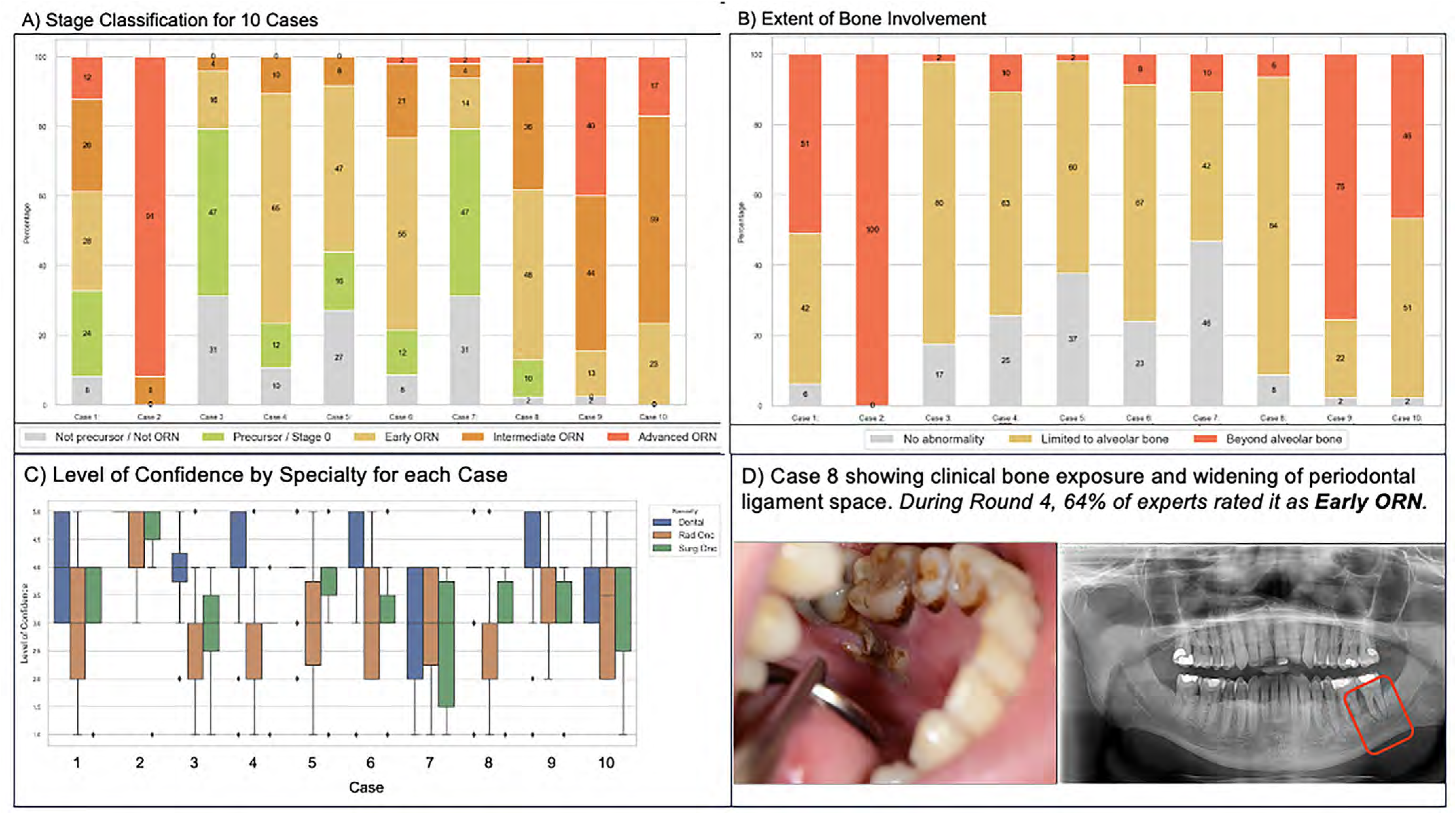
Stage and Bone Extent Classifications and Specialty-Based Levels of Confidence During Round 3.

**Table 2.**
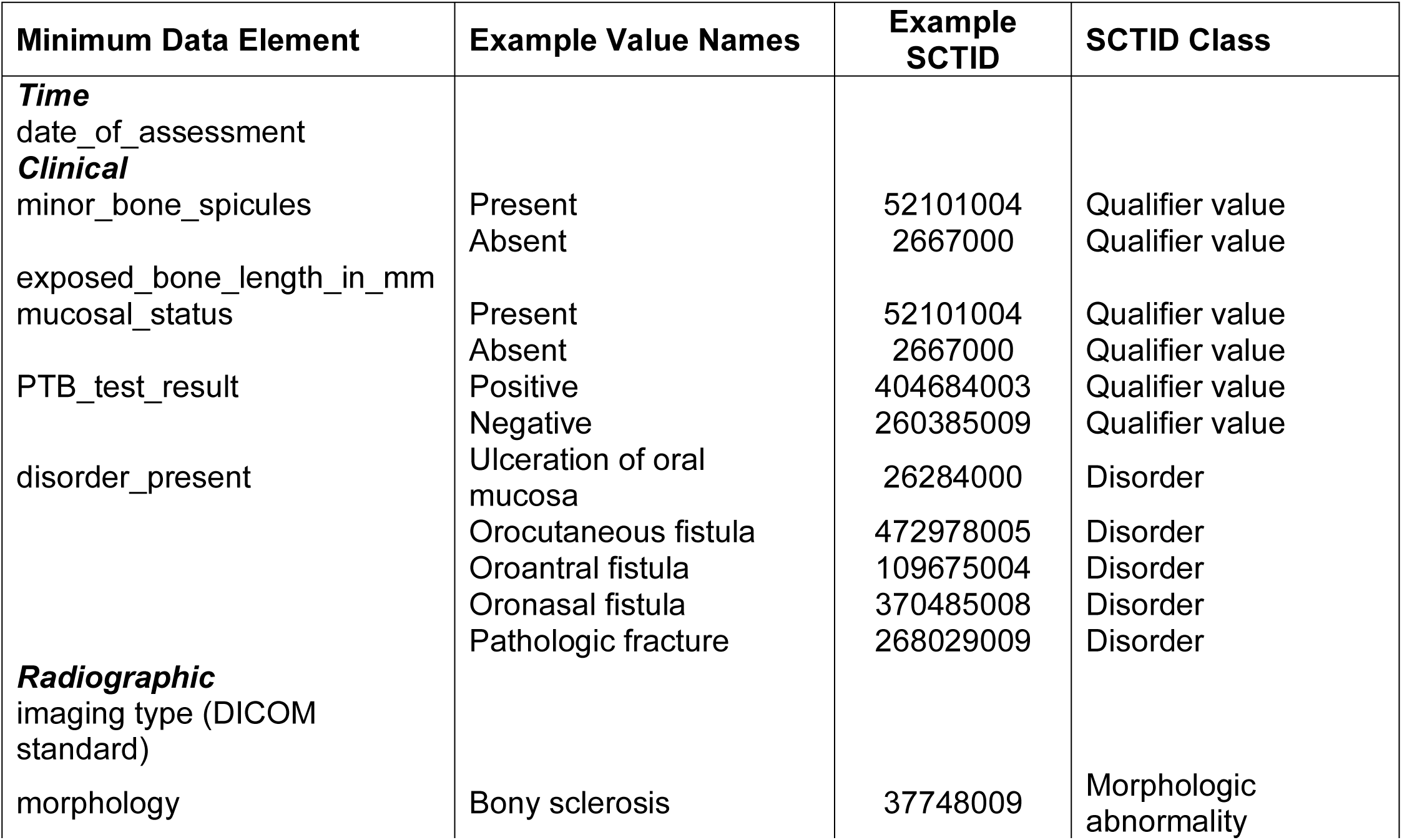
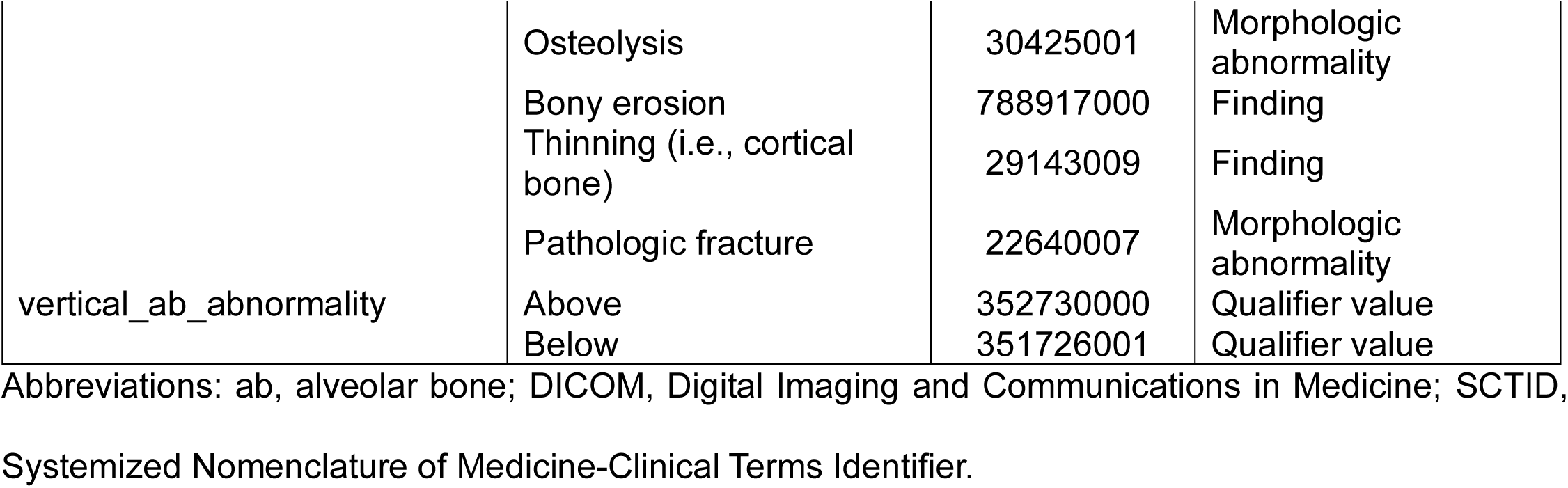
Minimum Data Element List.

**CS 8: All cases with clinical and/or radiographic evidence of a pathologic fracture or fistula formation (i.e., oro-cutaneous, oro-antral, oro-nasal) involving previously irradiated bone should be reported as advanced-stage ORNJ. These individual disorders are considered stage-defining MDEs, and each should be reported separately.**

### Specialty-Specific Knowledge Siloes & Inter-Rater Reliability

Siloes of knowledge may occur through different knowledge acquisition per specialty-based training programs and/or practice patterns. Within the ORAL Consortium, a significant difference was found in the clinical utilization of different imaging modalities (i.e., CT, MRI, panoramic radiograph; see S4_Results). Educational resources were provided to improve inter-rater reliability between rounds. With regards to the image-focused educational resources, 84% of experts found the resources helpful, and 92% were interested in having an updated, comprehensive, multidimensional atlas as a support tool for case classification.

### Recommendation for Multidisciplinary MDE Adoption

Given substantial variation in the classification of cases among experts, particularly ‘threshold’ cases which may be upstaged based on non-explicit clinical and/or radiographic imaging features (i.e., quantitative measurement of clinical bone exposure), the following consensus statements were developed:

**CS 9: The Consortium strongly recommends the adoption of ORNJ-focused MDEs in multidisciplinary clinical practice and clinical trial design to reduce misclassification risks and to facilitate ‘stage migration’ across classification models.**

**CS 10: Inclusion of serial photographs in a patient’s medical record during post-RT surveillance, especially once changes in the mucosa (i.e., ulceration) or bone (i.e., progressive bone exposure) are detected, is strongly recommended. Caliper or ruler-based measurements of clinical bone exposure should also be recorded for at least the longest dimension in millimeters under the MDE, clinical:exposed_bone_length_in_mm.**

## DISCUSSION

In this expert-based, iterative Delphi method study, we have generated an international, multidisciplinary-approved definition for ORNJ along with ten consensus statements and nine distinct minimum data elements that should be serially documented during dental and oncology post-RT appointments for cancer survivors undergoing ORNJ surveillance. These MDEs characterize static (i.e., date of assessment) and dynamic (i.e., progressive radiographic changes) features that can be used for meaningful classification of ORNJ and precursor stages.

During this Delphi modeling, several authors were simultaneously involved in the development and publication of the ClinRad model^1^ and the ISOO-MASCC-ASCO joint guideline for prevention and management of ORNJ.^9^ The Consortium’s definition for ORNJ is in alignment with the new ISOO-MASCC-ASCO guideline, which *operationally* characterizes ORNJ as a “radiographic lytic or mixed sclerotic lesion of bone and/or visibly exposed bone and/or bone probed through a periodontal pocket or fistula occurring within an anatomical site previously exposed to a therapeutic dose of head and neck radiation therapy.” Moreover, both definitions demonstrate a significant departure from using a time-limiting feature for diagnosing ORNJ, thereby addressing current issues with time bias.

The ClinRad system, which outperformed staging methods such as the Notani, LENT-SOMA, and Store systems, incorporates observable clinical (i.e., Probe-to-Bone [PTB] tests) and radiographic features and uses the alveolar bone as a distinguishable threshold for disease.^1^ This new staging model of ORNJ has been adopted by the new ISOO-MASCC-ASCO ORNJ guidelines.^9^ The ORAL Consortium’s favorable review of the ClinRad system prior to the publication of the guidelines further supports its utilization as it incorporates most of the MDEs identified in this study. However, we also demonstrate several staging challenges that should be addressed by providers prior to implementing the ClinRad system. There still exists a conceptualization discordance for MBS, namely whether or not it is related to ORNJ. ^1^The Consortium favors the classification of MBS as a precursor event to ORNJ that has a higher likelihood of resolving over time compared to other MDE features or feature combinations. More importantly, utilization of a quantitative measurement like **clinical:exposed_bone_length_in_mm** and inclusion of clinical photographs in the patient’s medical and dental records can facilitate data harmonization among interprofessional healthcare providers at different centers treating the same patient.

The concept of specialty-specific knowledge siloes is introduced in this study and is important to recognize since diagnosis and management of ORNJ is often a multidisciplinary task. Overall, providers tend to exhibit high agreement in perceiving severe presentations of ORNJ; however, subtle variations in physical exam or radiographic feature interpretations may cascade into differing classifications of disease for the same patient examined by different specialists. One approach to mitigate this discrepancy is by standardizing the use of MDEs across specialties and generating interdisciplinary, multi-modality (i.e., OPG, CT, MRI) image-focused educational materials. Biomarkers for ORNJ and its precursor stages are also being investigated, including dynamic contrast-enhanced (DCE) MRI parameters (K_trans_ and V_e_,) for assessing risk, diagnosis, and progression or treatment response of ORNJ.^45^ The Consortium supports the consideration of DCE-MRI changes indicative of vascular insufficiency in previously irradiated jaw bones as a precursor event to ORNJ. Subsequent clinical guidelines will be necessary for outlining optimal MRI-based ORNJ surveillance regimens and specification of MRI-specific MDEs.

While this interdisciplinary Delphi study has several strengths such as the Consortium size (n=69) and sustained level of engagement, there is underrepresentation of specialties (e.g. radiology (n=1) that may provide additional expertise on identifying stage-defining radiographic features across imaging modalities. Fleiss kappa statistics can provide substantial insight on how reliability experts classify cases (i.e., interrater reliability), but it does not provide information on whether those classifications represent the true disease state (i.e., validity).^46^ Lastly, while several consensus-defining methods exist,^47,48^ we chose a simple agreement threshold as it is commonly used, easy to interpret, and reinforced through iterative requestioning to produce metrics of reliability.

In conclusion, the Consortium’s definition of ORNJ and associated MDEs should be adopted as standards for reporting by head and neck surgery, oncology, radiology, and dental providers in clinical practice, research, and clinical studies. Collectively, these enable scalable health information exchange and AI/ML data readiness for rigorous modeling of the disease and its precursor stages. ORNJ-focused MDEs are also synergistic with recently published guidelines and newer risk-based ORNJ models that recognize the importance of combining clinical and radiographic features for ORNJ characterization. Lastly, additional efforts are underway to formalize an ORNJ ontology, develop radiology standards and automated imaging feature identification and reporting, and formulate and disseminate interdisciplinary educational resources to mitigate barriers to accurate ORNJ staging.

## Data Availability

Data availability statement: The data that support the findings of this study are openly available in an NIH-supported generalist scientific data repository [figshare] at http://doi.org/10.6084/m9.figshare.25546723; though embargoed pending peer-review, data is available upon request in the interim.

## Data availability statement

In accordance with the *Final NIH Policy for Data Management and Sharing* NOT-OD-21-013, data that support the findings of this study are openly available in an NIH-supported generalist scientific data repository (figshare) at http://doi.org/10.6084/m9.figshare.25546723 no later than the time of an associated publication.

## Reporting guideline compliance statement

In accordance with EQUATOR Network (*Enhancing the QUAlity and Transparency Of health Research*) guidance, we have utilized the ACCORD checklist *<u>(“ACcurate COnsensus Reporting Document): A reporting guideline for consensus methods in biomedicine developed via a modified Delphi”</u>,* Gattrell *et al.,* https://www.ismpp.org/accord); the completed ACCORD checklist is attached as Supplementary file and deposited via figshare at http://doi.org/10.6084/m9.figshare.25546723; during peer review, referees may access this checklist form via private link at https://figshare.com/s/625834718e80c583198d during the peer review process.

## CRediT statement

In accordance with the Contributor Roles Taxonomy (CRediT, https://credit.niso.org/), the contributing authors have designated responsibilities and individual author attribution. The corresponding authors (ACM, CDF) assume responsibility for role assignment, and all contributors have been given the opportunity to review and confirm assigned roles:

*Conceptualization*: ACM and CDF; *Data curation*: ACM; *Formal analysis*: ACM; *Funding acquisition*: ACM and CDF; *Investigation*: ACM, EEW, LH, DEP, LVV, TG, LVdB, AJH, MK, FH, RAA, JEB, PB, AFD, MD, ERF, TJG, DYG, IAH, CH, SK, AK, NYL, CML, CDL, AAM, MM, YMM, CM, JM, EO, JP, JMR, DS, JDS, US, ES, VT, JT, GV, AV, NW, MW, AW, MEW, CMKLY, SY, KA, CEAB, DB, FJD, TH, AK, AO, JCR, LH, JJ, RL, AL, VP, RS, MC, DM, MS, NY, ASRM, KAH, SYL, and CDF; *Methodology*: ACM and CDF; *Project administration*: ACM and CDF; *Resources*: ACM, EEW, LH, DEP, LVV, TG, LVdB, AJH, MK, FH, RAA, JEB, PB, AFD, MD, ERF, TJG, DYG, IAH, CH, SK, AK, NYL, CML, CDL, AAM, MM, YMM, CM, JM, EO, JP, JMR, DS, JDS, US, ES, VT, JT, GV, AV, NW, MW, AW, MEW, CMKLY, SY, KA, CEAB, DB, FJD, TH, AK, AO, JCR, LH, JJ, RL, AL, VP, RS, MC, DM, MS, NY, ASRM, KAH, SYL, and CDF; *Supervision*: ACM and CDF; *Writing – original draft*: ACM, EEW, DEP, KAH, and CDF; *Writing - review & editing*: ACM, EEW, LH, DEP, LVV, TG, LVdB, AJH, MK, FH, RAA, JEB, PB, AFD, MD, ERF, TJG, DYG, IAH, CH, SK, AK, NYL, CML, CDL, AAM, MM, YMM, CM, JM, EO, JP, JMR, DS, JDS, US, ES, VT, JT, GV, AV, NW, MW, AW, MEW, CMKLY, SY, KA, CEAB, DB, FJD, TH, AK, AO, JCR, LH, JJ, RL, AL, VP, RS, MC, DM, MS, NY, ASRM, KAH, SYL, and CDF.

## Acknowledgements

We would like to acknowledge Dr. Barbara Murphy for her contributions to round 1 of this study.

## ICJME author statement

In accordance with International Committee of Medical Journal Editors (ICJME, https://www.icmje.org/) recommendations, all authors affirm qualification for authorship via the following criteria: *“Substantial contributions to the conception or design of the work; or the acquisition, analysis, or interpretation of data for the work; AND Drafting the work or reviewing it critically for important intellectual content; AND Final approval of the version to be published; AND Agreement to be accountable for all aspects of the work in ensuring that questions related to the accuracy or integrity of any part of the work are appropriately investigated and resolved.”*

---

**Figure S1.**
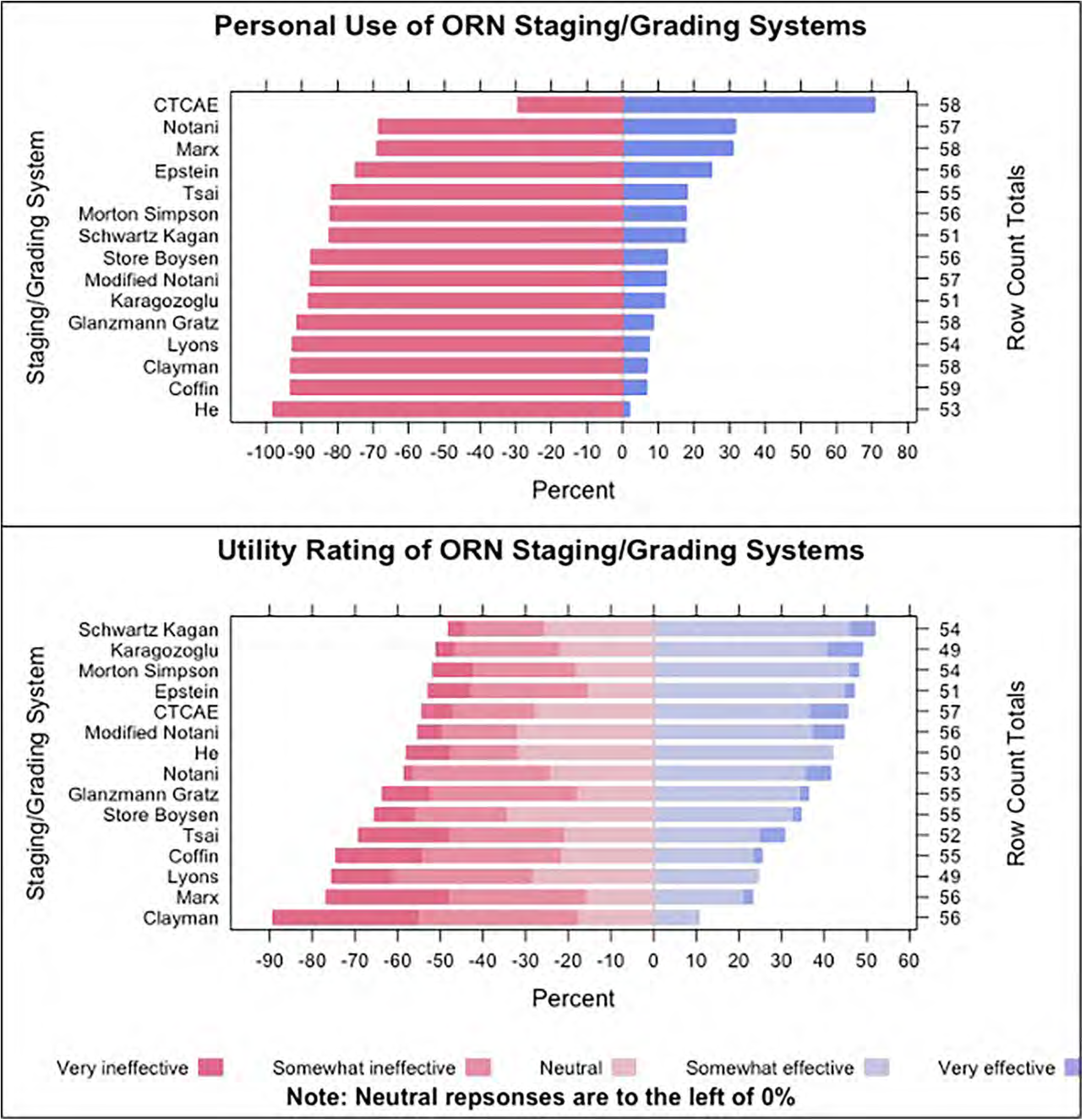
Personal Use and Utility Rating of Existing ORN Staging/Grading Systems.

**Figure S2.**
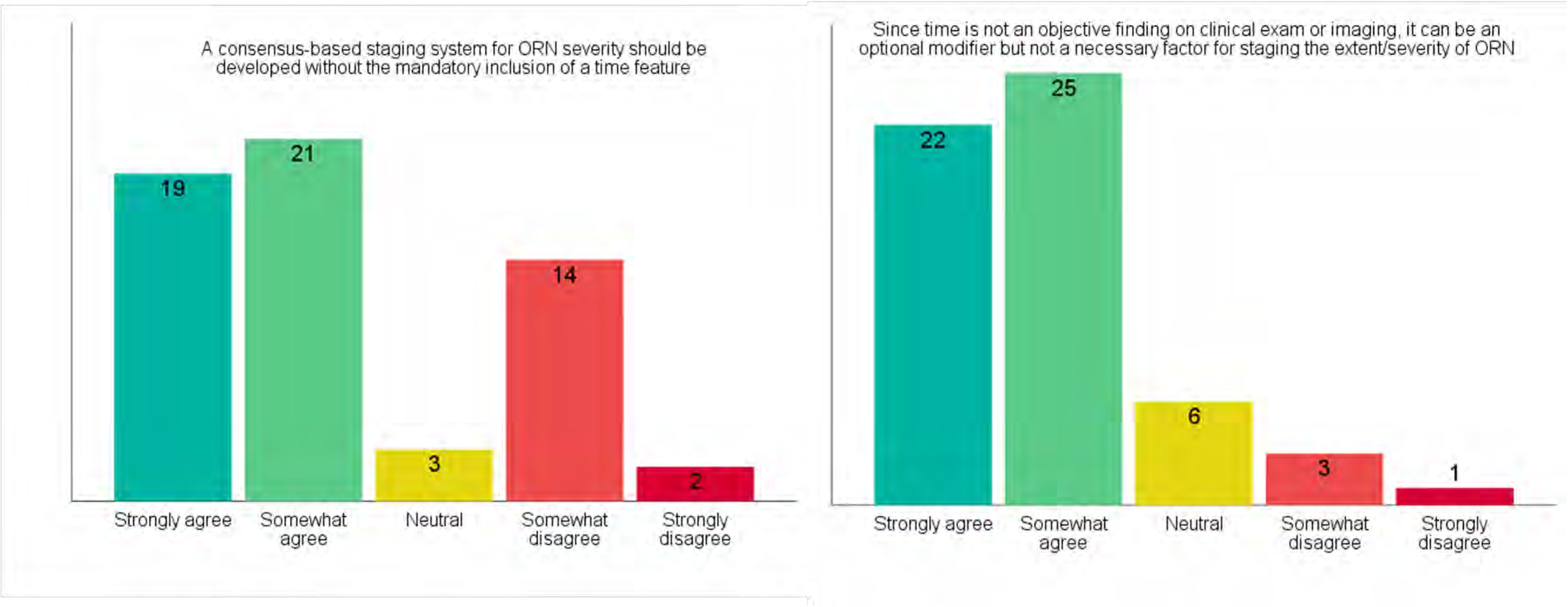
The Time Feature and ORN Staging.

**Figure S3.**
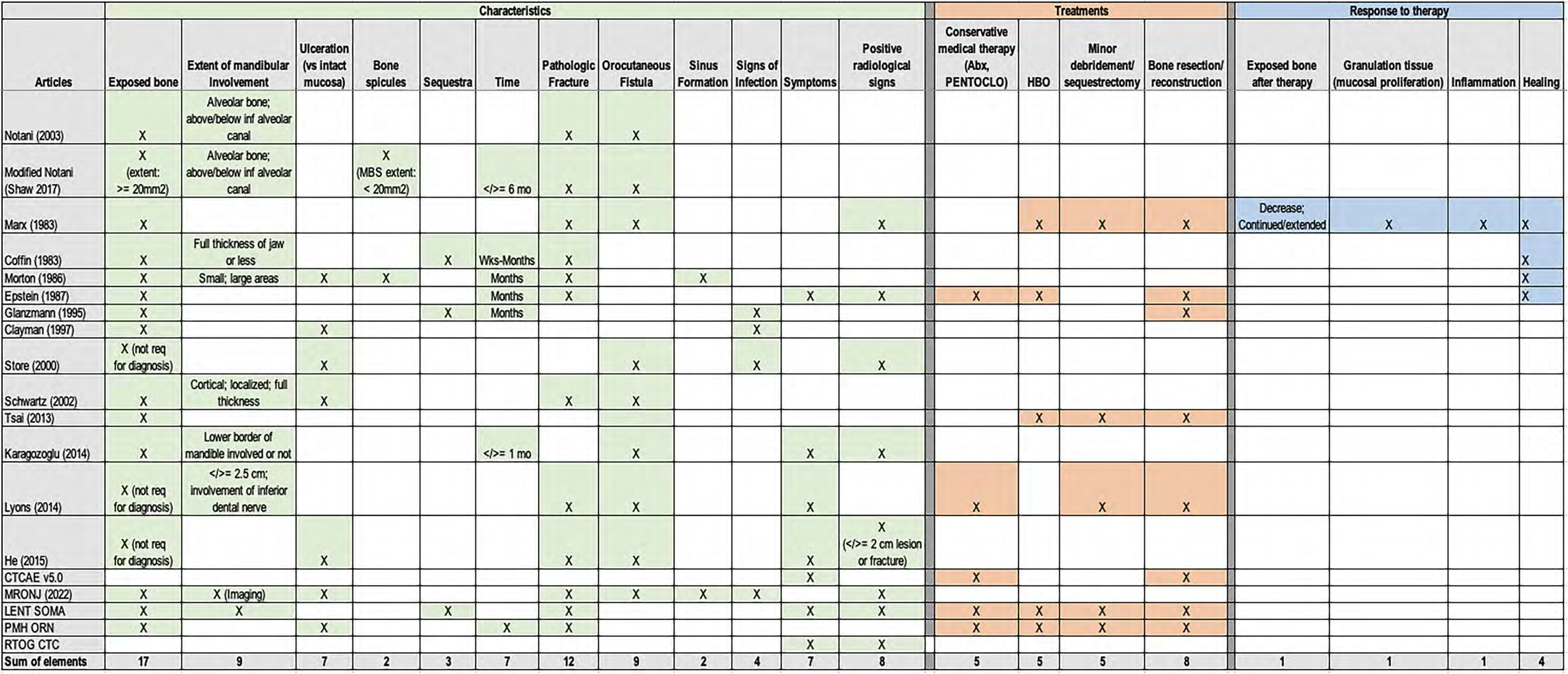
Data Element Tracker.

**Figure S4.**
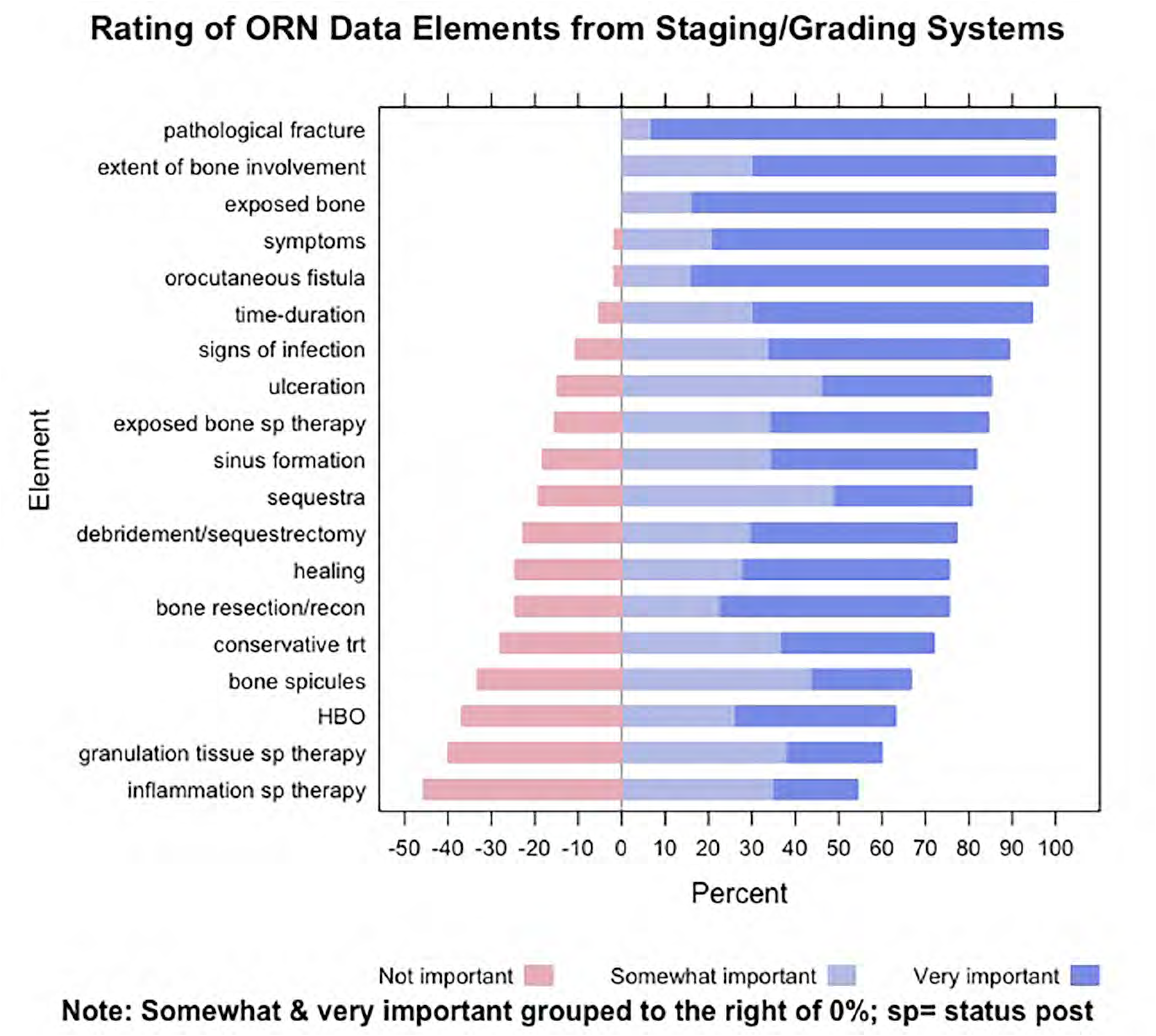
Rating of ORN Data Elements from Staging/Grading Systems.

**Figure S5.**
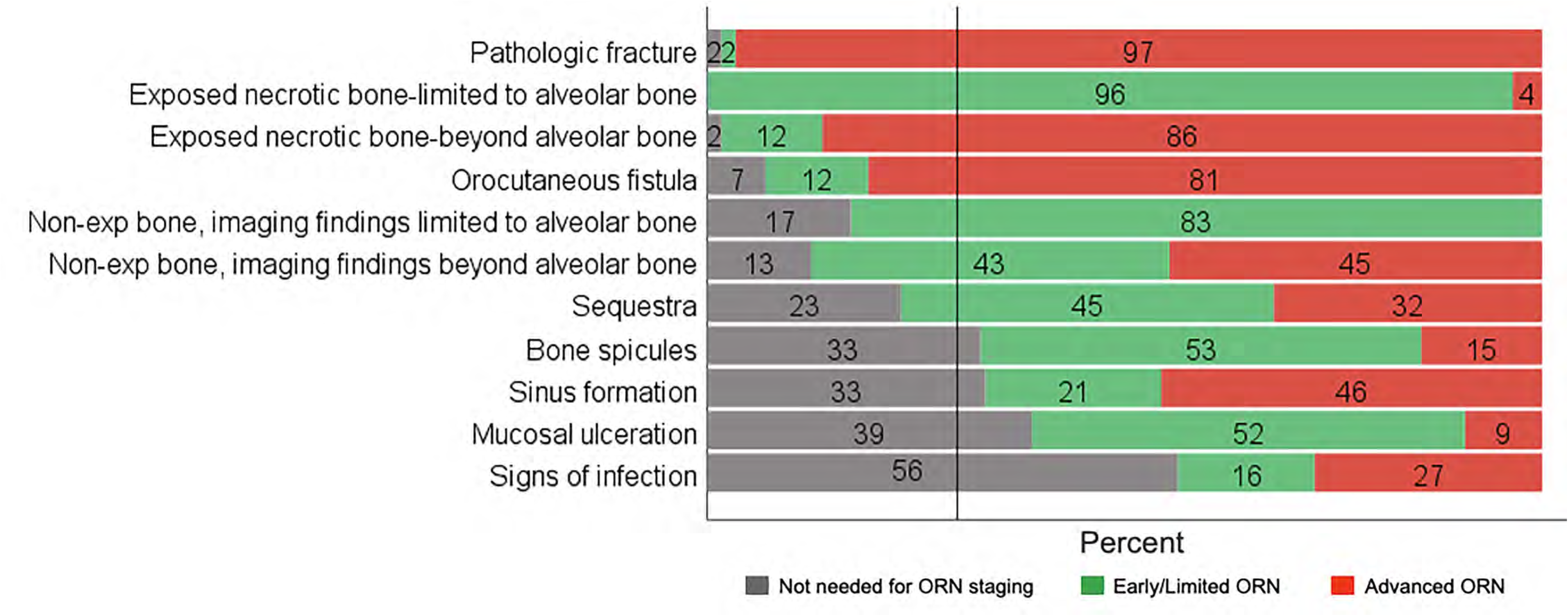
Round 2 Data Element Classification.

**Figure S6.**
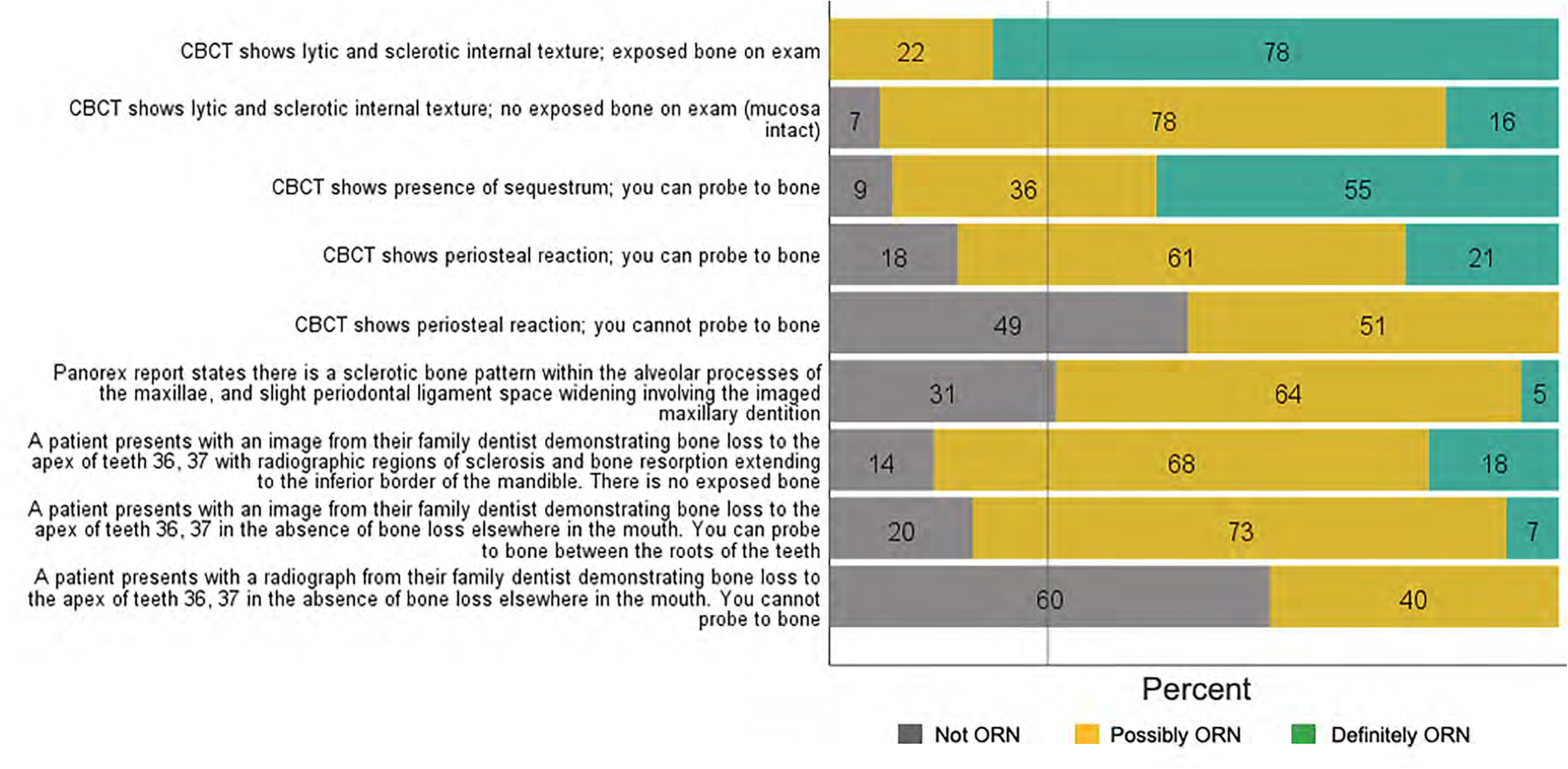
Round 2 Scenario Classification.

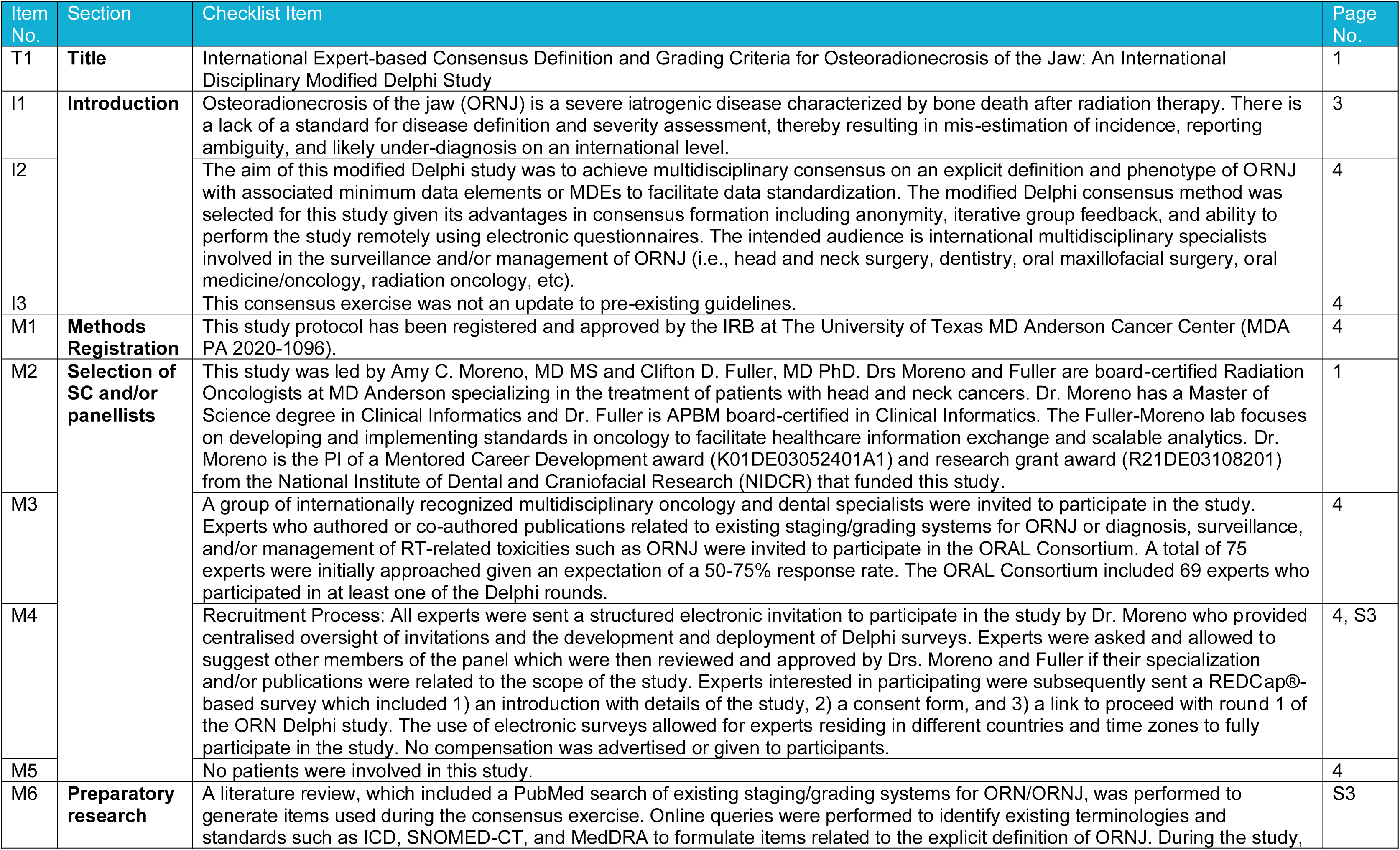
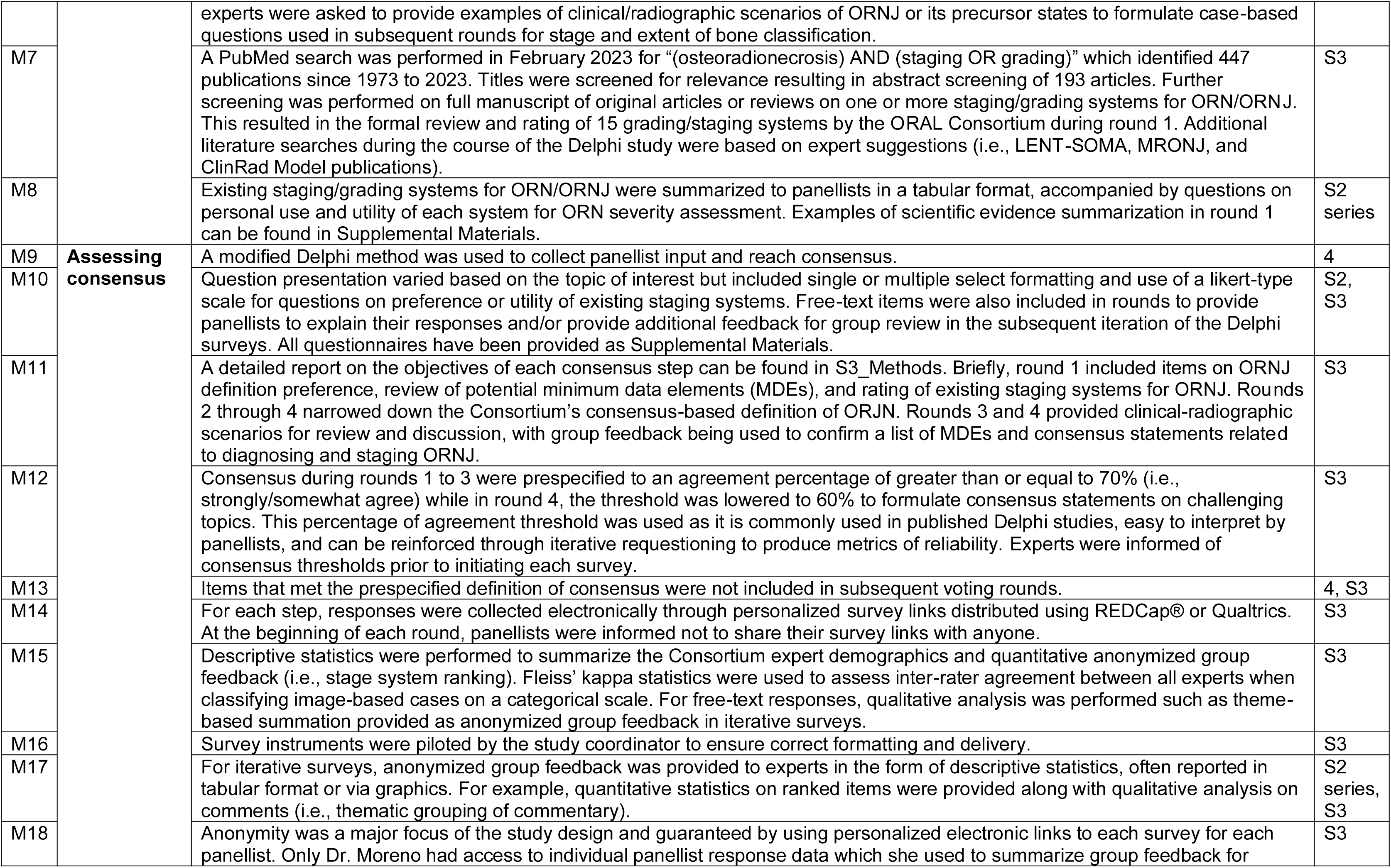
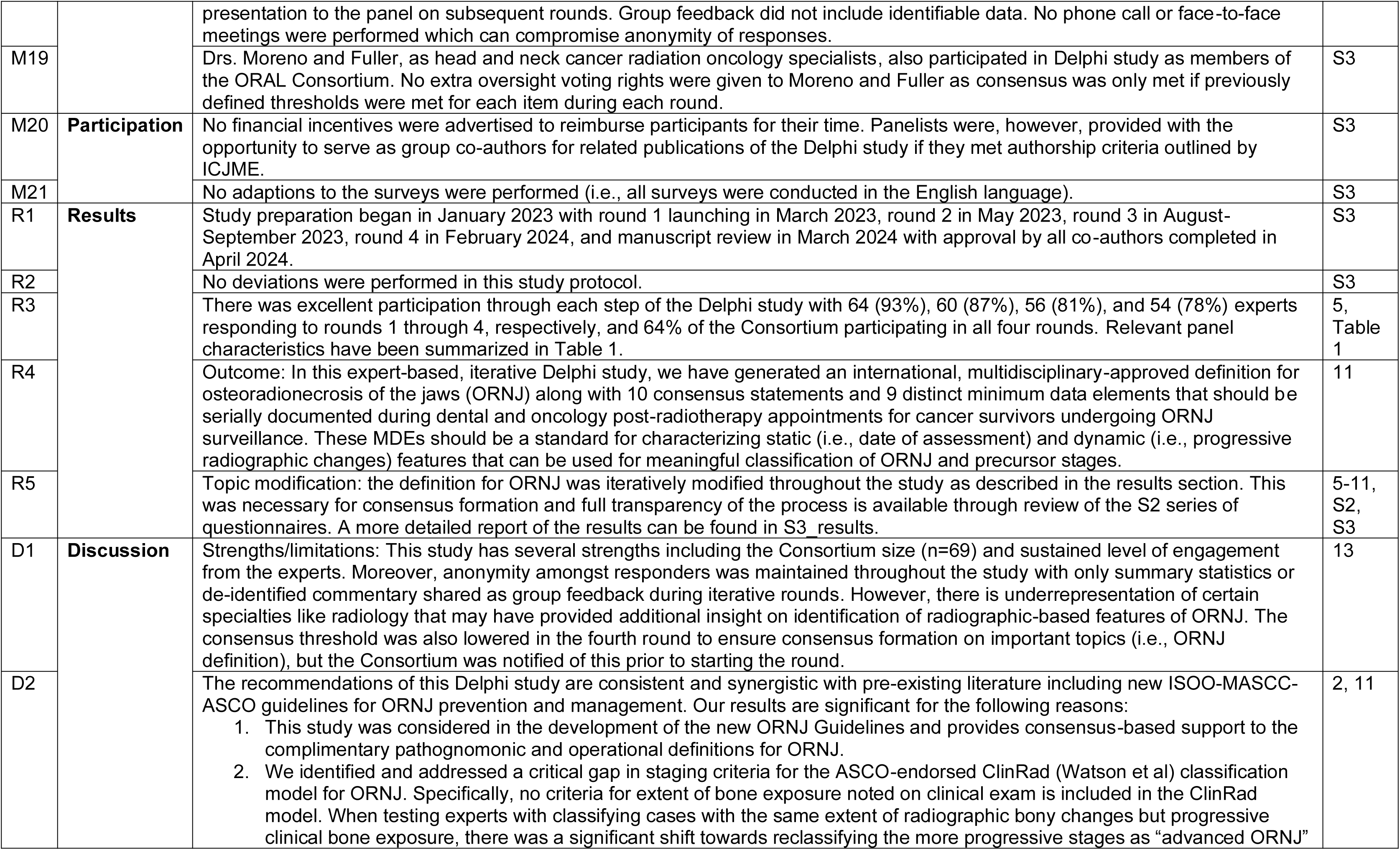
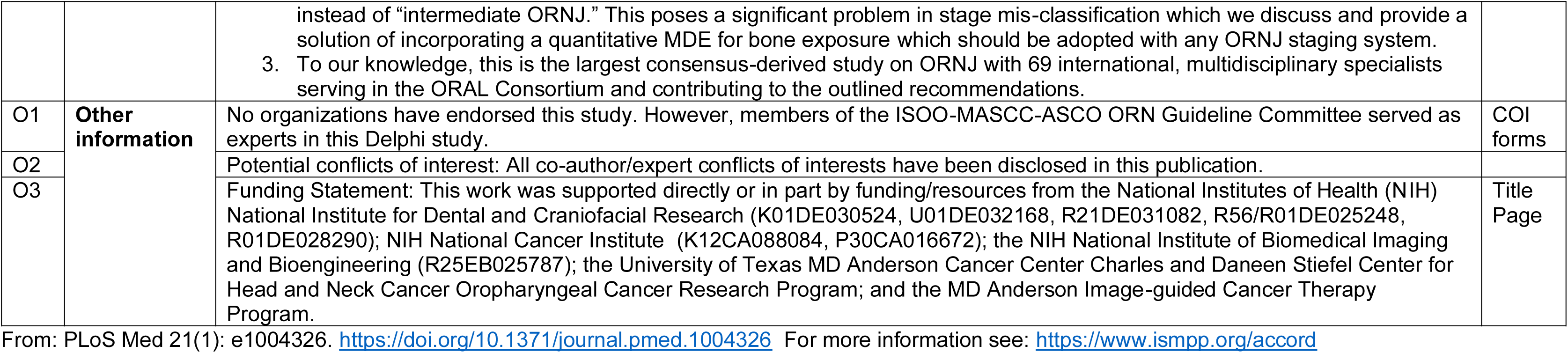

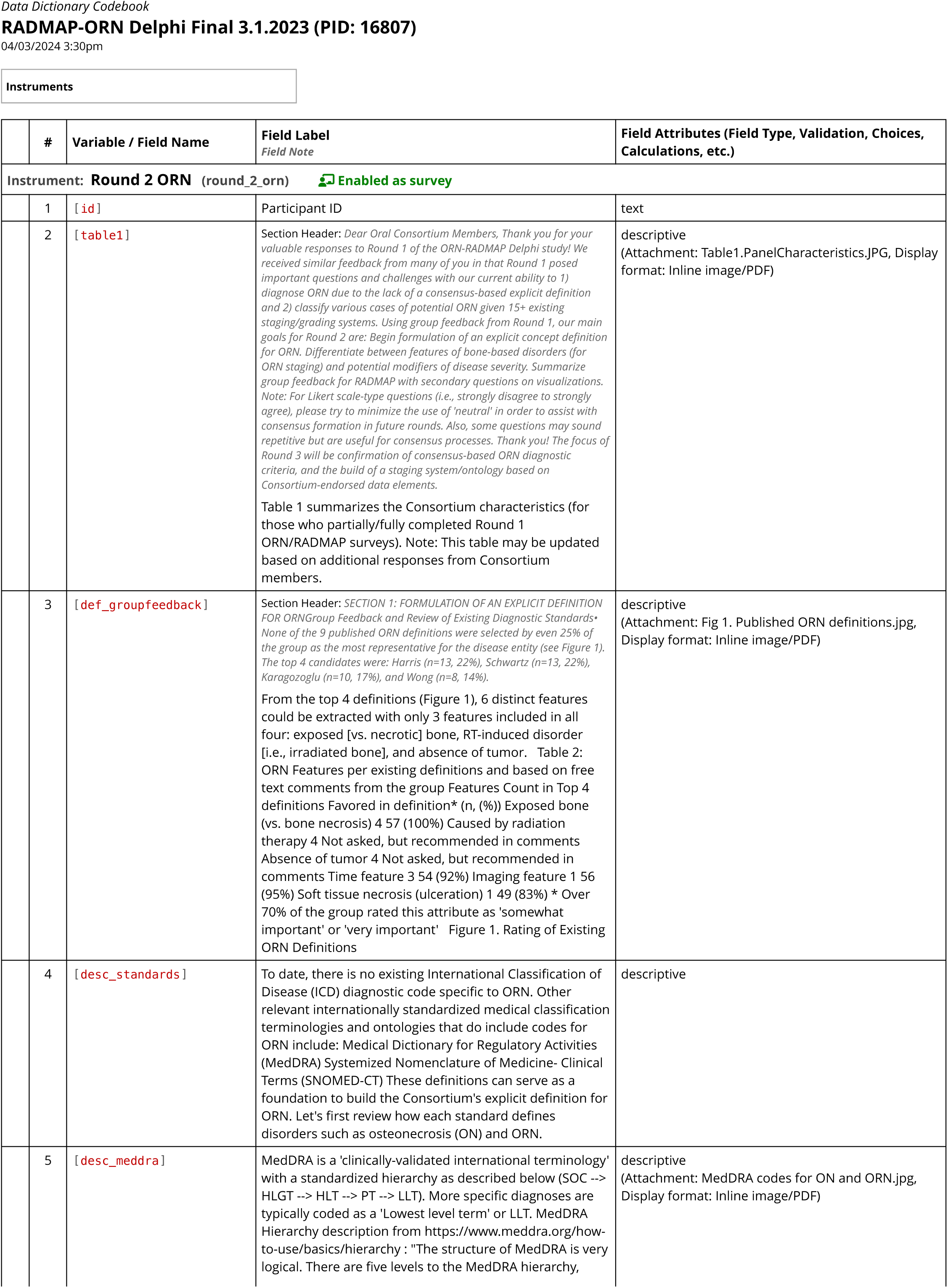
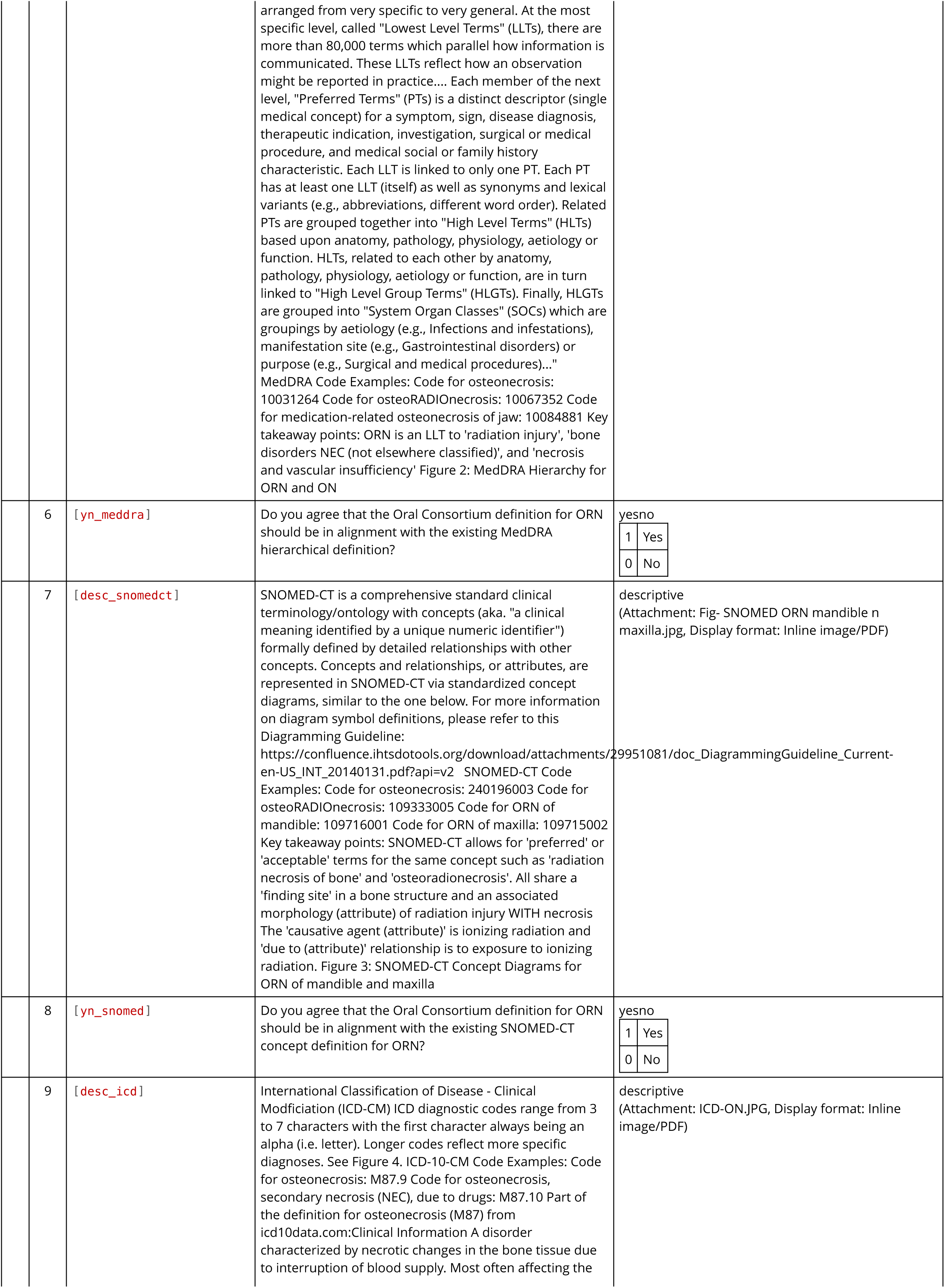
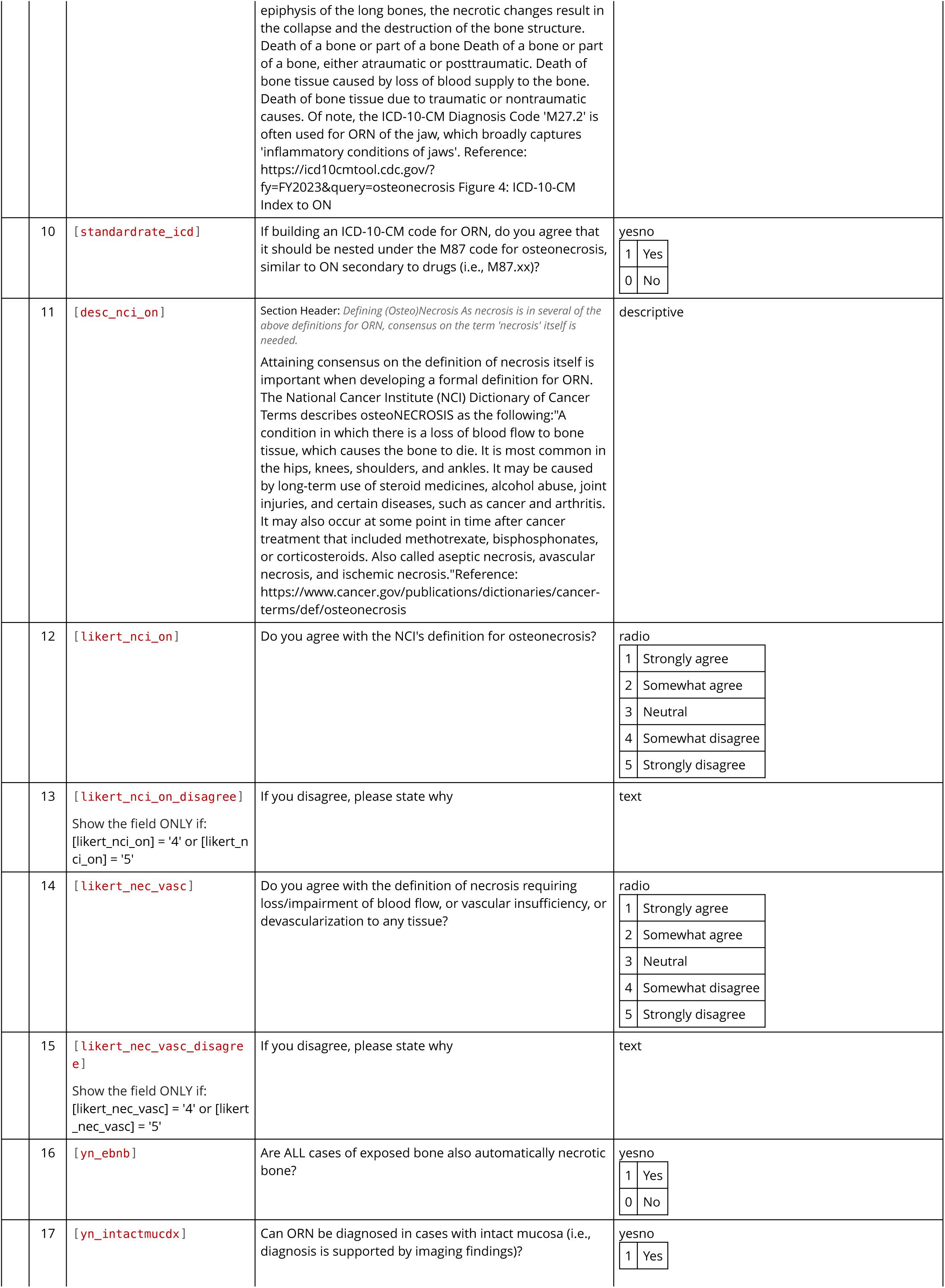
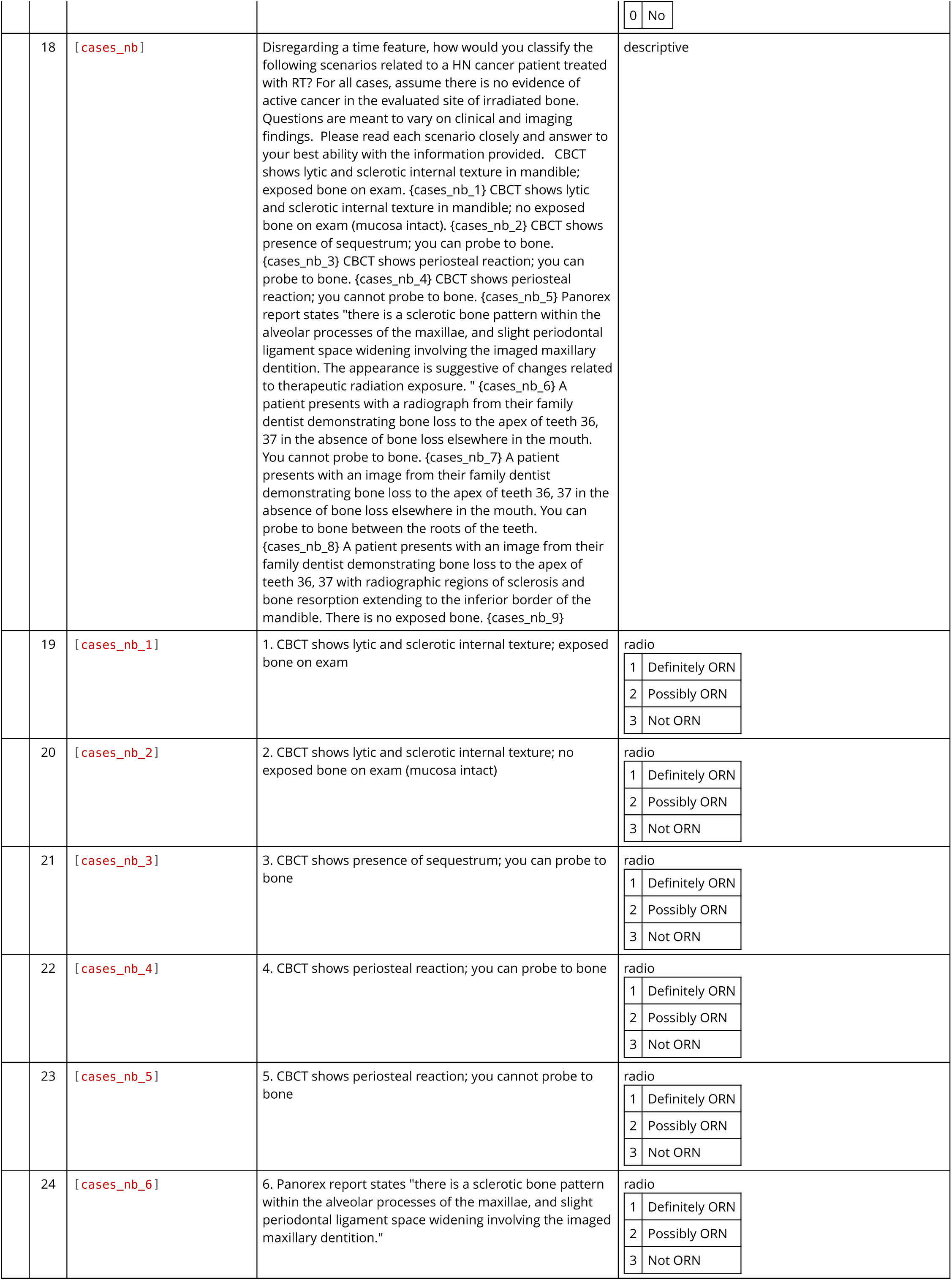
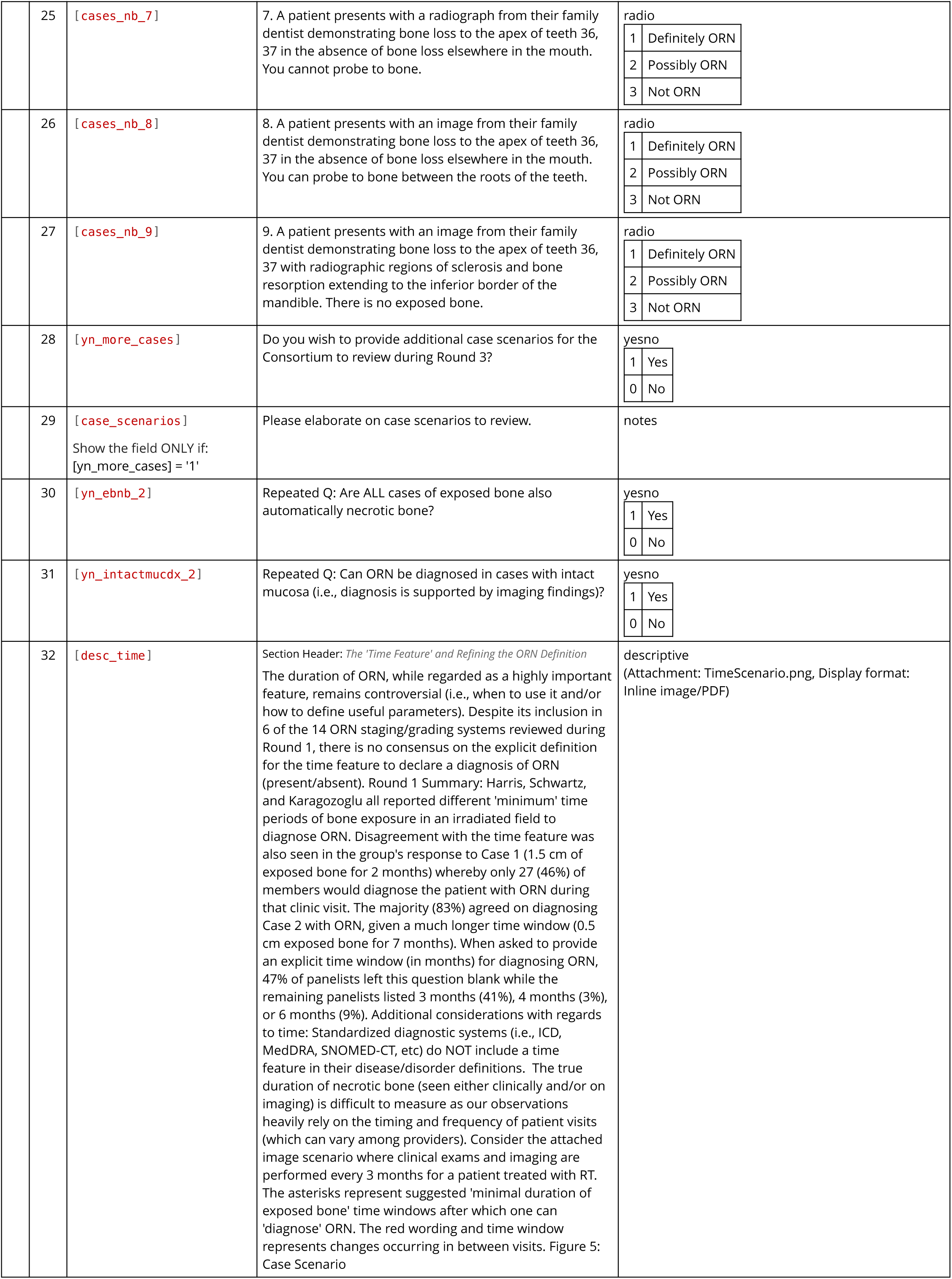
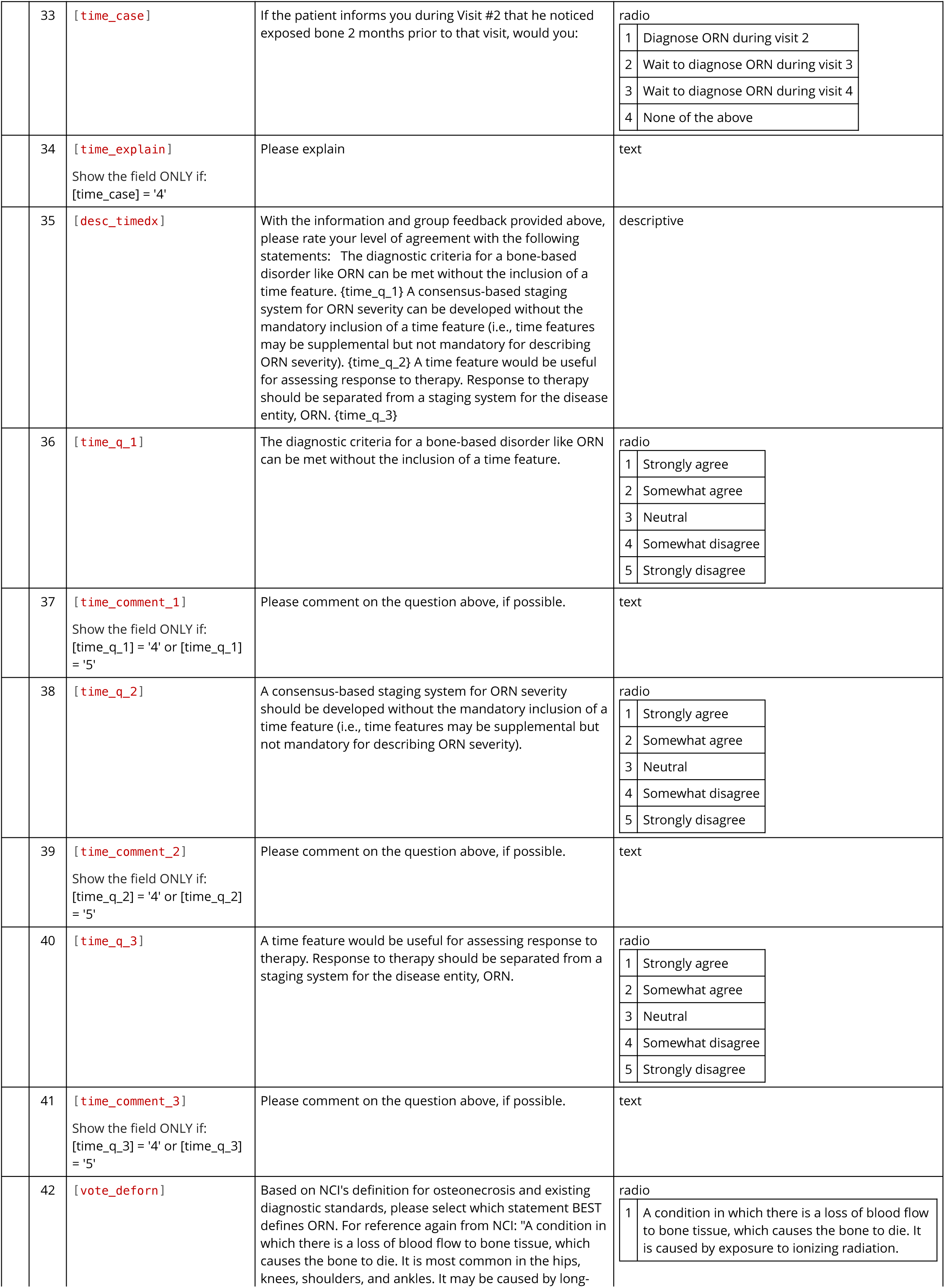
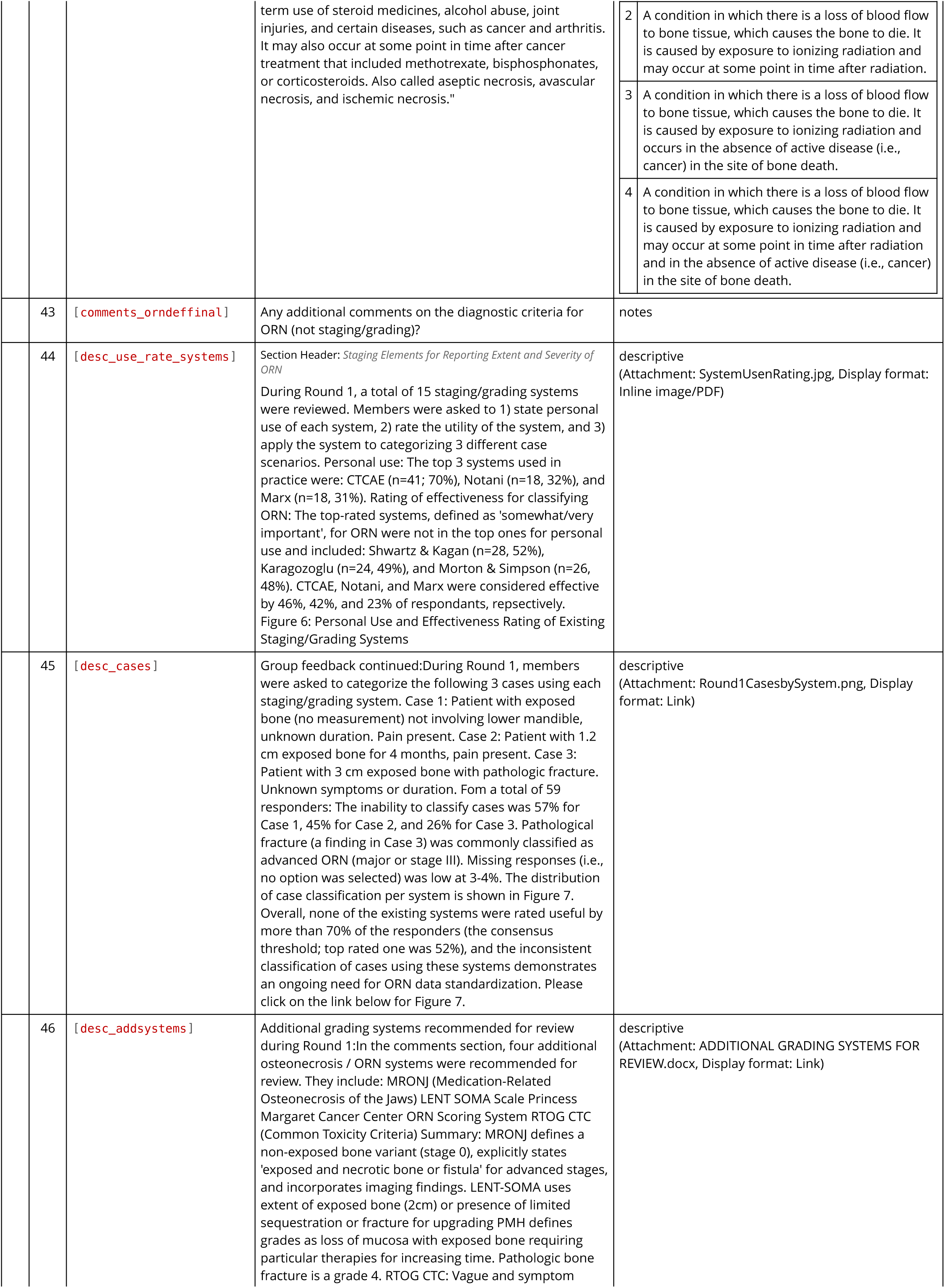
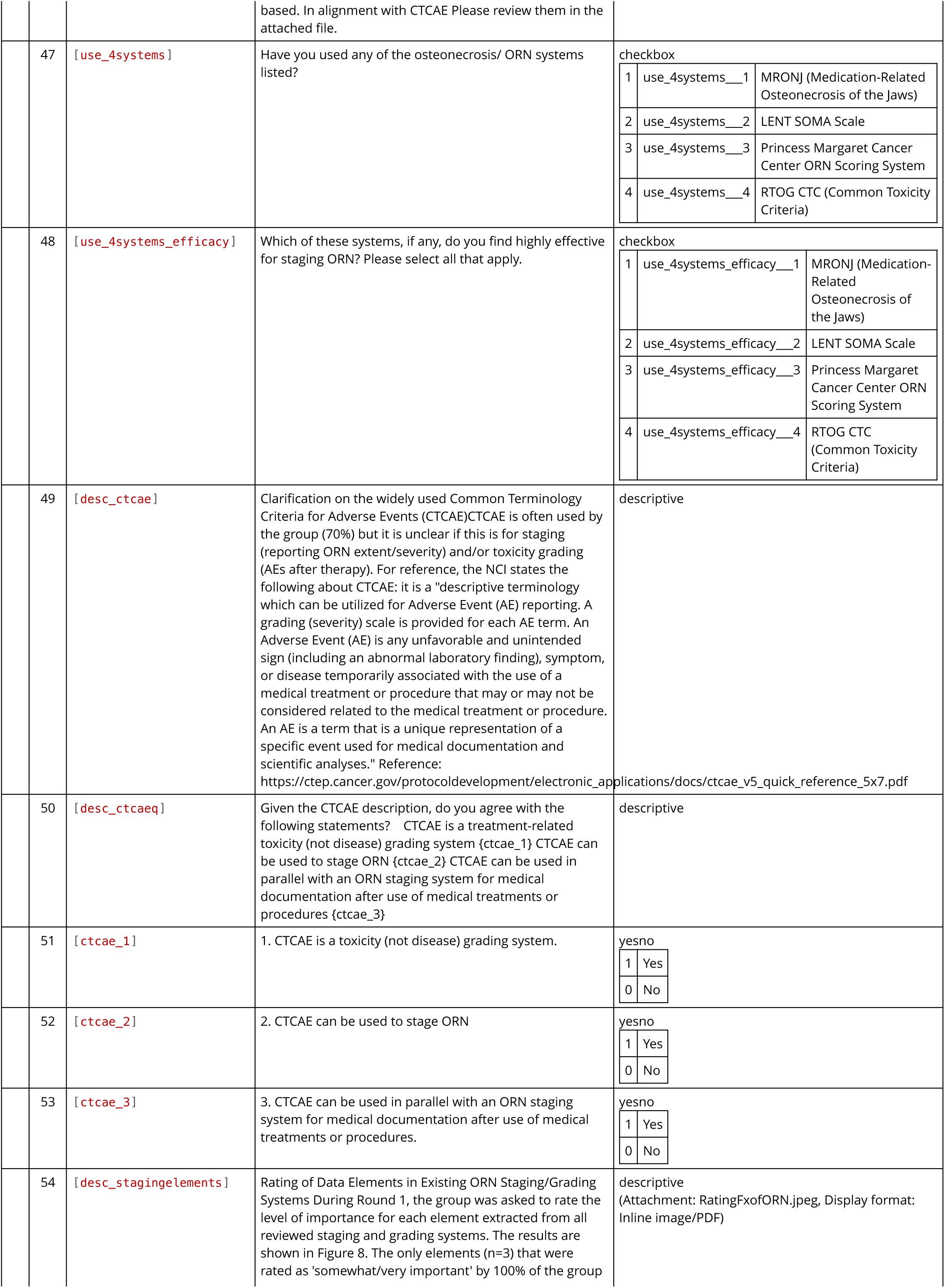
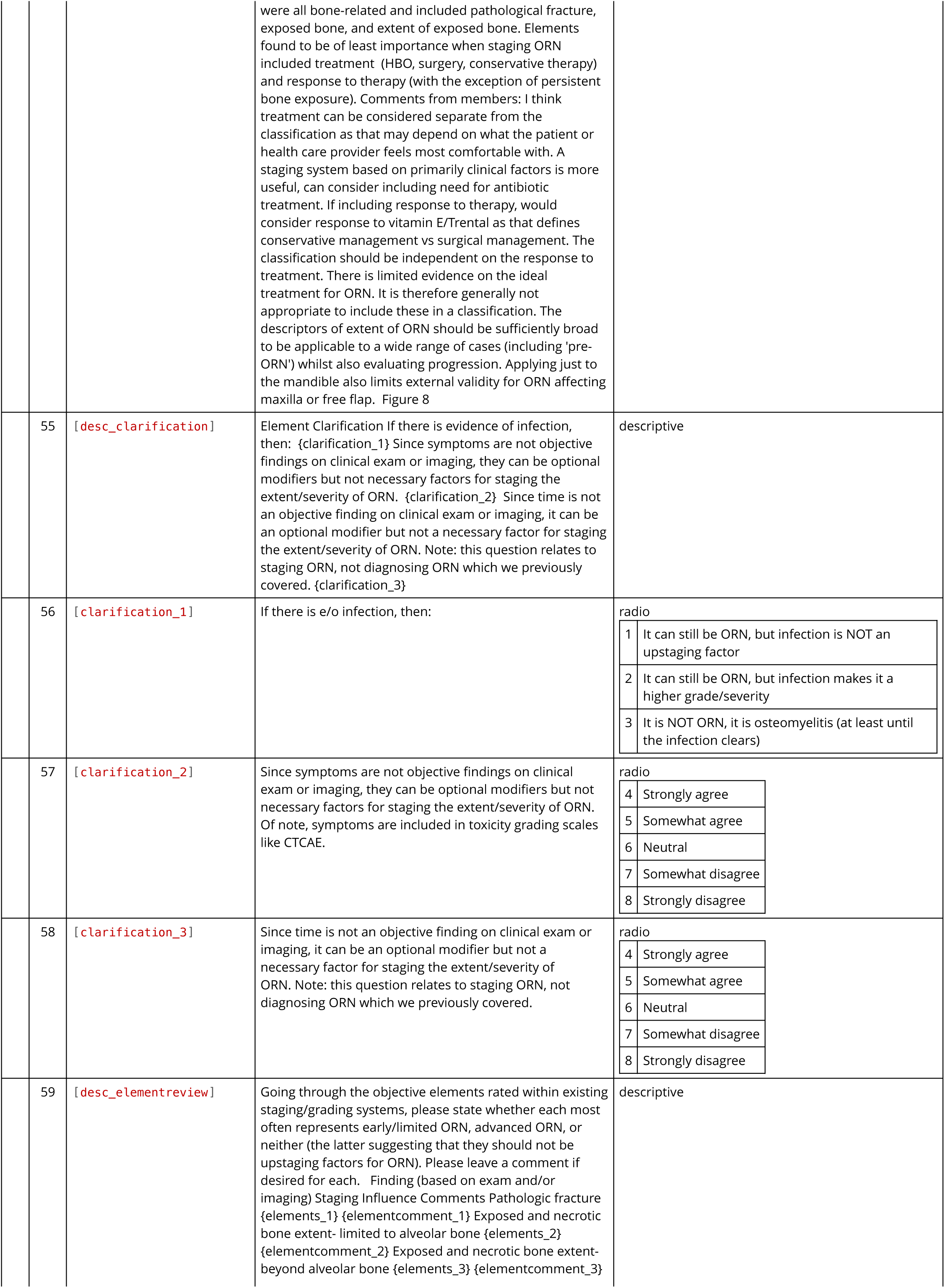
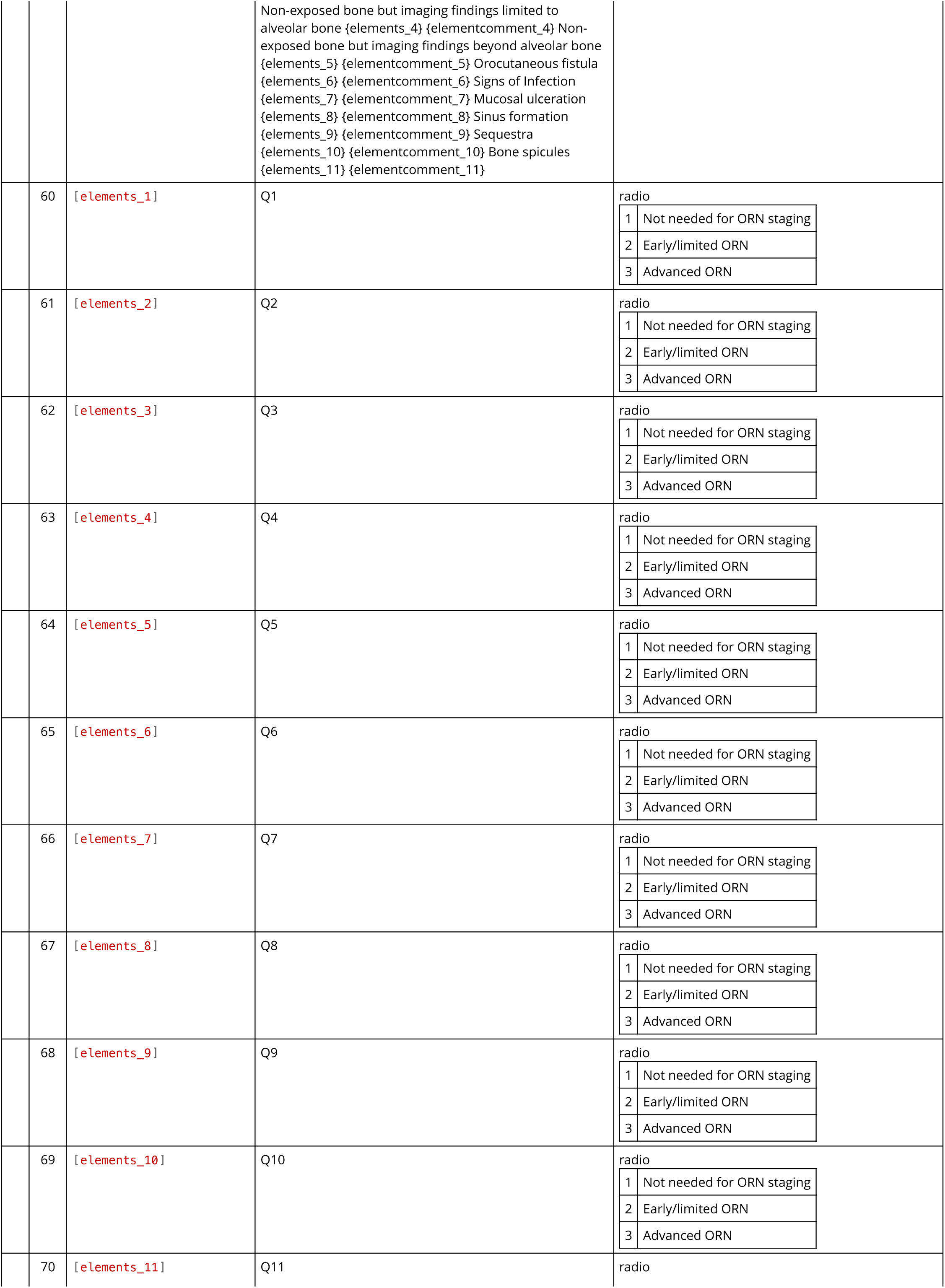
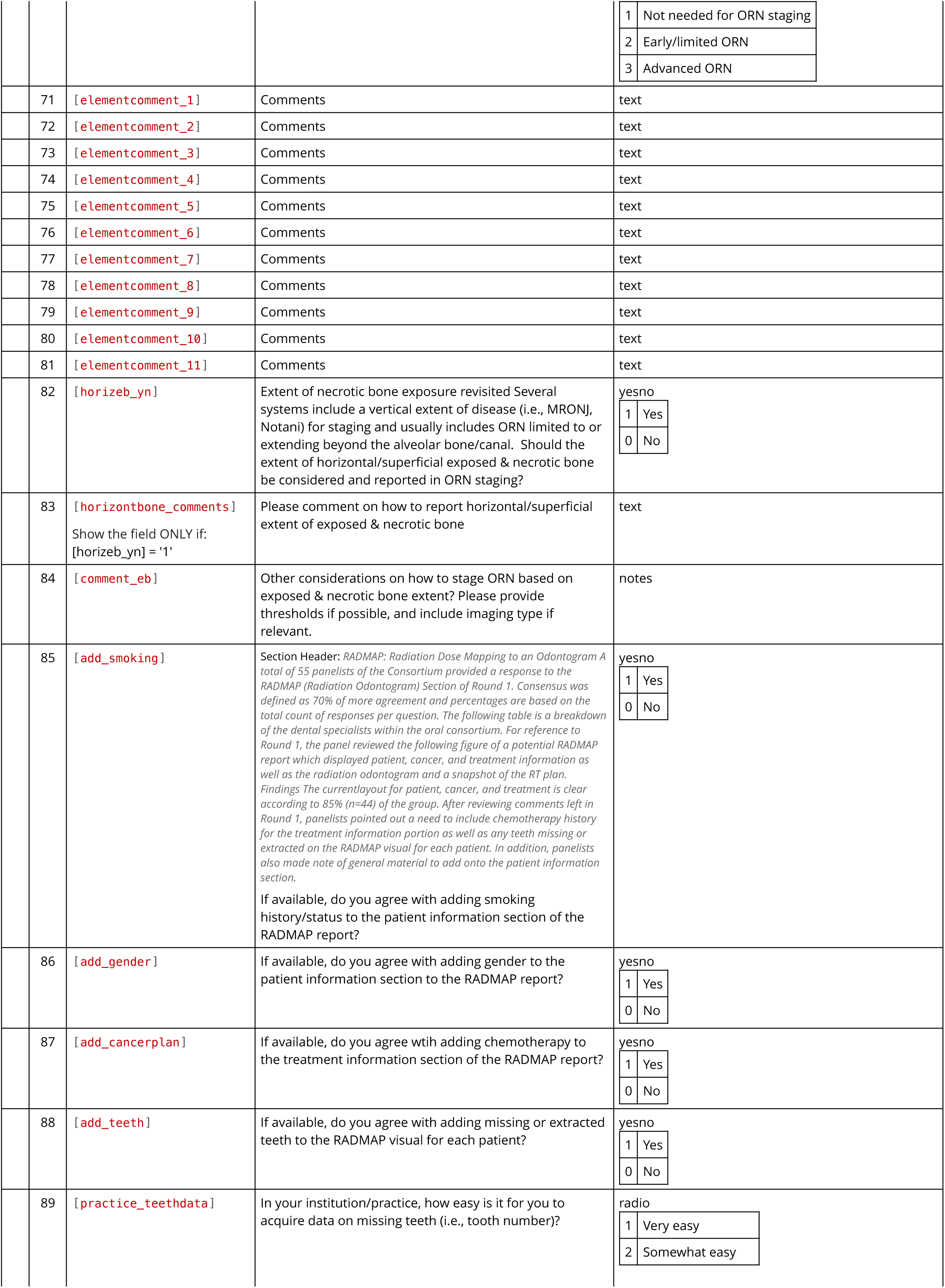
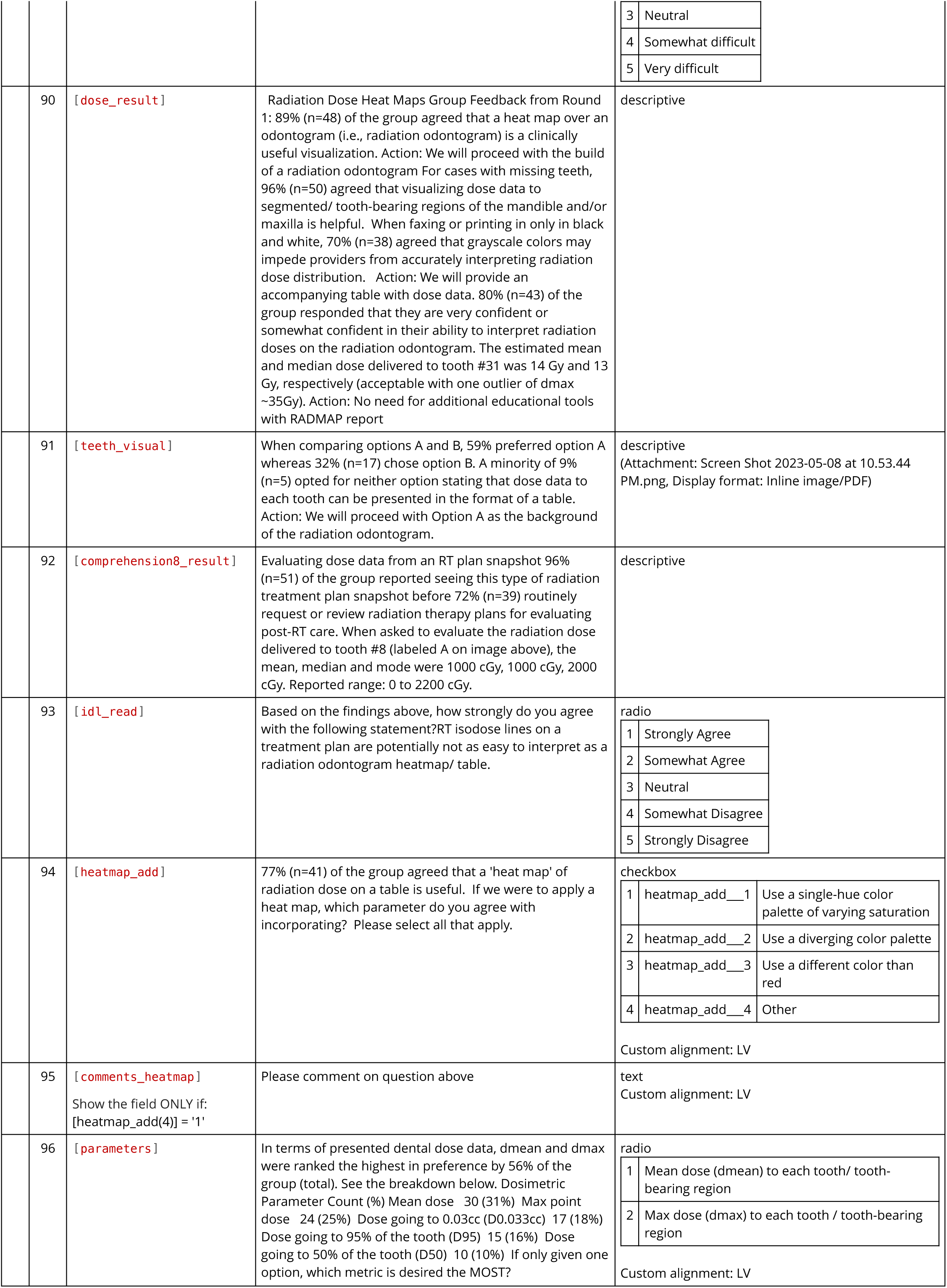
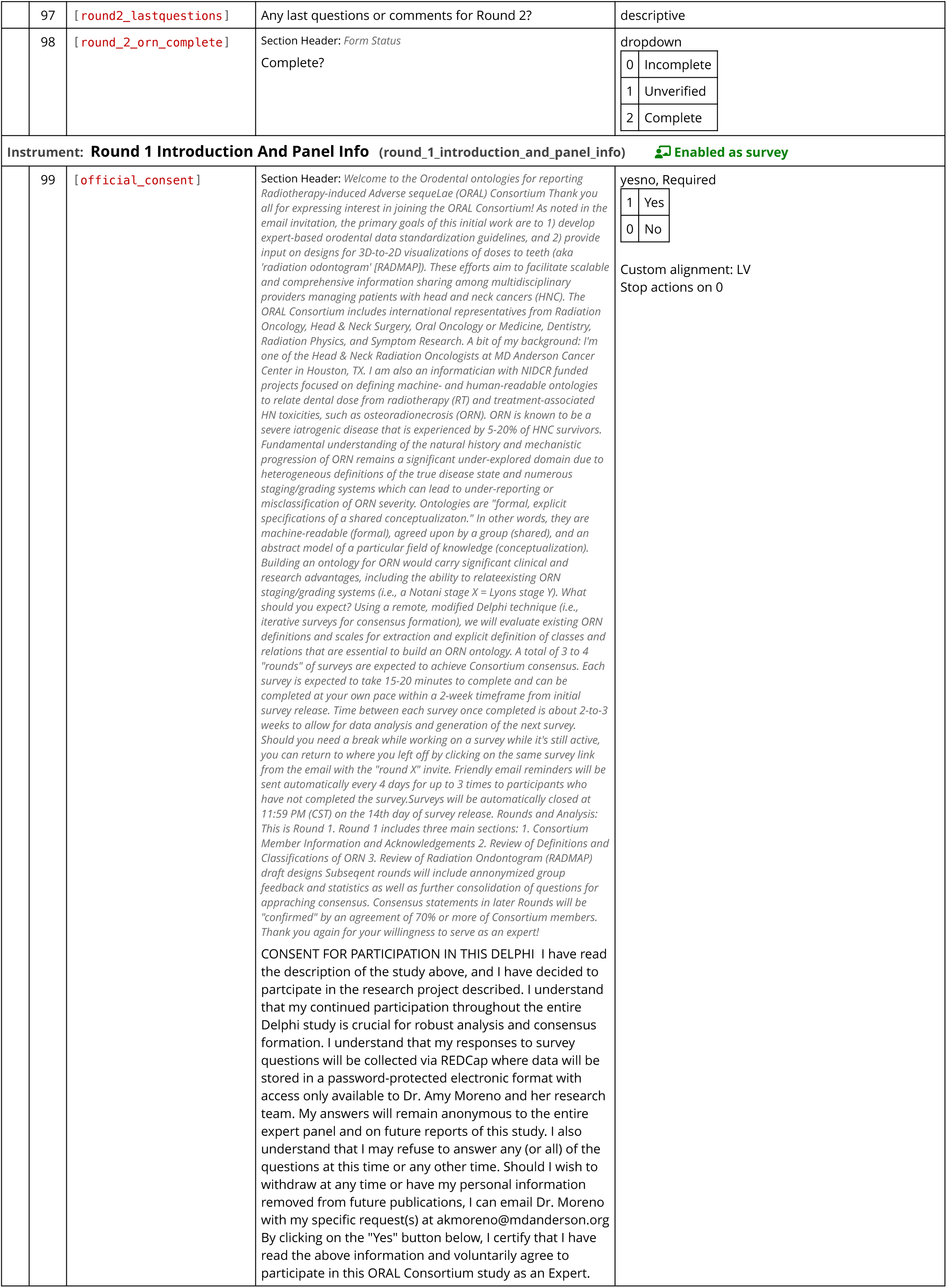
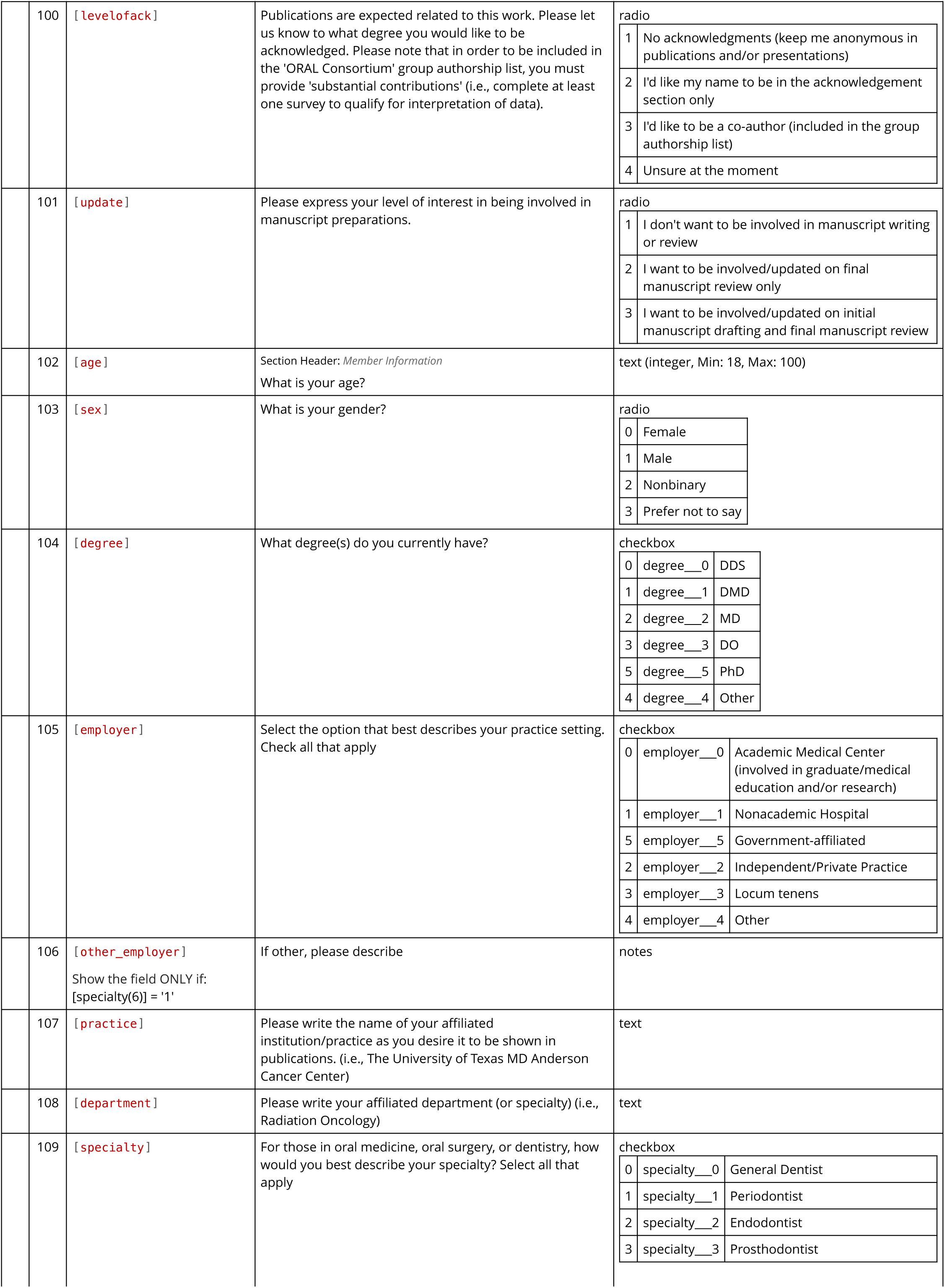
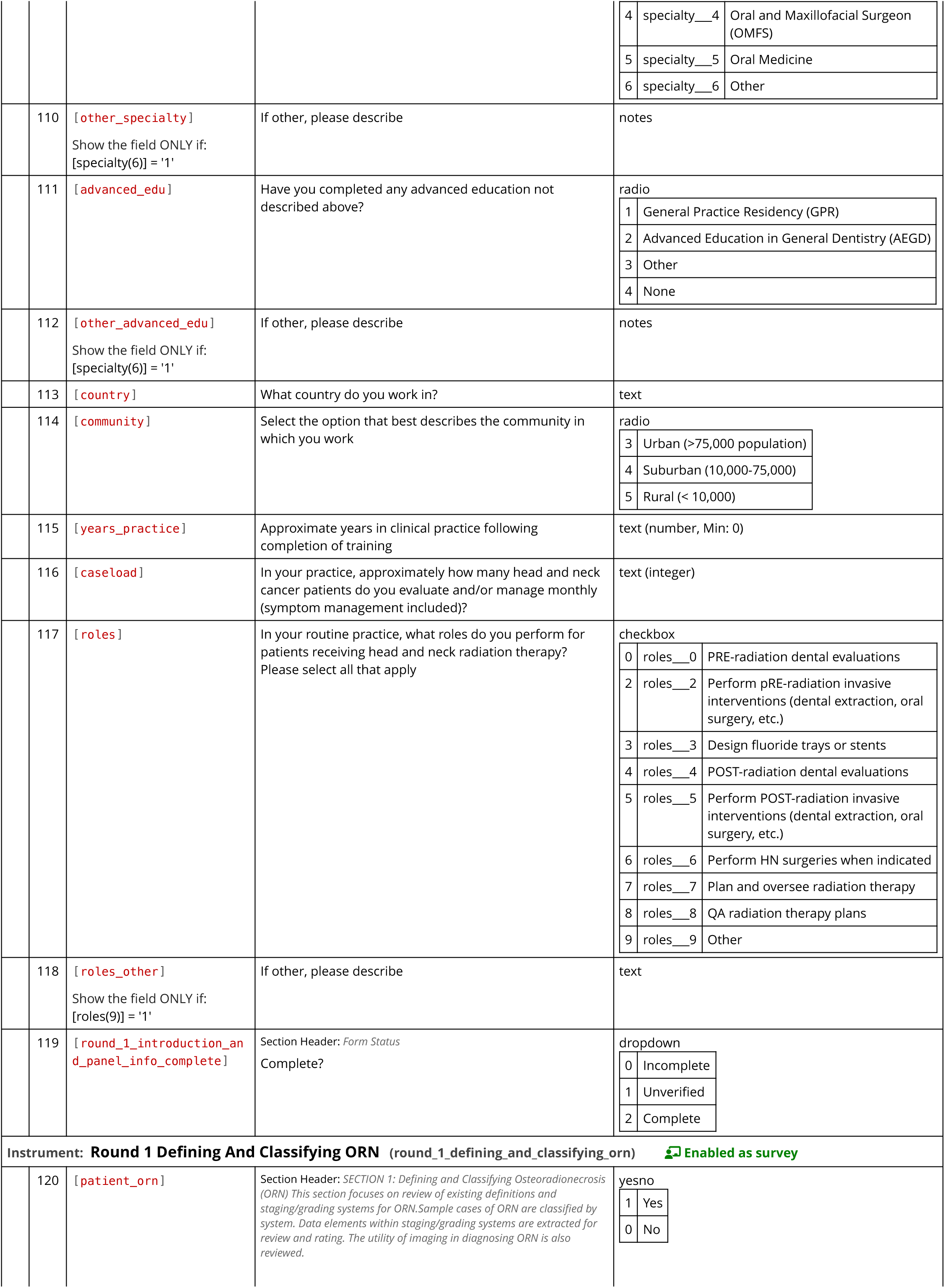
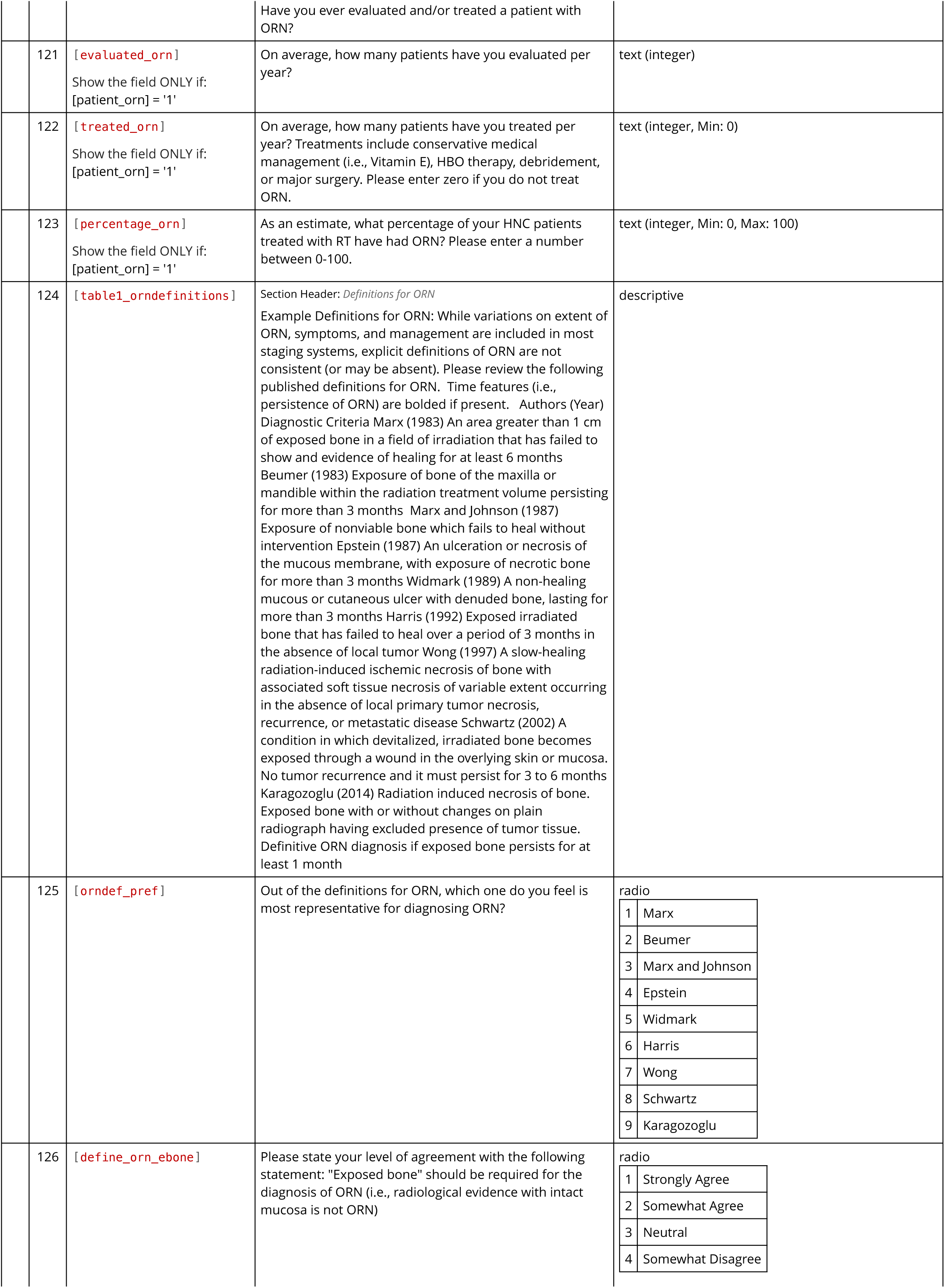
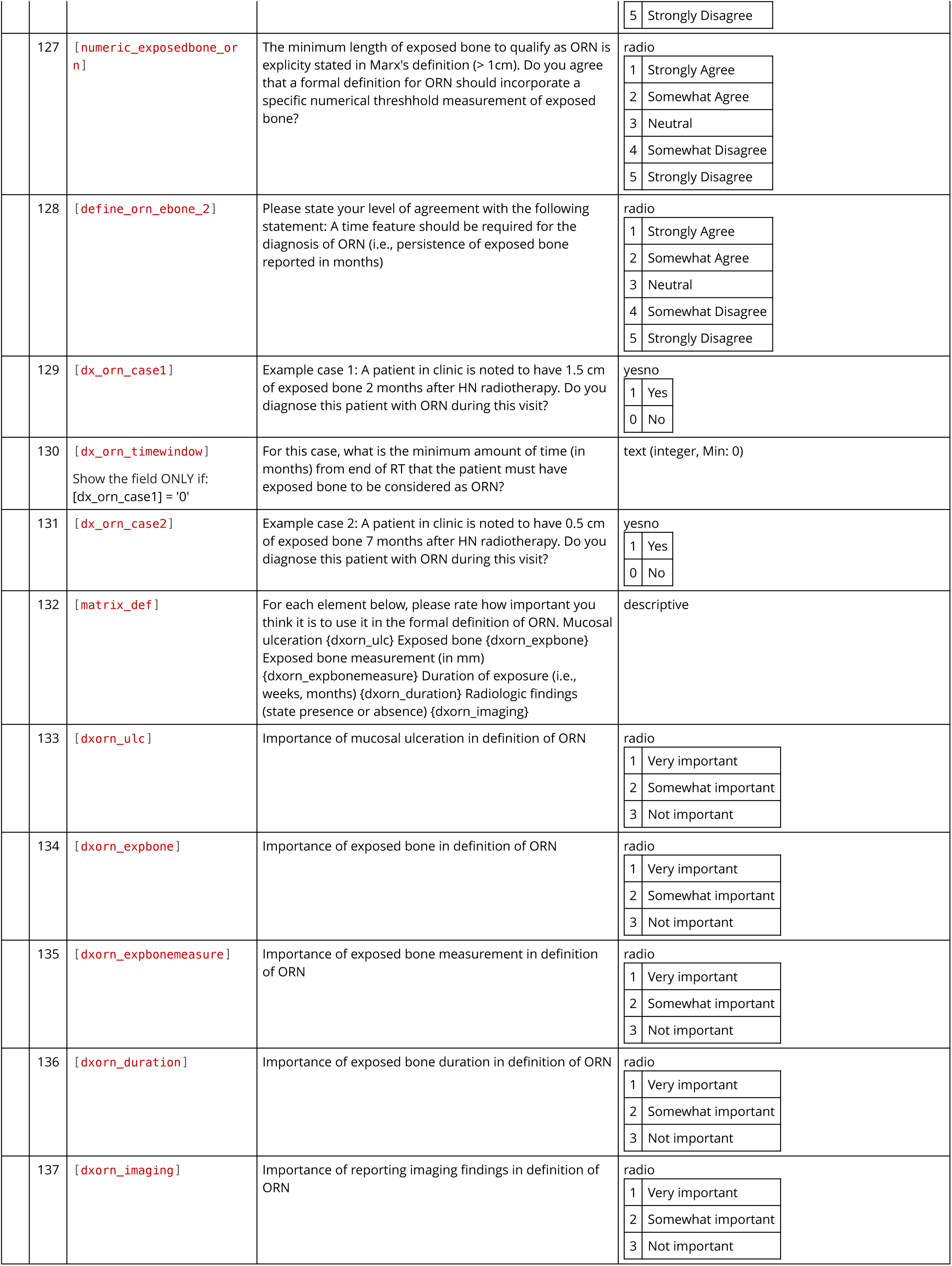
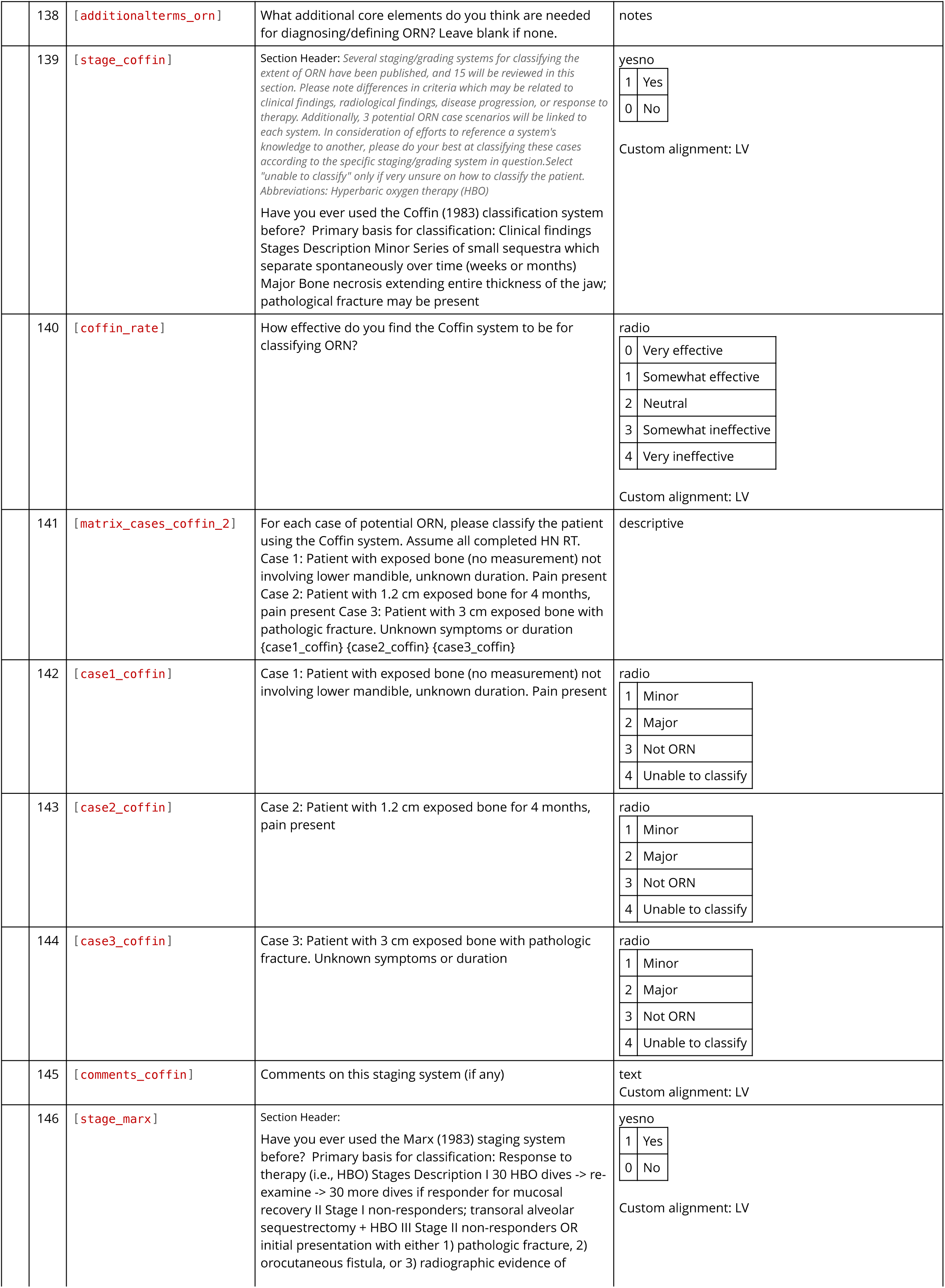
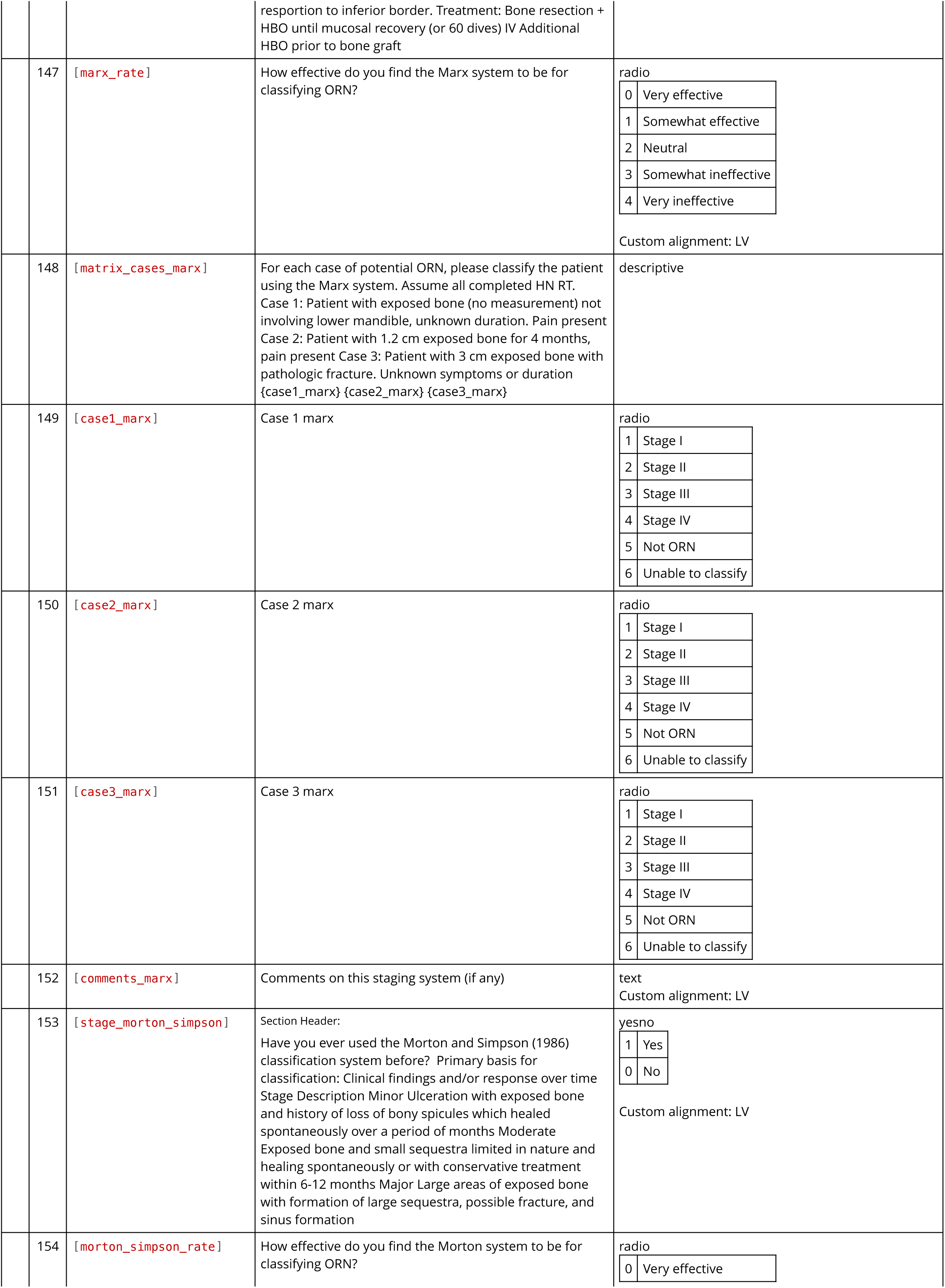
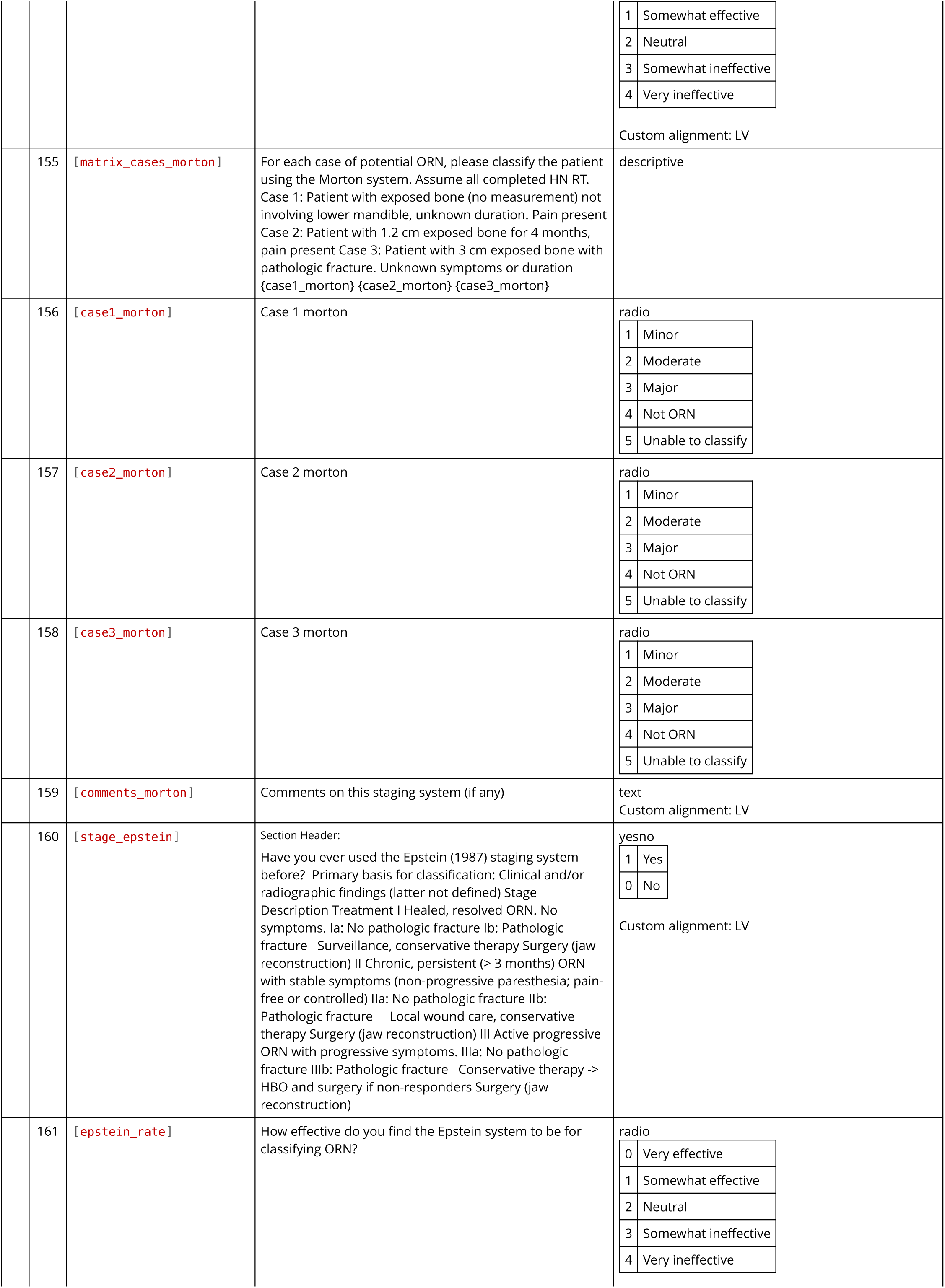
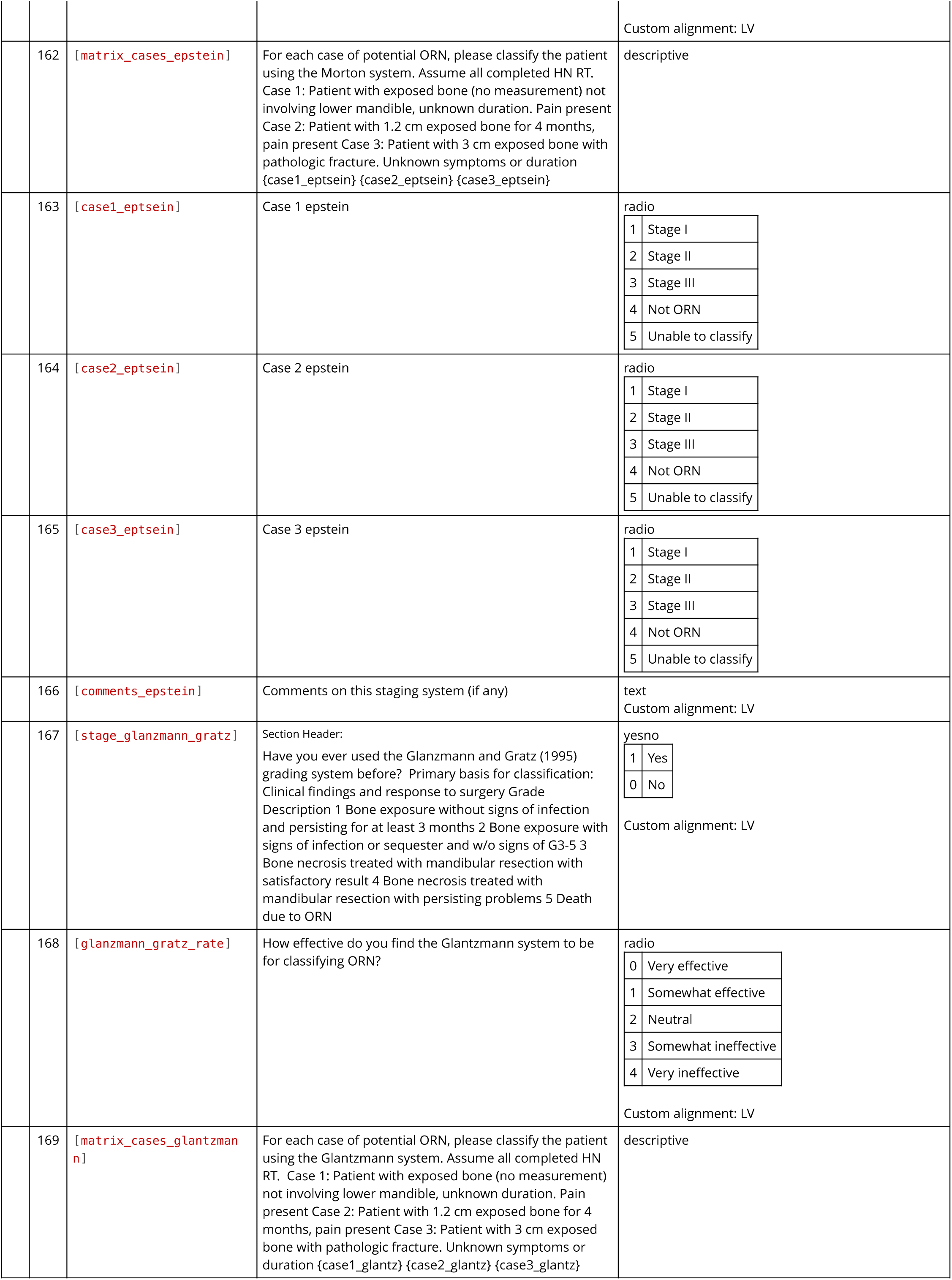
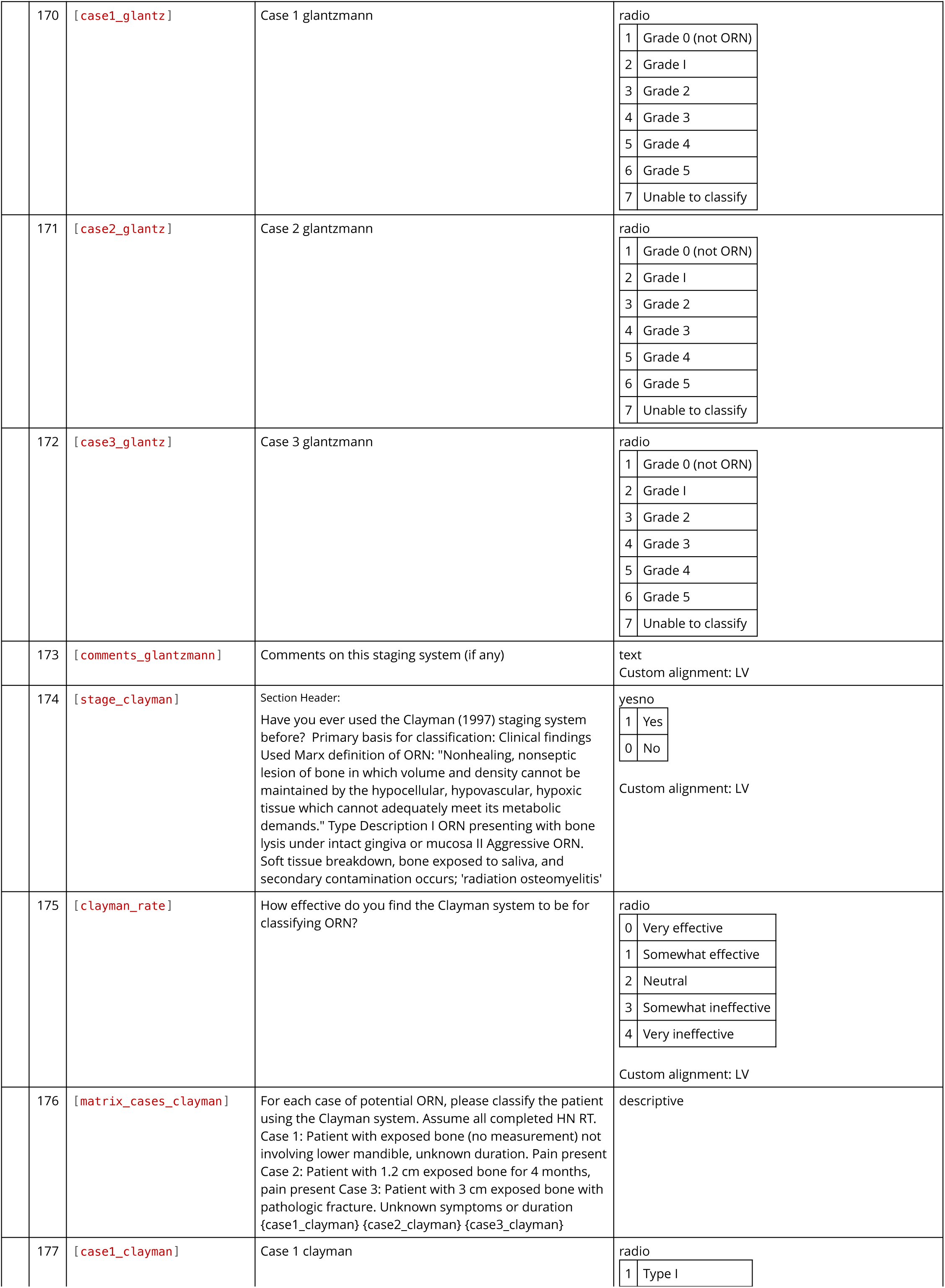
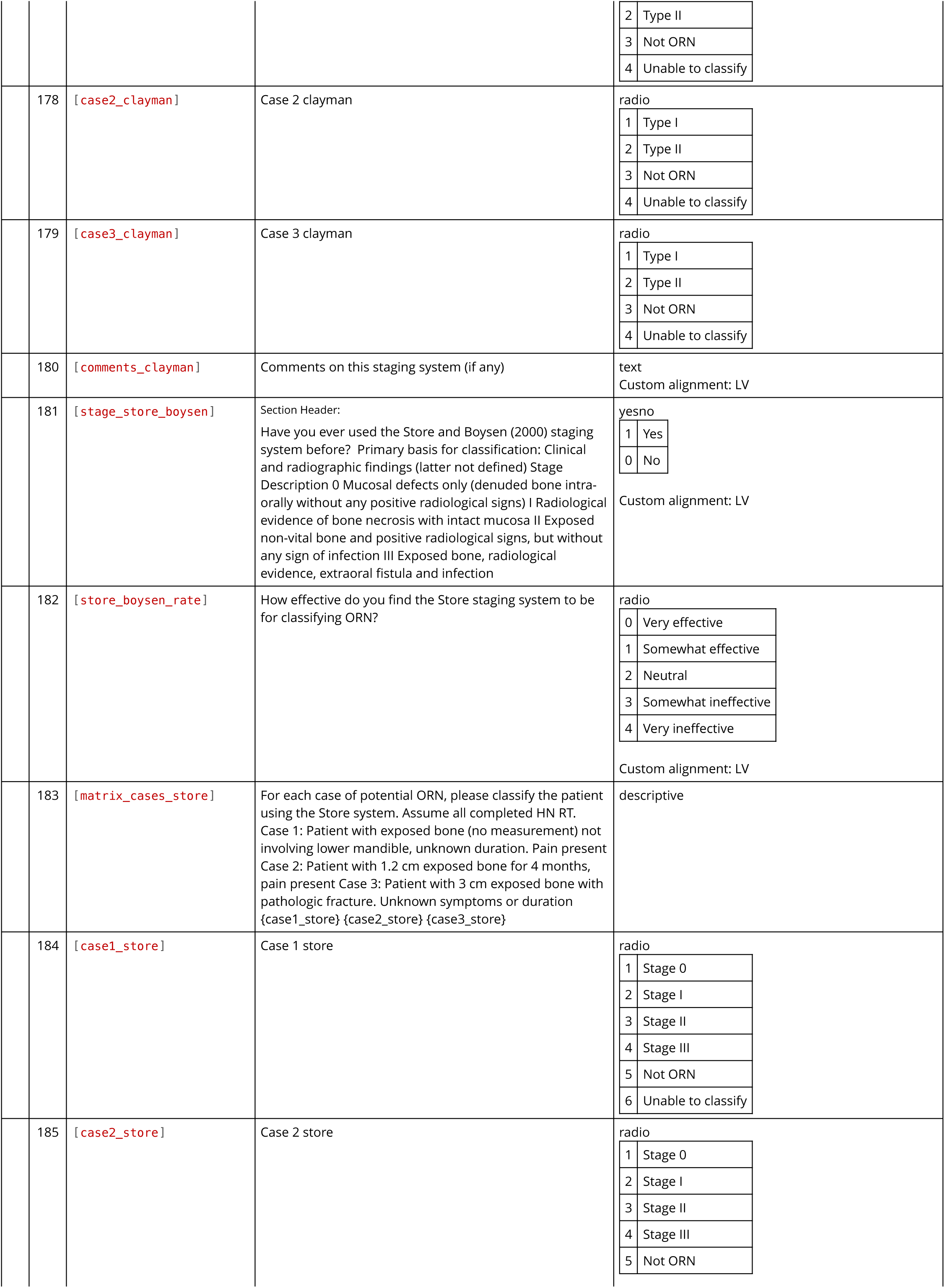
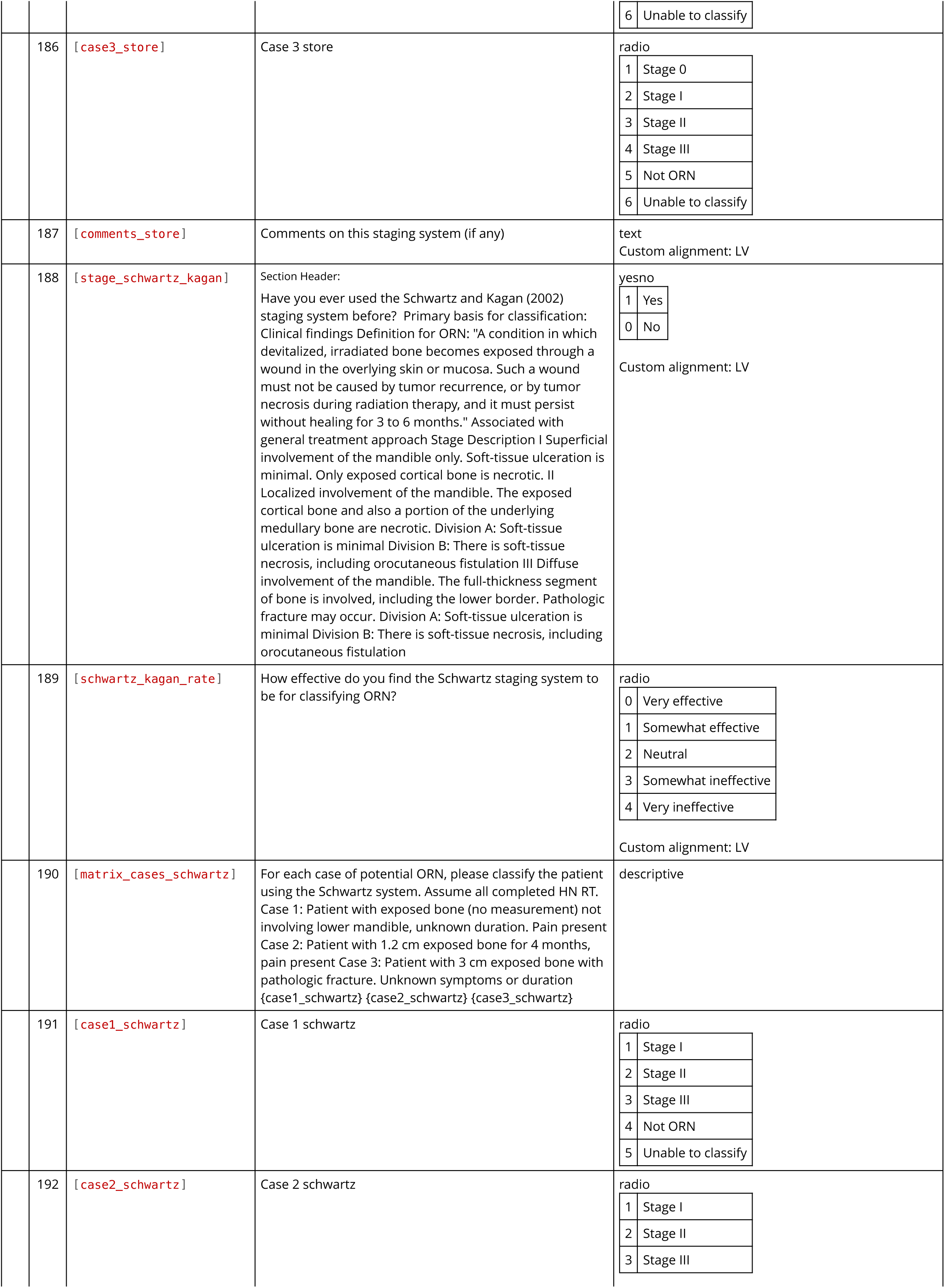
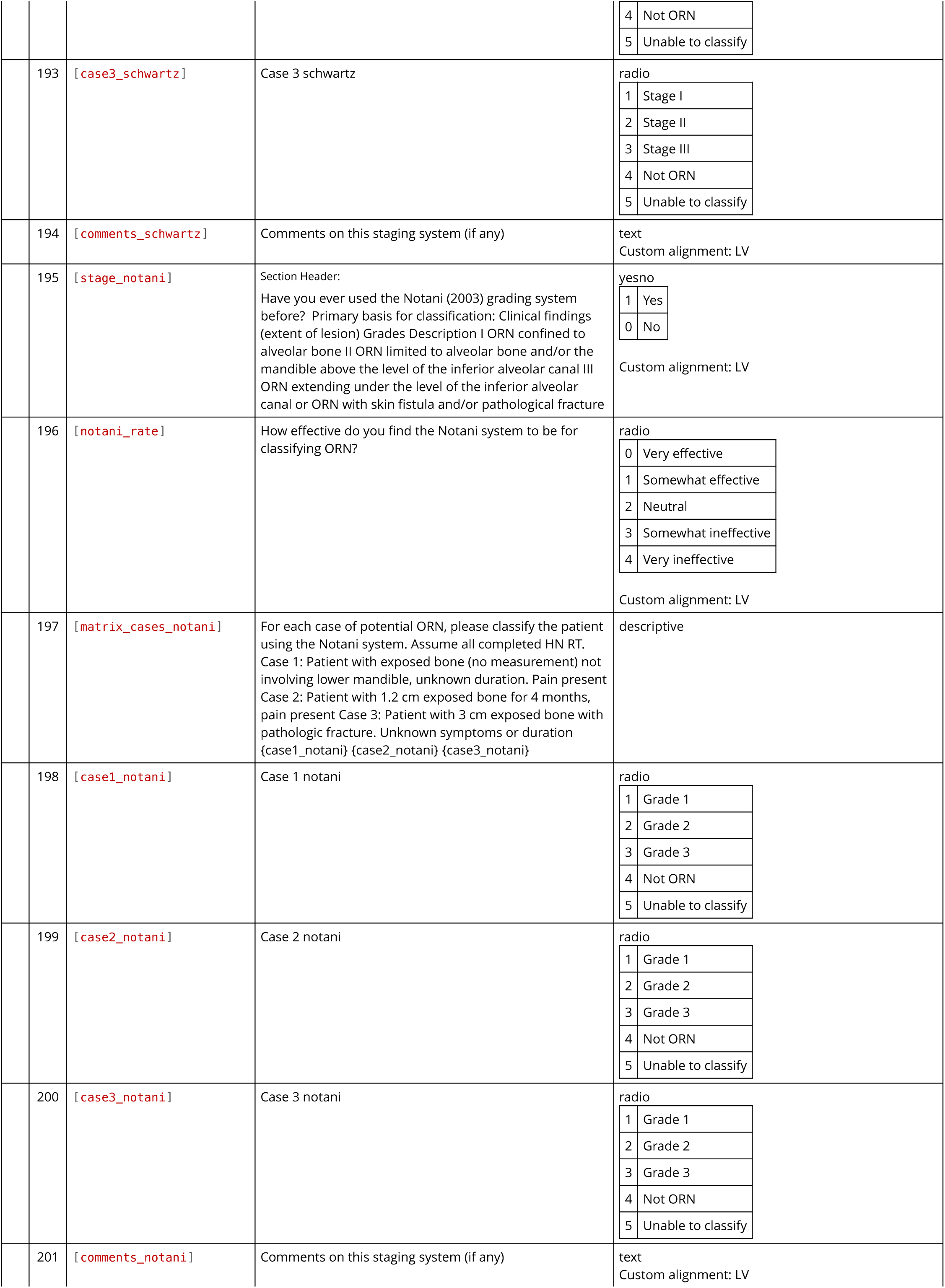
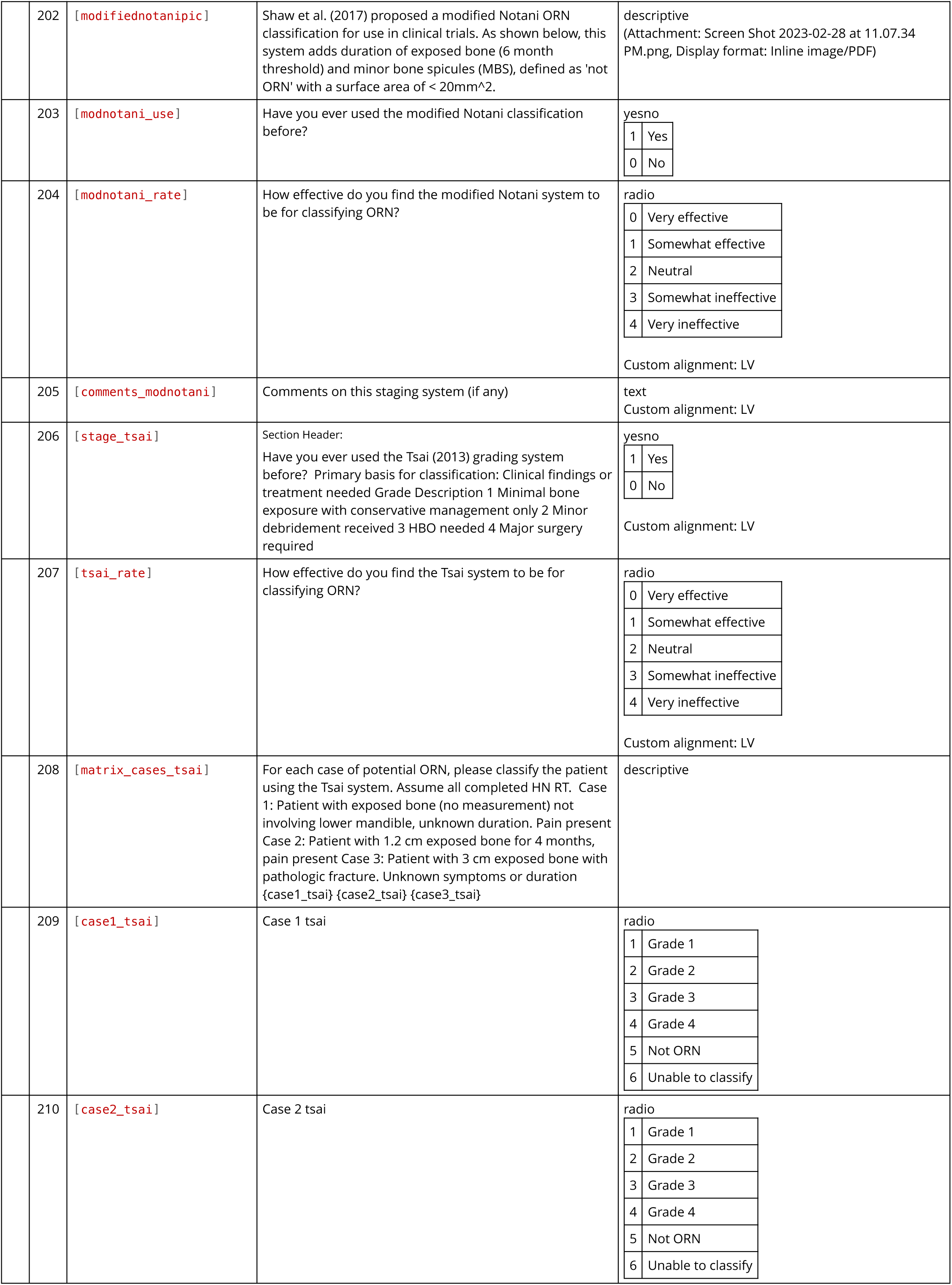
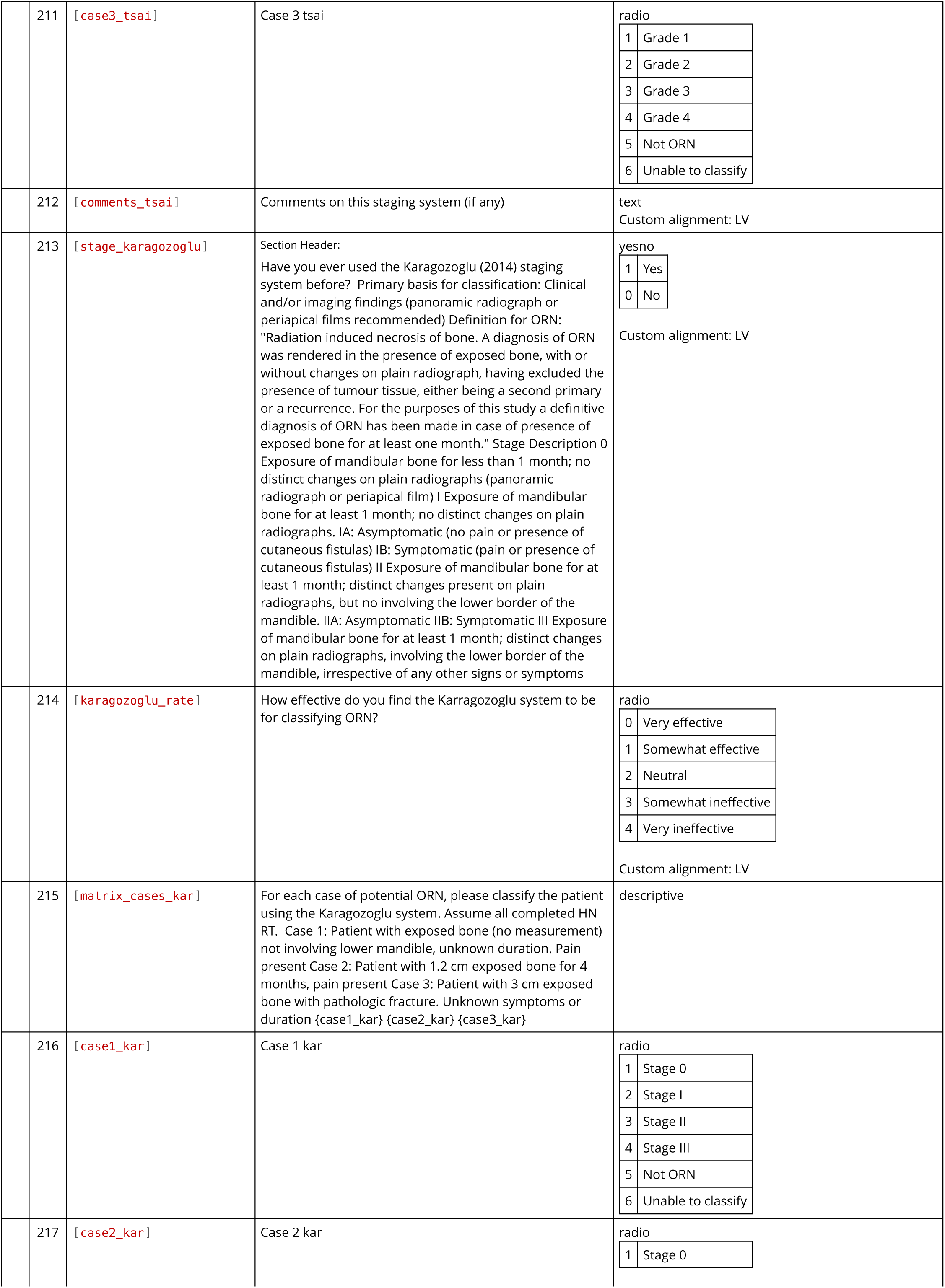
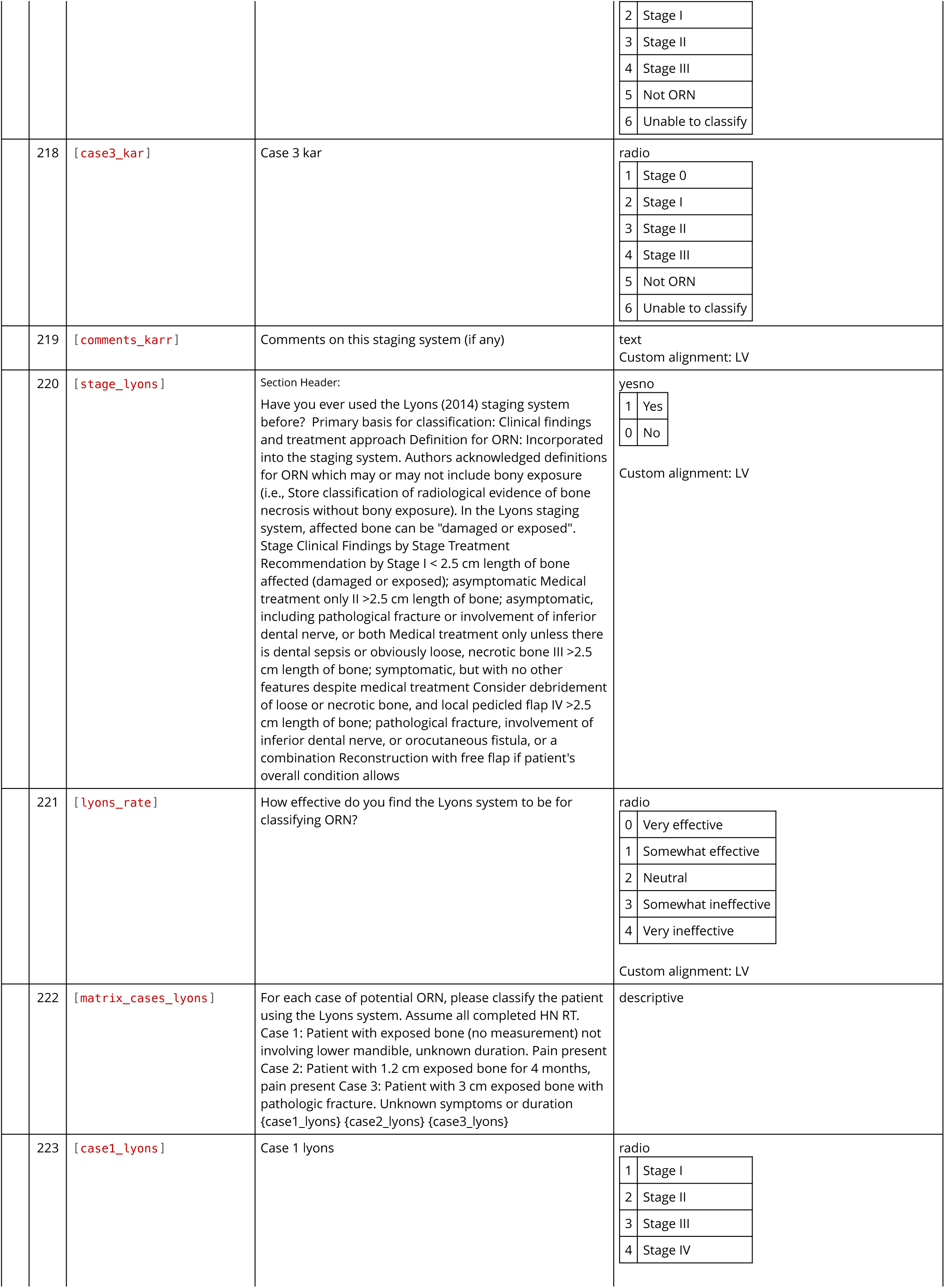
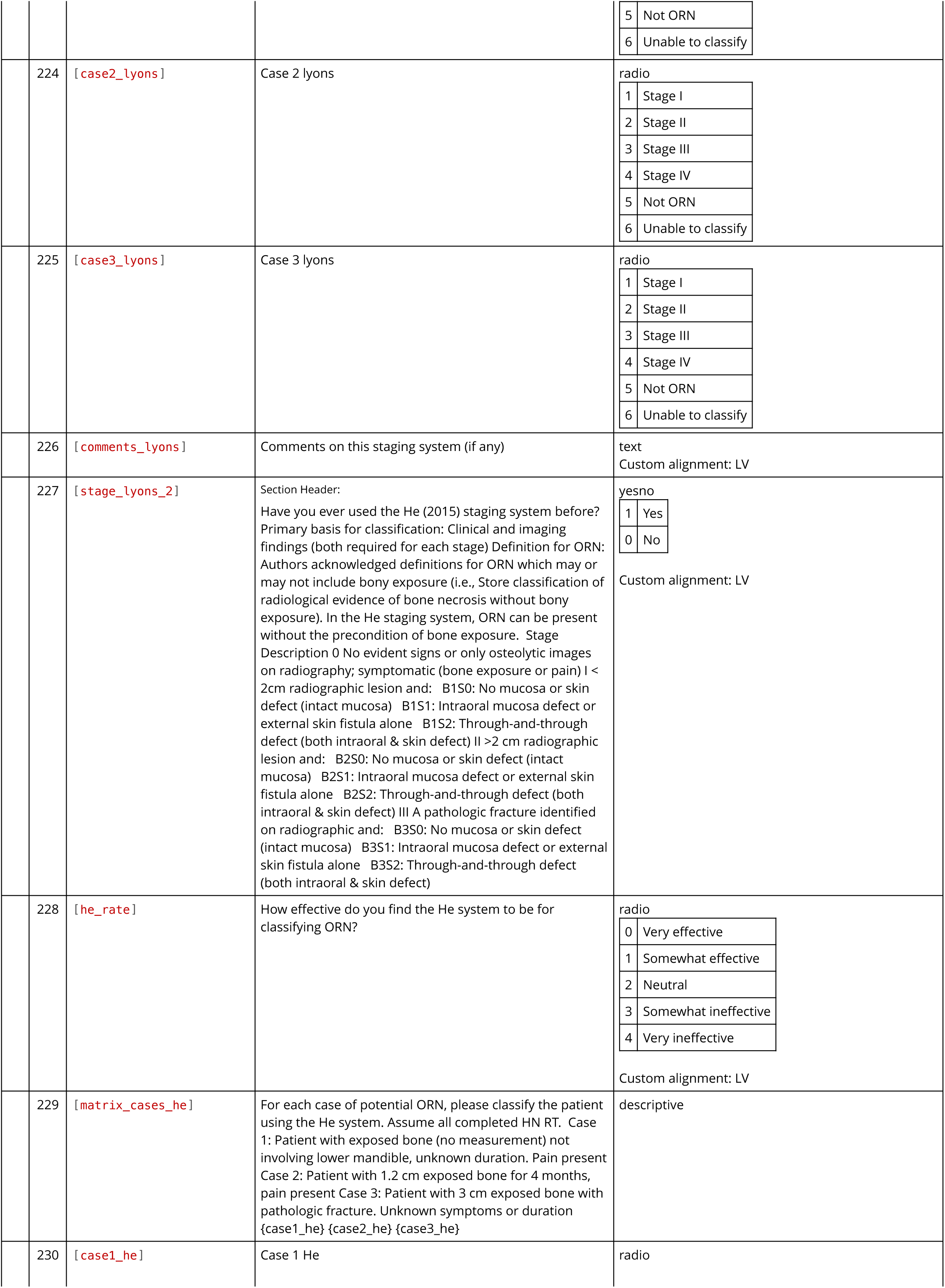
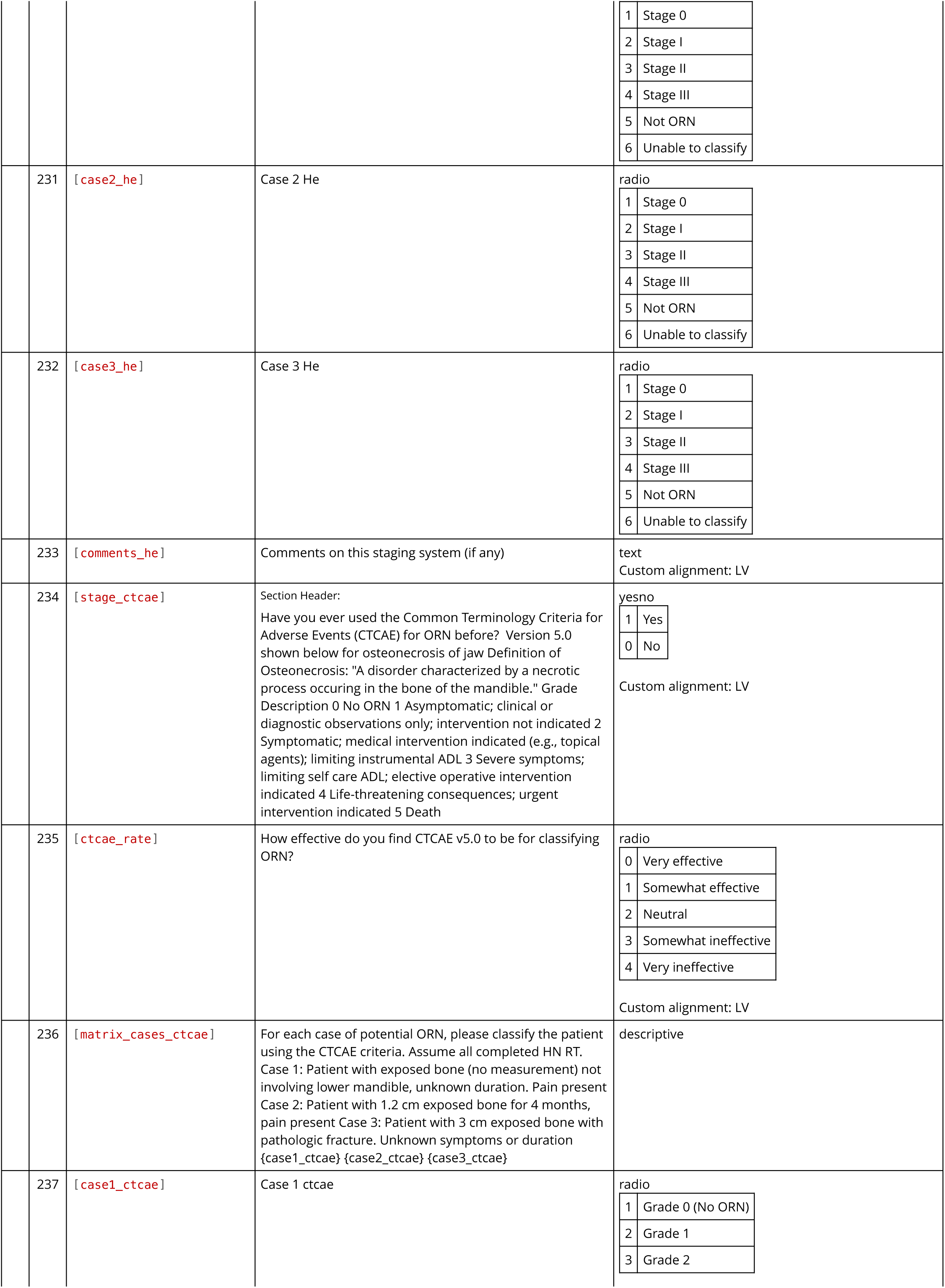
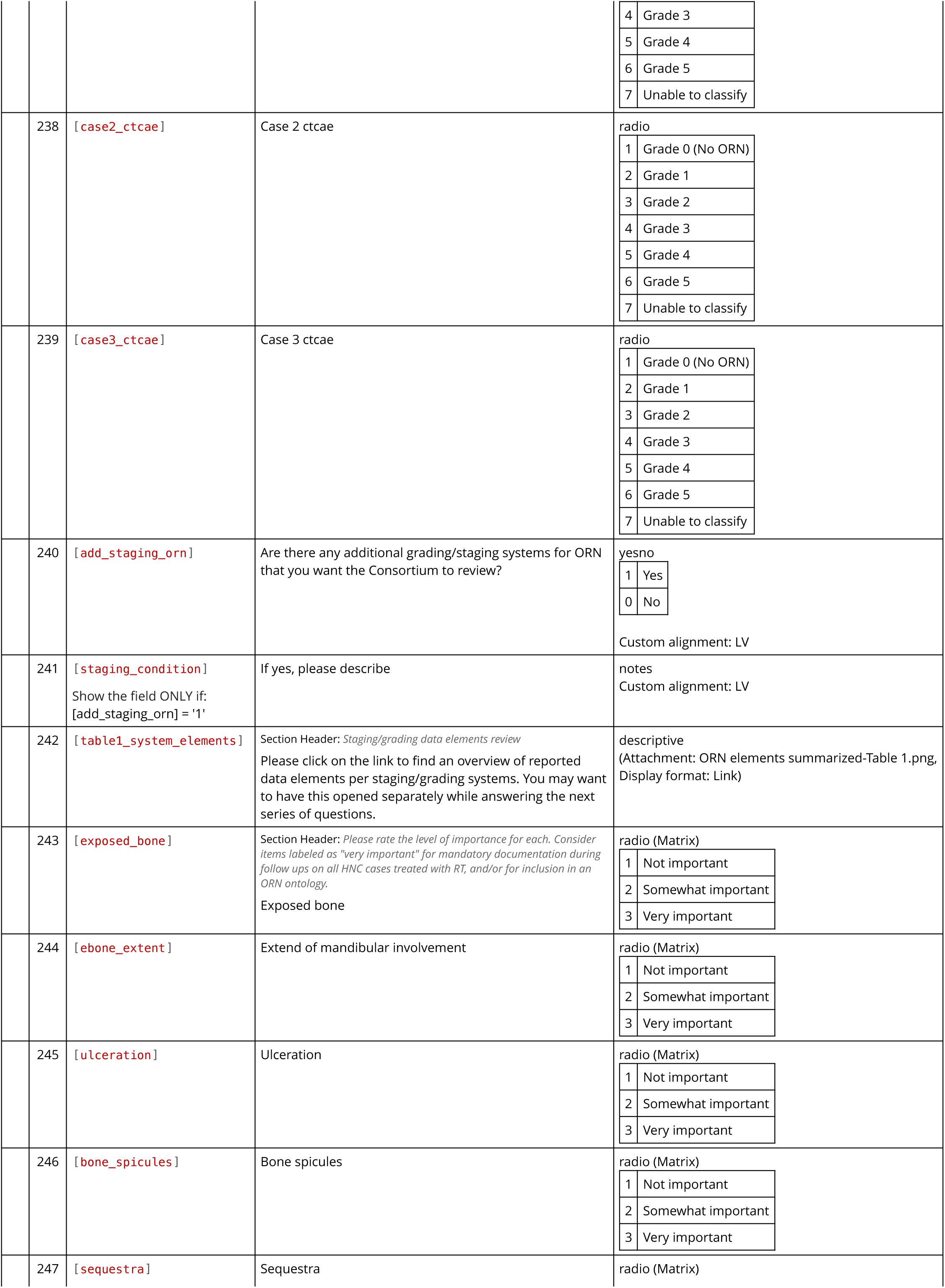
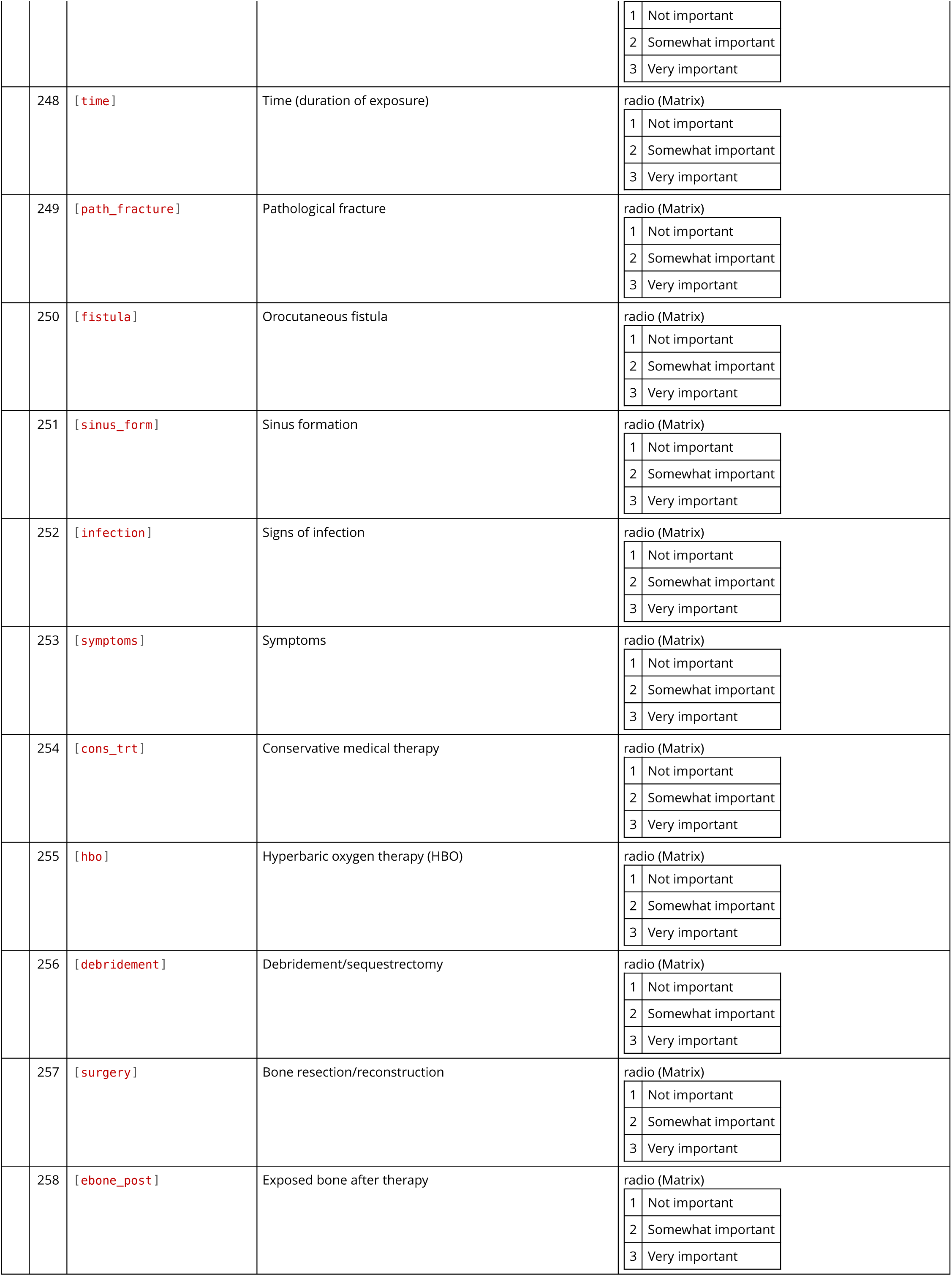
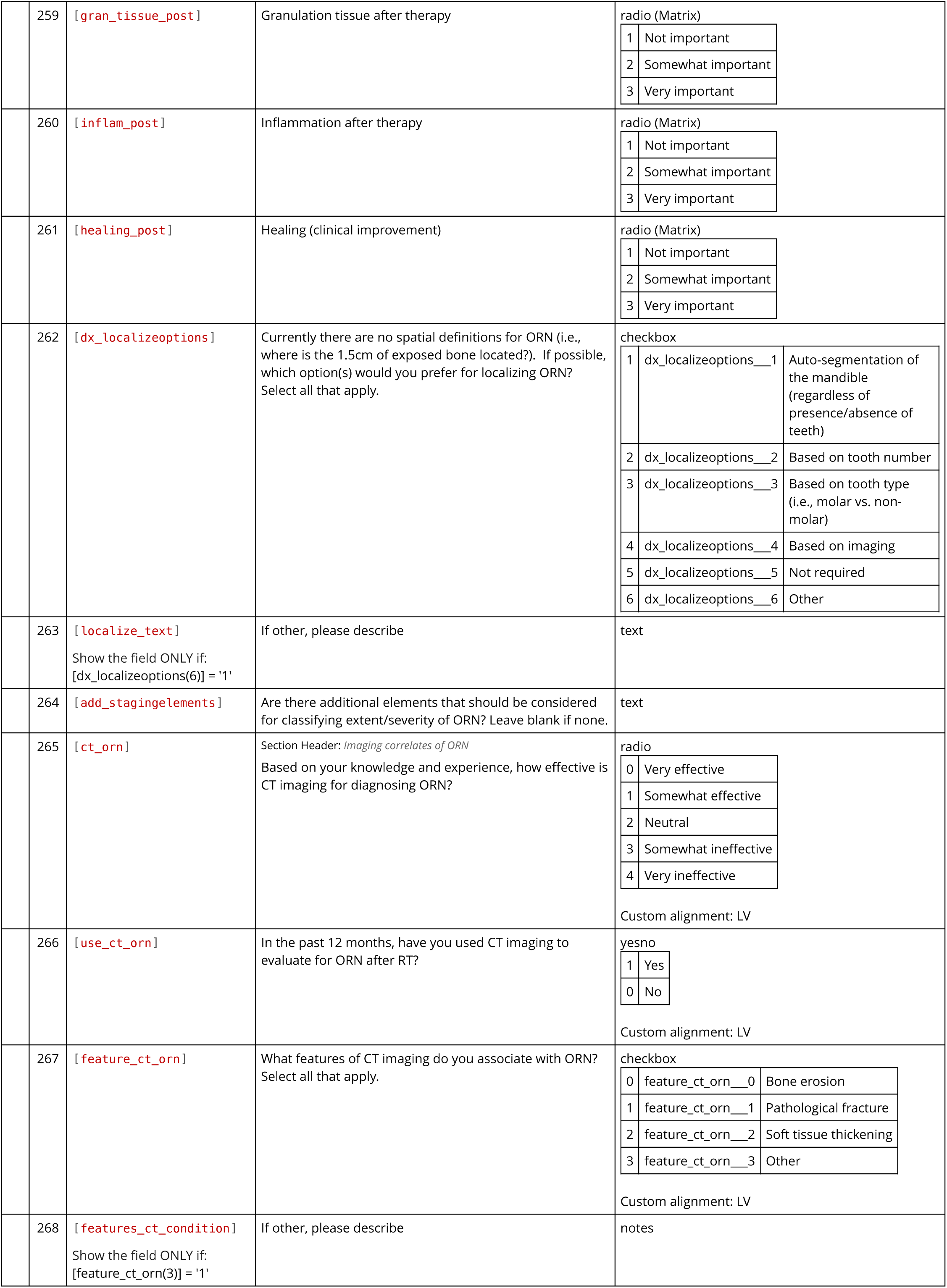
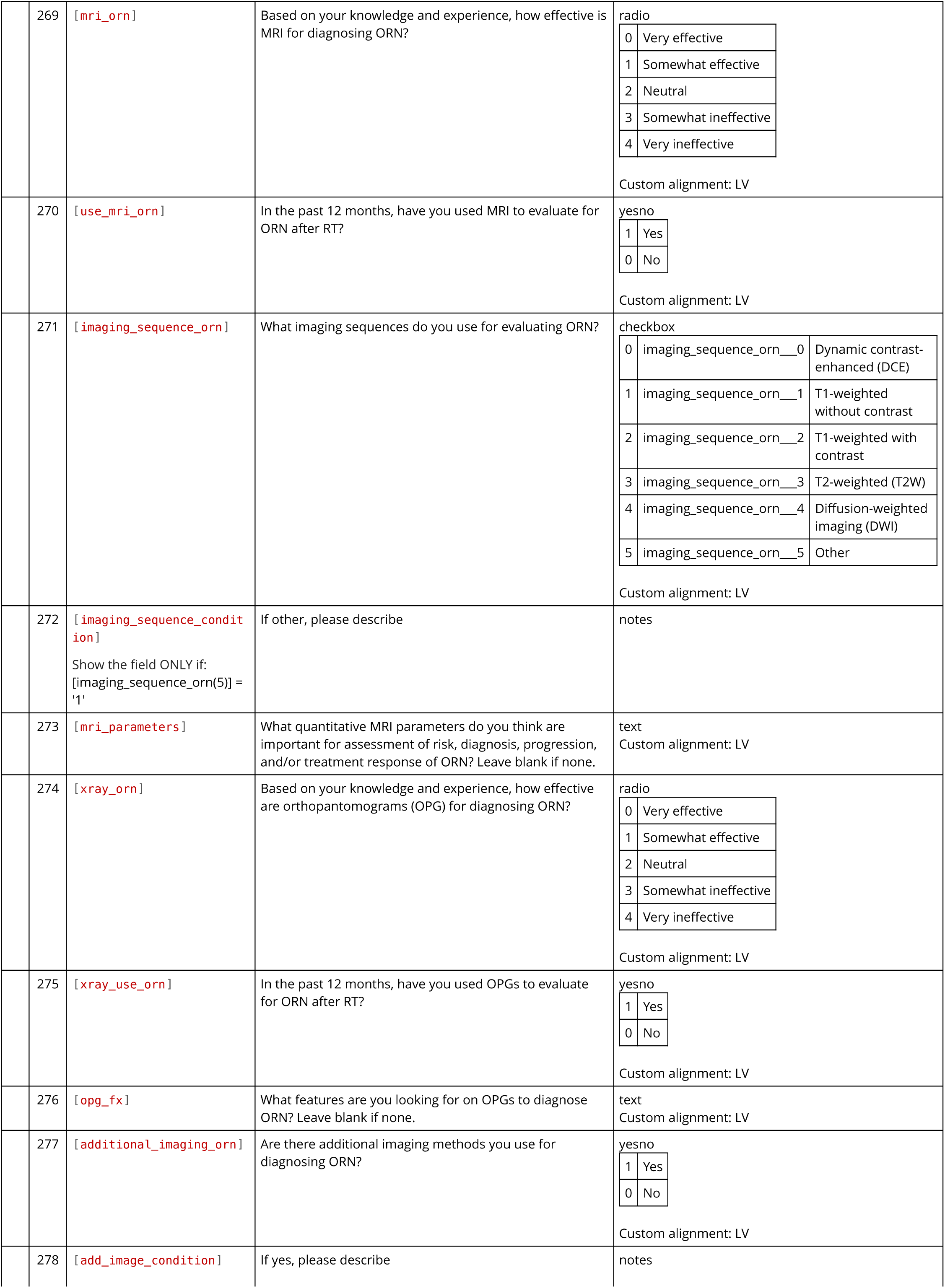
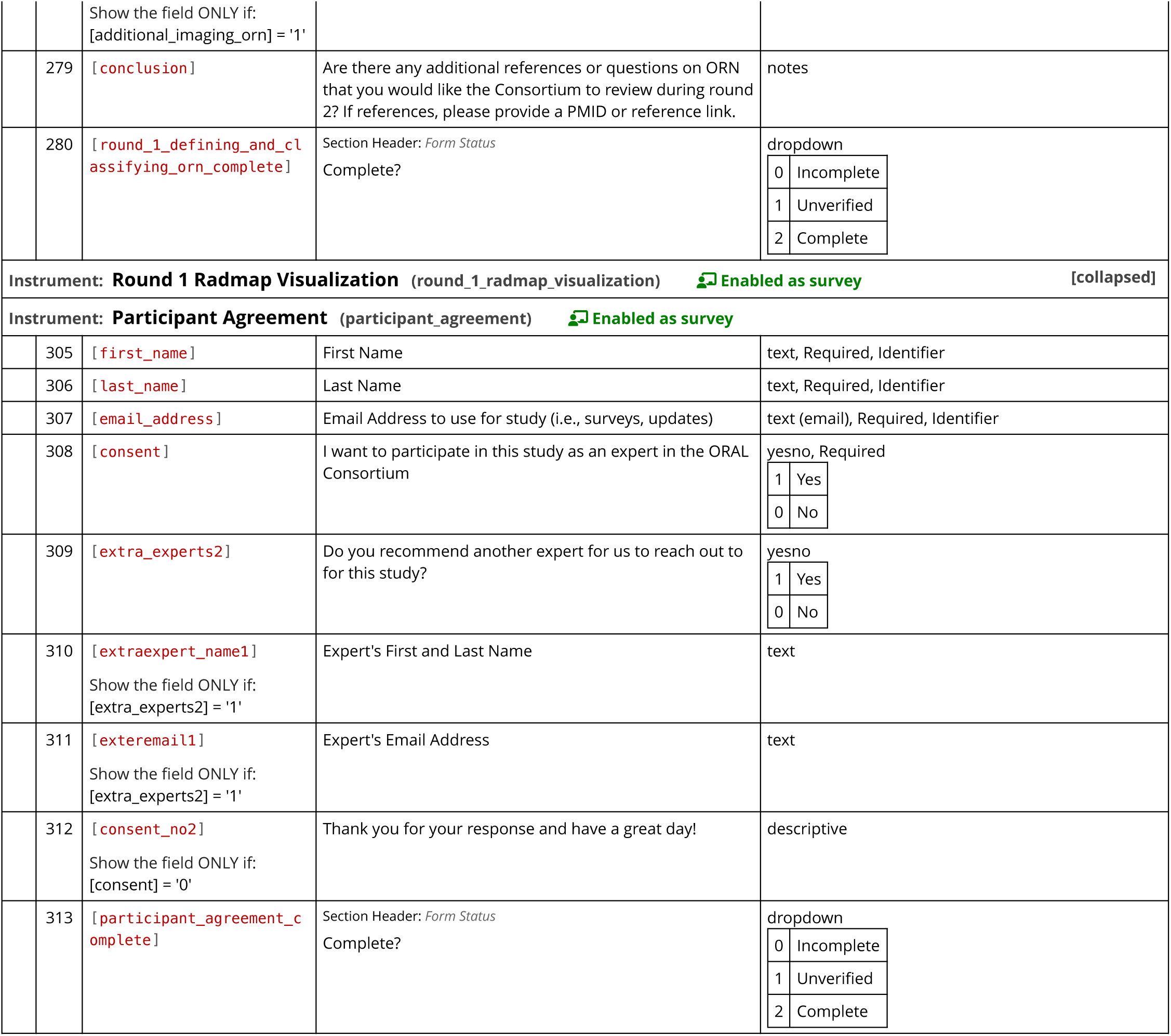

## Round 1: Introduction and Panel Info

Welcome to the

Orodental ontologies for reporting Radiotherapy-induced Adverse sequeLae (ORAL) Consortium

Thank you all for expressing interest in joining the ORAL Consortium! As noted in the email invitation, the primary goals of this initial work are to 1) develop expert-based orodental data standardization guidelines, and 2) provide input on designs for 3D-to-2D visualizations of doses to teeth (aka ‘radiation odontogram’ [RADMAP]). These efforts aim to facilitate scalable and comprehensive information sharing among multidisciplinary providers managing patients with head and neck cancers (HNC).

The ORAL Consortium includes international representatives from Radiation Oncology, Head & Neck Surgery, Oral Oncology or Medicine, Dentistry, Radiation Physics, and Symptom Research. A bit of my background: I’m one of the Head & Neck Radiation Oncologists at MD Anderson Cancer Center in Houston, TX. I am also an informatician with NIDCR funded projects focused on defining machine- and human-readable ontologies to relate dental dose from radiotherapy (RT) and treatment-associated HN toxicities, such as osteoradionecrosis (ORN).

ORN is known to be a severe iatrogenic disease that is experienced by 5-20% of HNC survivors. Fundamental understanding of the natural history and mechanistic progression of ORN remains a significant under-explored domain due to heterogeneous definitions of the true disease state and numerous staging/grading systems which can lead to under-reporting or misclassification of ORN severity.

Ontologies are “formal, explicit specifications of a shared conceptualizaton.” In other words, they are machine-readable (formal), agreed upon by a group (shared), and an abstract model of a particular field of knowledge (conceptualization). Building an ontology for ORN would carry significant clinical and research advantages, including the ability to relate existing ORN staging/grading systems (i.e., a Notani stage X = Lyons stage Y).

What should you expect?

Using a remote, modified Delphi technique (i.e., iterative surveys for consensus formation), we will evaluate existing ORN definitions and scales for extraction and explicit definition of classes and relations that are essential to build an ORN ontology. A total of 3 to 4 “rounds” of surveys are expected to achieve Consortium consensus. Each survey is expected to take 15-20 minutes to complete and can be completed at your own pace within a 2-week timeframe from initial survey release. Time between each survey once completed is about 2-to-3 weeks to allow for data analysis and generation of the next survey.

Should you need a break while working on a survey while it’s still active, you can return to where you left off by clicking on the same survey link from the email with the “round X” invite.

Friendly email reminders will be sent automatically every 4 days for up to 3 times to participants who have not completed the survey. Surveys will be automatically closed at 11:59 PM (CST) on the 14th day of survey release.

Rounds and Analysis:

This is Round 1. Round 1 includes three main sections:

1. Consortium Member Information and Acknowledgements
2. Review of Definitions and Classifications of ORN
3. Review of Radiation Ondontogram (RADMAP) draft designs

Subseqent rounds will include annonymized group feedback and statistics as well as further consolidation of questions for appraching consensus. Consensus statements in later Rounds will be “confirmed” by an agreement of 70% or more of Consortium members.

Thank you again for your willingness to serve as an expert!

### CONSENT FOR PARTICIPATION IN THIS DELPHI

I have read the description of the study above, and I have decided to partcipate in the research project described. I understand that my continued participation throughout the entire Delphi study is crucial for robust analysis and consensus formation. I understand that my responses to survey questions will be collected via REDCap where data will be stored in a password-protected electronic format with access only available to Dr. Amy Moreno and her research team. My answers will remain anonymous to the entire expert panel and on future reports of this study. I also understand that I may refuse to answer any (or all) of the questions at this time or any other time. Should I wish to withdraw at any time or have my personal information removed from future publications, I can email Dr. Moreno with my specific request(s) at

By clicking on the “Yes” button below, I certify that I have read the above information and voluntarily agree to participate in this ORAL Consortium study as an Expert.

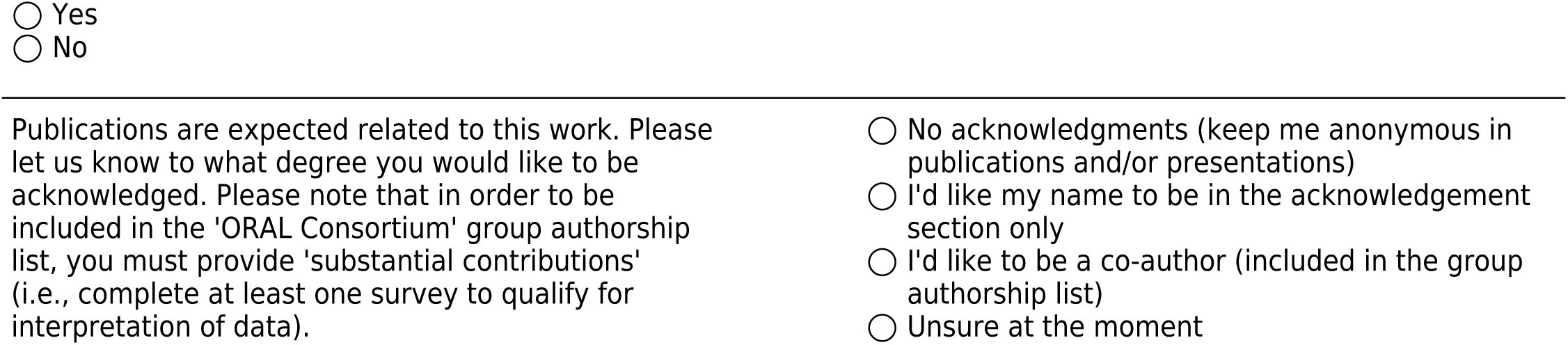
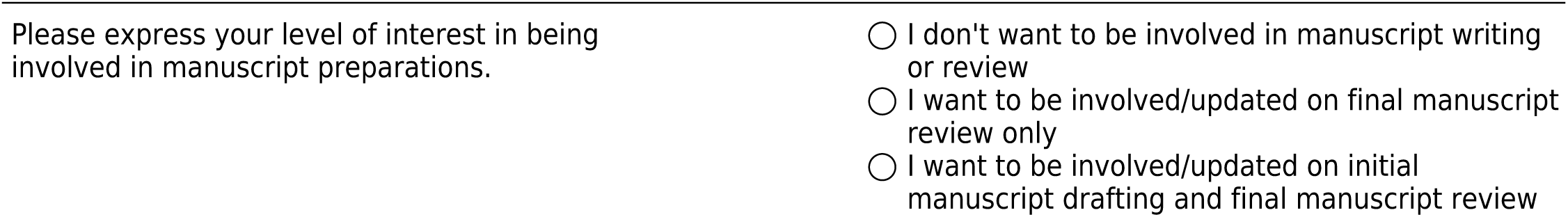

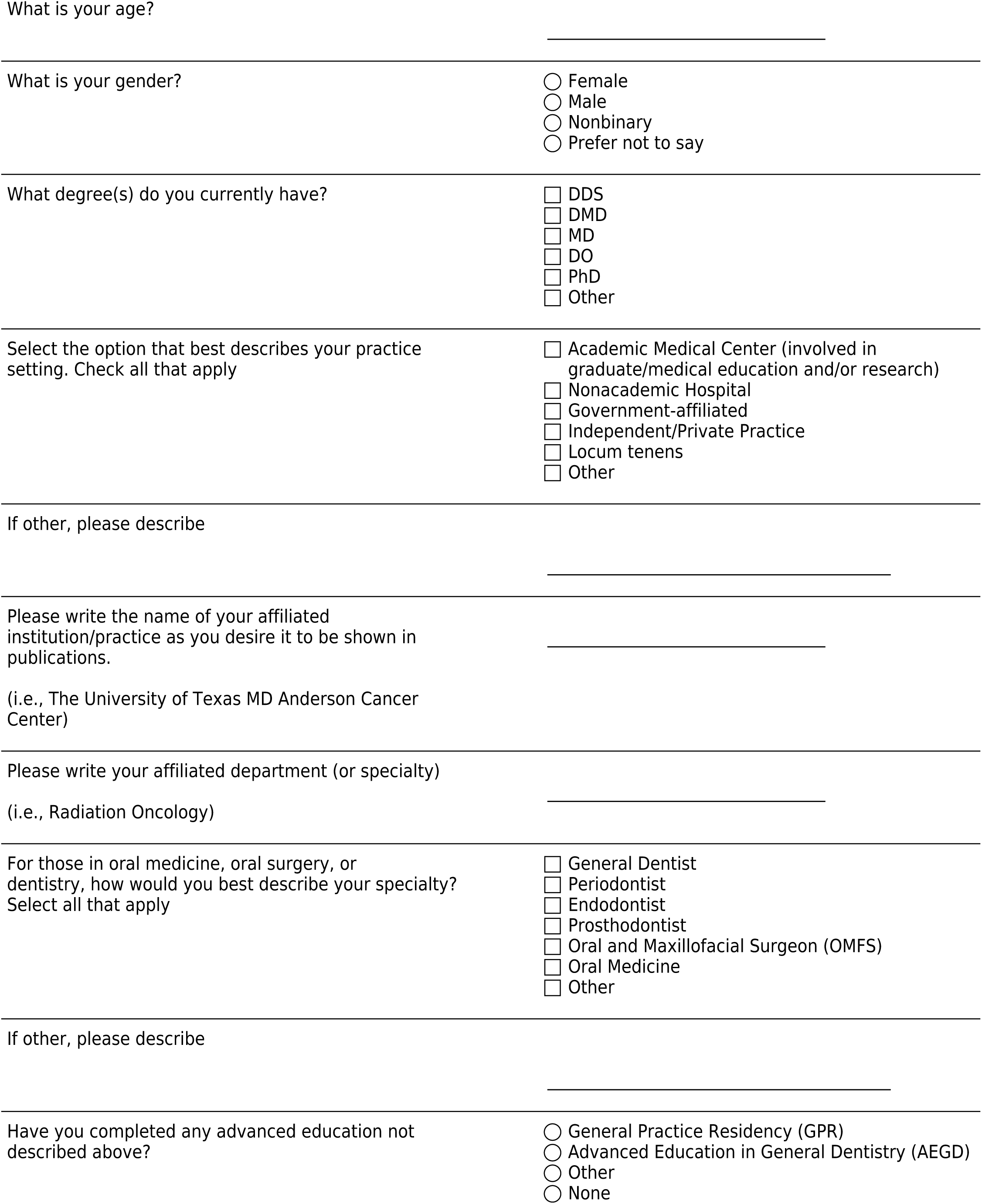
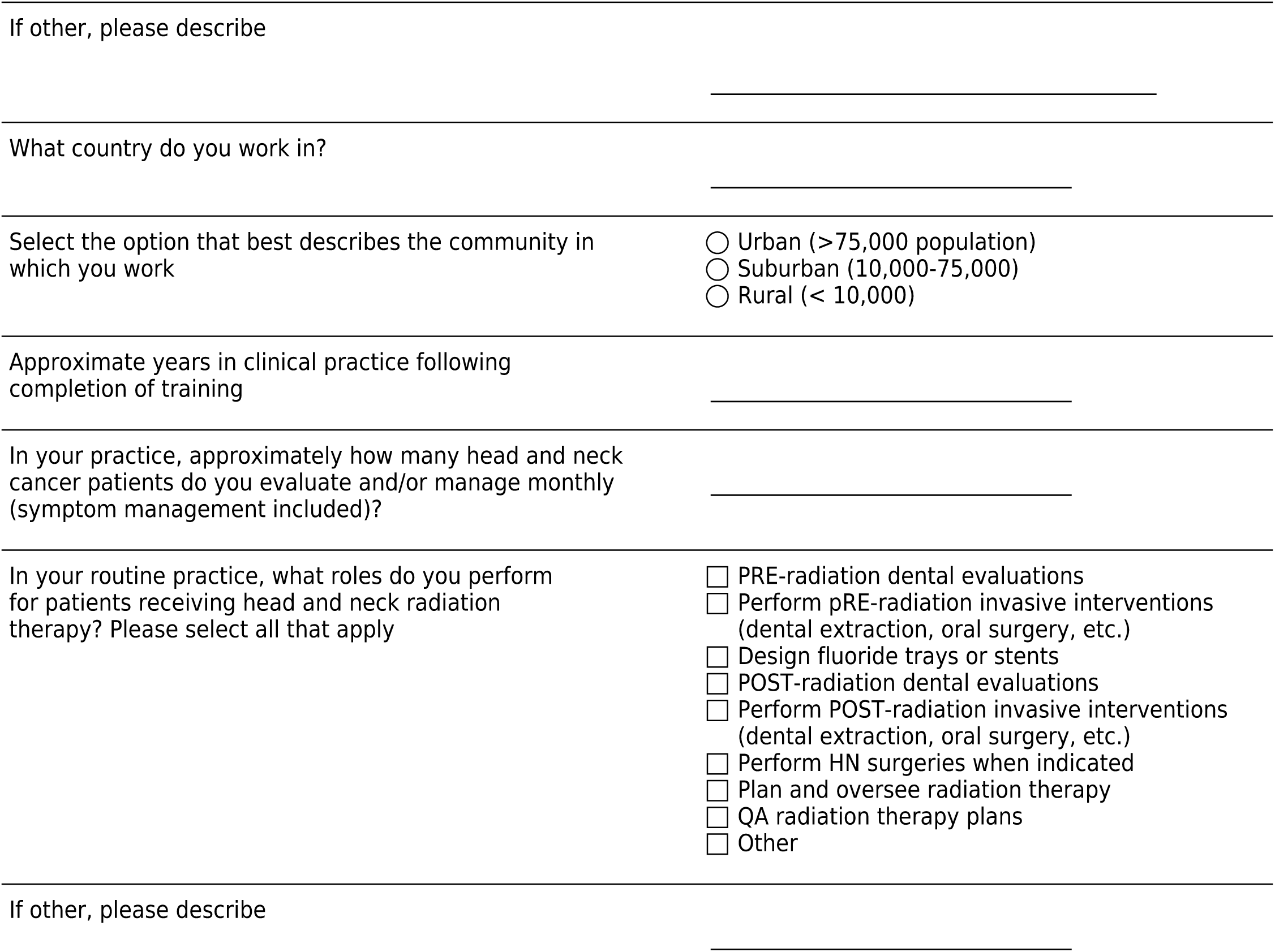

## Round 1 Defining And Classifying ORN

Please complete the survey below.

Thank you!

### SECTION 1: Defining and Classifying Osteoradionecrosis (ORN)

This section focuses on review of existing definitions and staging/grading systems for ORN. Sample cases of ORN are classified by system. Data elements within staging/grading systems are extracted for review and rating. The utility of imaging in diagnosing ORN is also reviewed.

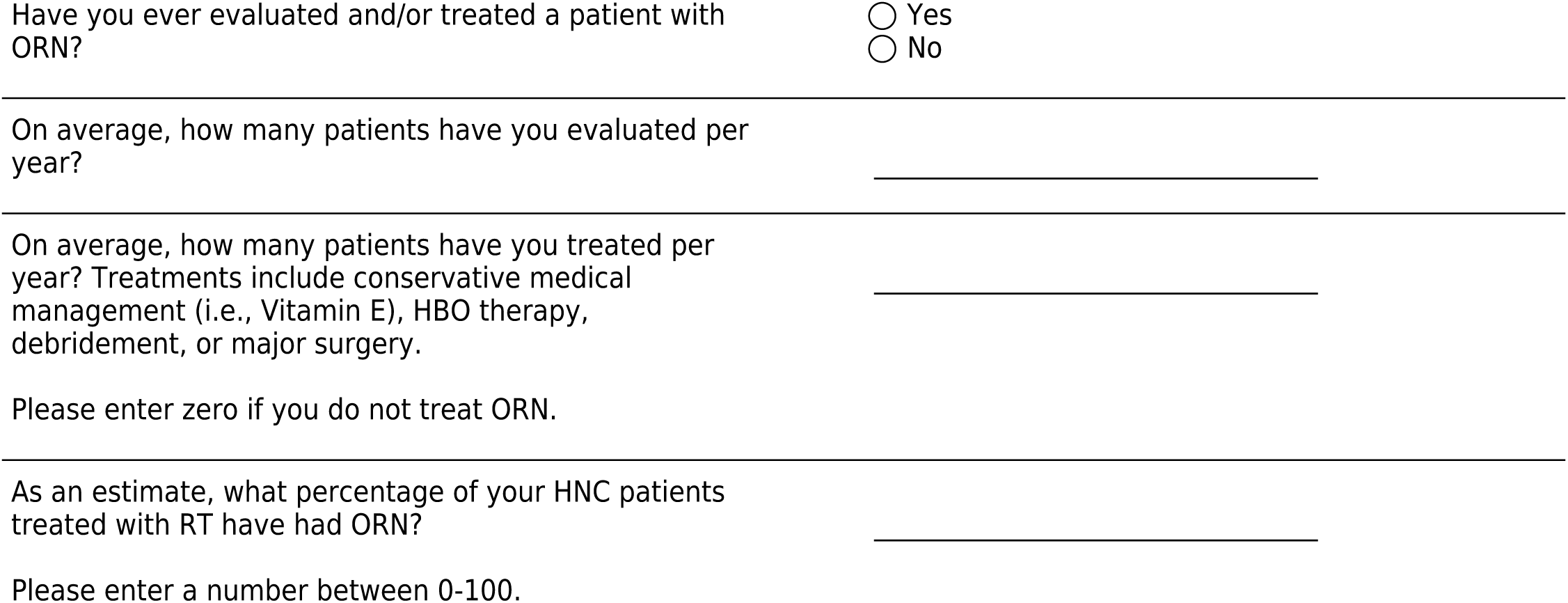

### Definitions for ORN

Example Definitions for ORN: While variations on extent of ORN, symptoms, and management are included in most staging systems, explicit definitions of ORN are not consistent (or may be absent). Please review the following published definitions for ORN. Time features (i.e., persistence of ORN) are bolded if present.

Authors (Year)

Diagnostic Criteria

Marx (1983)

An area greater than 1 cm of exposed bone in a field of irradiation that has failed to show and evidence of healing for at least 6 months

Beumer (1983)

Exposure of bone of the maxilla or mandible within the radiation treatment volume persisting for more than 3 months

Marx and Johnson (1987)

Exposure of nonviable bone which fails to heal without intervention

Epstein (1987)

An ulceration or necrosis of the mucous membrane, with exposure of necrotic bone for more than 3 months

Widmark (1989)

A non-healing mucous or cutaneous ulcer with denuded bone, lasting for more than 3 months

Harris (1992)

Exposed irradiated bone that has failed to heal over a period of 3 months in the absence of local tumor

Wong (1997)

A slow-healing radiation-induced ischemic necrosis of bone with associated soft tissue necrosis of variable extent occurring in the absence of local primary tumor necrosis, recurrence, or metastatic disease

Schwartz (2002)

A condition in which devitalized, irradiated bone becomes exposed through a wound in the overlying skin or mucosa. No tumor recurrence and it must persist for 3 to 6 months

Karagozoglu (2014)

Radiation induced necrosis of bone. Exposed bone with or without changes on plain radiograph having excluded presence of tumor tissue. Definitive ORN diagnosis if exposed bone persists for at least 1 month

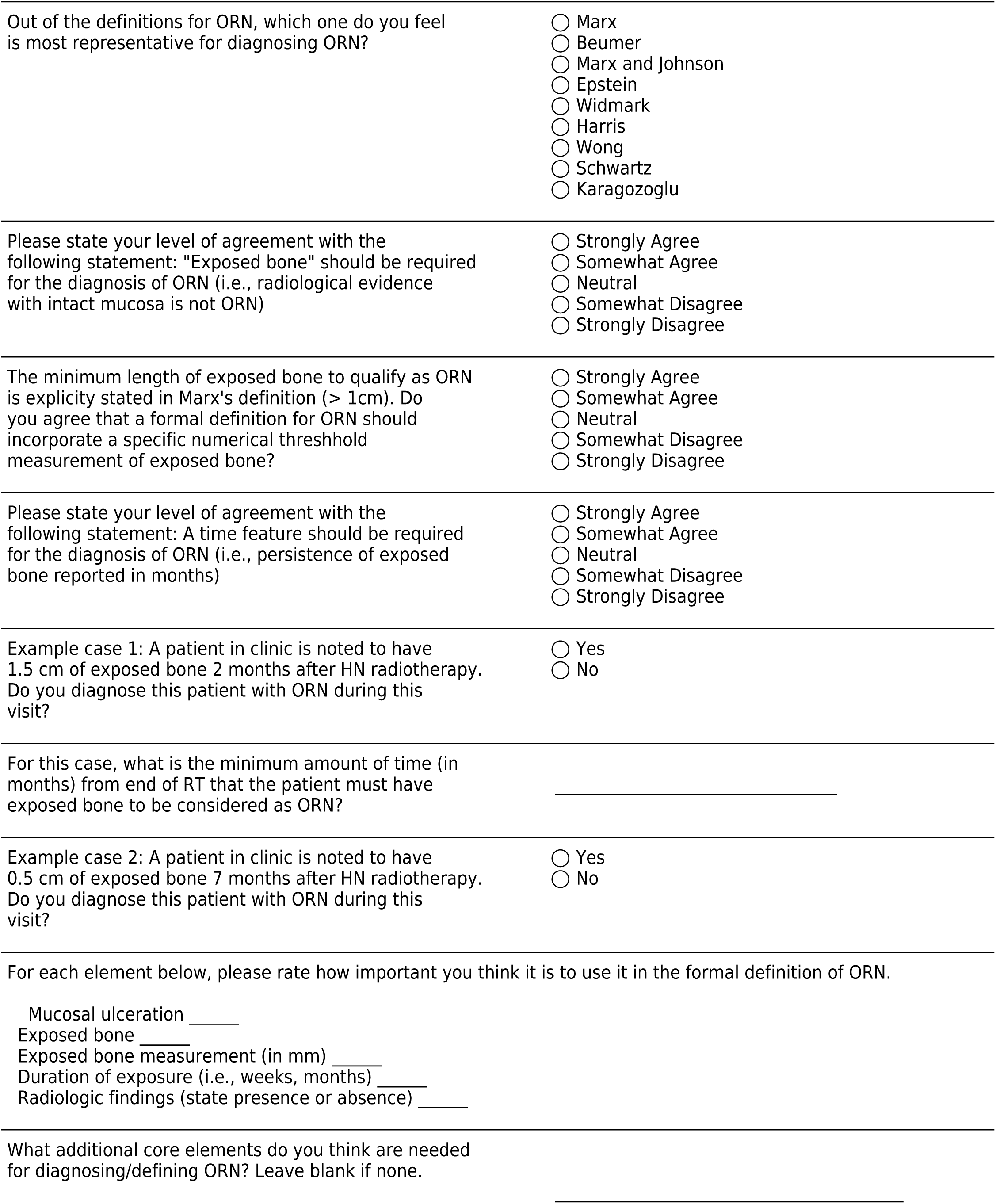

Several staging/grading systems for classifying the extent of ORN have been published, and 15 will be reviewed in this section. Please note differences in criteria which may be related to clinical findings, radiological findings, disease progression, or response to therapy.

Additionally, 3 potential ORN case scenarios will be linked to each system. In consideration of efforts to reference a system’s knowledge to another, please do your best at classifying these cases according to the specific staging/grading system in question. Select “unable to classify” only if very unsure on how to classify the patient.

Abbreviations: Hyperbaric oxygen therapy (HBO)

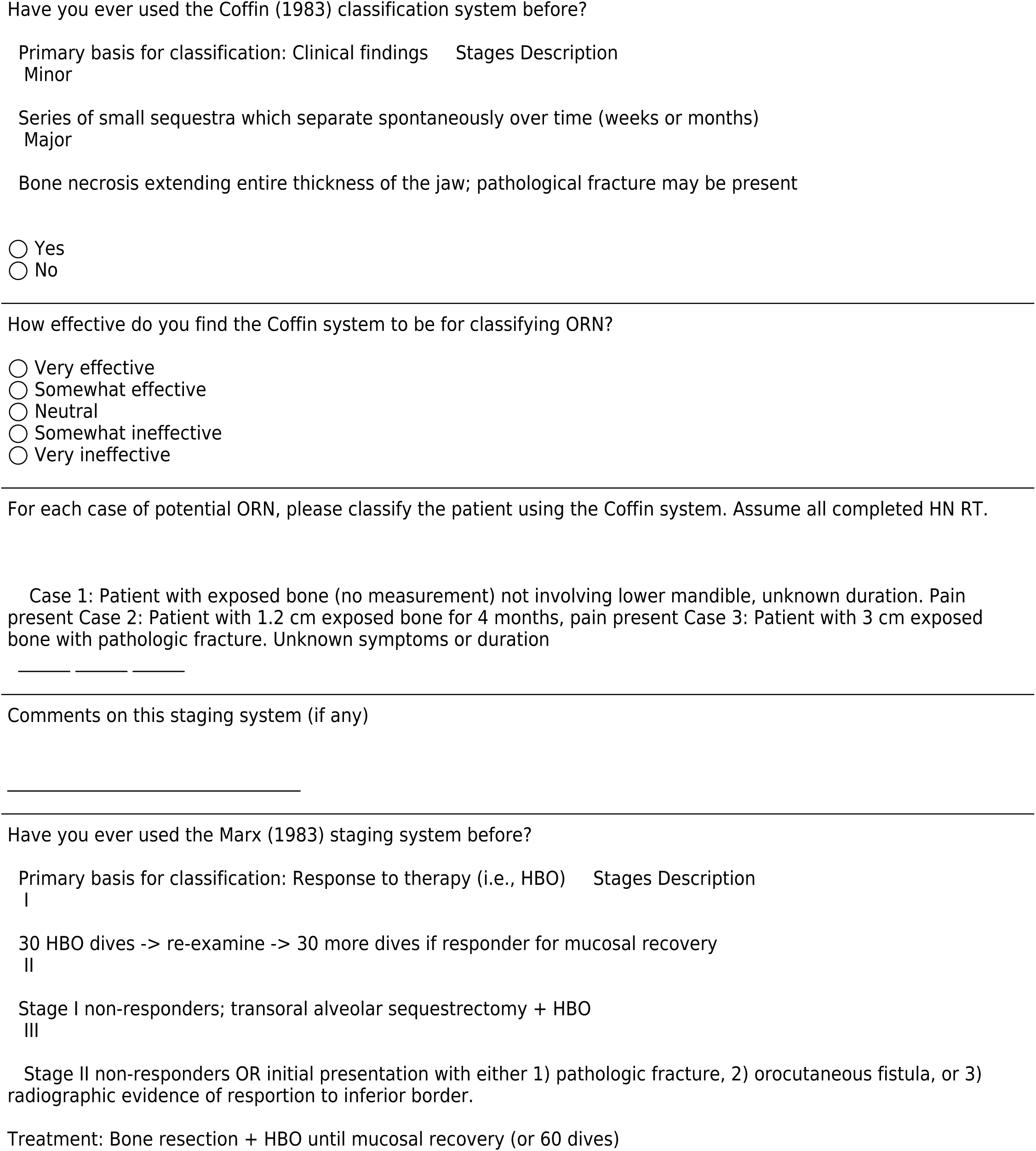
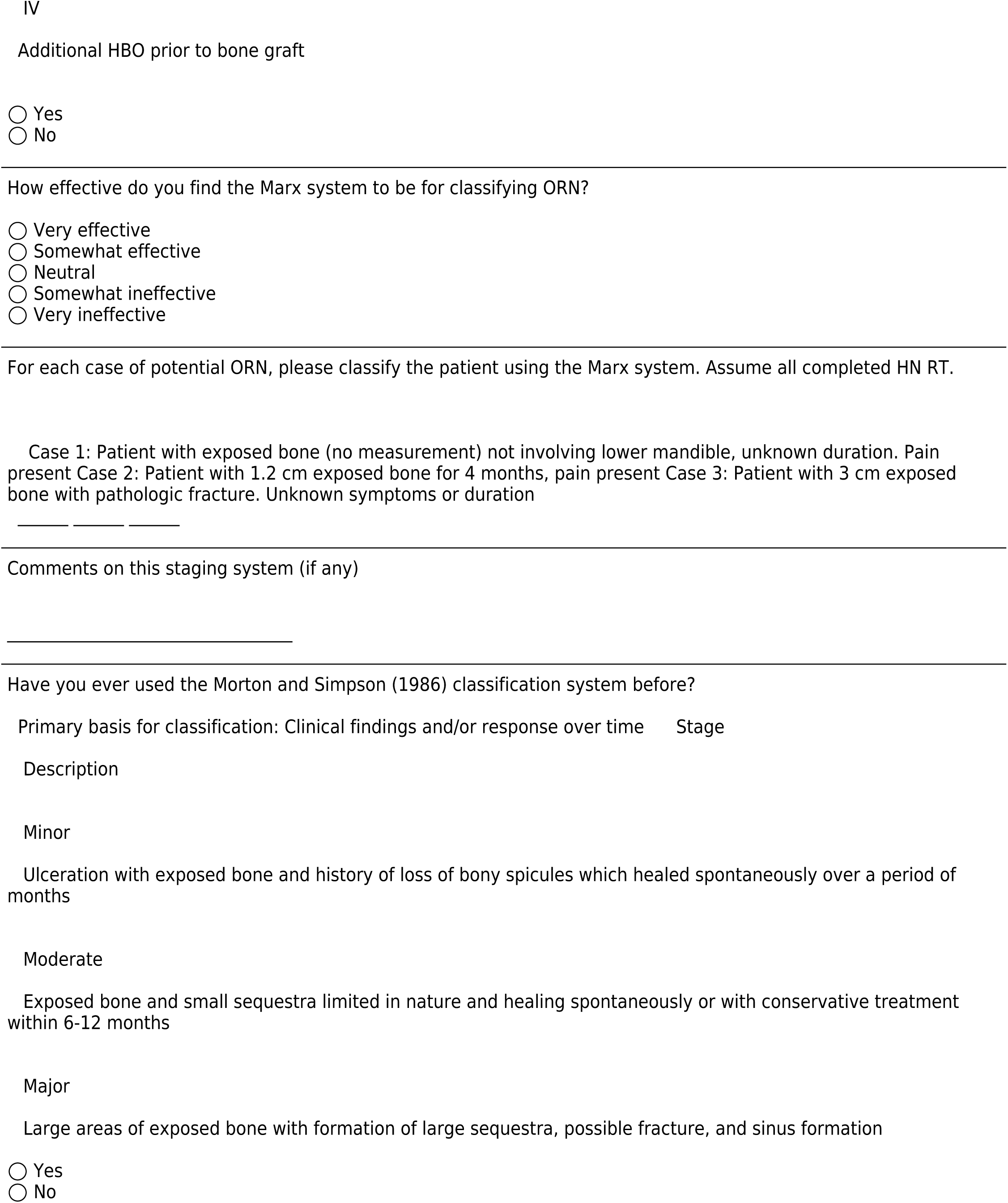
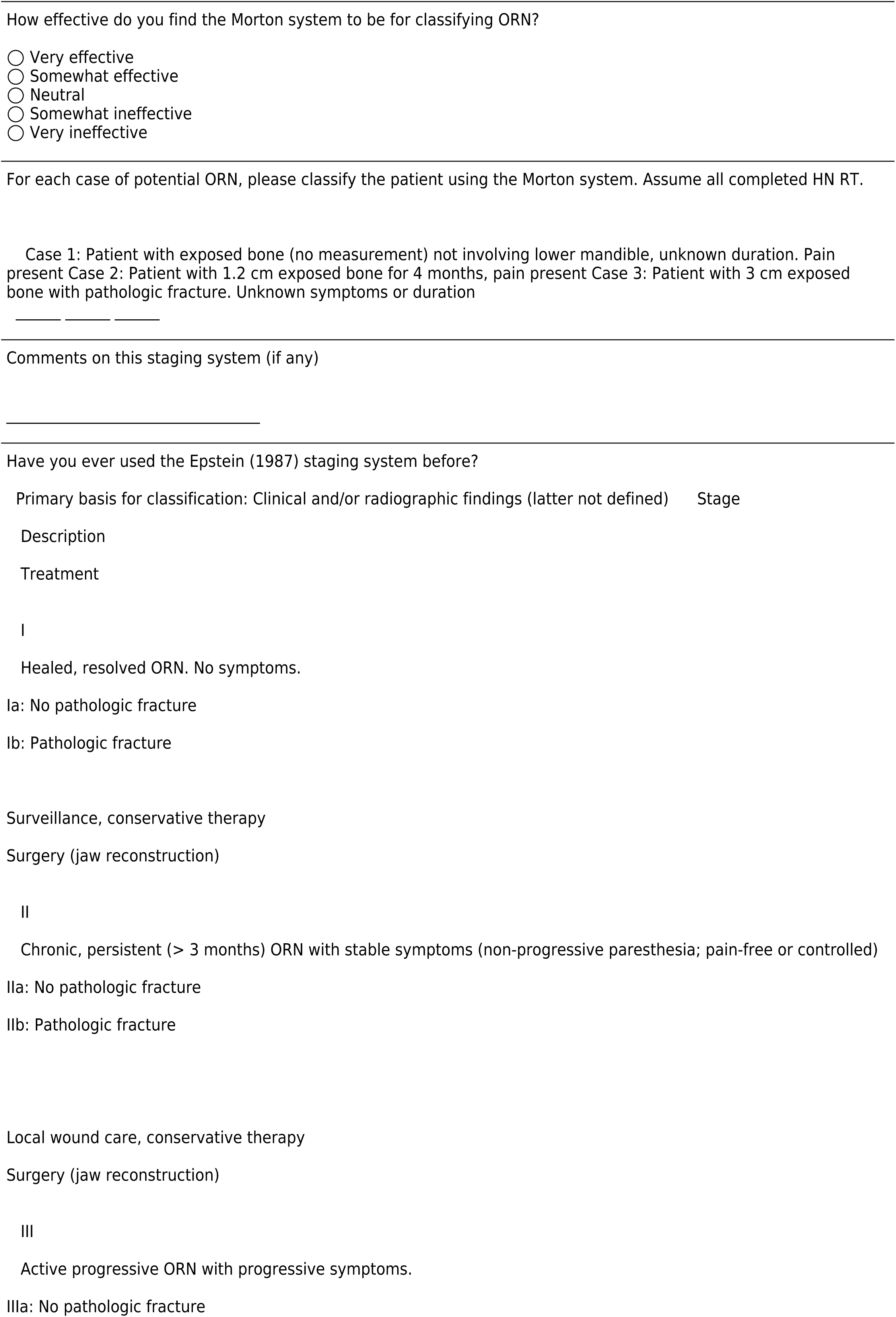
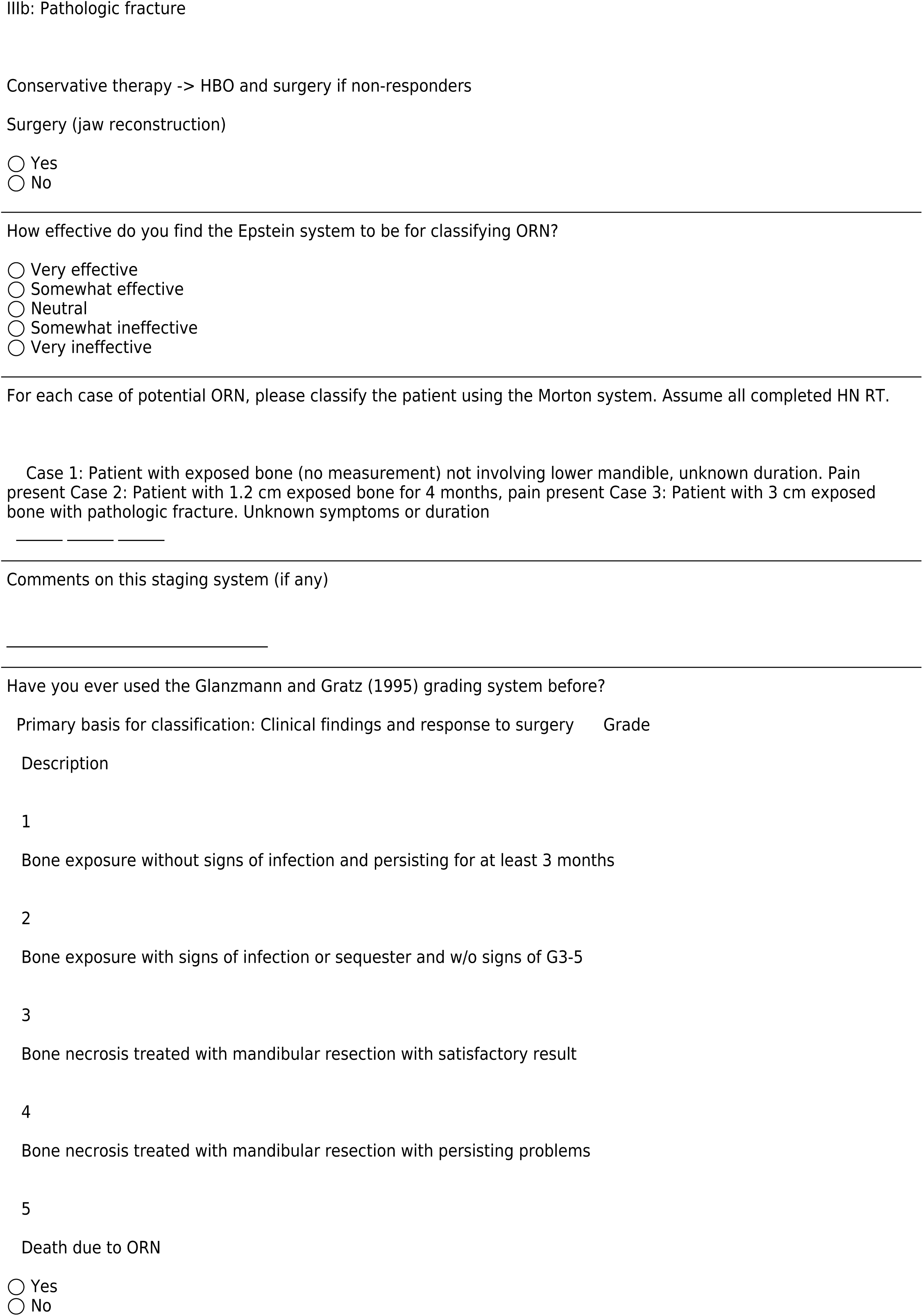
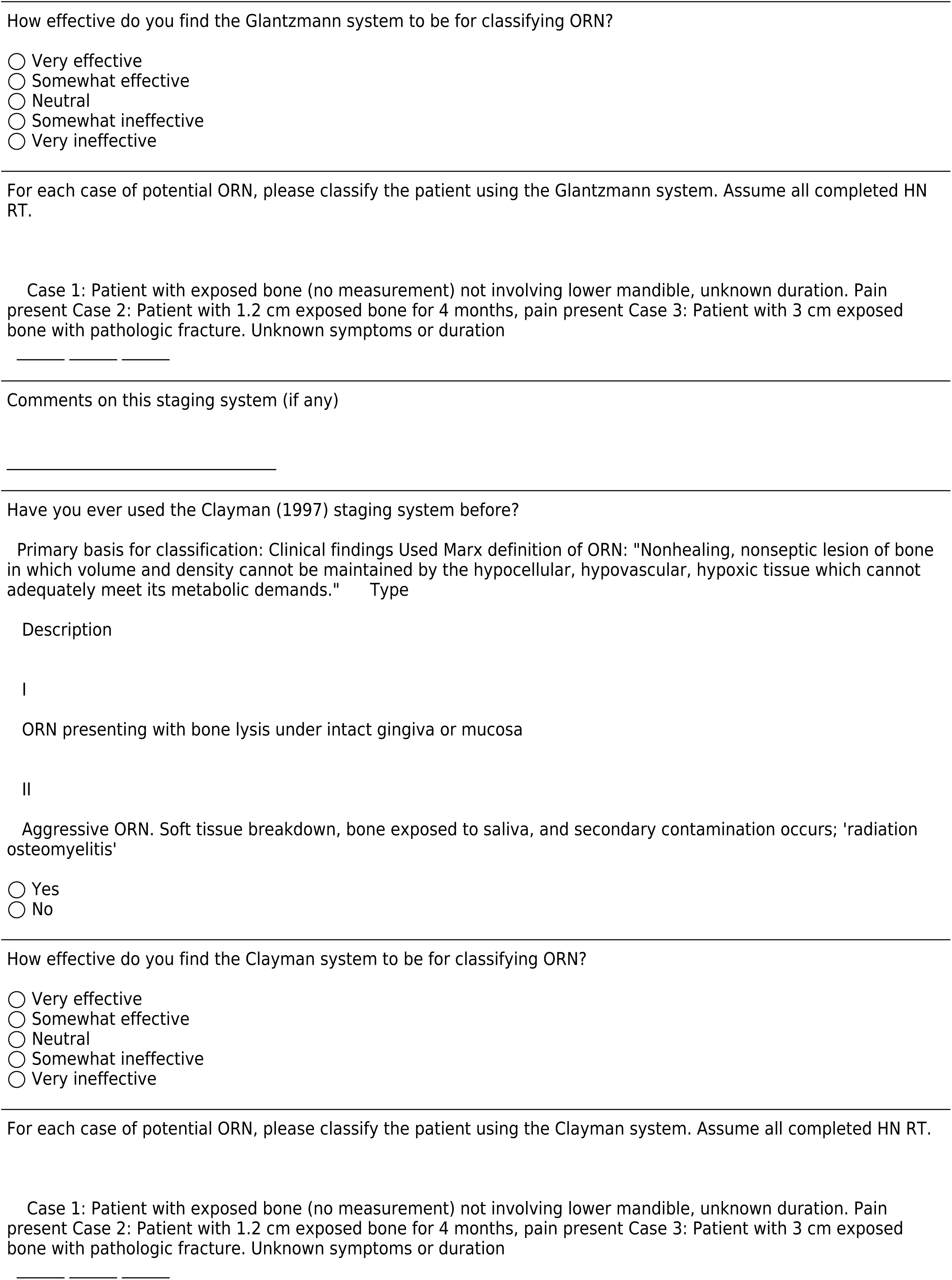
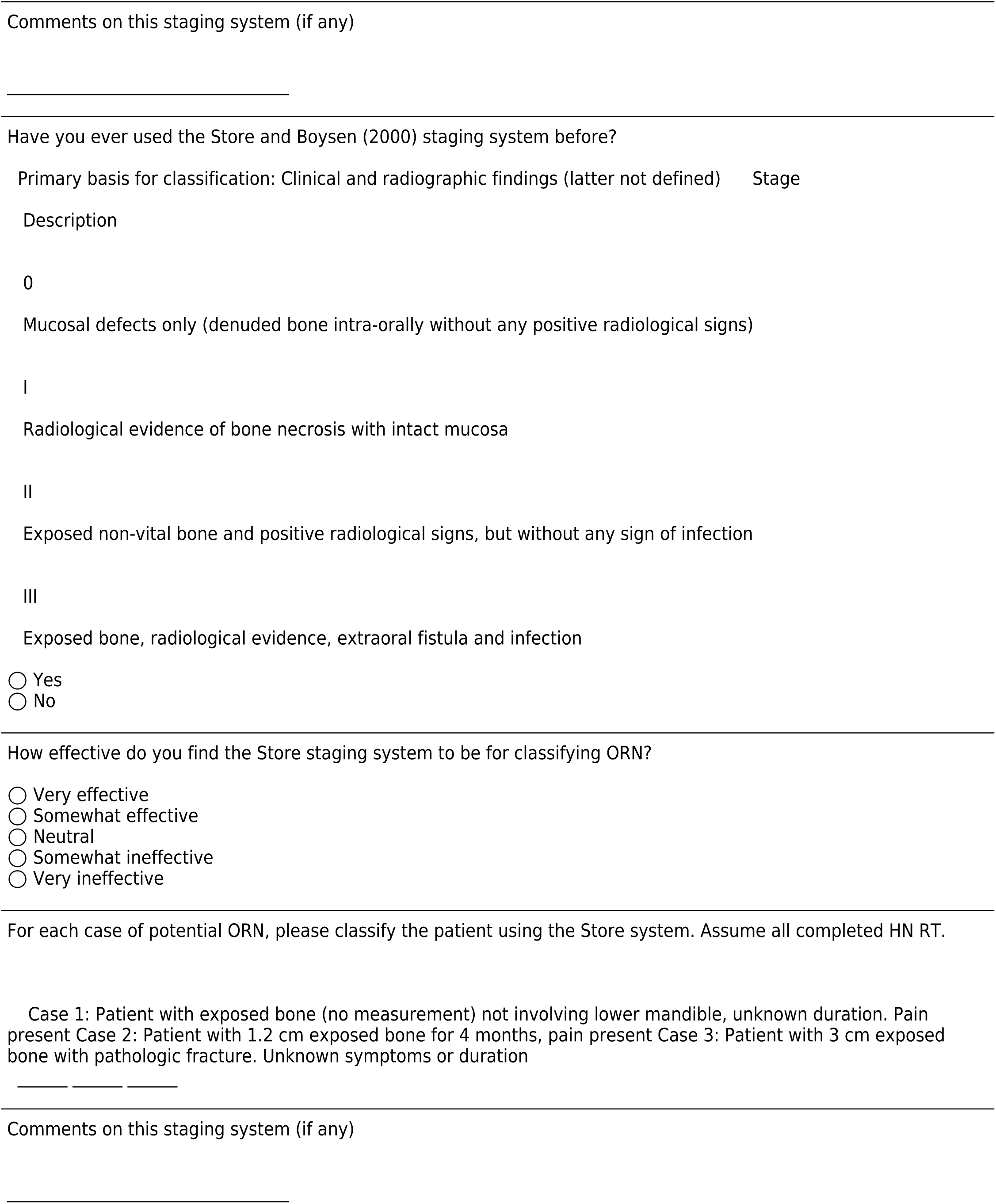
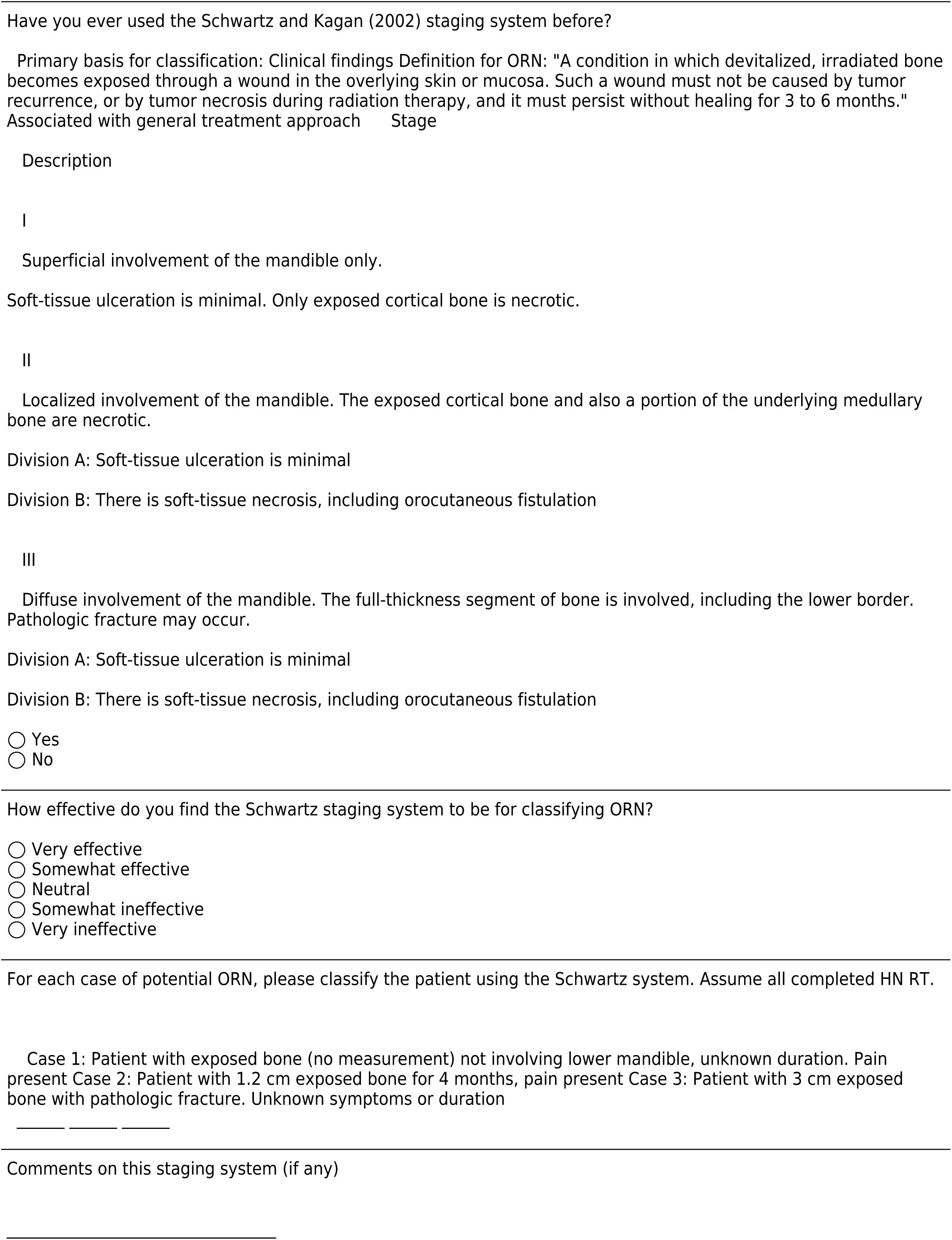
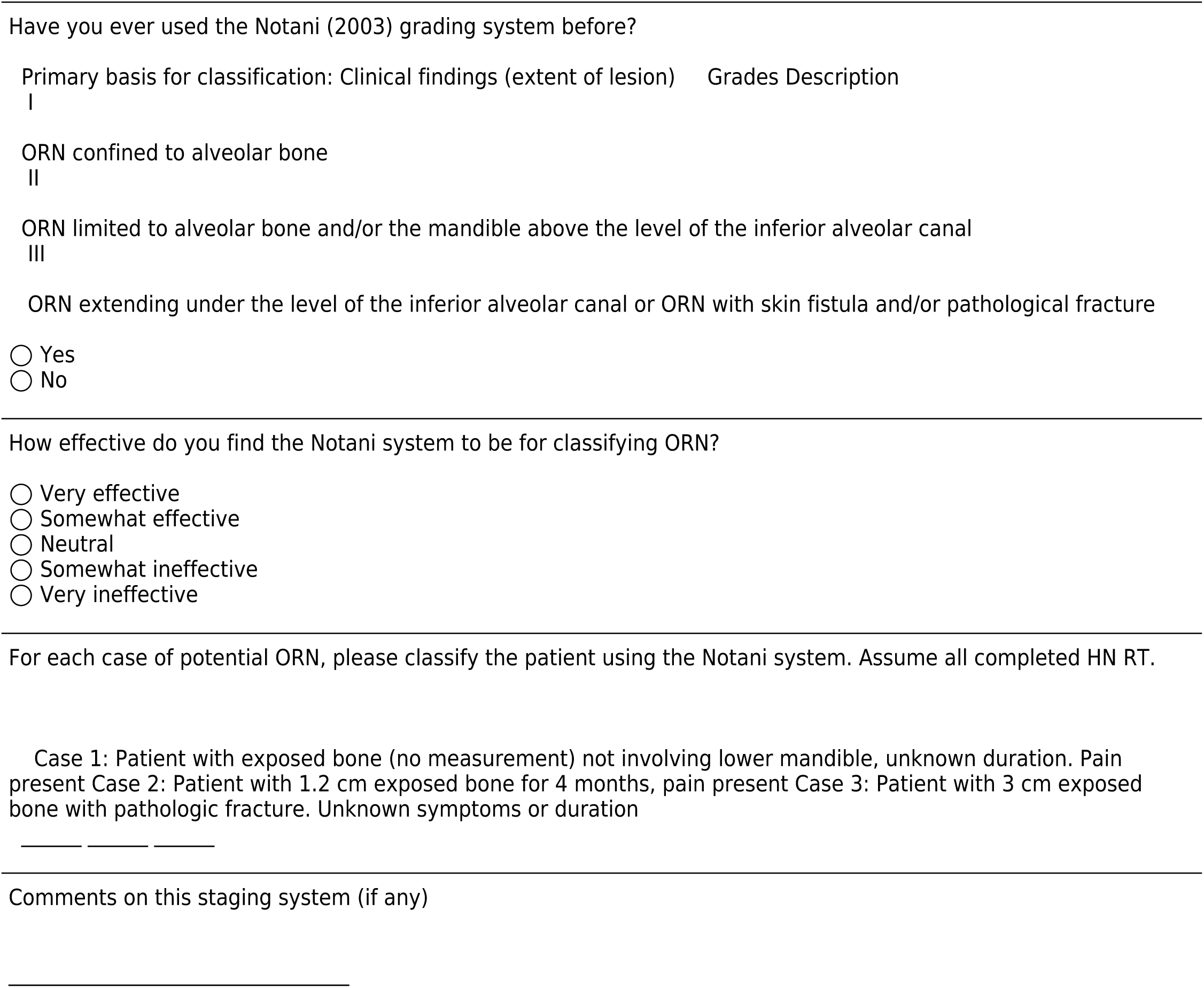

Shaw et al. (2017) proposed a modified Notani ORN classification for use in clinical trials. As shown below, this system adds duration of exposed bone (6 month threshold) and minor bone spicules (MBS), defined as ‘not ORN’ with a surface area of < 20mm^2.

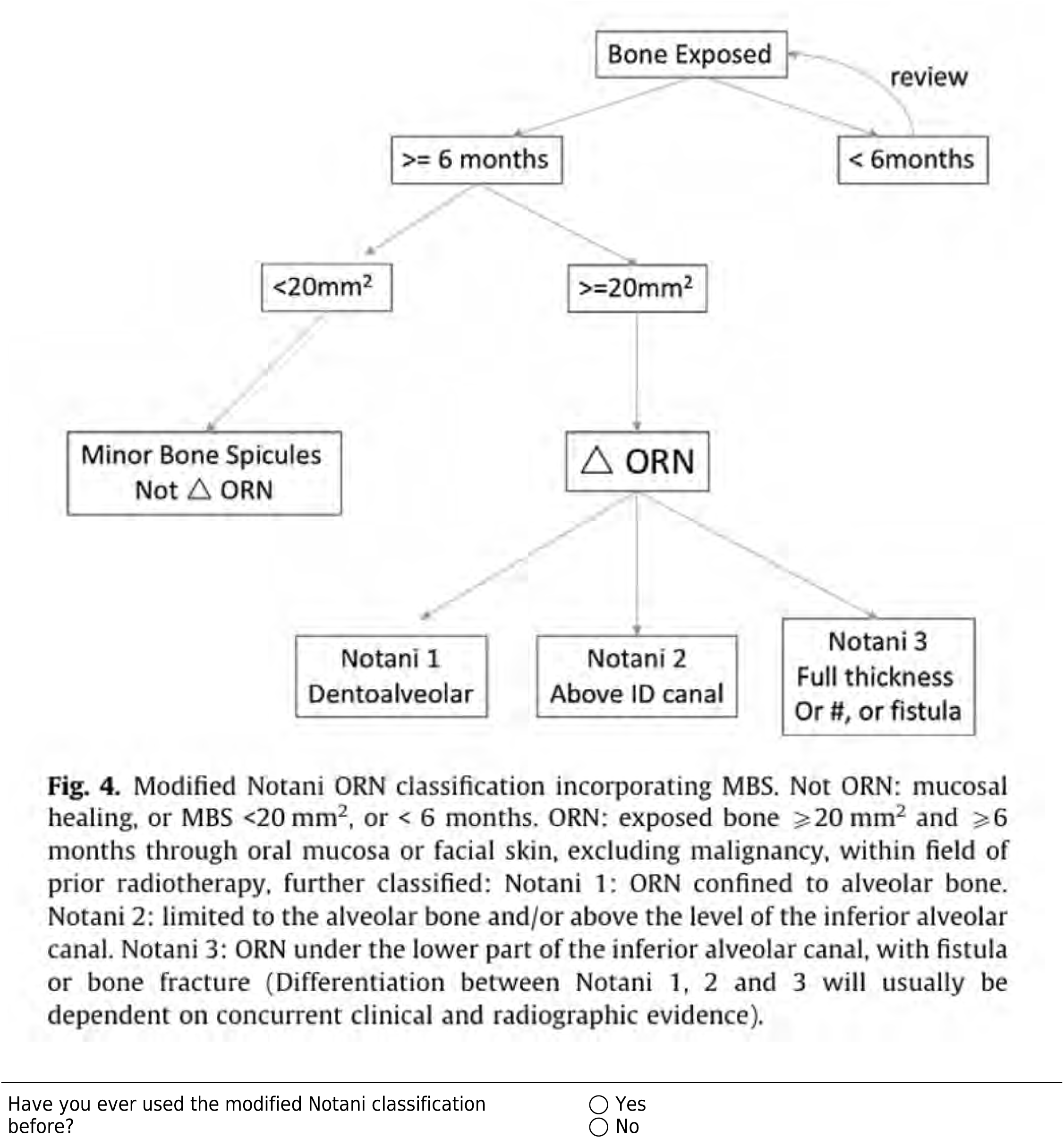
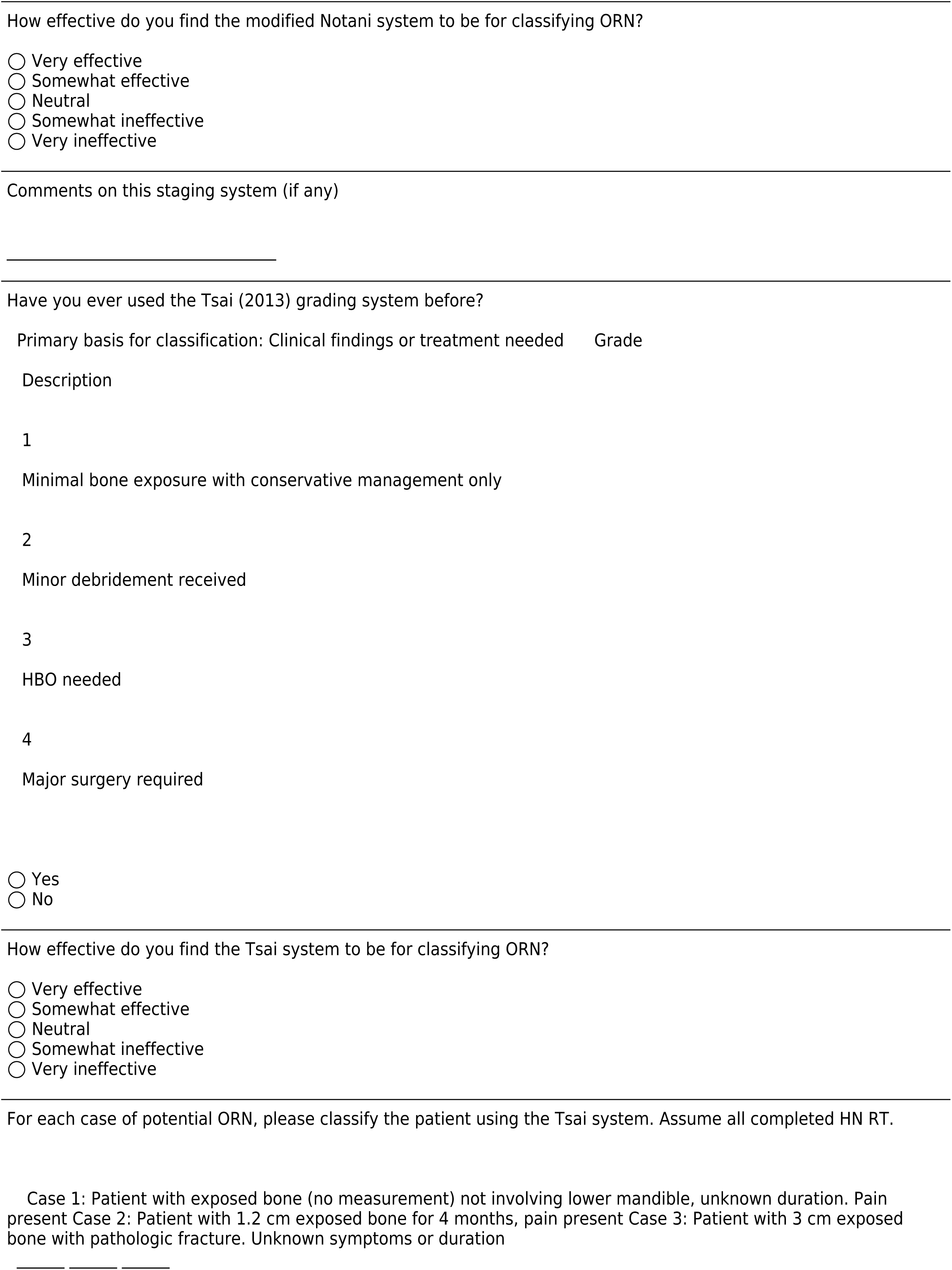
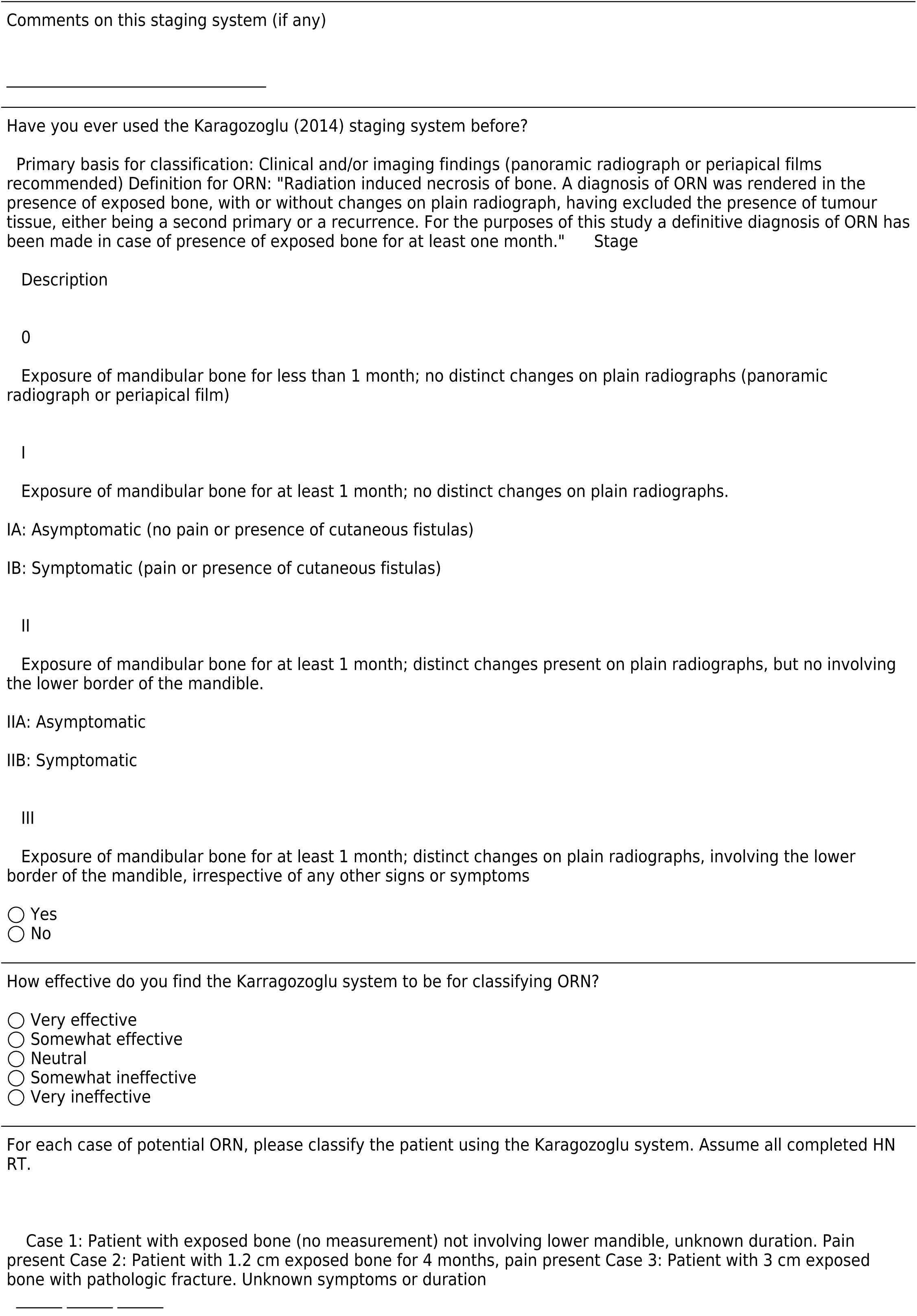
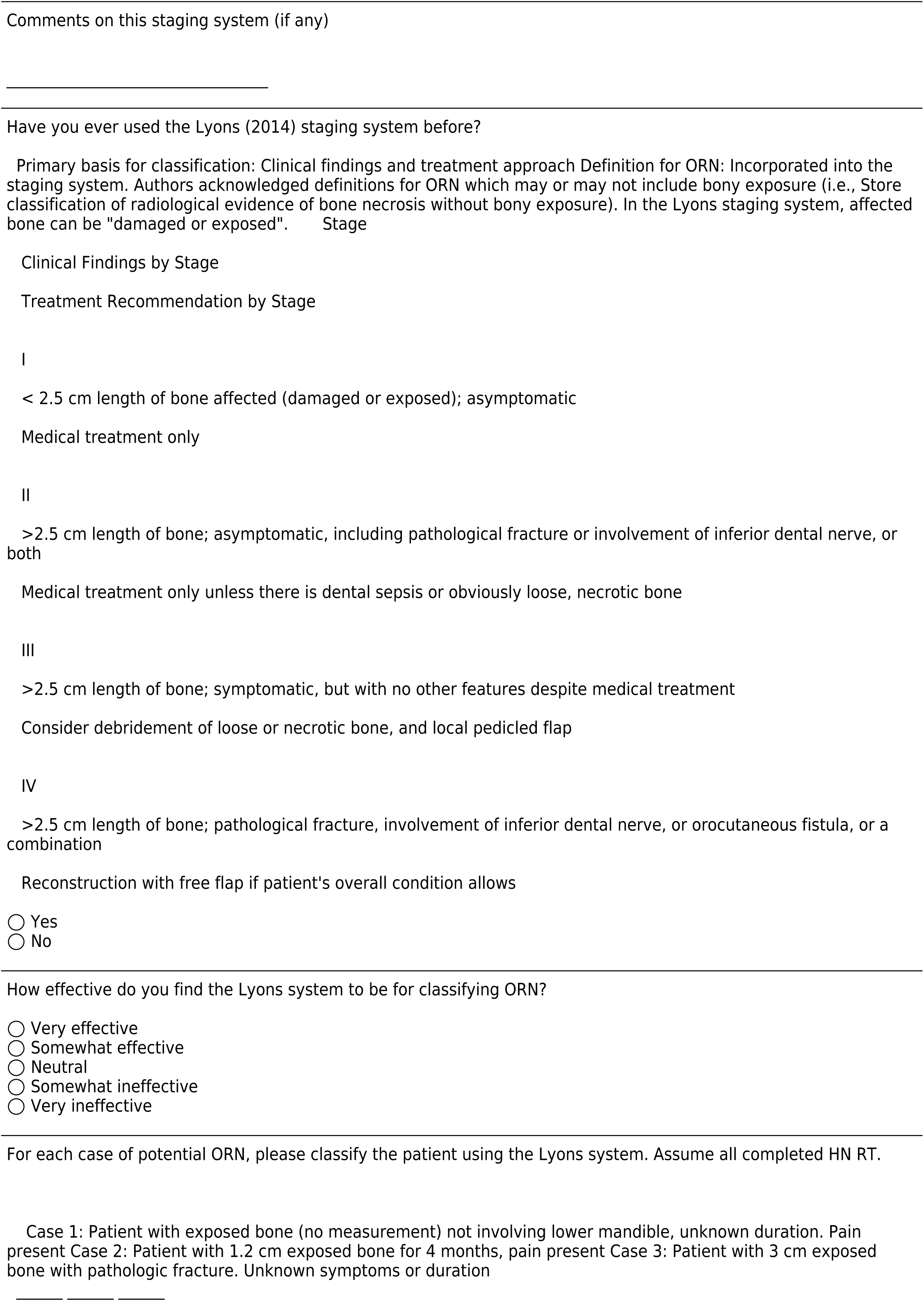
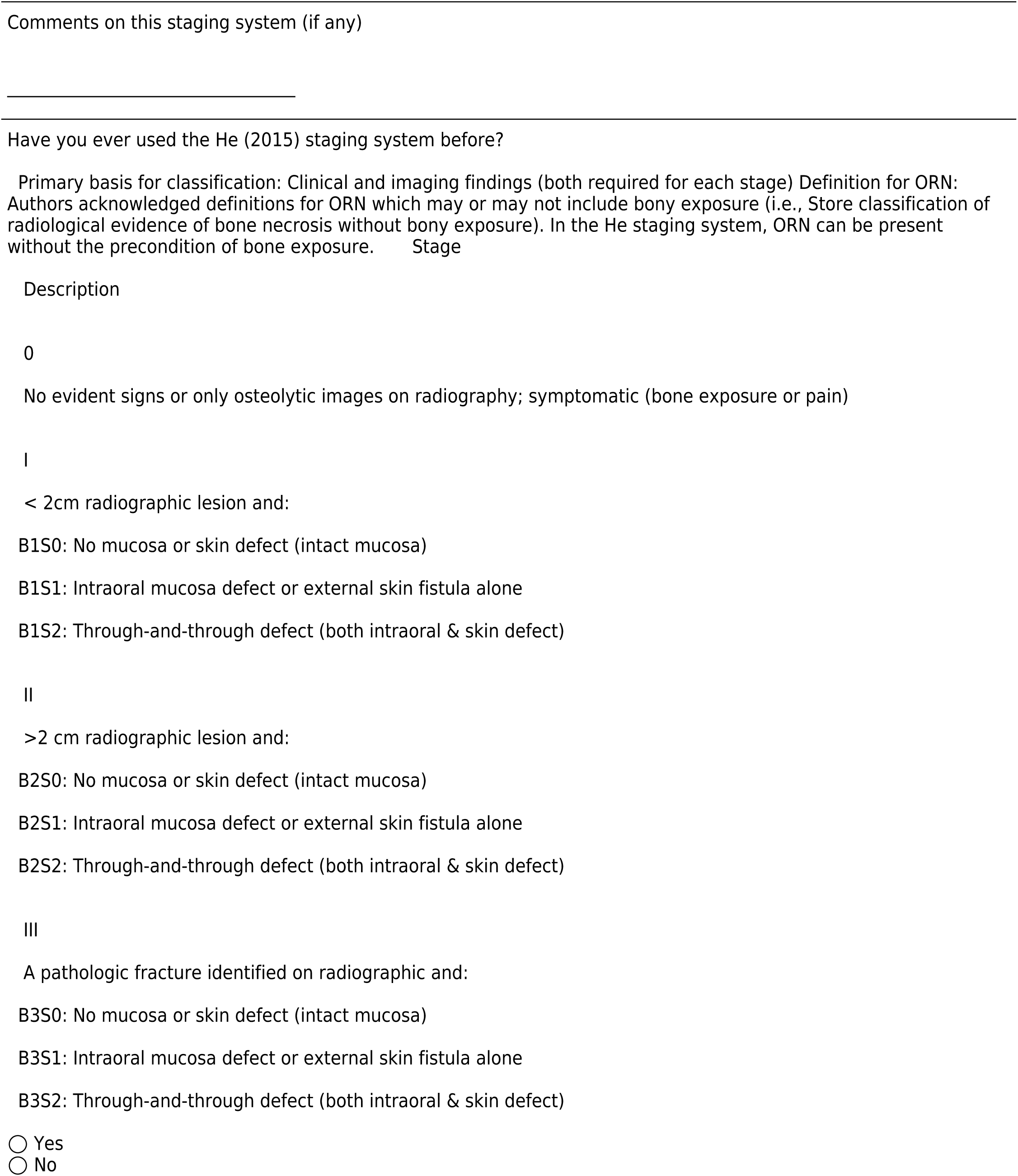
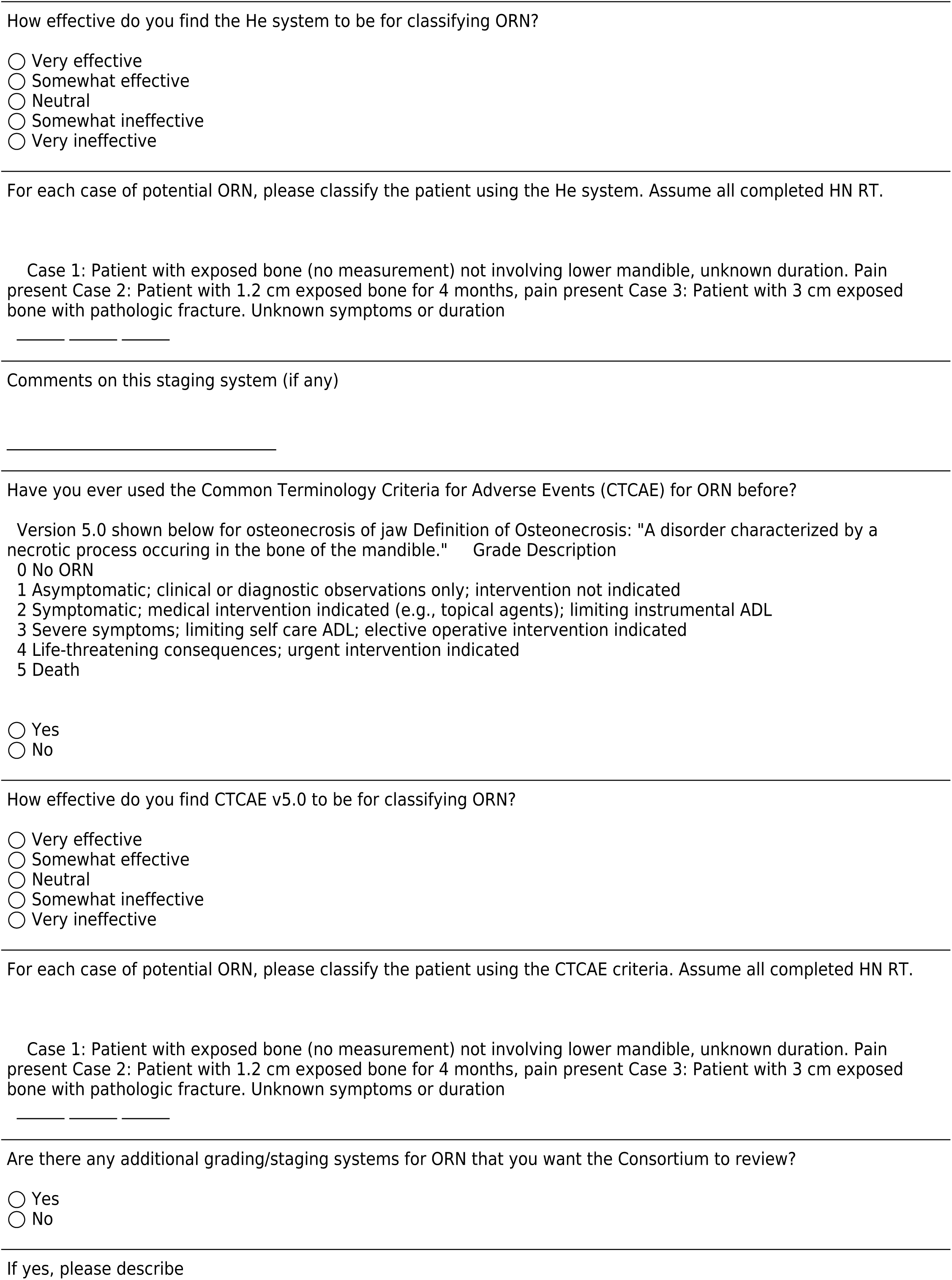

### Staging/grading data elements review

Please click on the link to find an overview of reported data elements per staging/grading systems. You may want to have this opened separately while answering the next series of questions.

[Attachment: “ORN elements summarized-Table 1.png”]

Please rate the level of importance for each. Consider items labeled as “very important” for mandatory documentation during follow ups on all HNC cases treated with RT, and/or for inclusion in an ORN ontology.

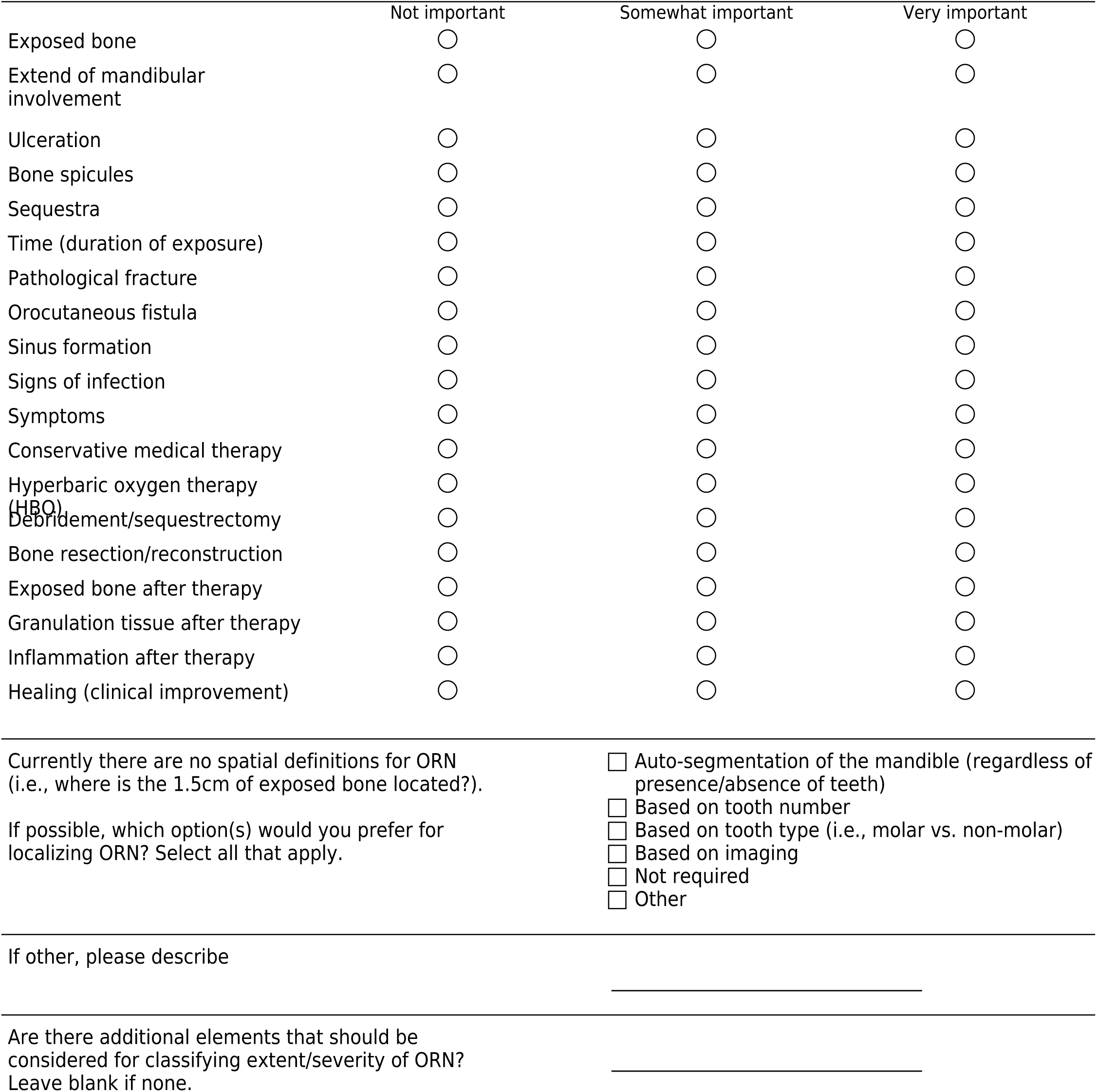

### Imaging correlates of ORN

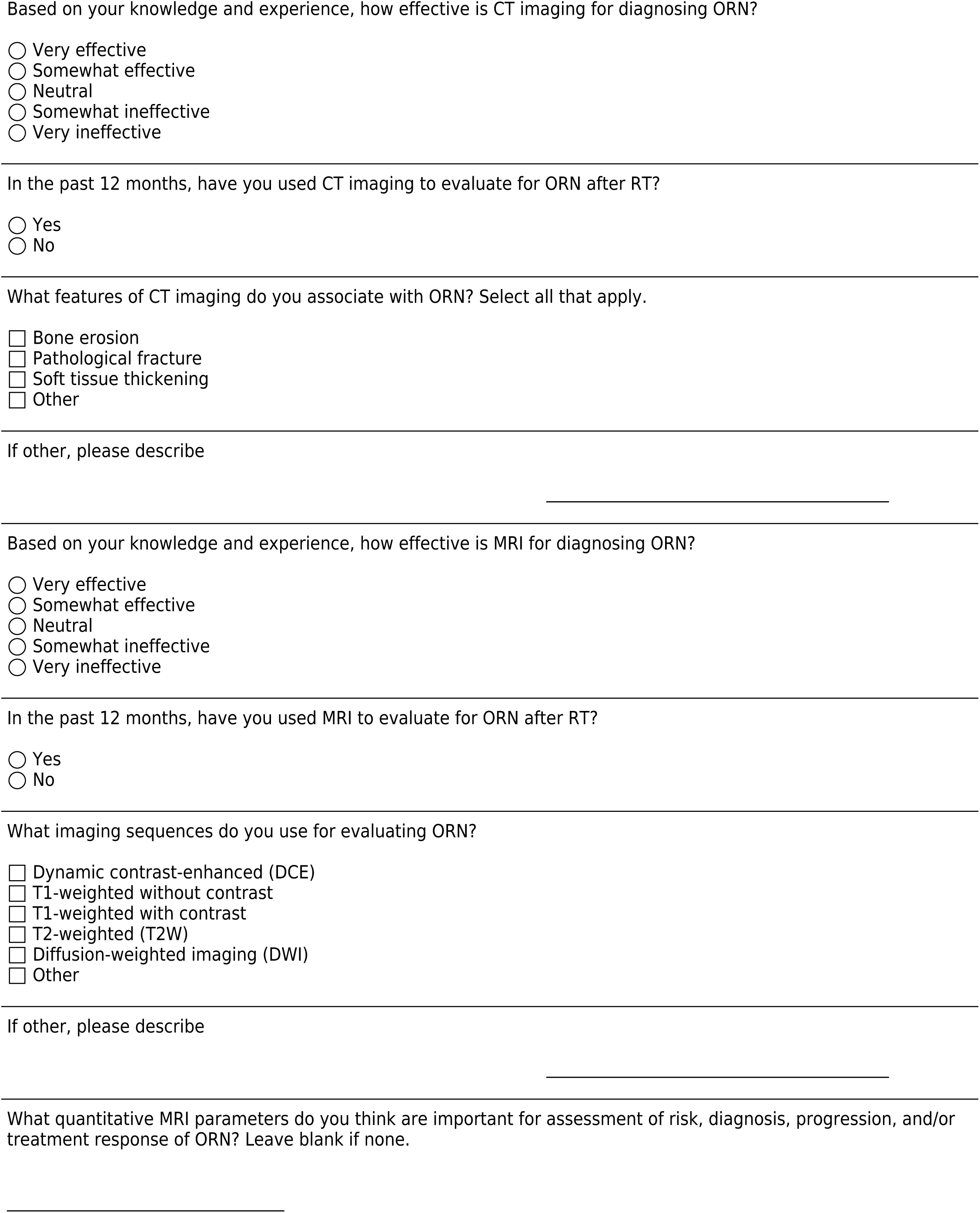
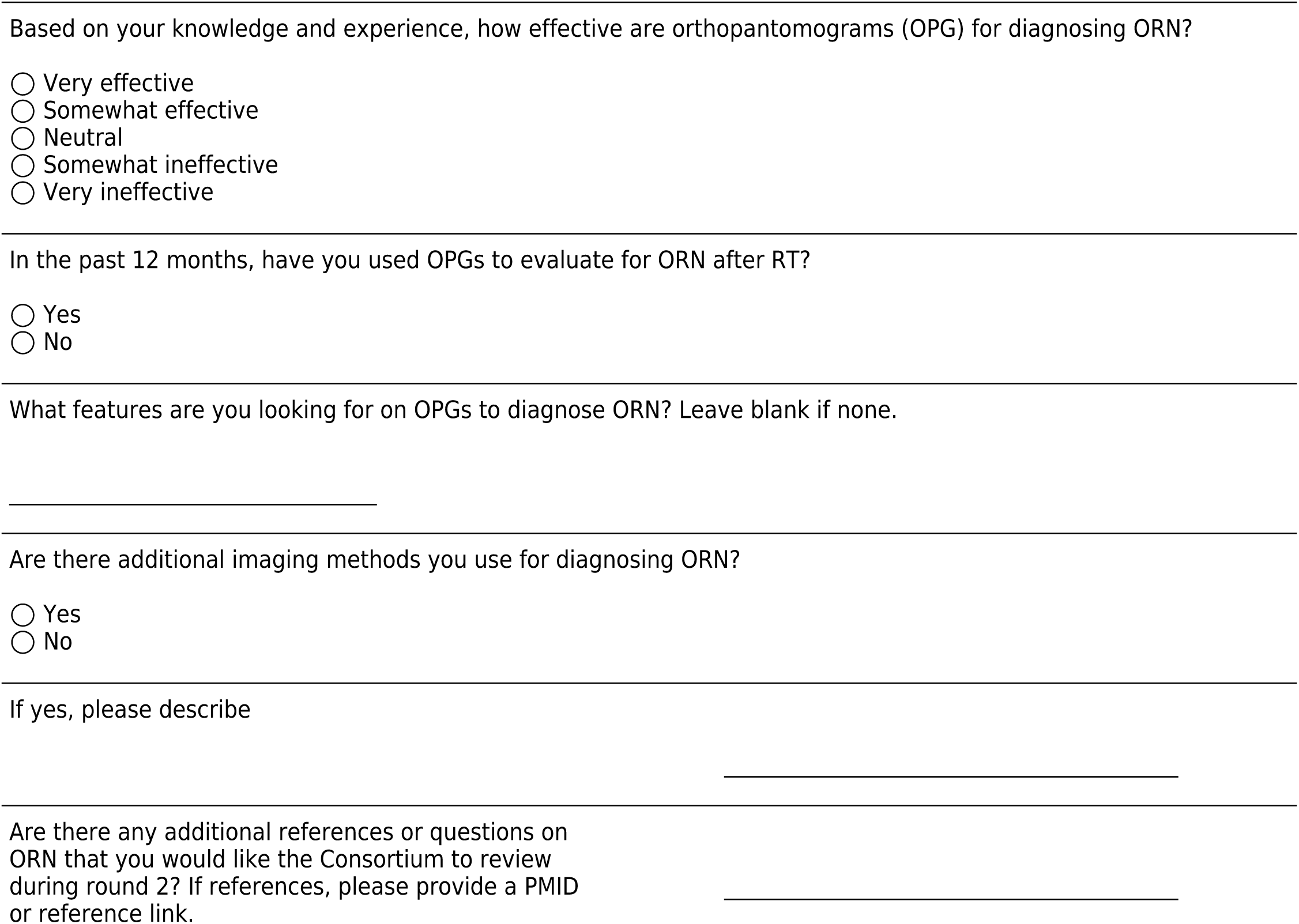

## Round 2 ORN

Please complete the survey below.

Thank you!

Dear Oral Consortium Members,

Thank you for your valuable responses to Round 1 of the ORN-RADMAP Delphi study!

We received similar feedback from many of you in that Round 1 posed important questions and challenges with our current ability to 1) diagnose ORN due to the lack of a consensus-based explicit definition and 2) classify various cases of potential ORN given 15+ existing staging/grading systems.

Using group feedback from Round 1, our main goals for Round 2 are:

Begin formulation of an explicit concept definition for ORN. Differentiate between features of bone-based disorders (for ORN staging) and potential modifiers of disease severity. Summarize group feedback for RADMAP with secondary questions on visualizations. Note: For Likert scale-type questions (i.e., strongly disagree to strongly agree), please try to minimize the use of ‘neutral’ in order to assist with consensus formation in future rounds. Also, some questions may sound repetitive but are useful for consensus processes. Thank you!

The focus of Round 3 will be confirmation of consensus-based ORN diagnostic criteria, and the build of a staging system/ontology based on Consortium-endorsed data elements.

**Table 1.** summarizes the Consortium characteristics (for those who partially/fully completed Round 1 ORN/RADMAP surveys). Note: This table may be updated based on additional responses from Consortium members.

|  | Overall<br>(N=59) |
| --- | --- |
| **Sex** |  |
| Female | 27 (45.8%) |
| Male | 32 (54.2%) |
| Nonbinary | 0 (0%) |
| Prefer not to say | 0 (0%) |
| **Age (years)** |  |
| Mean (SD) | 46.2 (8.53) |
| Median [Min, Max] | 46.0 [33.0, 68.0] |
| Missing | 3 (5.1%) |
| **Specialty** |  |
| HN Surgery | 9 (15.3%) |
| Medical Oncology | 4 (6.8%) |
| OMFS | 11 (18.6%) |
| Oral Oncology/Medicine | 8 (13.6%) |
| Physics | 2 (3.4%) |
| Radiation Oncology | 24 (40.7%) |
| Radiology | 1 (1.7%) |
| **Practice Community** |  |
| Urban (>75,000 population) | 57 (96.6%) |
| Suburban (10,000-75,000) | 2 (3.4%) |
| Rural (< 10,000) | 0 (0%) |
| **Time in practice (years)** |  |
| Mean (SD) | 15.3 (8.02) |
| Median [Min, Max] | 16.0 [1.00, 35.0] |
| **Approx. HNC patient caseload (per month)** |  |
| Mean (SD) | 47.7 (42.3) |
| Median [Min, Max] | 45.0 [0, 200] |
| Missing | 1 (1.7%) |
| **Have you evaluated/treated ORN?** |  |
| Yes | 55 (93.2%) |
| No | 4 (6.8%) |
| **Avg. pts evaluated for ORN annually (n)** |  |
| Mean (SD) | 52.6 (182) |
| Median [Min, Max] | 11.0 [0, 1320] |
| Missing | 5 (8.5%) |
| **Avg. pts treated for ORN annually (%)** |  |
| Mean (SD) | 13.4 (21.8) |
| Median [Min, Max] | 4.00 [0, 100] |
| Missing | 4 (6.8%) |
| **Avg. HNC pts with ORN (%)** |  |
| Mean (SD) | 7.06 (4.84) |
| Median [Min, Max] | 6.00 [0, 25.0] |
| Missing | 5 (8.5%) |

### SECTION 1: FORMULATION OF AN EXPLICIT DEFINITION FOR ORN

**Group Feedback and Review of Existing Diagnostic Standards**

- None of the 9 published ORN definitions were selected by even 25% of the group as the most representative for the disease entity (see Figure 1). The top 4 candidates were: Harris (n=13, 22%), Schwartz (n=13, 22%), Karagozoglu (n=10, 17%), and Wong (n=8, 14%).

From the top 4 definitions (Figure 1), 6 distinct features could be extracted with only 3 features included in all four: exposed [vs. necrotic] bone, RT-induced disorder [i.e., irradiated bone], and absence of tumor.

**Table 2:** ORN Features per existing definitions and based on free text comments from the group.

| Features |
| --- |
| Count in Top 4 definitions |
| Favored in definition* (n, (%)) |
| Exposed bone (vs. bone necrosis) 4 57 (100%) |
| Caused by radiation therapy 4 Not asked, but recommended in comments |
| Absence of tumor 4 Not asked, but recommended in comments |
| Time feature 3 54 (92%) |
| Imaging feature 1 56 (95%) |
| Soft tissue necrosis (ulceration) 1 49 (83%) |
| * Over 70% of the group rated this attribute as 'somewhat important' or 'very important' |

**Figure 1.**
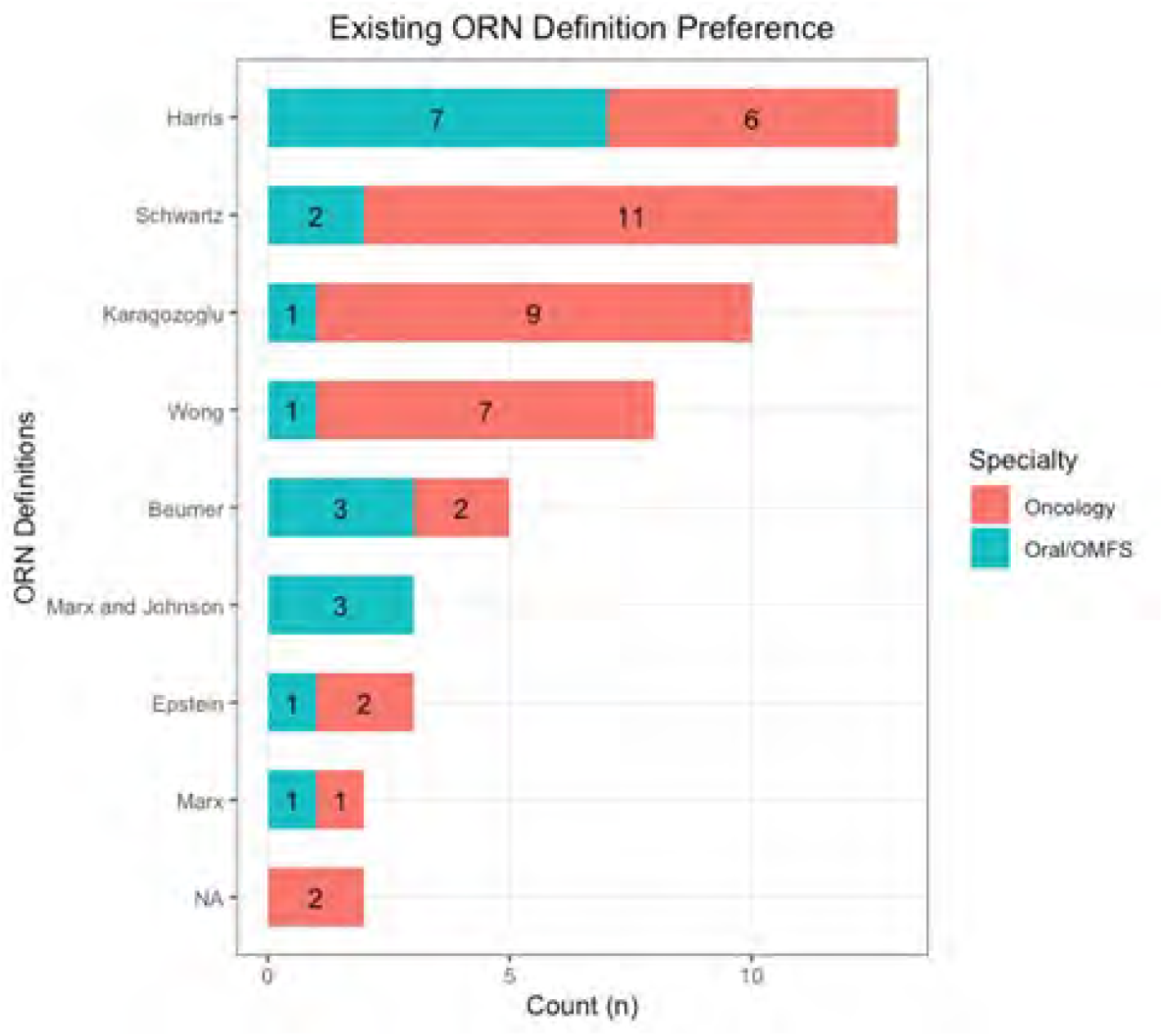
Rating of Existing ORN Definitions.

To date, there is no existing International Classification of Disease (ICD) diagnostic code specific to ORN. Other relevant internationally standardized medical classification terminologies and ontologies that do include codes for ORN include:

Medical Dictionary for Regulatory Activities (MedDRA) Systemized Nomenclature of Medicine-Clinical Terms (SNOMED-CT) These definitions can serve as a foundation to build the Consortium’s explicit definition for ORN. Let’s first review how each standard defines disorders such as osteonecrosis (ON) and ORN.

MedDRA is a ‘clinically-validated international terminology’ with a standardized hierarchy as described below (SOC --> HLGT --> HLT --> PT --> LLT). More specific diagnoses are typically coded as a ‘Lowest level term’ or LLT.

MedDRA Hierarchy description from https://www.meddra.org/how-to-use/basics/hierarchy :

”The structure of MedDRA is very logical. There are five levels to the MedDRA hierarchy, arranged from very specific to very general. At the most specific level, called “Lowest Level Terms” (LLTs), there are more than 80,000 terms which parallel how information is communicated. These LLTs reflect how an observation might be reported in practice. Each member of the next level, “Preferred Terms” (PTs) is a distinct descriptor (single medical concept) for a symptom, sign, disease diagnosis, therapeutic indication, investigation, surgical or medical procedure, and medical social or family history characteristic. Each LLT is linked to only one PT. Each PT has at least one LLT (itself) as well as synonyms and lexical variants (e.g., abbreviations, different word order). Related PTs are grouped together into “High Level Terms” (HLTs) based upon anatomy, pathology, physiology, aetiology or function. HLTs, related to each other by anatomy, pathology, physiology, aetiology or function, are in turn linked to “High Level Group Terms” (HLGTs). Finally, HLGTs are grouped into “System Organ Classes” (SOCs) which are groupings by aetiology (e.g., Infections and infestations), manifestation site (e.g., Gastrointestinal disorders) or purpose (e.g., Surgical and medical procedures)…” MedDRA Code Examples:

Code for osteonecrosis: 10031264 Code for osteoRADIOnecrosis: 10067352 Code for medication-related osteonecrosis of jaw: 10084881 Key takeaway points:

ORN is an LLT to ‘radiation injury’, ‘bone disorders NEC (not elsewhere classified)’, and ‘necrosis and vascular insufficiency’ Figure 2: MedDRA Hierarchy for ORN and ON

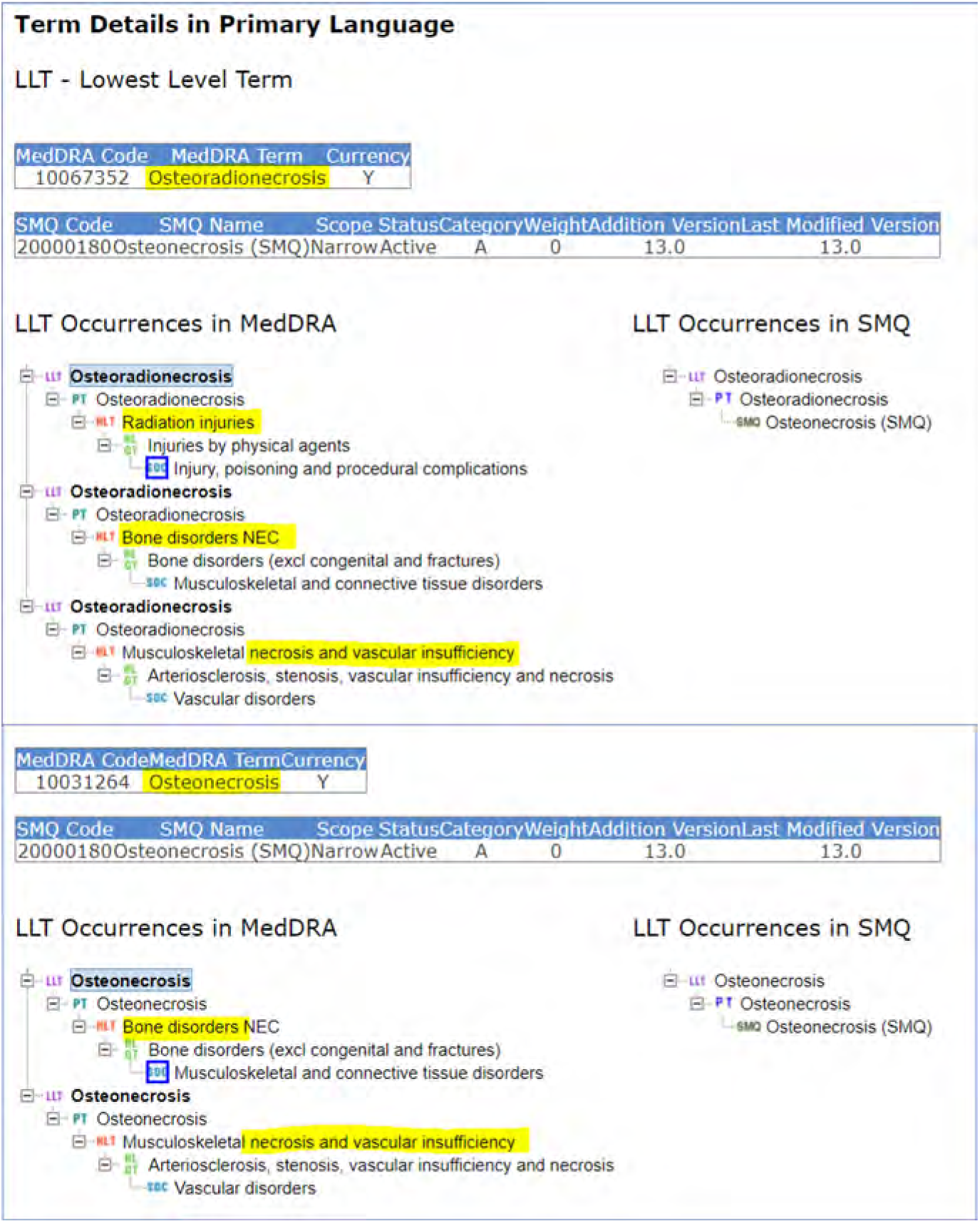
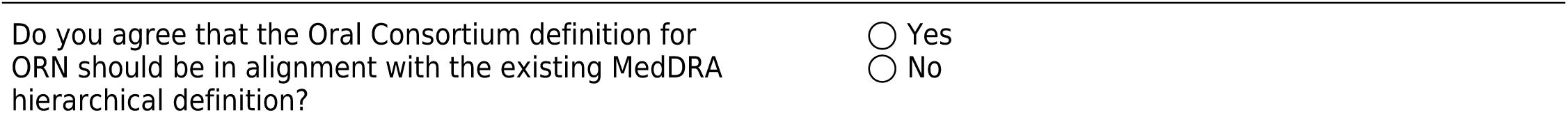

SNOMED-CT is a comprehensive standard clinical terminology/ontology with concepts (aka. “a clinical meaning identified by a unique numeric identifier”) formally defined by detailed relationships with other concepts.

Concepts and relationships, or attributes, are represented in SNOMED-CT via standardized concept diagrams, similar to the one below. For more information on diagram symbol definitions, please refer to this Diagramming Guideline: https://confluence.ihtsdotools.org/download/attachments/29951081/doc_DiagrammingGuideline_Current-en-US_INT_2 0140131.pdf?api=v2

SNOMED-CT Code Examples:

Code for osteonecrosis: 240196003 Code for osteoRADIOnecrosis: 109333005 Code for ORN of mandible: 109716001 Code for ORN of maxilla: 109715002 Key takeaway points:

SNOMED-CT allows for ‘preferred’ or ‘acceptable’ terms for the same concept such as ‘radiation necrosis of bone’ and ‘osteoradionecrosis’. All share a ‘finding site’ in a bone structure and an associated morphology (attribute) of radiation injury WITH necrosis The ‘causative agent (attribute)’ is ionizing radiation and ‘due to (attribute)’ relationship is to exposure to ionizing radiation. Figure 3: SNOMED-CT Concept Diagrams for ORN of mandible and maxilla

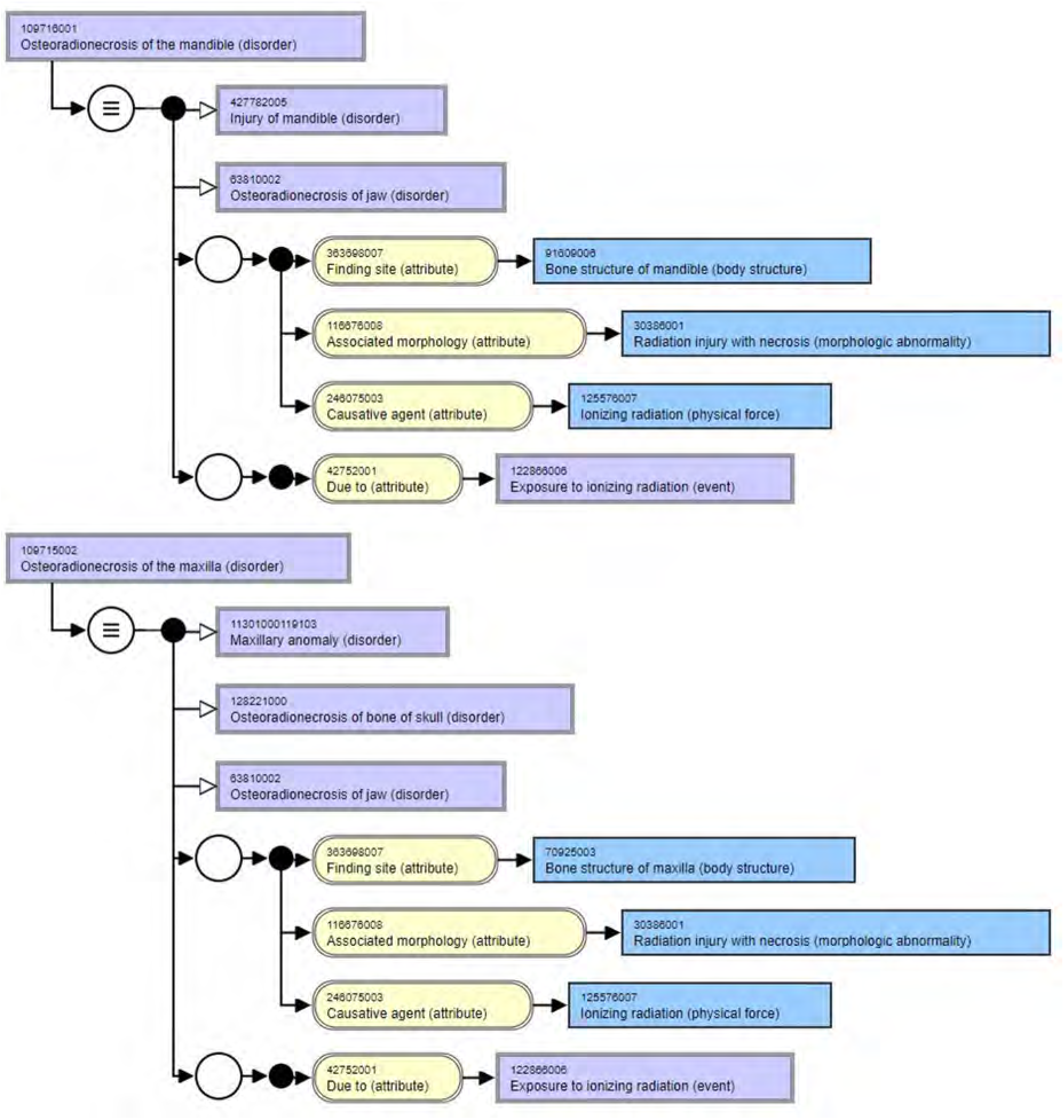
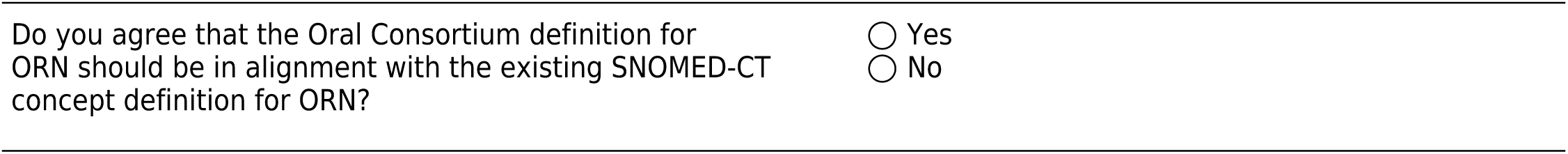

International Classification of Disease - Clinical Modficiation (ICD-CM)

ICD diagnostic codes range from 3 to 7 characters with the first character always being an alpha (i.e. letter). Longer codes reflect more specific diagnoses. See Figure 4.

ICD-10-CM Code Examples:

Code for osteonecrosis: M87.9 Code for osteonecrosis, secondary necrosis (NEC), due to drugs: M87.10 Part of the definition for osteonecrosis (M87) from icd10data.com:

Clinical Information

A disorder characterized by necrotic changes in the bone tissue due to interruption of blood supply. Most often affecting the epiphysis of the long bones, the necrotic changes result in the collapse and the destruction of the bone structure. Death of a bone or part of a bone Death of a bone or part of a bone, either atraumatic or posttraumatic. Death of bone tissue caused by loss of blood supply to the bone. Death of bone tissue due to traumatic or nontraumatic causes. Of note, the ICD-10-CM Diagnosis Code ‘M27.2’ is often used for ORN of the jaw, which broadly captures ‘inflammatory conditions of jaws’.

Reference: https://icd10cmtool.cdc.gov/?fy=FY2023&query=osteonecrosis

**Figure 4:**
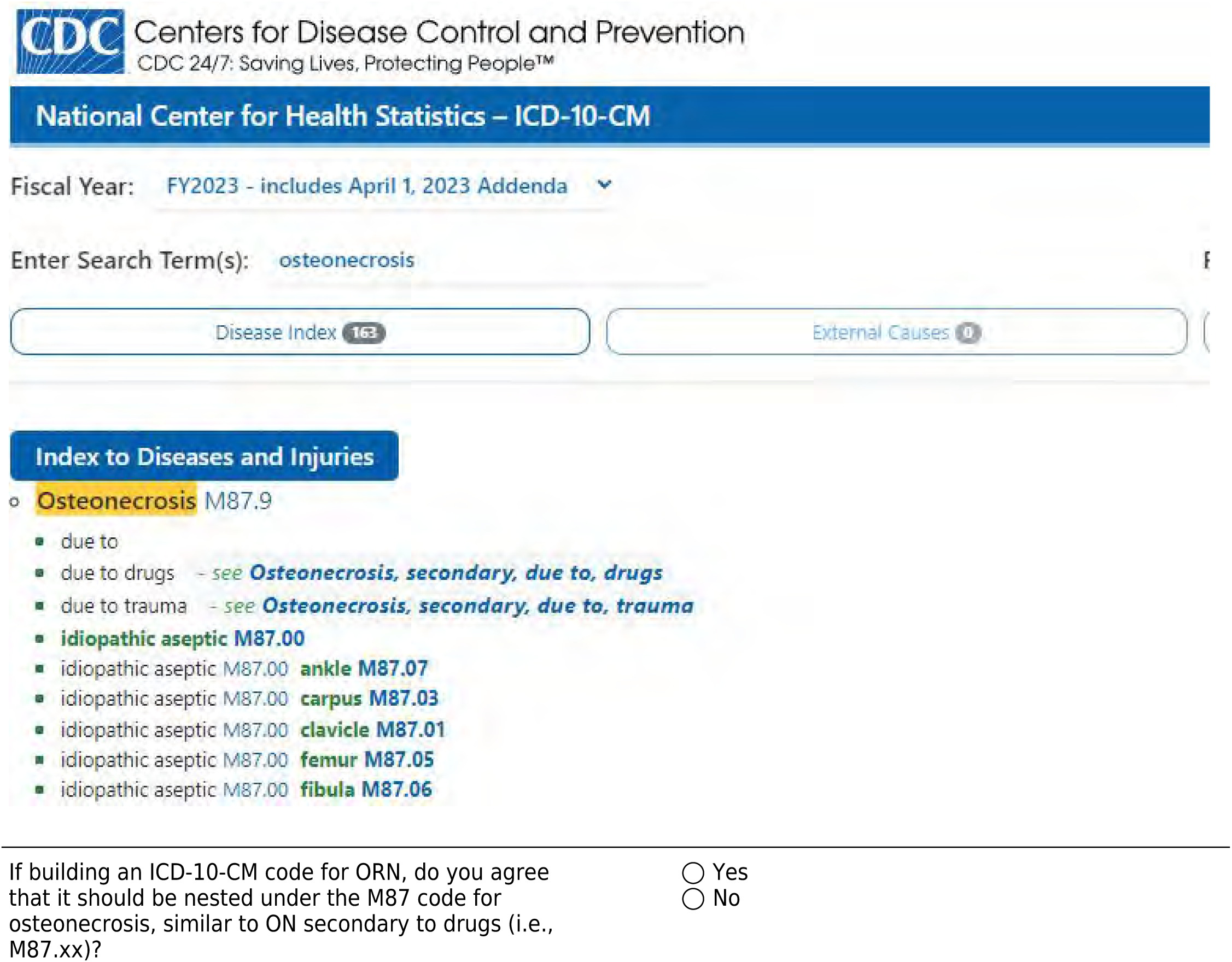
ICD-10-CM Index to ON.

### Defining (Osteo)Necrosis

As necrosis is in several of the above definitions for ORN, consensus on the term ‘necrosis’ itself is needed.

Attaining consensus on the definition of necrosis itself is important when developing a formal definition for ORN. The National Cancer Institute (NCI) Dictionary of Cancer Terms describes osteoNECROSIS as the following:

”A condition in which there is a loss of blood flow to bone tissue, which causes the bone to die. It is most common in the hips, knees, shoulders, and ankles. It may be caused by long-term use of steroid medicines, alcohol abuse, joint injuries, and certain diseases, such as cancer and arthritis. It may also occur at some point in time after cancer treatment that included methotrexate, bisphosphonates, or corticosteroids. Also called aseptic necrosis, avascular necrosis, and ischemic necrosis.”

Reference: https://www.cancer.gov/publications/dictionaries/cancer-terms/def/osteonecrosis

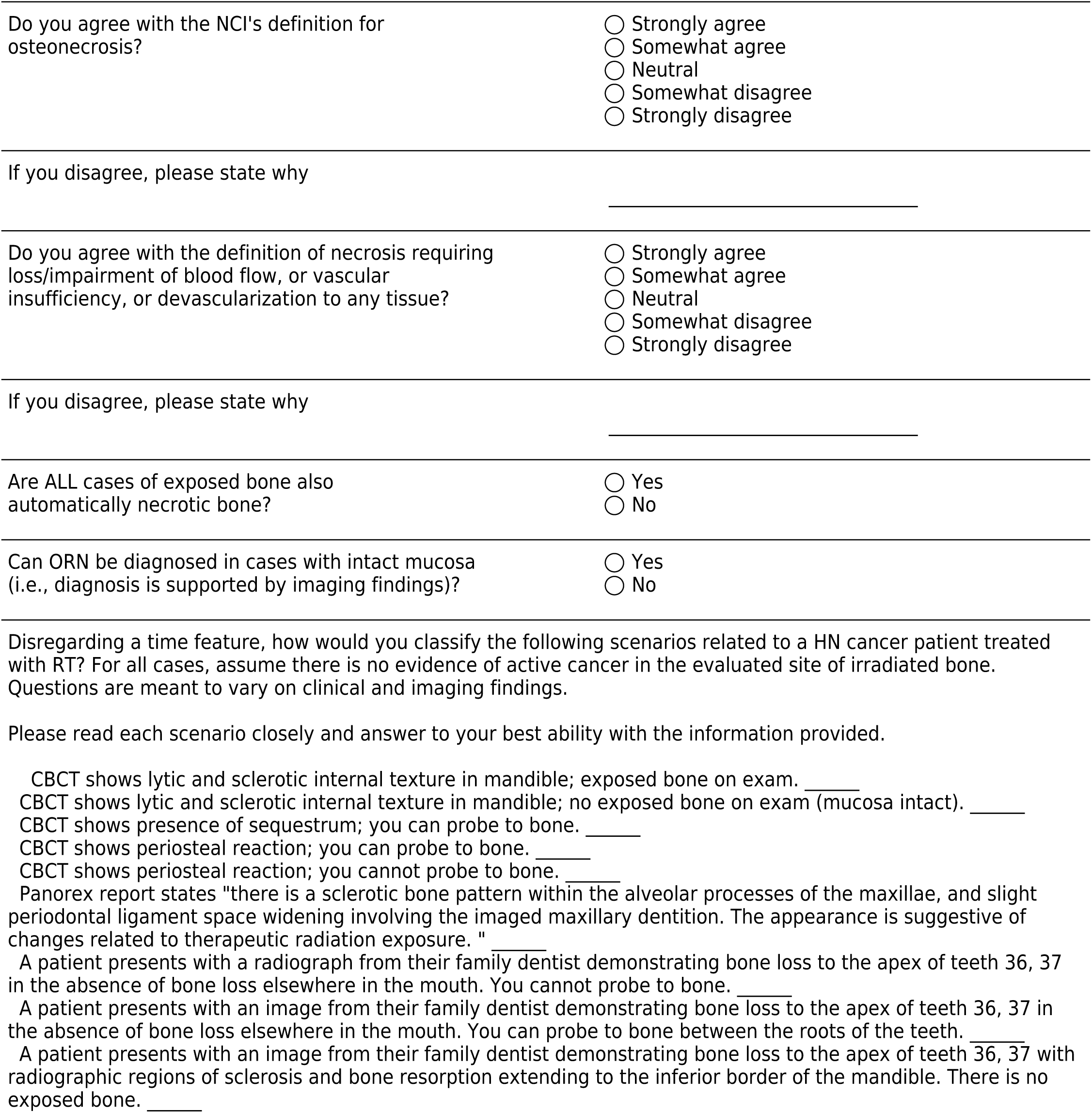
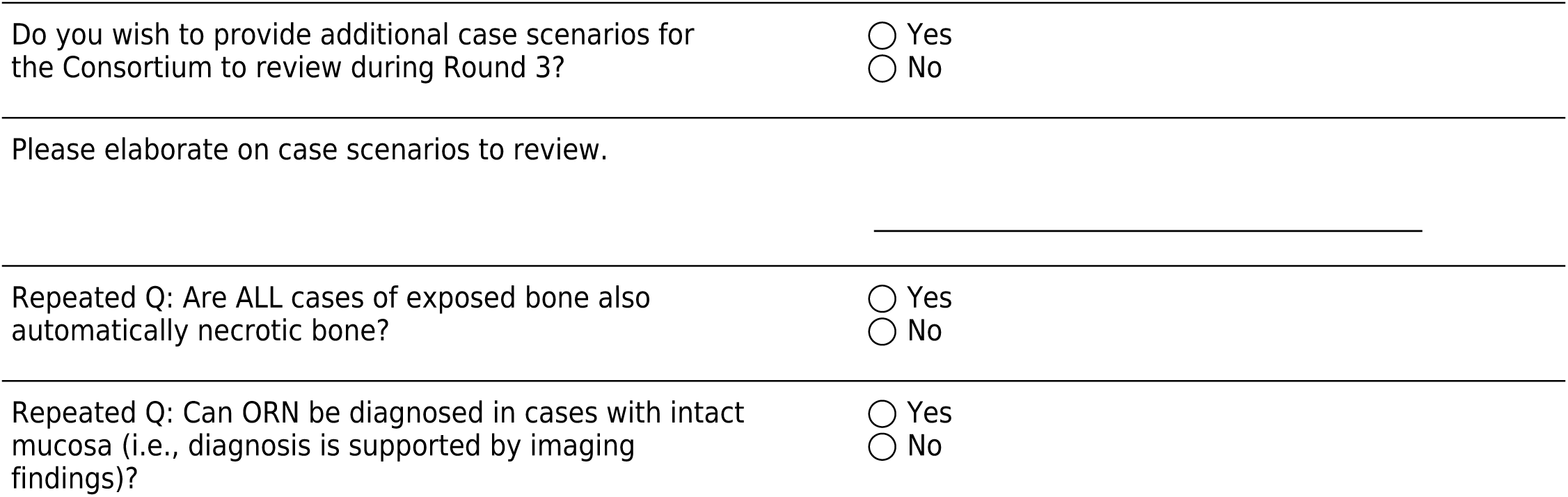

### The ‘Time Feature’ and Refining the ORN Definition

The duration of ORN, while regarded as a highly important feature, remains controversial (i.e., when to use it and/or how to define useful parameters). Despite its inclusion in 6 of the 14 ORN staging/grading systems reviewed during Round 1, there is no consensus on the explicit definition for the time feature to declare a diagnosis of ORN (present/absent).

Round 1 Summary:

Harris, Schwartz, and Karagozoglu all reported different ‘minimum’ time periods of bone exposure in an irradiated field to diagnose ORN. Disagreement with the time feature was also seen in the group’s response to Case 1 (1.5 cm of exposed bone for 2 months) whereby only 27 (46%) of members would diagnose the patient with ORN during that clinic visit. The majority (83%) agreed on diagnosing Case 2 with ORN, given a much longer time window (0.5 cm exposed bone for 7 months). When asked to provide an explicit time window (in months) for diagnosing ORN, 47% of panelists left this question blank while the remaining panelists listed 3 months (41%), 4 months (3%), or 6 months (9%). Additional considerations with regards to time:

Standardized diagnostic systems (i.e., ICD, MedDRA, SNOMED-CT, etc) do NOT include a time feature in their disease/disorder definitions. The true duration of necrotic bone (seen either clinically and/or on imaging) is difficult to measure as our observations heavily rely on the timing and frequency of patient visits (which can vary among providers). Consider the attached image scenario where clinical exams and imaging are performed every 3 months for a patient treated with RT. The asterisks represent suggested ‘minimal duration of exposed bone’ time windows after which one can ‘diagnose’ ORN. The red wording and time window represents changes occurring in between visits.

**Figure 5:**
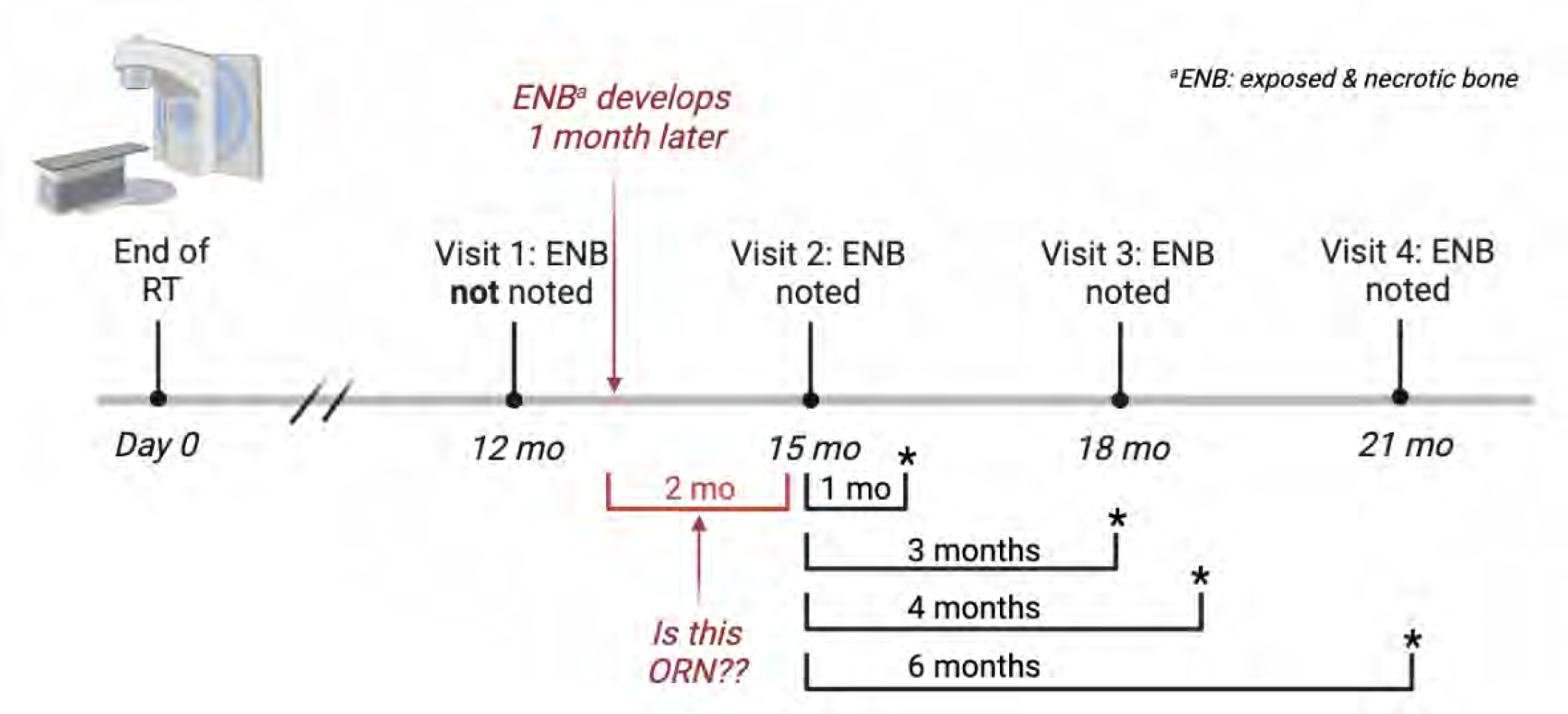
Case Scenario.

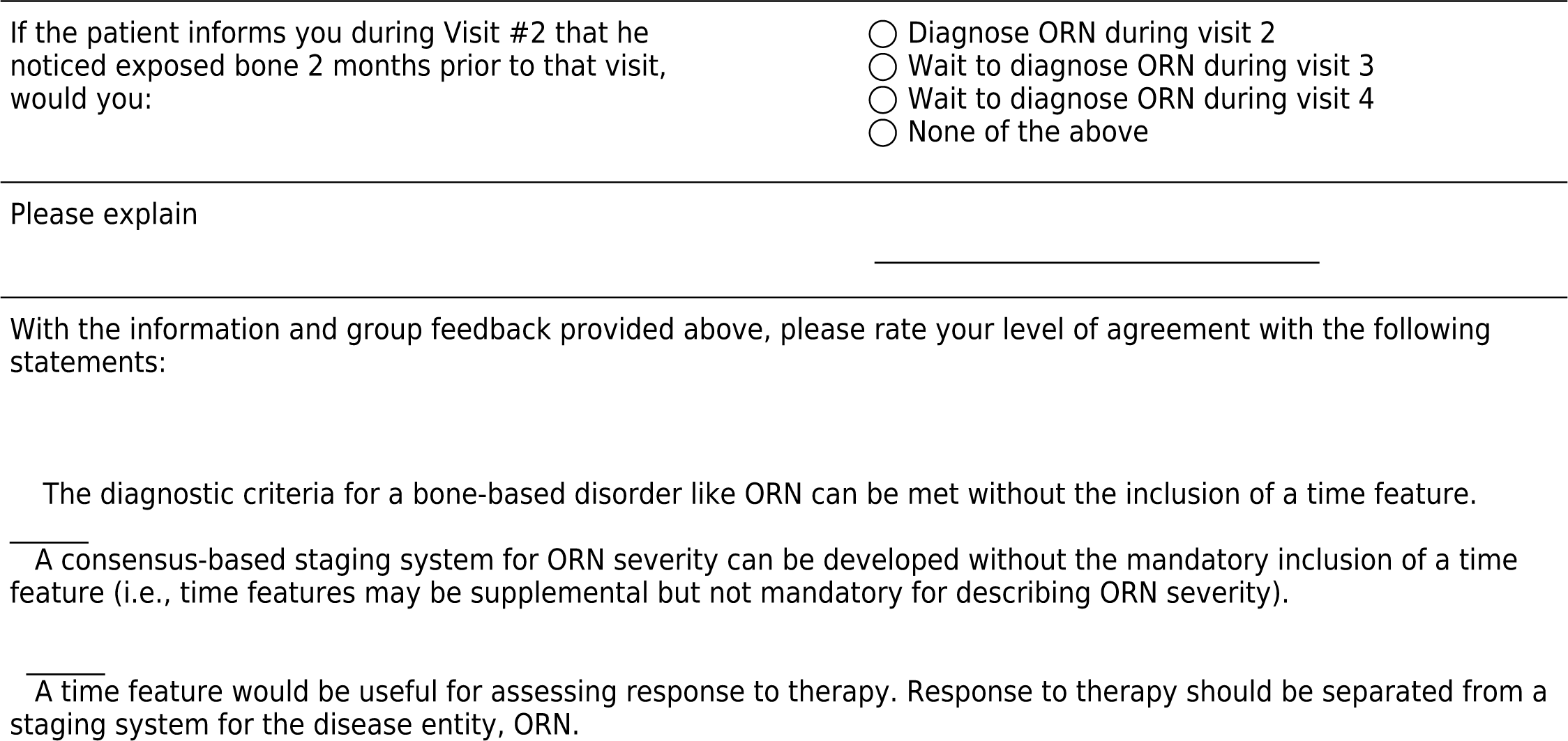
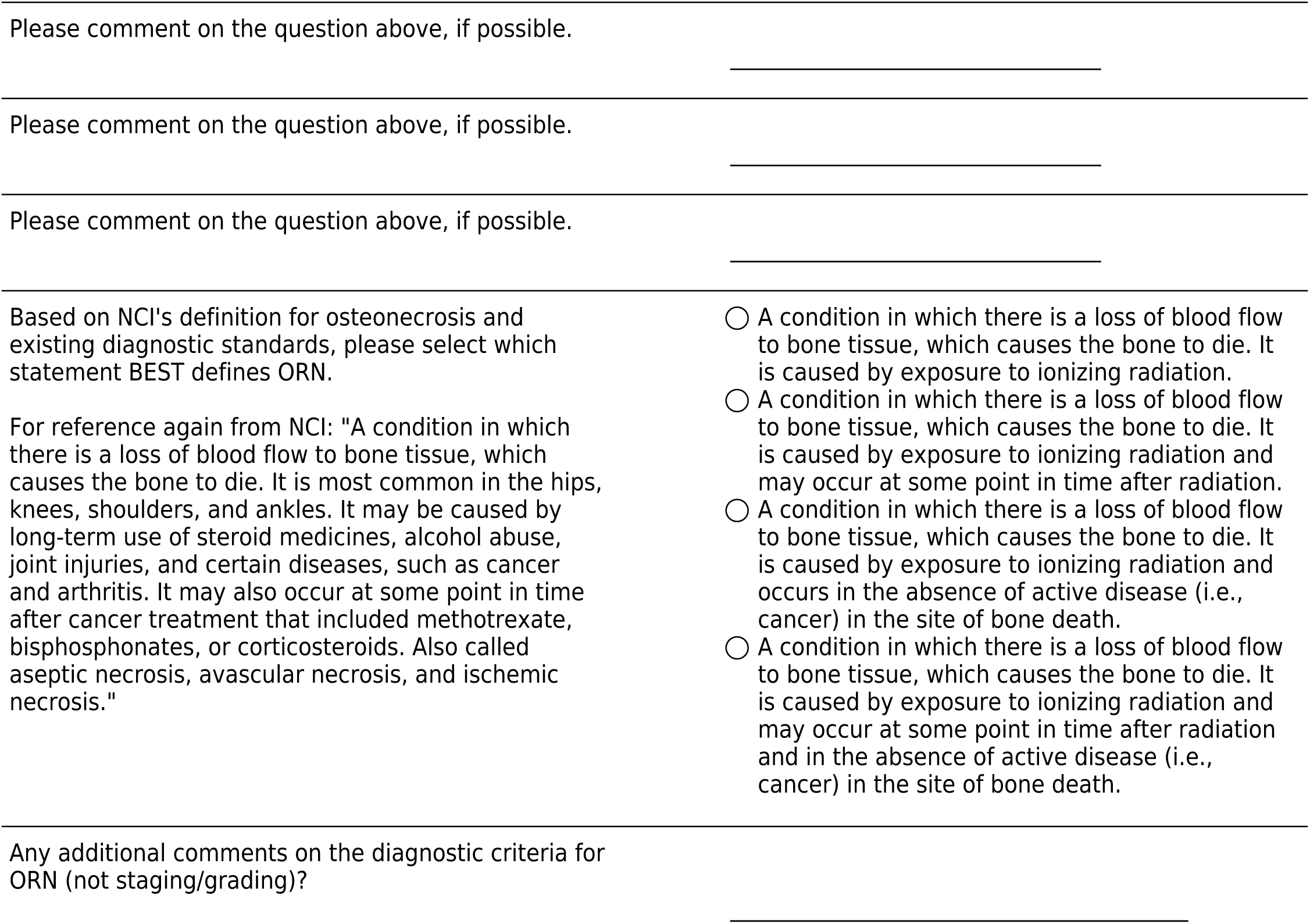

### Staging Elements for Reporting Extent and Severity of ORN

During Round 1, a total of 15 staging/grading systems were reviewed. Members were asked to 1) state personal use of each system, 2) rate the utility of the system, and 3) apply the system to categorizing 3 different case scenarios.

Personal use: The top 3 systems used in practice were: CTCAE (n=41; 70%), Notani (n=18, 32%), and Marx (n=18, 31%).

Rating of effectiveness for classifying ORN: The top-rated systems, defined as ‘somewhat/very important’, for ORN were not in the top ones for personal use and included: Shwartz & Kagan (n=28, 52%), Karagozoglu (n=24, 49%), and Morton & Simpson (n=26, 48%). CTCAE, Notani, and Marx were considered effective by 46%, 42%, and 23% of respondants, repsectively.

**Figure 6:**
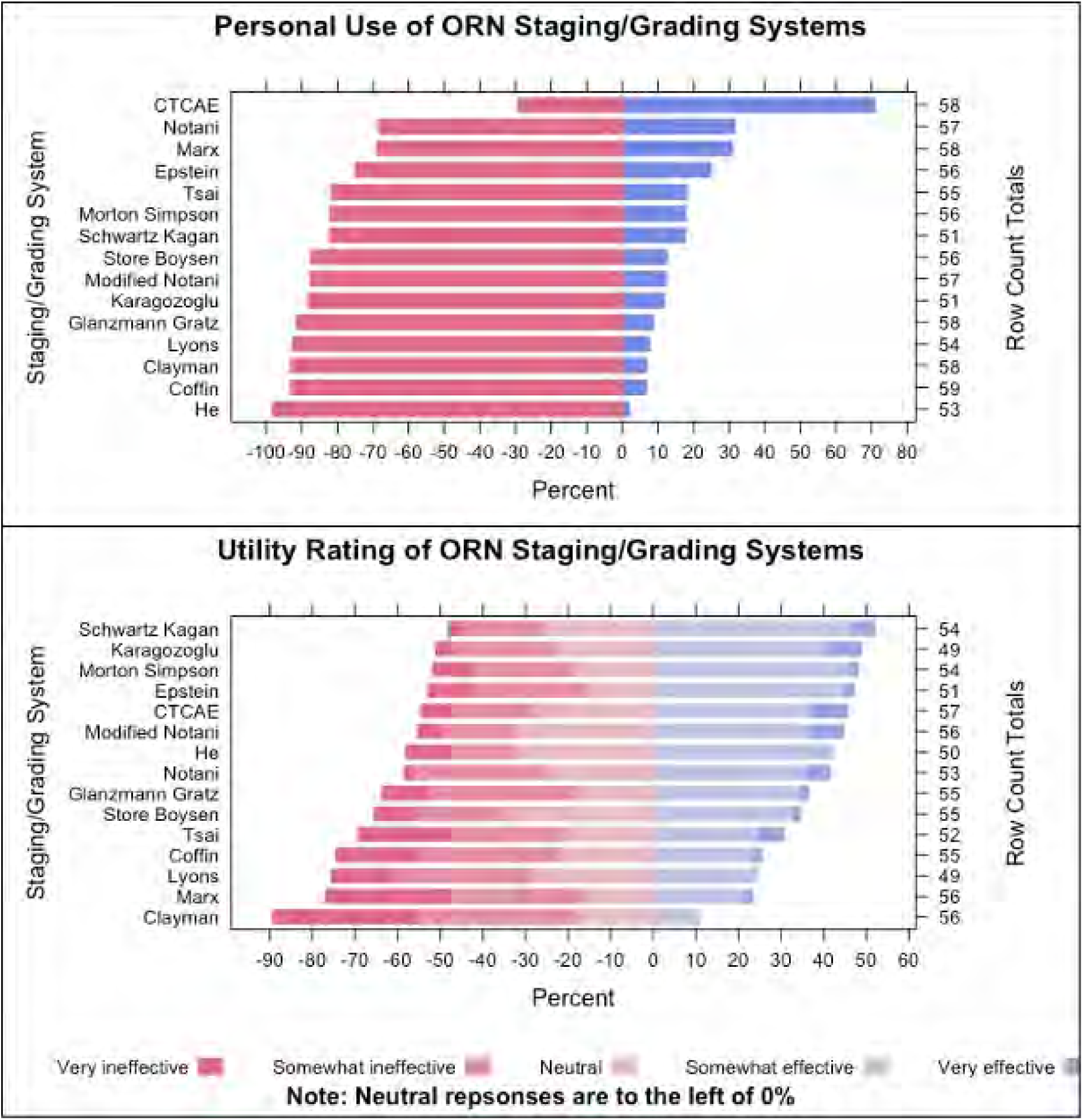
Personal Use and Effectiveness Rating of Existing Staging/Grading Systems.

Group feedback continued:

During Round 1, members were asked to categorize the following 3 cases using each staging/grading system.

Case 1: Patient with exposed bone (no measurement) not involving lower mandible, unknown duration. Pain present. Case 2: Patient with 1.2 cm exposed bone for 4 months, pain present. Case 3: Patient with 3 cm exposed bone with pathologic fracture. Unknown symptoms or duration. Fom a total of 59 responders:

The inability to classify cases was 57% for Case 1, 45% for Case 2, and 26% for Case 3. Pathological fracture (a finding in Case 3) was commonly classified as advanced ORN (major or stage III). Missing responses (i.e., no option was selected) was low at 3-4%. The distribution of case classification per system is shown in Figure 7. Overall, none of the existing systems were rated useful by more than 70% of the responders (the consensus threshold; top rated one was 52%), and the inconsistent classification of cases using these systems demonstrates an ongoing need for ORN data standardization.

Please click on the link below for Figure 7.

[Attachment: “Round1CasesbySystem.png”]

Additional grading systems recommended for review during Round 1:

In the comments section, four additional osteonecrosis / ORN systems were recommended for review. They include:

MRONJ (Medication-Related Osteonecrosis of the Jaws) LENT SOMA Scale Princess Margaret Cancer Center ORN Scoring System RTOG CTC (Common Toxicity Criteria) Summary:

MRONJ defines a non-exposed bone variant (stage 0), explicitly states ‘exposed and necrotic bone or fistula’ for advanced stages, and incorporates imaging findings. LENT-SOMA uses extent of exposed bone (2cm) or presence of limited sequestration or fracture for upgrading PMH defines grades as loss of mucosa with exposed bone requiring particular therapies for increasing time. Pathologic bone fracture is a grade 4. RTOG CTC: Vague and symptom based. In alignment with CTCAE Please review them in the attached file.

[Attachment: “ADDITIONAL GRADING SYSTEMS FOR REVIEW.docx”]

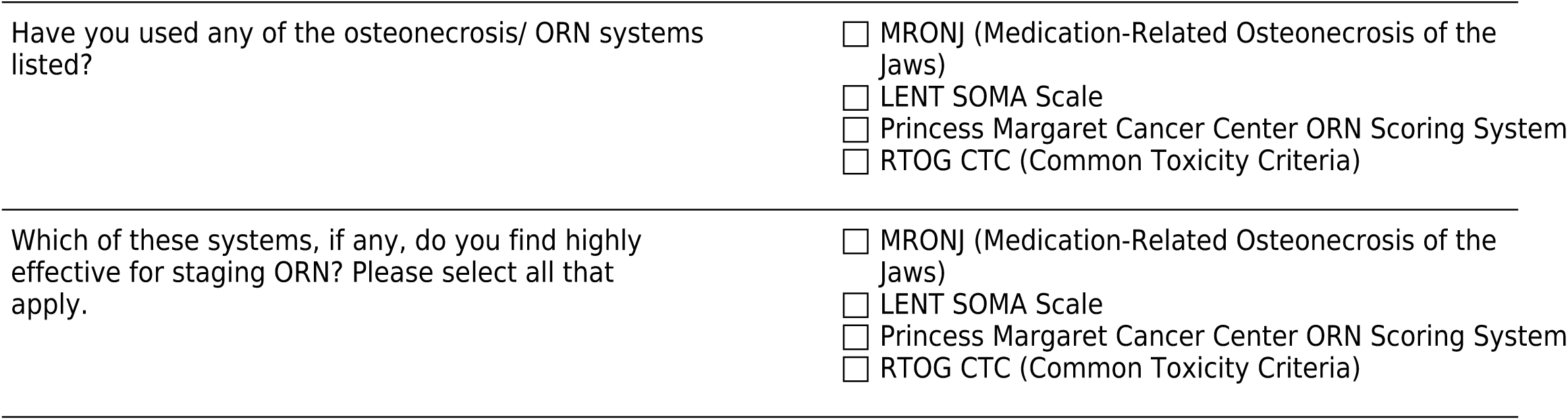

Clarification on the widely used Common Terminology Criteria for Adverse Events (CTCAE)

CTCAE is often used by the group (70%) but it is unclear if this is for staging (reporting ORN extent/severity) and/or toxicity grading (AEs after therapy).

For reference, the NCI states the following about CTCAE: it is a “descriptive terminology which can be utilized for Adverse Event (AE) reporting. A grading (severity) scale is provided for each AE term. An Adverse Event (AE) is any unfavorable and unintended sign (including an abnormal laboratory finding), symptom, or disease temporarily associated with the use of a medical treatment or procedure that may or may not be considered related to the medical treatment or procedure. An AE is a term that is a unique representation of a specific event used for medical documentation and scientific analyses.”

Reference: https://ctep.cancer.gov/protocoldevelopment/electronic_applications/docs/ctcae_v5_quick_reference_5x7.pdf

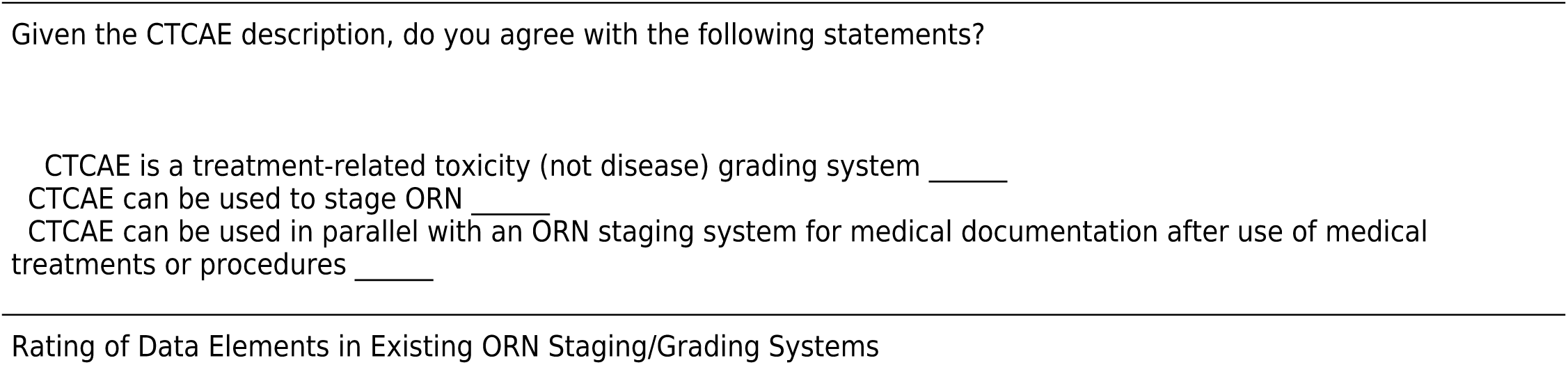

Rating of Data Elements in Existing ORN Staging/Grading Systems

During Round 1, the group was asked to rate the level of importance for each element extracted from all reviewed staging and grading systems. The results are shown in Figure 8.

The only elements (n=3) that were rated as ‘somewhat/very important’ by 100% of the group were all bone-related and included pathological fracture, exposed bone, and extent of exposed bone. Elements found to be of least importance when staging ORN included treatment (HBO, surgery, conservative therapy) and response to therapy (with the exception of persistent bone exposure). Comments from members: I think treatment can be considered separate from the classification as that may depend on what the patient or health care provider feels most comfortable with. A staging system based on primarily clinical factors is more useful, can consider including need for antibiotic treatment. If including response to therapy, would consider response to vitamin E/Trental as that defines conservative management vs surgical management. The classification should be independent on the response to treatment. There is limited evidence on the ideal treatment for ORN. It is therefore generally not appropriate to include these in a classification. The descriptors of extent of ORN should be sufficiently broad to be applicable to a wide range of cases (including ‘pre-ORN’) whilst also evaluating progression. Applying just to the mandible also limits external validity for ORN affecting maxilla or free flap.

**Figure 8.**
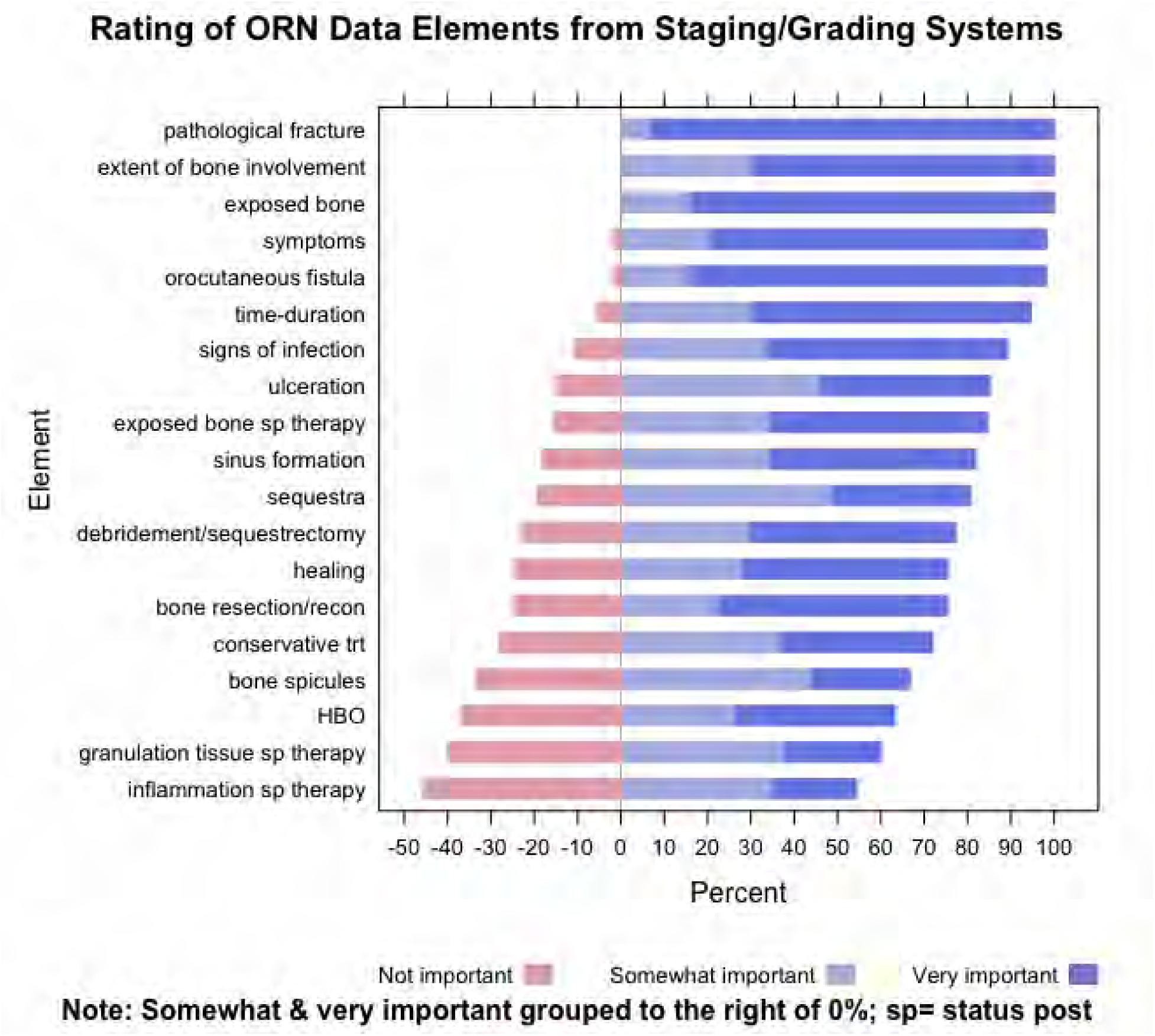
Rating of ORN Data Elements from Staging/Grading Systems.

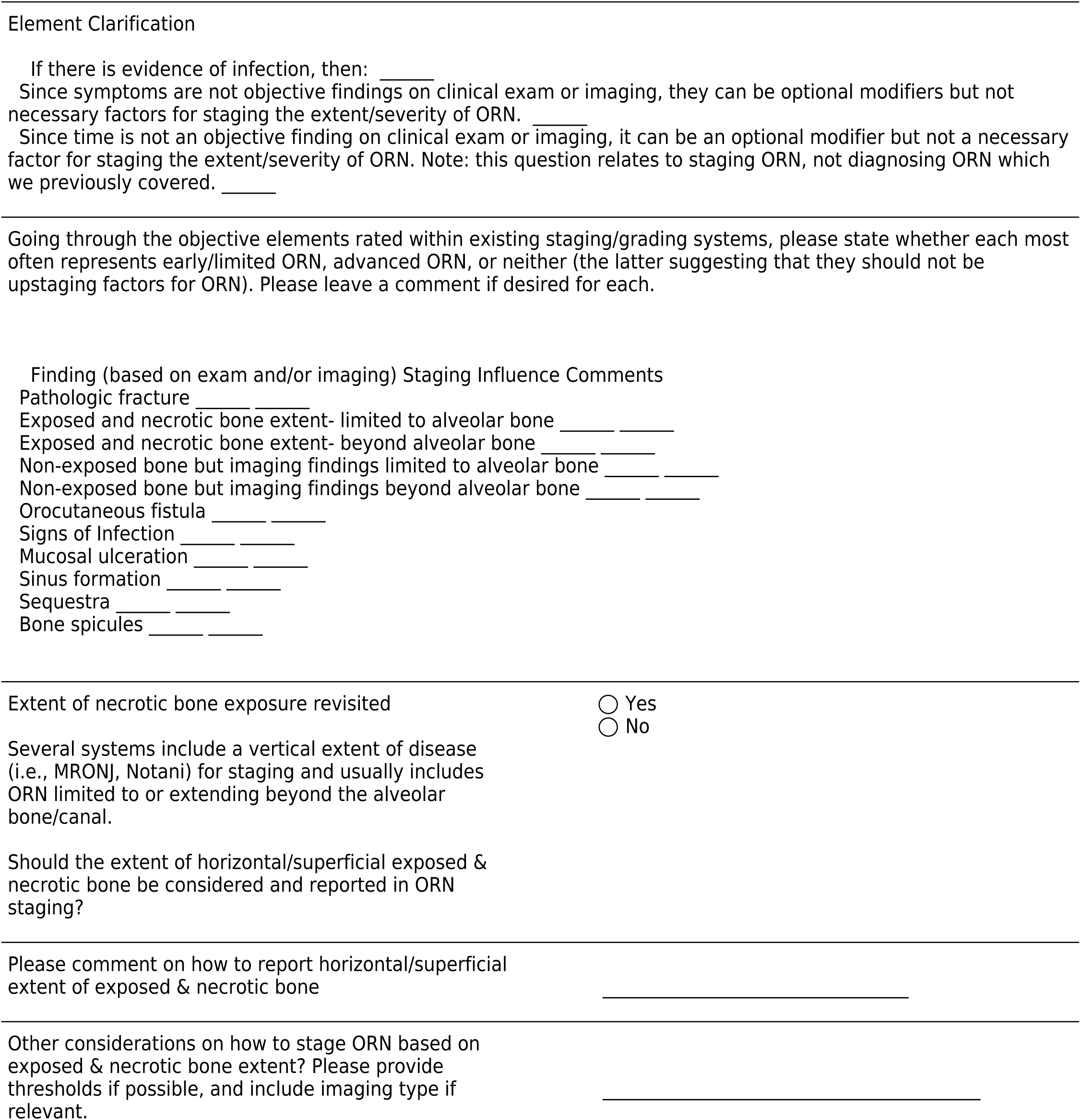

### RADMAP: Radiation Dose Mapping to an Odontogram

A total of 55 panelists of the Consortium provided a response to the RADMAP (Radiation Odontogram) Section of Round 1. Consensus was defined as 70% of more agreement and percentages are based on the total count of responses per question.

The following table is a breakdown of the dental specialists within the oral consortium.

For reference to Round 1, the panel reviewed the following figure of a potential RADMAP report which displayed patient, cancer, and treatment information as well as the radiation odontogram and a snapshot of the RT plan.

Findings

The current layout for patient, cancer, and treatment is clear according to 85% (n=44) of the group. After reviewing comments left in Round 1, panelists pointed out a need to include chemotherapy history for the treatment information portion as well as any teeth missing or extracted on the RADMAP visual for each patient. In addition, panelists also made note of general material to add onto the patient information section.

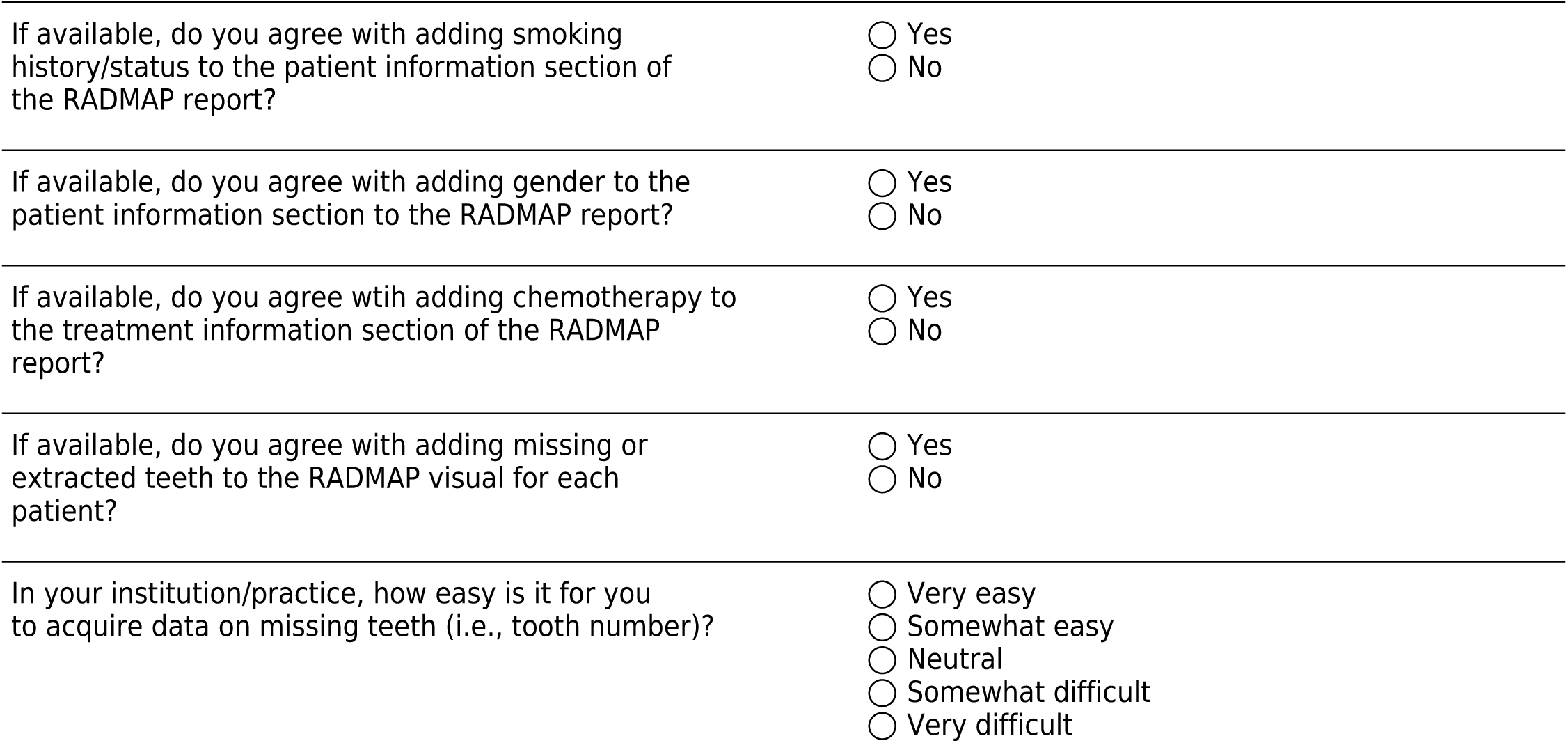

Radiation Dose Heat Maps

Group Feedback from Round 1:

89% (n=48) of the group agreed that a heat map over an odontogram (i.e., radiation odontogram) is a clinically useful visualization. Action: We will proceed with the build of a radiation odontogram For cases with missing teeth, 96% (n=50) agreed that visualizing dose data to segmented/ tooth-bearing regions of the mandible and/or maxilla is helpful. When faxing or printing in only in black and white, 70% (n=38) agreed that grayscale colors may impede providers from accurately interpreting radiation dose distribution. Action: We will provide an accompanying table with dose data. 80% (n=43) of the group responded that they are very confident or somewhat confident in their ability to interpret radiation doses on the radiation odontogram. The estimated mean and median dose delivered to tooth #31 was 14 Gy and 13 Gy, respectively (acceptable with one outlier of dmax ∼35Gy). Action: No need for additional educational tools with RADMAP report

When comparing options A and B, 59% preferred option A whereas 32% (n=17) chose option B. A minority of 9% (n=5) opted for neither option stating that dose data to each tooth can be presented in the format of a table. Action: We will proceed with Option A as the background of the radiation odontogram.

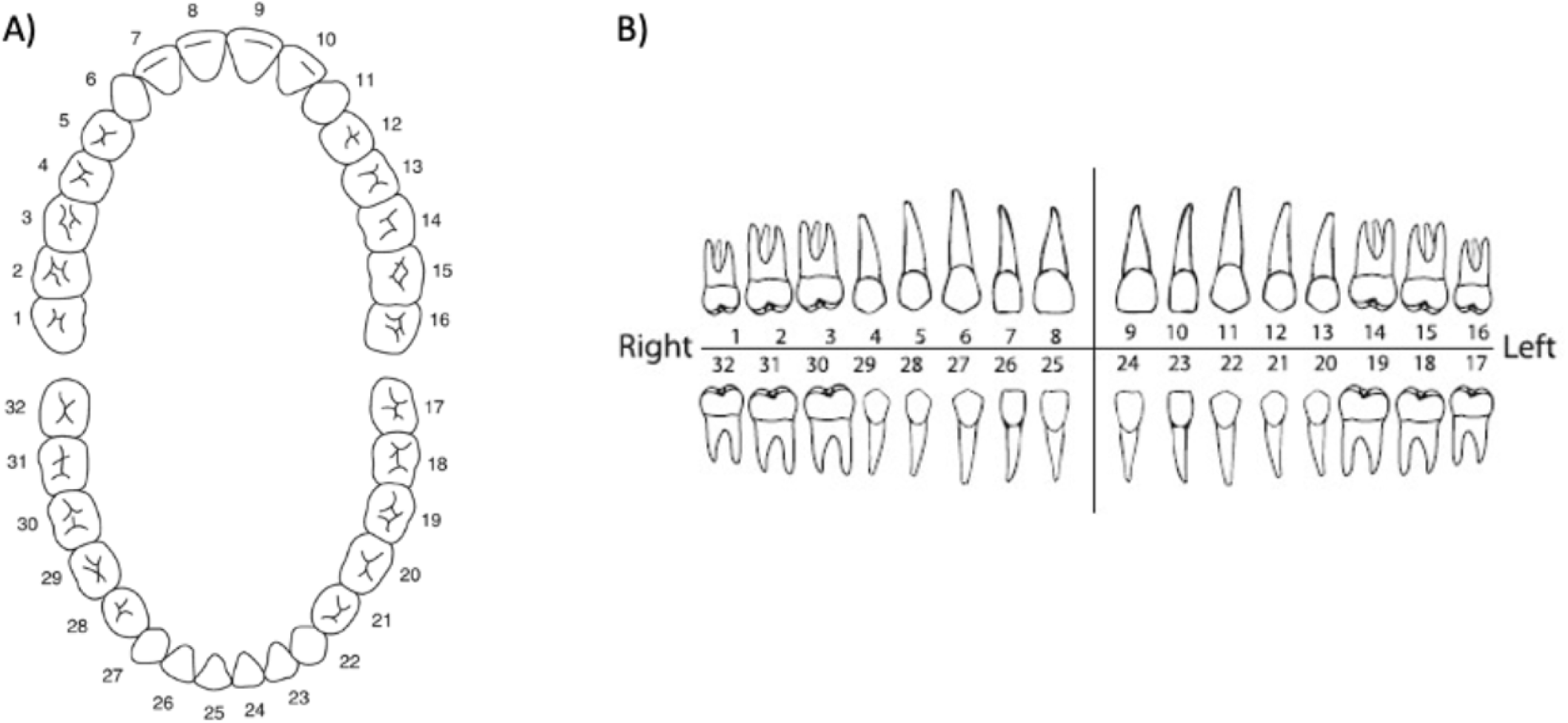

Evaluating dose data from an RT plan snapshot

96% (n=51) of the group reported seeing this type of radiation treatment plan snapshot before 72% (n=39) routinely request or review radiation therapy plans for evaluating post-RT care. When asked to evaluate the radiation dose delivered to tooth #8 (labeled A on image above), the mean, median and mode were 1000 cGy, 1000 cGy, 2000 cGy. Reported range: 0 to 2200 cGy.

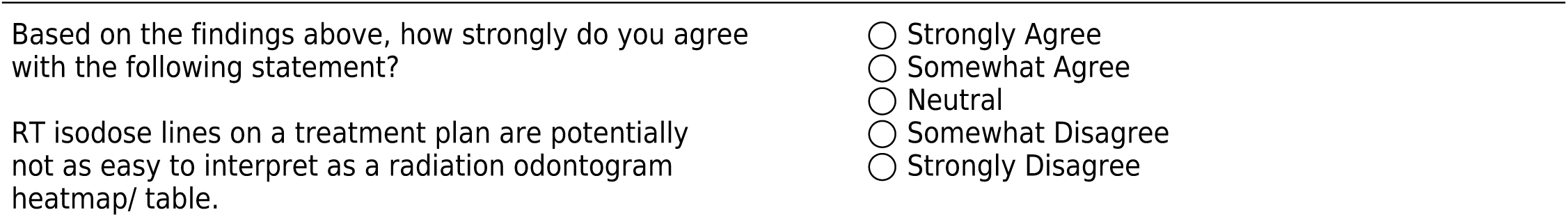
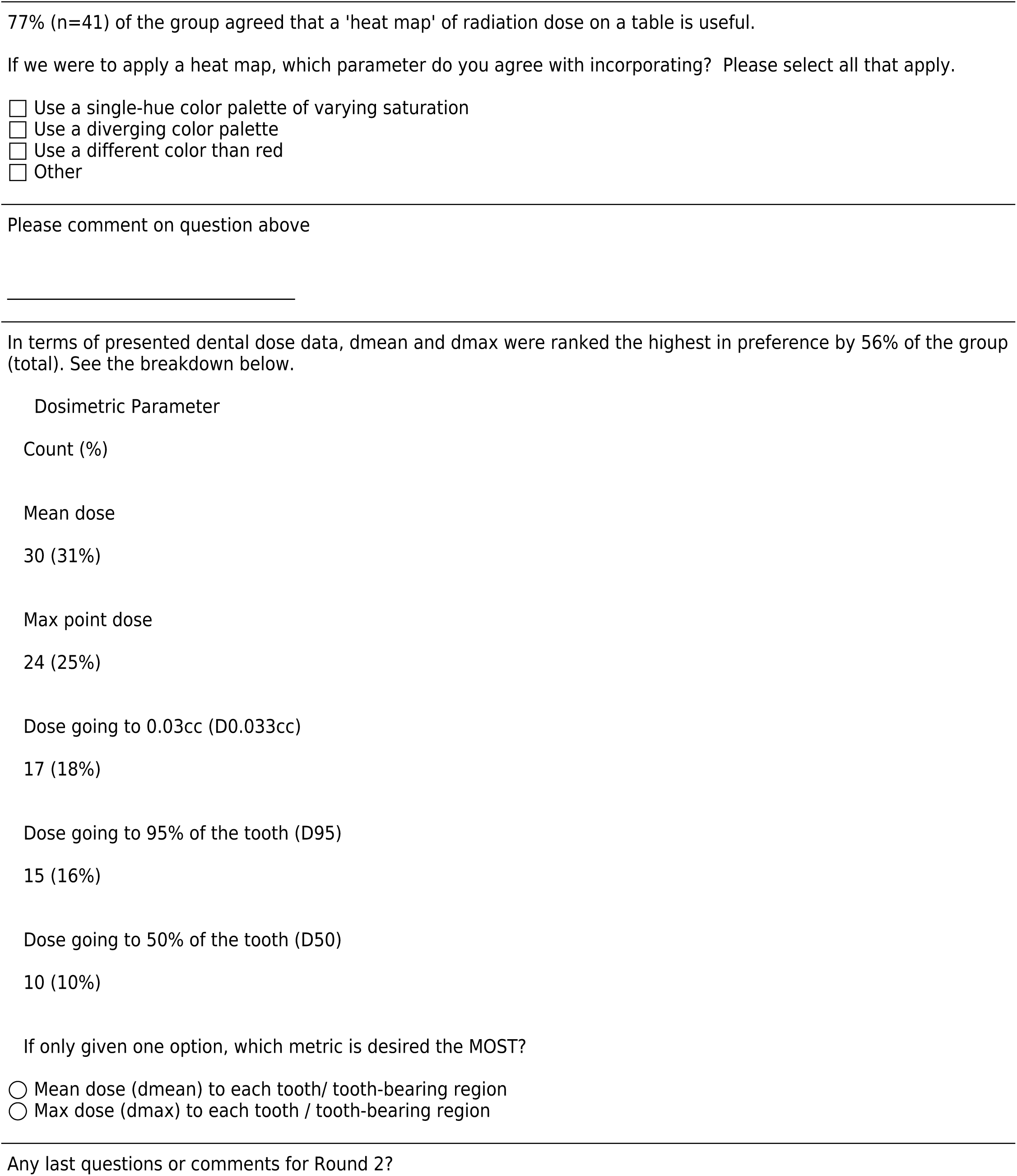

## Intro

### ORAL Consortium Round 3 Survey

This round of the ORAL Consortium Delphi study will build upon the prior round by providing group feedback on the definition of osteoradionecrosis (ORN), the diagnostic criteria for ORN, and the utility and categorization of clinical and radiographic features associated with ORN.

The goal of Round 3 is to focus on the nuances of the definition of ORN as well as classification criteria (clinical and/or radiographic) for ORN staging. Any topics that have met consensus based on the pre-defined consensus threshold of 70% will be reported in blue font.

## ORN Definition

### The Definition of ORN

In Round 2, 83% (43/52) and 85% (45/53) of panelists agreed that the ORAL Consortium’s definition for ORN should align with existing MedDRA and SNOMED-CT ORN definitions, respectively, and 80% (48/60) strongly/somewhat agreed with NCI’s definition for osteonecrosis. 87% (52/60) strongly/somewhat agreed that the term ‘vascular insufficiency’ or ‘loss/impairment of blood flow’ should be included in definitions of necrosis (i.e., ORN).

To review these definitions again, click on the links below.

[MedDRA](https://mdanderson.co1.qualtrics.com/ControlPanel/Graphic.php?IM=IM_EFn1R50uEqXc1A9)

[SNOMED-CT](https://mdanderson.co1.qualtrics.com/ControlPanel/Graphic.php?IM=IM_gTpKJ4hX59n7BUX)

NCI

Consensus: The Consortium’s definition for ORN will reflect features in these existing definitions, including:

1. Bone disorder
2. Radiation injury and/or caused by ionizing radiation
3. Loss of blood flow or vascular insufficiency AND findings of bone death/necrosis

### Time Feature

To refine the nuances of time and its relevance as a diagnostic feature for ORN, the case below was presented to the panel along with a question of when they would diagnose the patient with ORN. 79% (44/56) of panelists would diagnose this patient during visit 2 (the first time observing features of exposed and necrotic bone) while 0 panelists would wait 6 months to diagnose during visit 4.

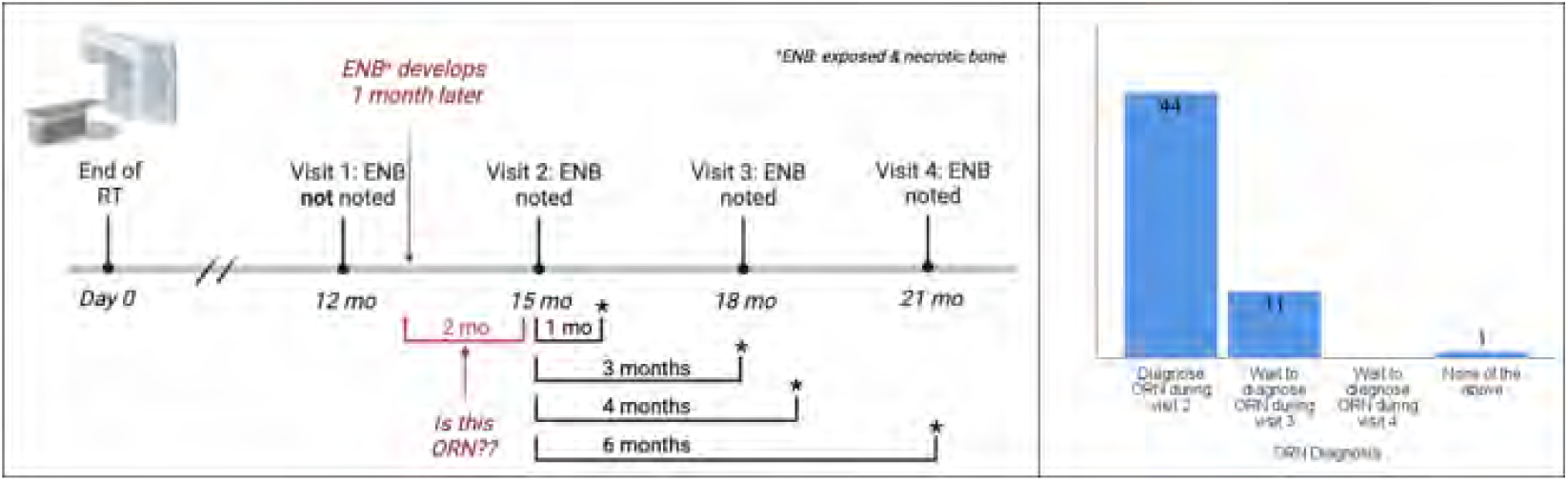

A time feature is not present in existing diagnostic codes for ORN, and 70% of the panel strongly/somewhat agreed that a diagnosis of ORN could be met without a time feature.

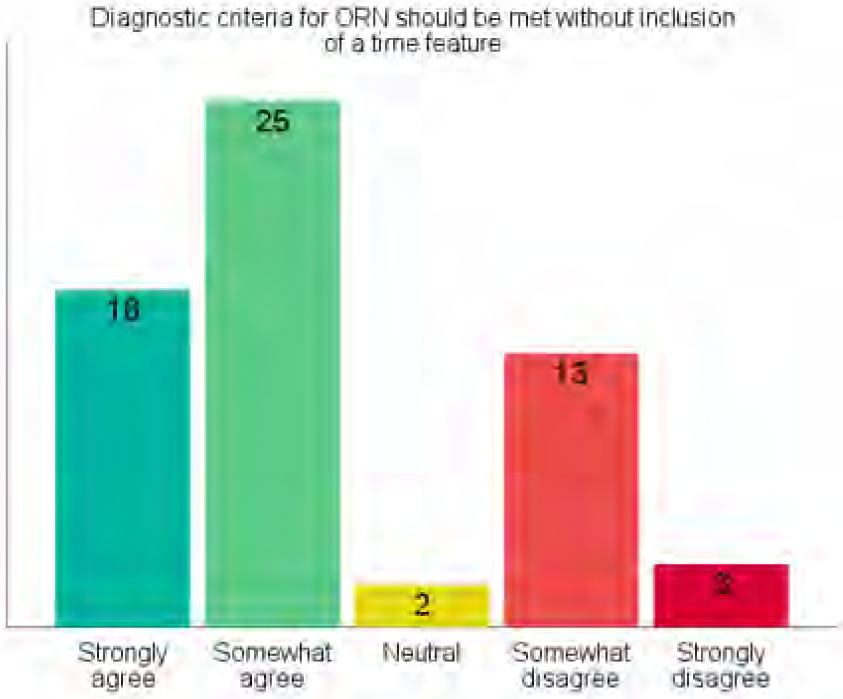

Consensus: While potentially valuable to report and reflect the duration of non-healing changes observed in bone after RT, time is not a defining diagnostic feature for ORN.

### Consensus Statements

- 87% do not consider all cases of exposed bone to automatically also be necrotic bone (i.e., minor bone spicules)
- 93% agreed that a diagnosis of ORN can be made with intact mucosa

Consensus: Not all cases of exposed bone are automatically considered to be necrotic bone.

Consensus: A <u>diagnosis</u> of ORN can be made in a patient treated with RT and with intact mucosa (i.e., no bone exposure) if there is supporting radiographic evidence of bone death/necrosis.

### Working ORN Definition

The working definition for ORN selected by 73% (41/56) of the panel during Round 2 is:

A condition in which there is a loss of blood flow to bone tissue which causes the bone to die. It is caused by ionizing radiation and may occur at some point in time after radiation and in the absence of active disease (i.e., cancer) in the site of bone death.

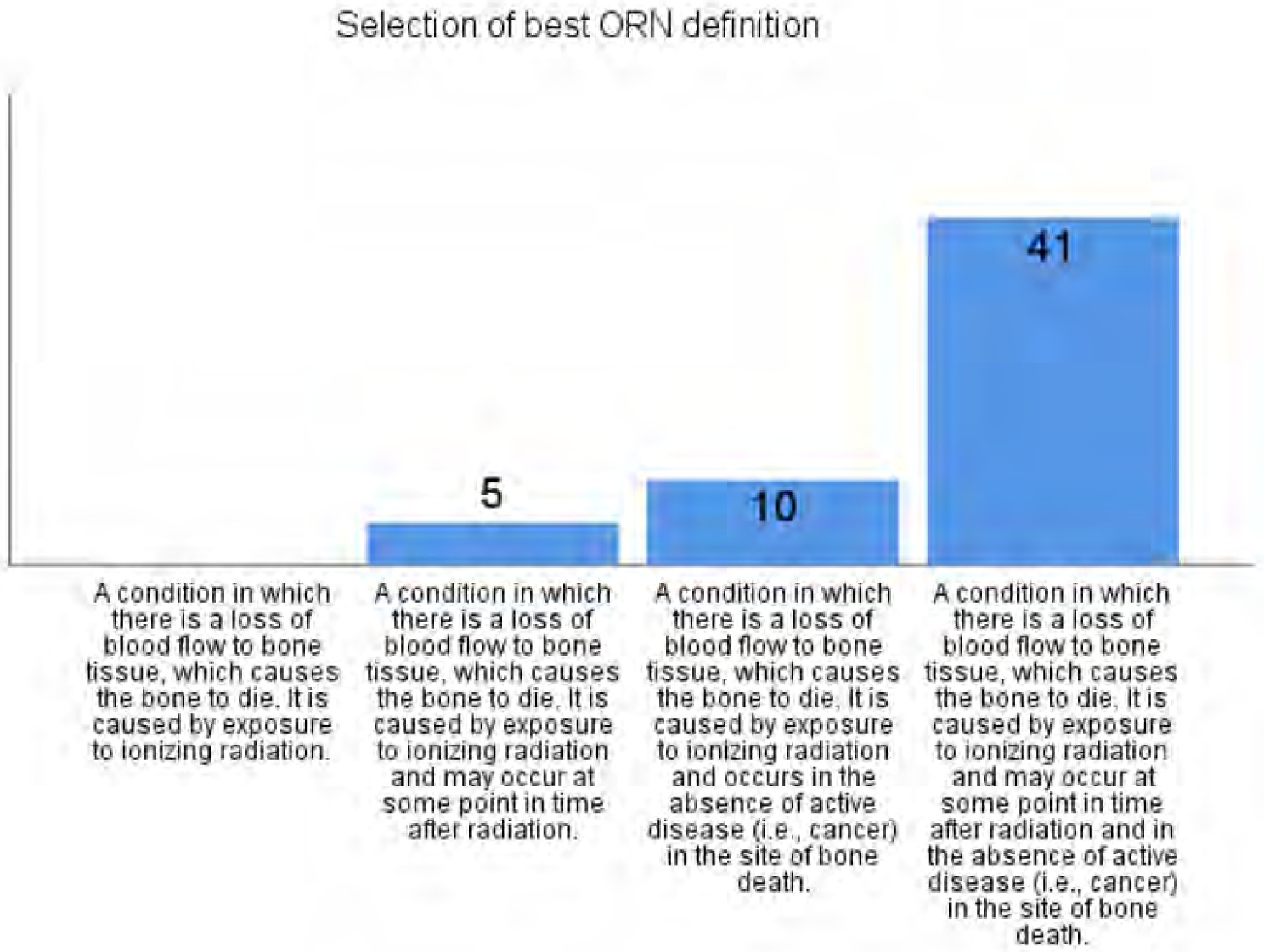

Comments by panelists (and comments by moderators) are listed below. Select any you agree should <u>definitely be considered</u> for addition in the official ORN definition.

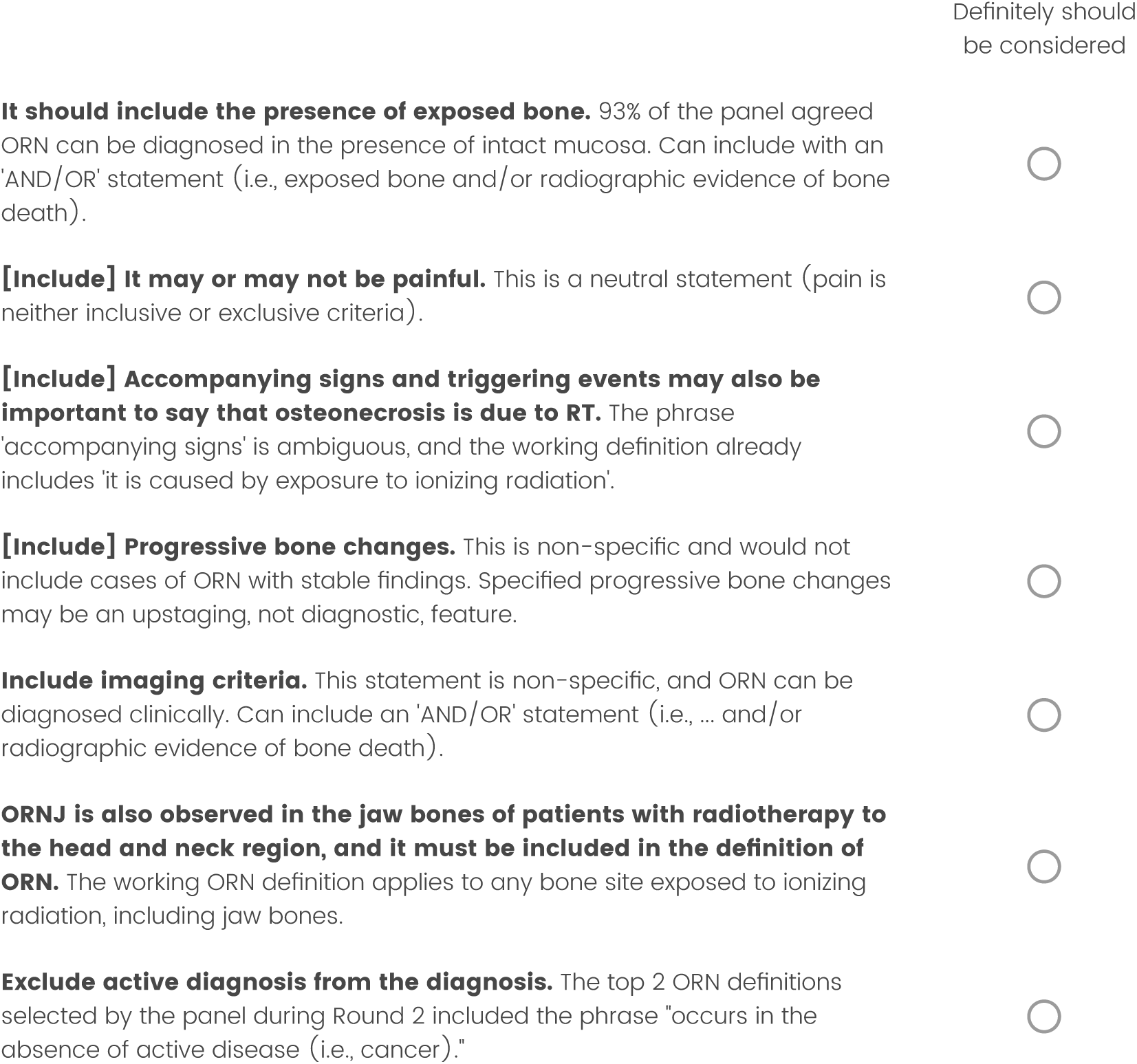

Revisions to the working ORN definition are shown below. Choose the best and second best definition and drag them to the appropriate box on the right (only one per box).

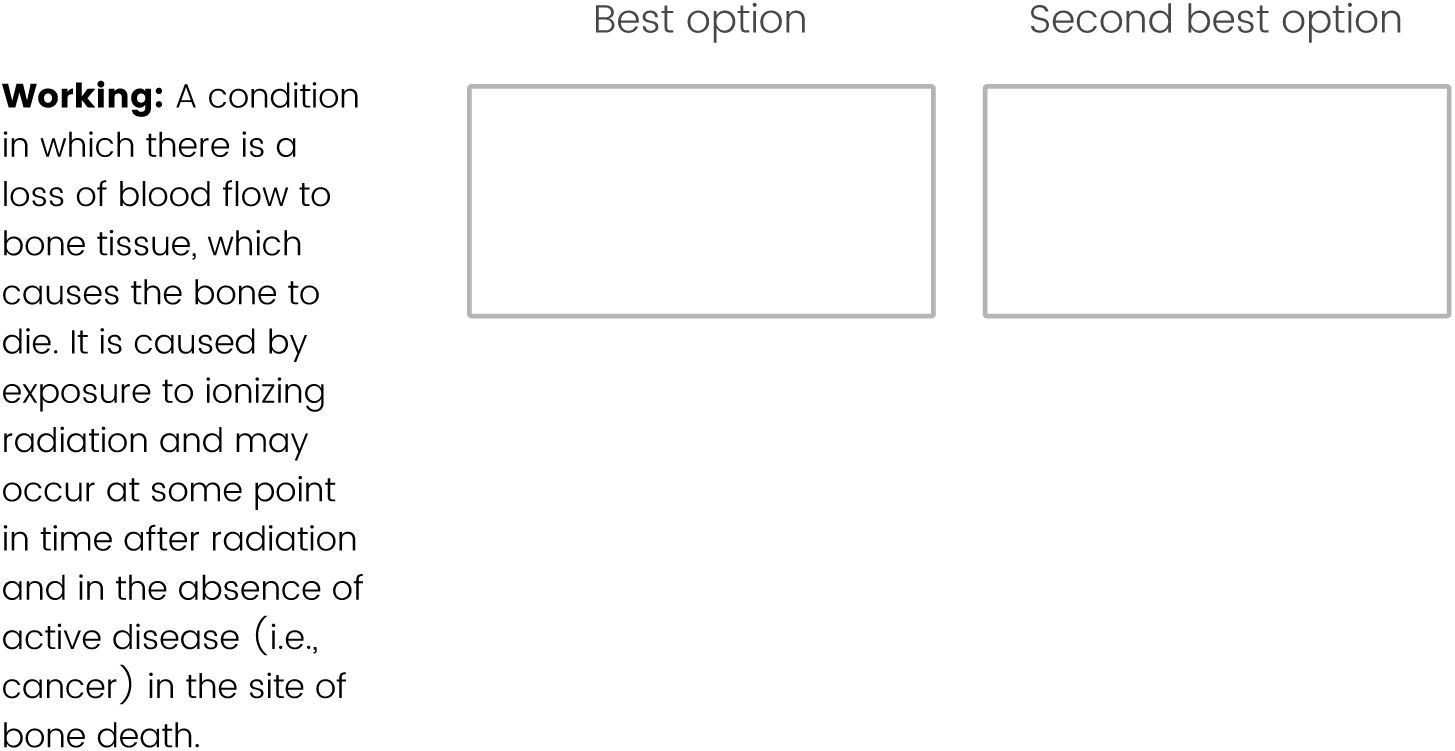
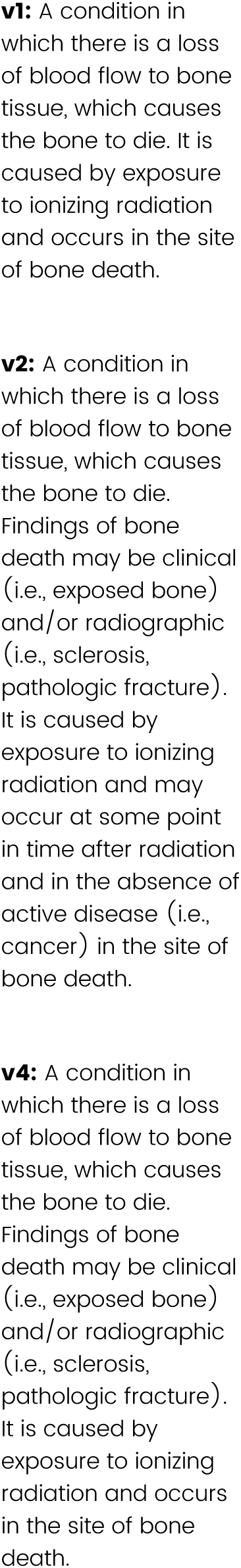

## Staging Features

### Consensus on CTCAE

CTCAE is the most commonly used grading system for ORN by the panel but was only rated 4th in terms of utility (Round 1). Three questions were asked for clarification on how to use CTCAE.

1. CTCAE can be used to stage ORN. Only 52% agreed.
2. CTCAE is a toxicity (not disease grading system). High consensus with 90% agreeing.
3. CTCAE can be used in parallel with an ORN staging system after use of treatments. High consensus with 95% agreeing.

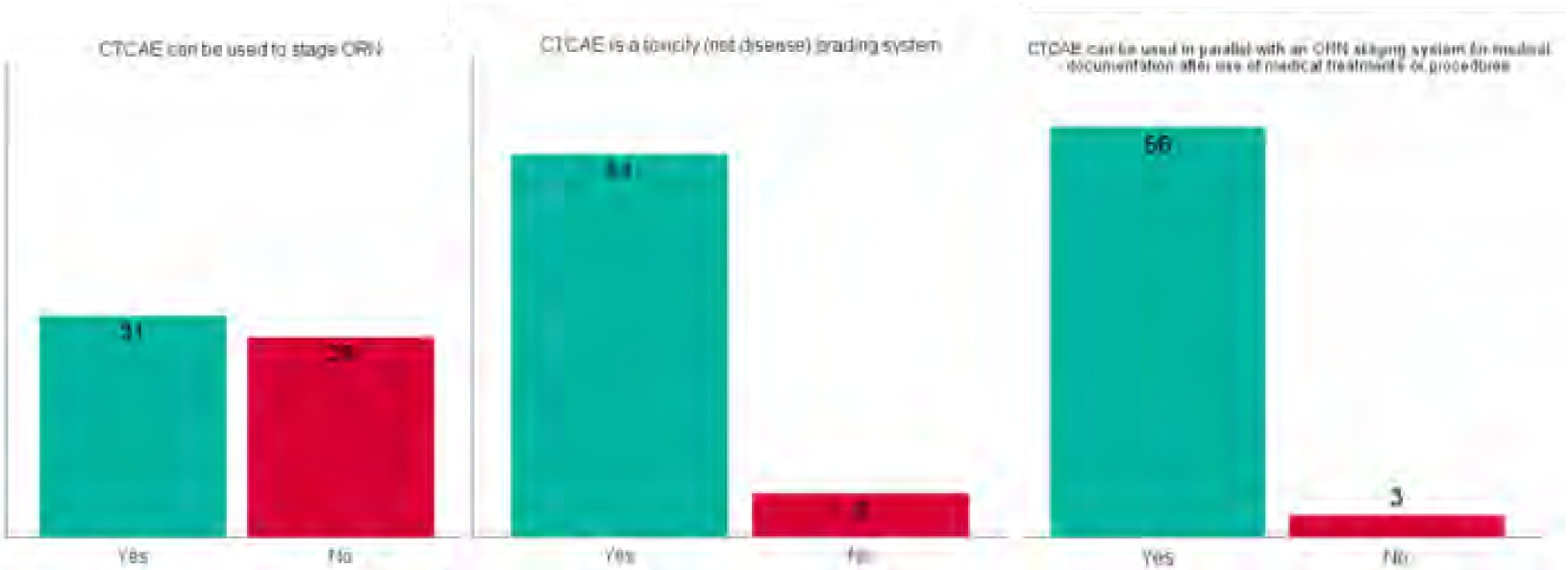

Consensus: CTCAE is a valuable toxicity grading system that should be used in parallel with, but not replace, an ORN staging system.

The following items focus on features relevant to staging the severity of ORN once it has been diagnosed.

### Time Feature and Staging

Two questions related to time and its relevance as a staging feature for ORN were asked during Round 2. In the first question, 68% (40/59) agreed that a staging system for ORN should be developed without a mandatory inclusion of a time feature. When rephrased to state that a time feature could be an optional modifier but not a necessary factor, 83% (47/57) agreed.

As far as reporting time for assessing response to therapy (considered to be distinct from reporting time as a staging feature for ORN), 85% (49/58) agreed.

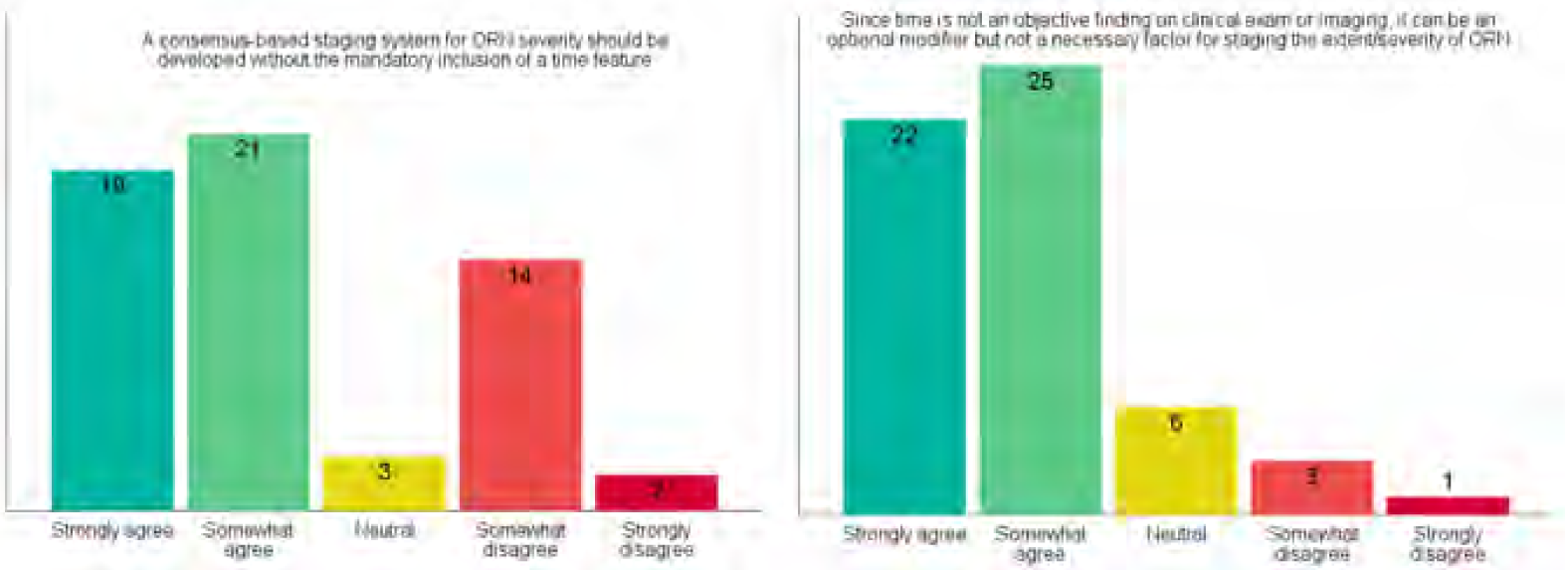

Consensus: A time feature is not necessary for staging ORN. However, reporting time is still beneficial for reporting the duration of observed ORN or response to therapy.

### Symptoms and Staging

Symptoms such as pain are often reported in patients with ORN. For the purposes of staging the severity of ORN, 72% (41/57) of the panel agreed that symptoms are not necessary features.

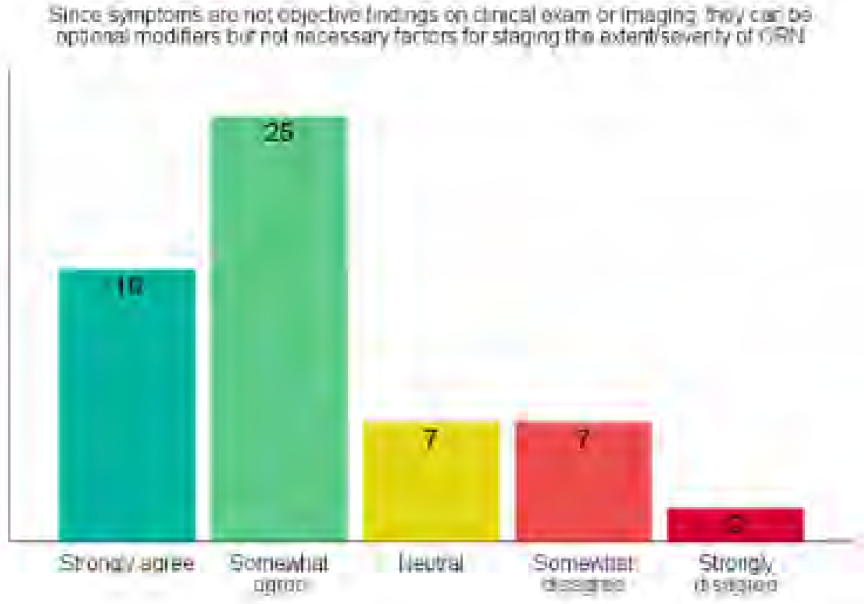

Consensus: Symptoms are not necessary for staging ORN. They can be reported as additional information but will not be considered as an upstaging feature.

## Case Reviews

### Case Review Summary

Several cases regarding a head & neck cancer patient treated with RT (with no evidence of cancer during surveillance) were reviewed during Round 2. Distribution of percent responses are shown per case.

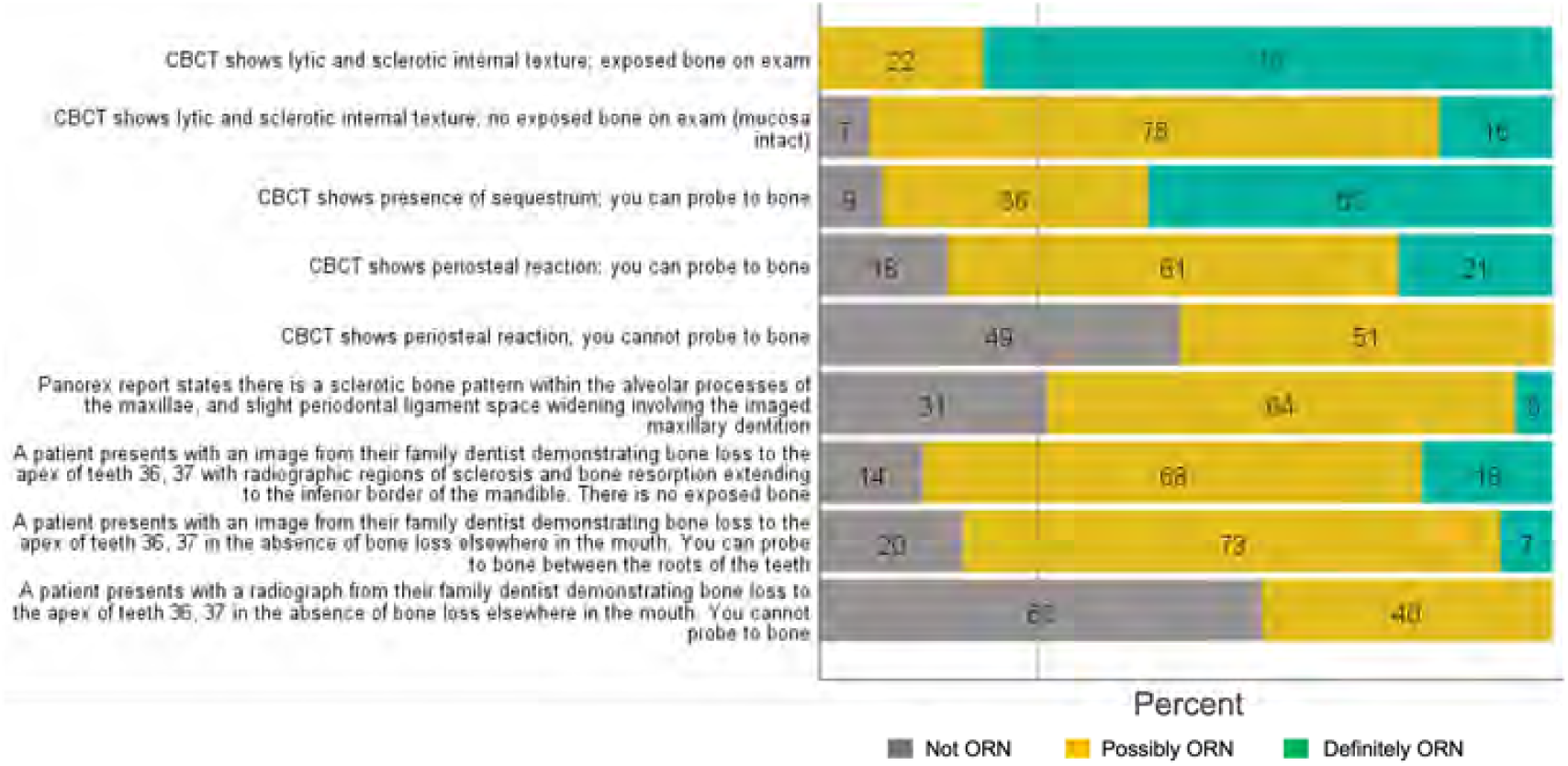

Reformatting cases based on clinical and imaging findings is shown below. Minor variations to the same case are shaded in the same color. For example, exposed bone with lytic or sclerotic changes was considered ORN by 78% of the group whereas the same imaging findings with a clinical finding of intact mucosa (i.e., non-exposed bone) were rated as possible ORN (?ORN, 78%) or definitely ORN (16%).

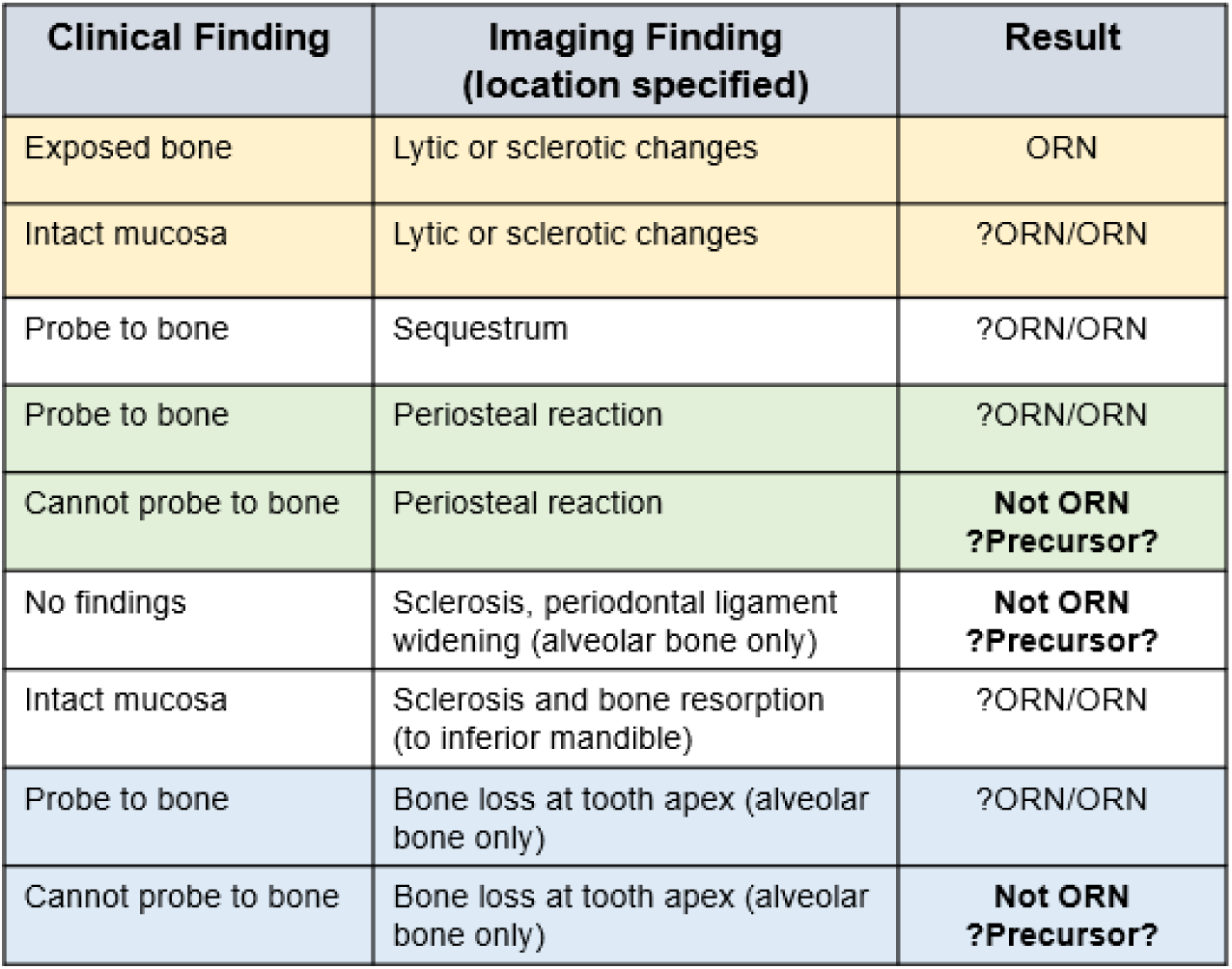

### Early vs. Advanced Stage ORN

Another series of scenarios or specific features were reviewed and classified as ‘not needed for ORN staging’, ‘early / limited ORN’, or ‘advanced ORN’. Group responses are shown below.

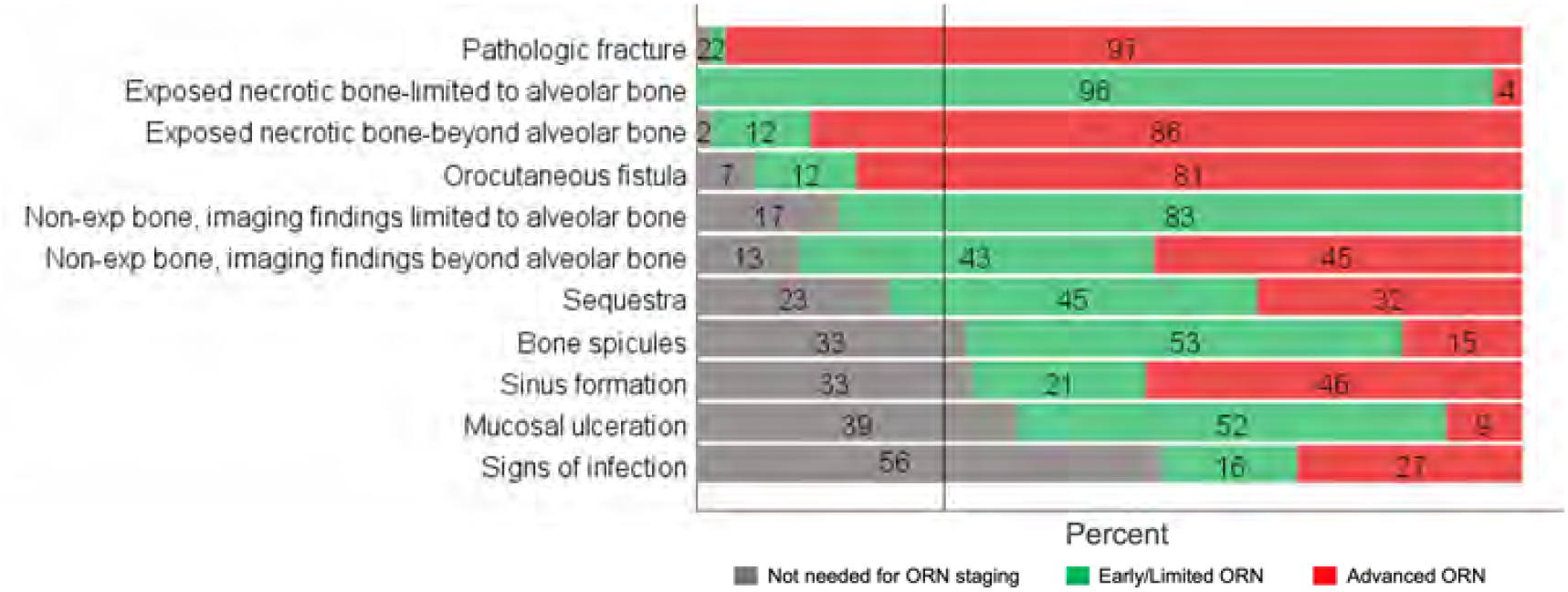

**Features and feature combinations meeting consensus are:**

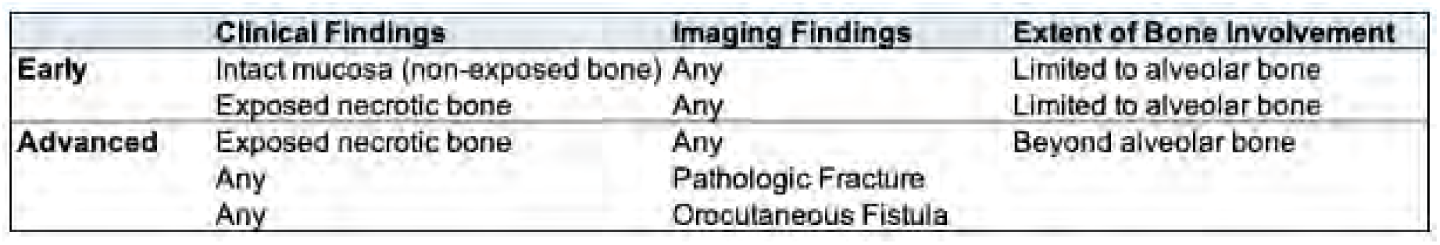

These items have been classified as advanced ORN. For further clarification, drag and drop <u>every item</u> to an intermediate or advanced stage box. If you consider all to be of same severity, drag them all to the advanced stage box.

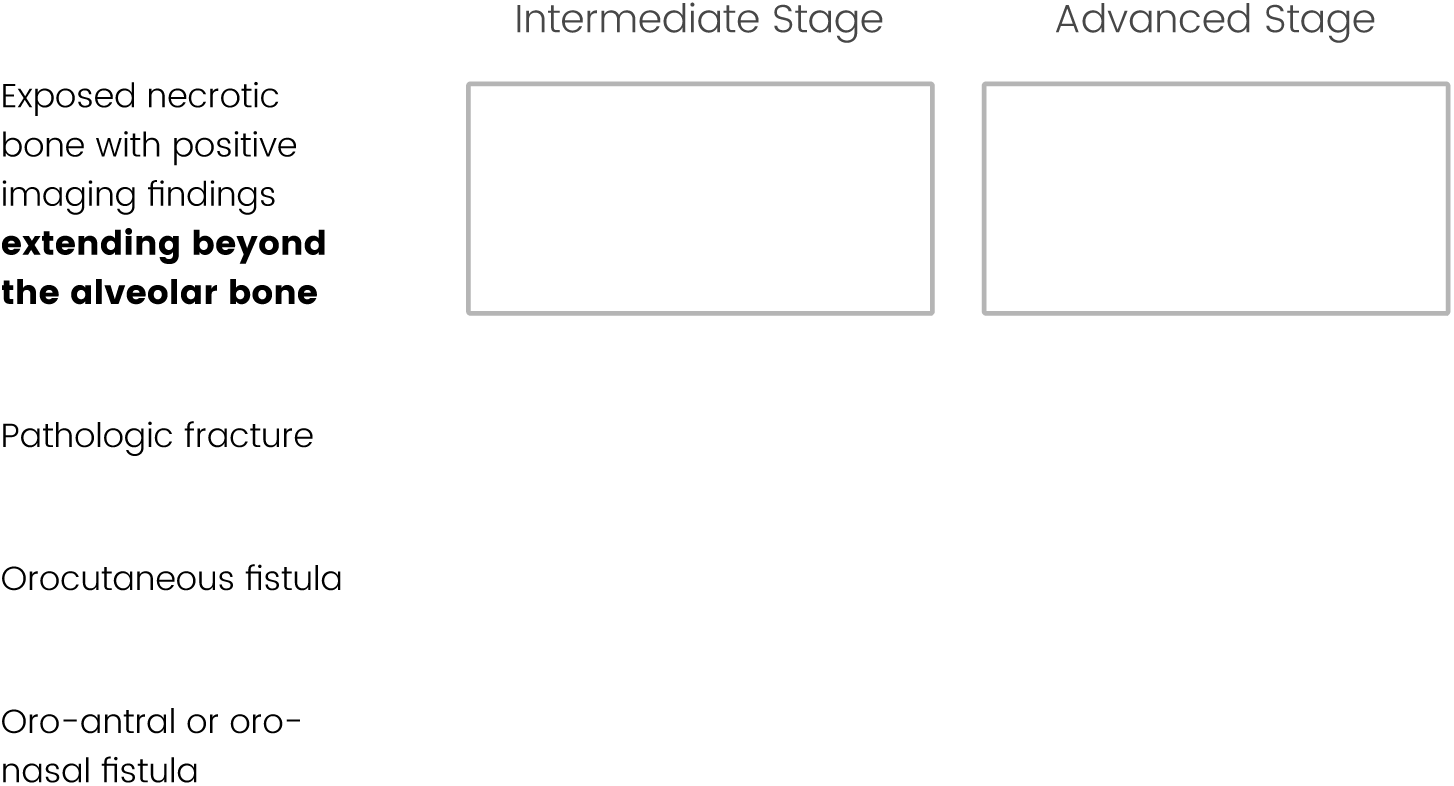

These items have been classified as possible or early ORN due to findings limited to alveolar bone. For further clarification, drag and drop <u>every item</u> to a specific staging box. If you consider all to be of same severity, drag them all to the same stage box. Note: ‘Advanced ORN’ is not an option for these items.

*Abbreviations: AB, alveolar bone; PTB, probe-to-bone test*

A PTB test example is shown below. A positive PTB test is when a hard, bone-like surface is felt after gentle probing of a wound or ulcer.

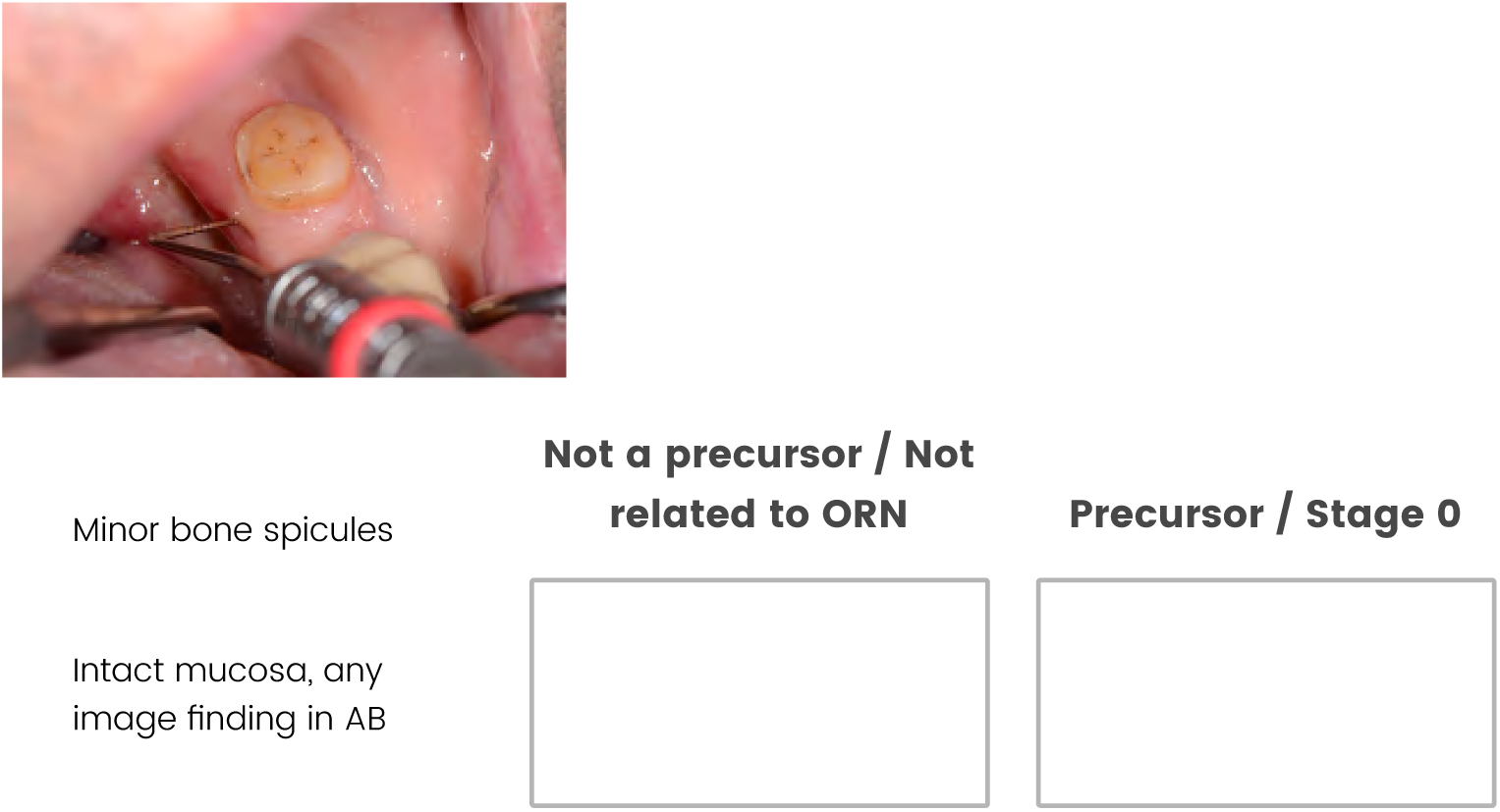
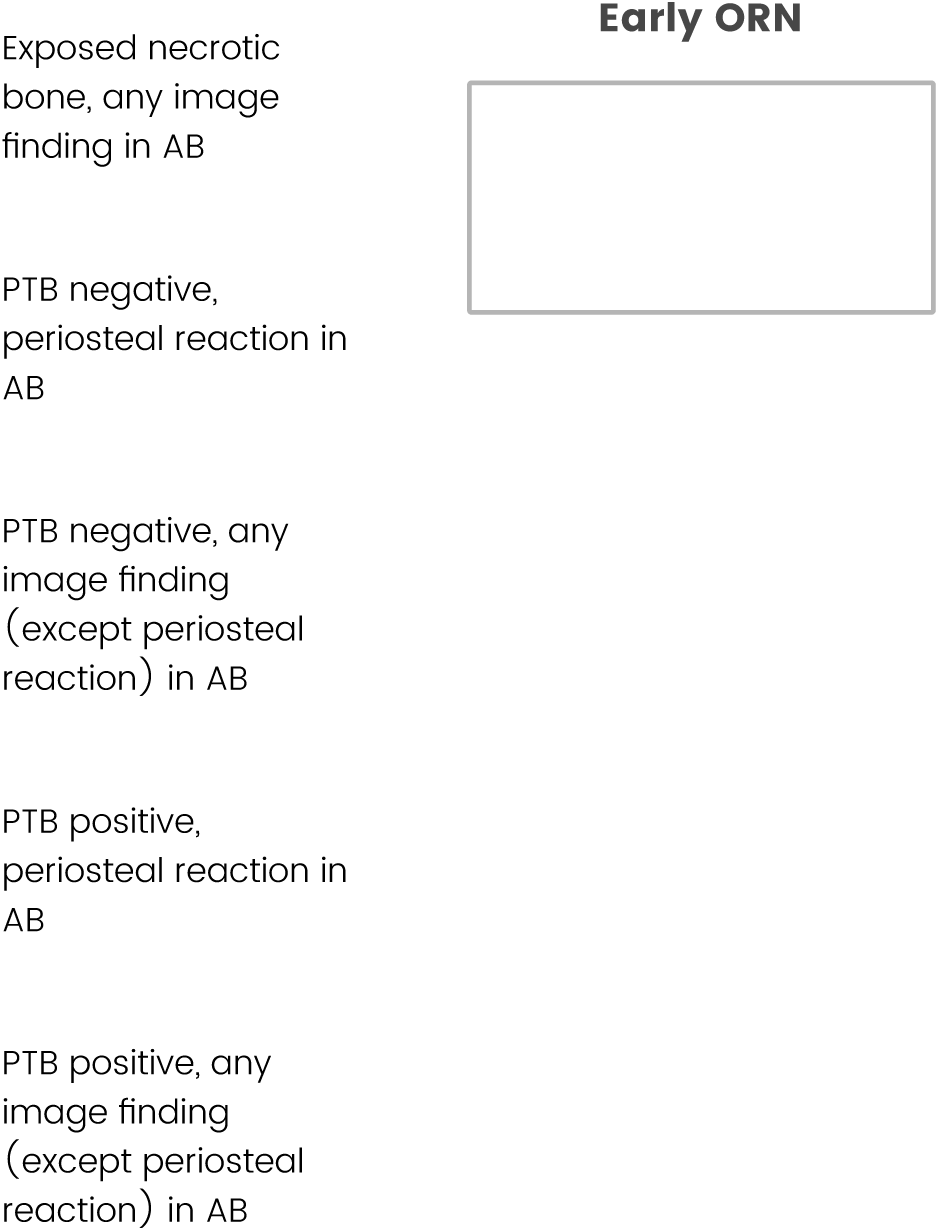

Vascular damage after radiation therapy can be measured in patients with ORN using DCE-MRI (PMID: 32712257). How should we classify a case with vascular damage in bone seen on MRI without exposed bone and without other imaging findings (i.e., CT shows no bony abnormalities)?

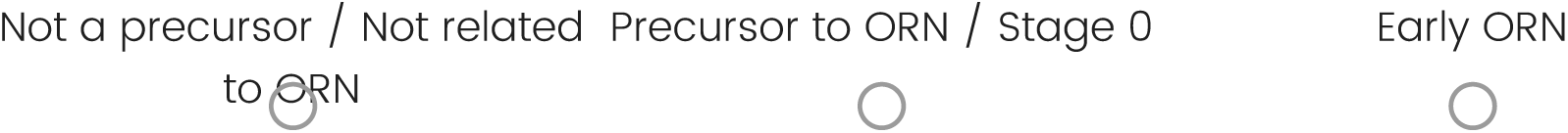

How should we classify a case with vascular damage in bone seen on MRI without exposed bone and with other imaging findings limited to alveolar bone (i.e., x-ray shows sclerosis limited to alveolar bone)?

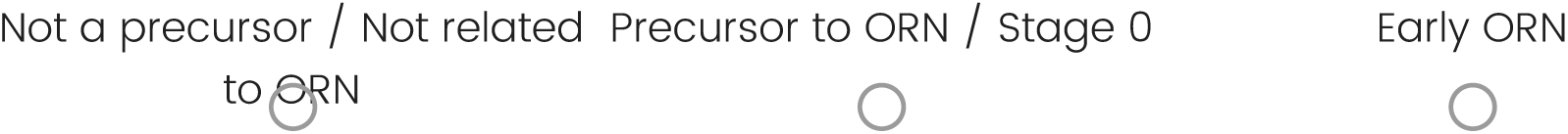

### Cases with Imaging

These items are case scenarios with clinical and radiographic information. Classify the severity of ORN and extent of bone involvement based on the information provided, and rate your level of confidence. *Note: Some images have the bony changes outlined in a red box while others do not*.

## Case 1

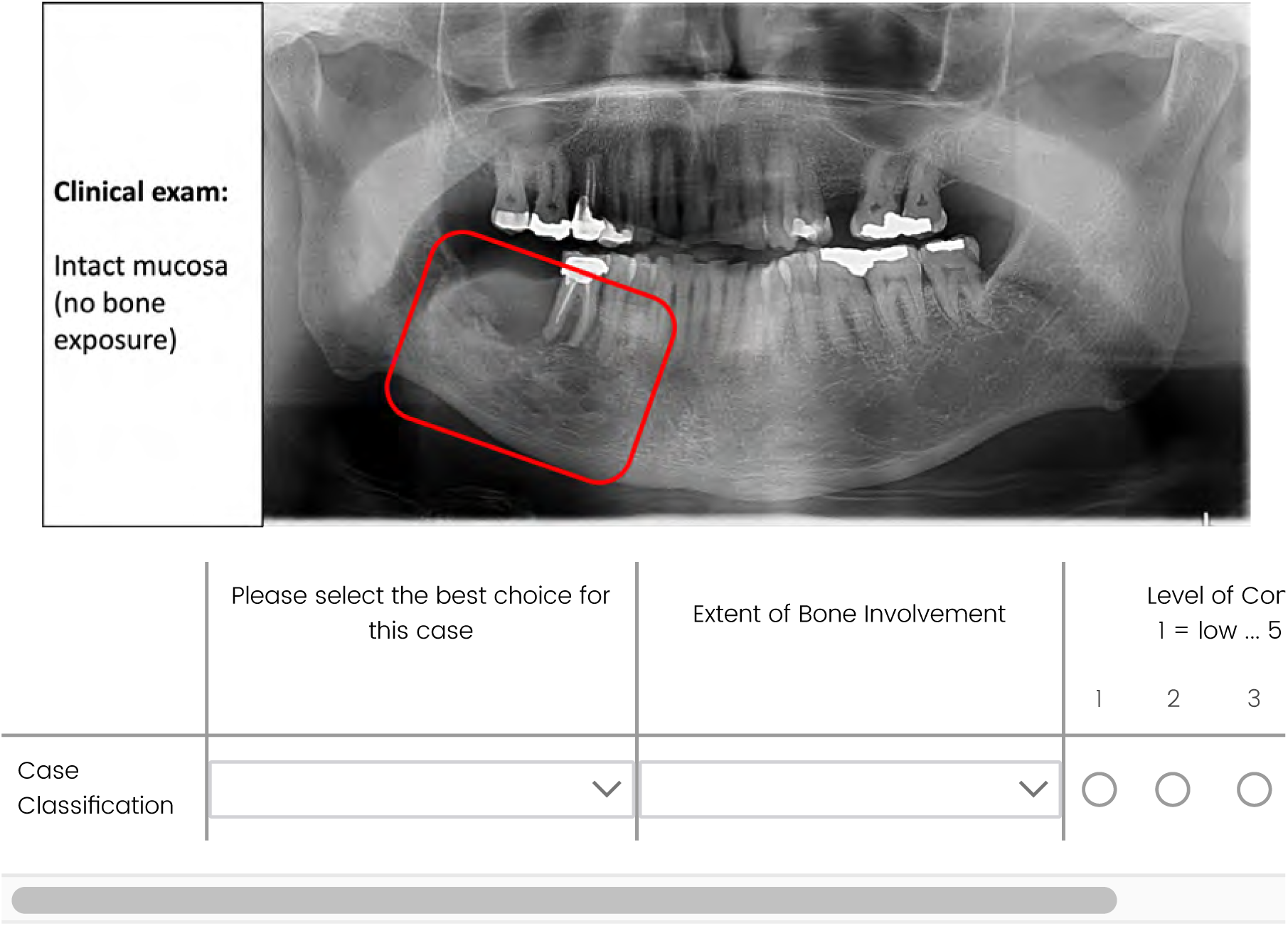

## Case 2

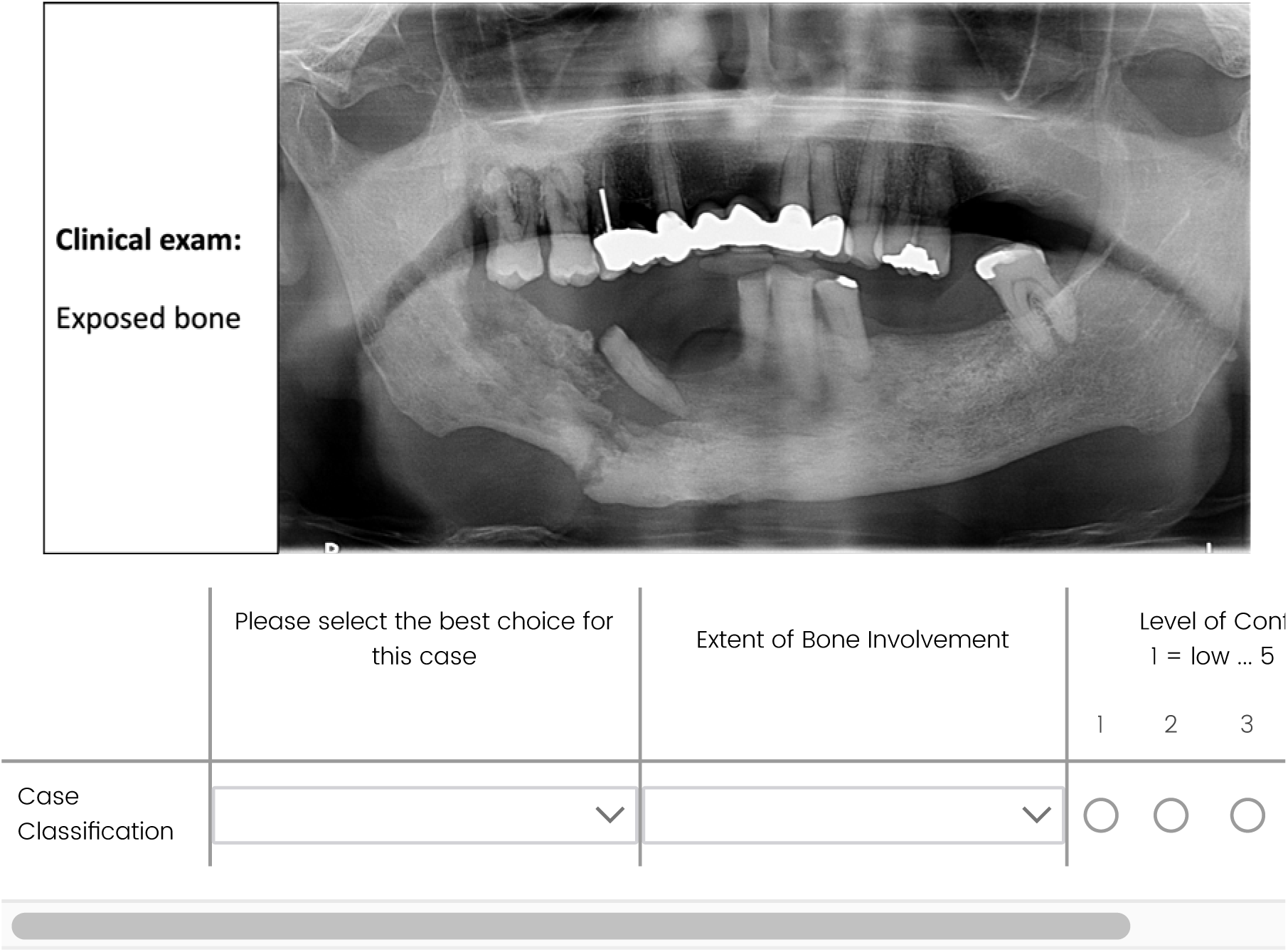

## Case 3

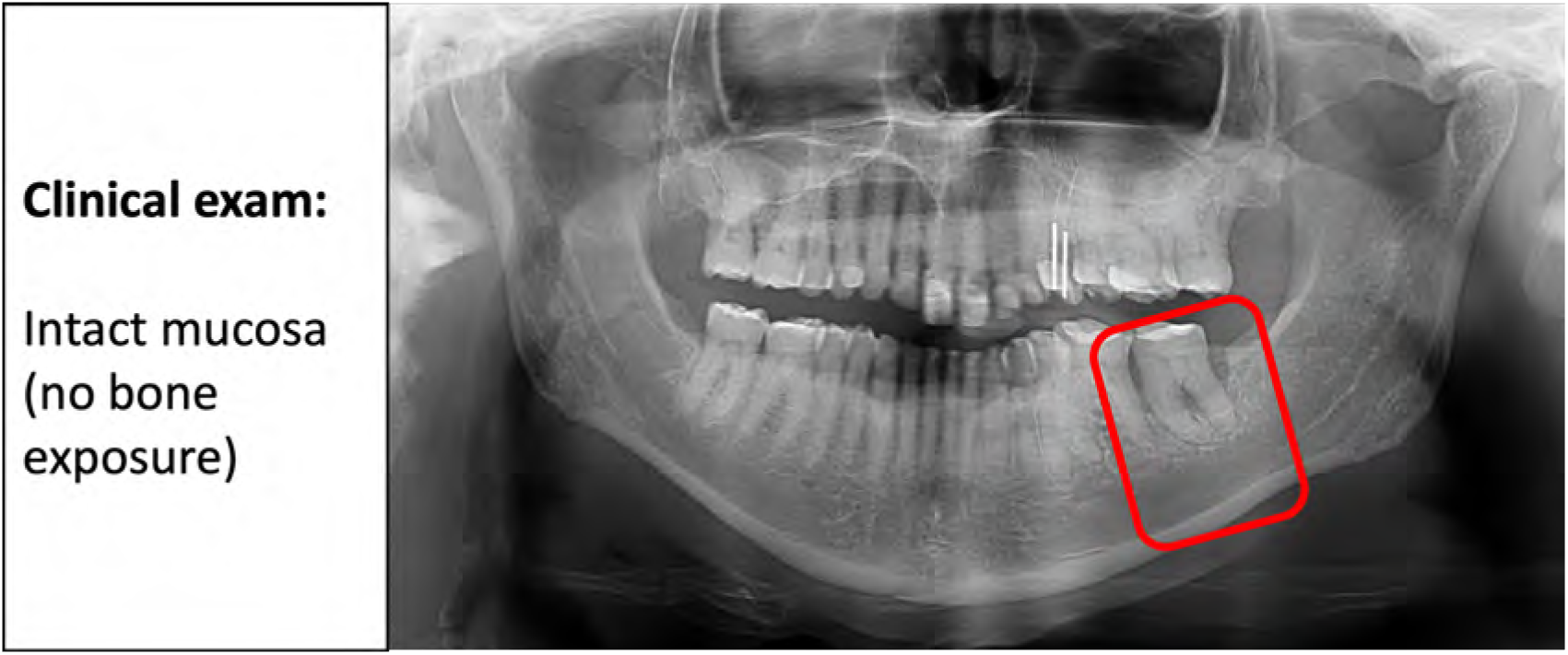
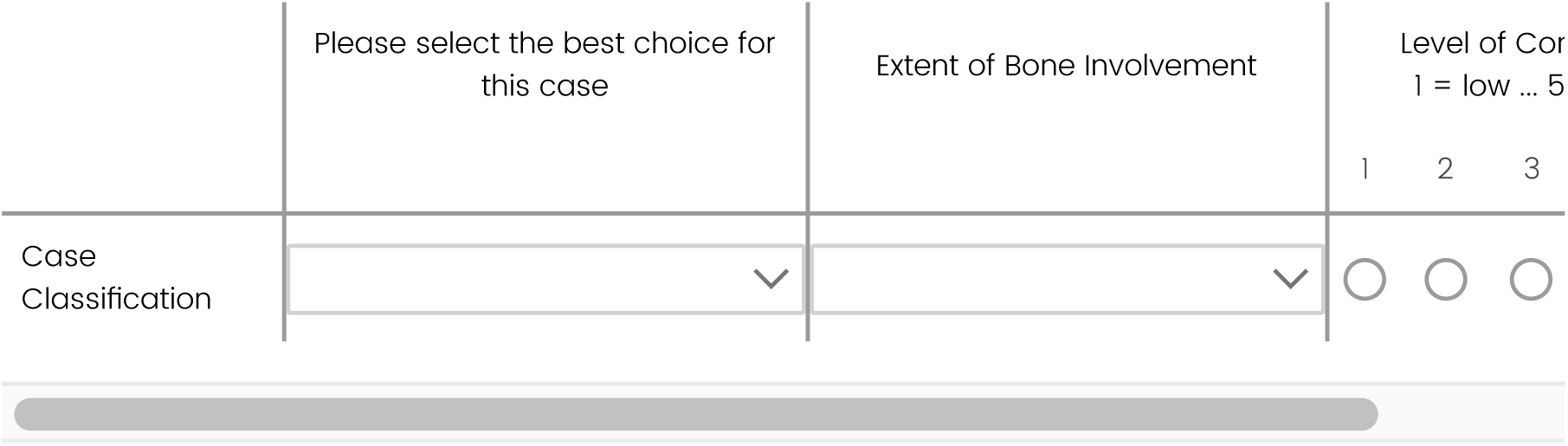

## Case 4

PTB test is positive.

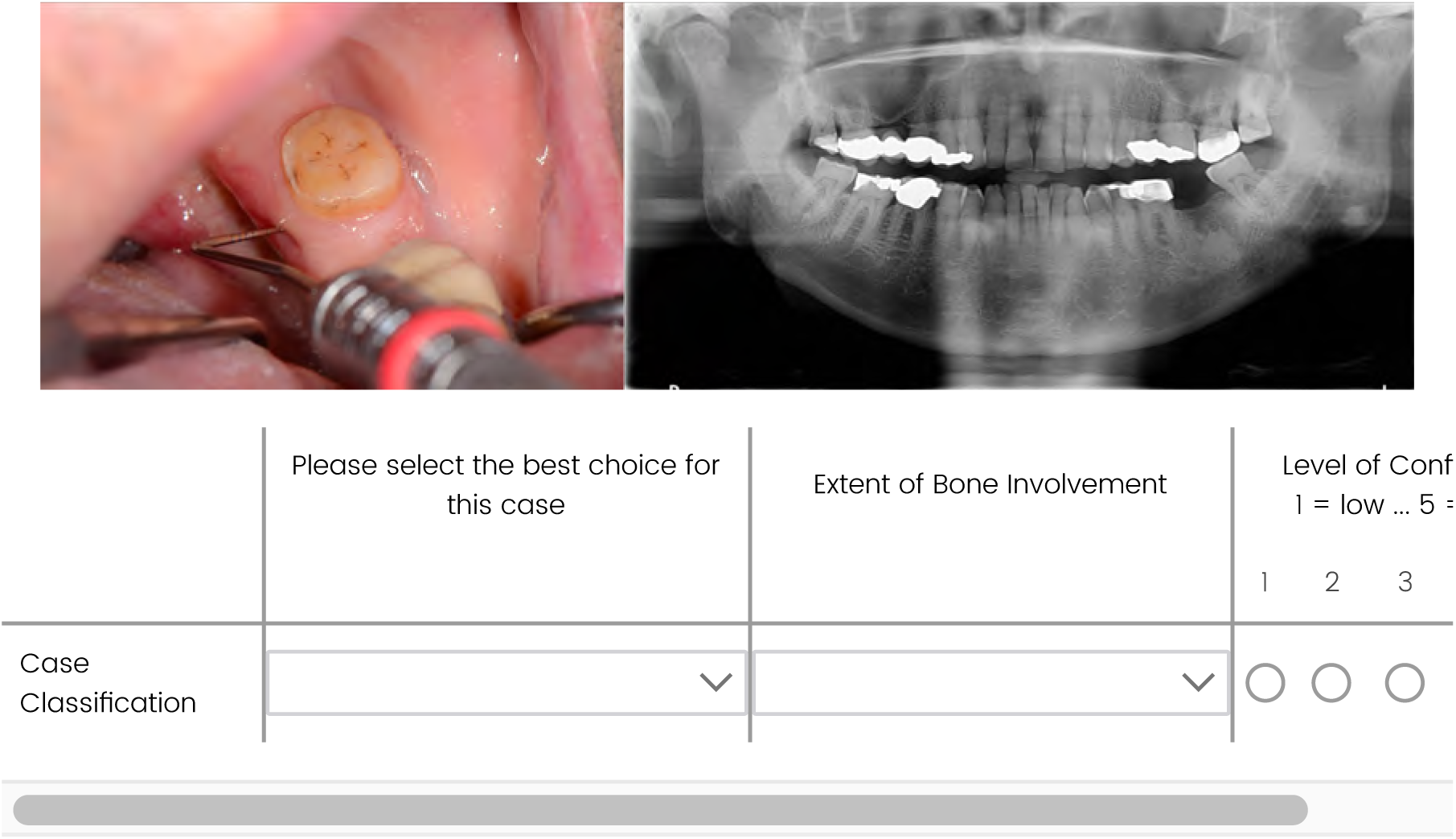

## Case 5

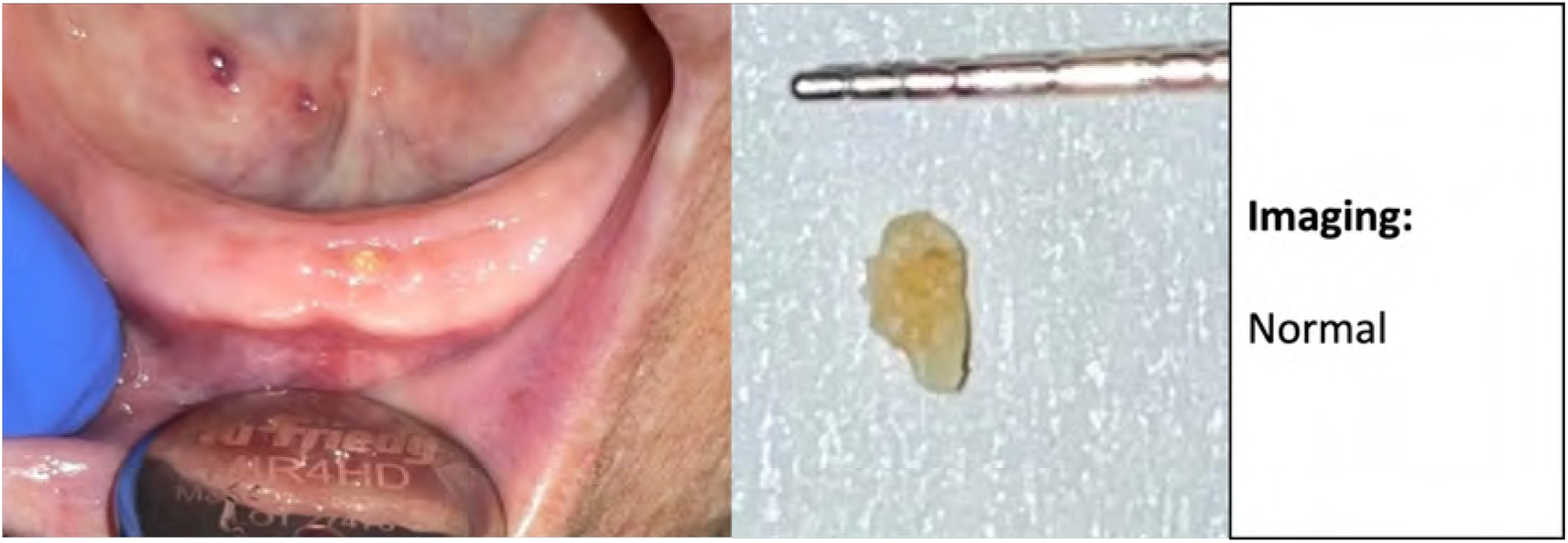
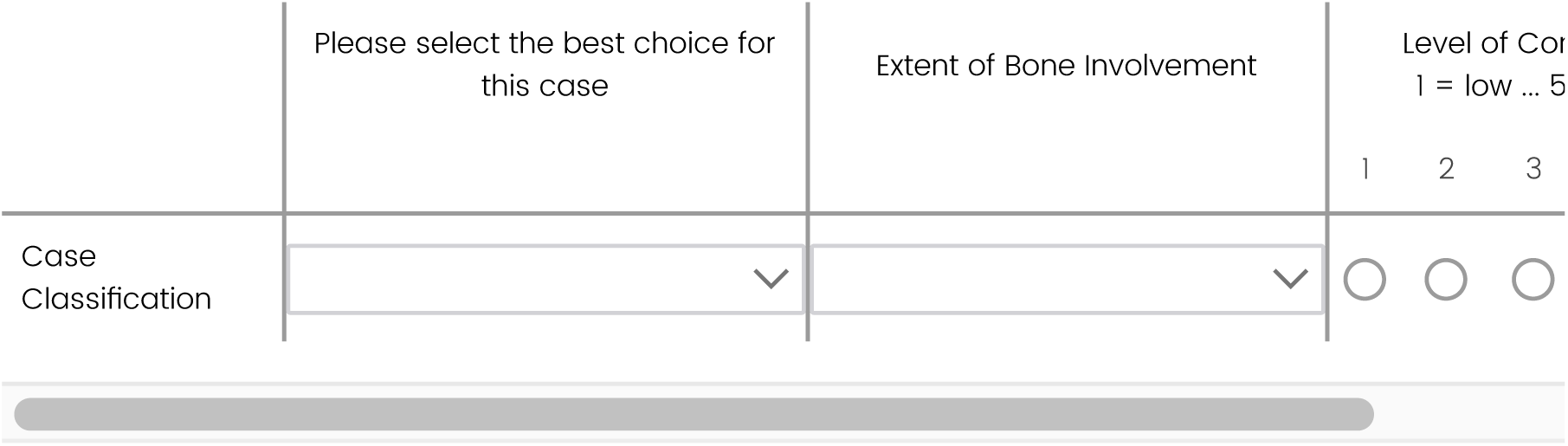

## Case 6

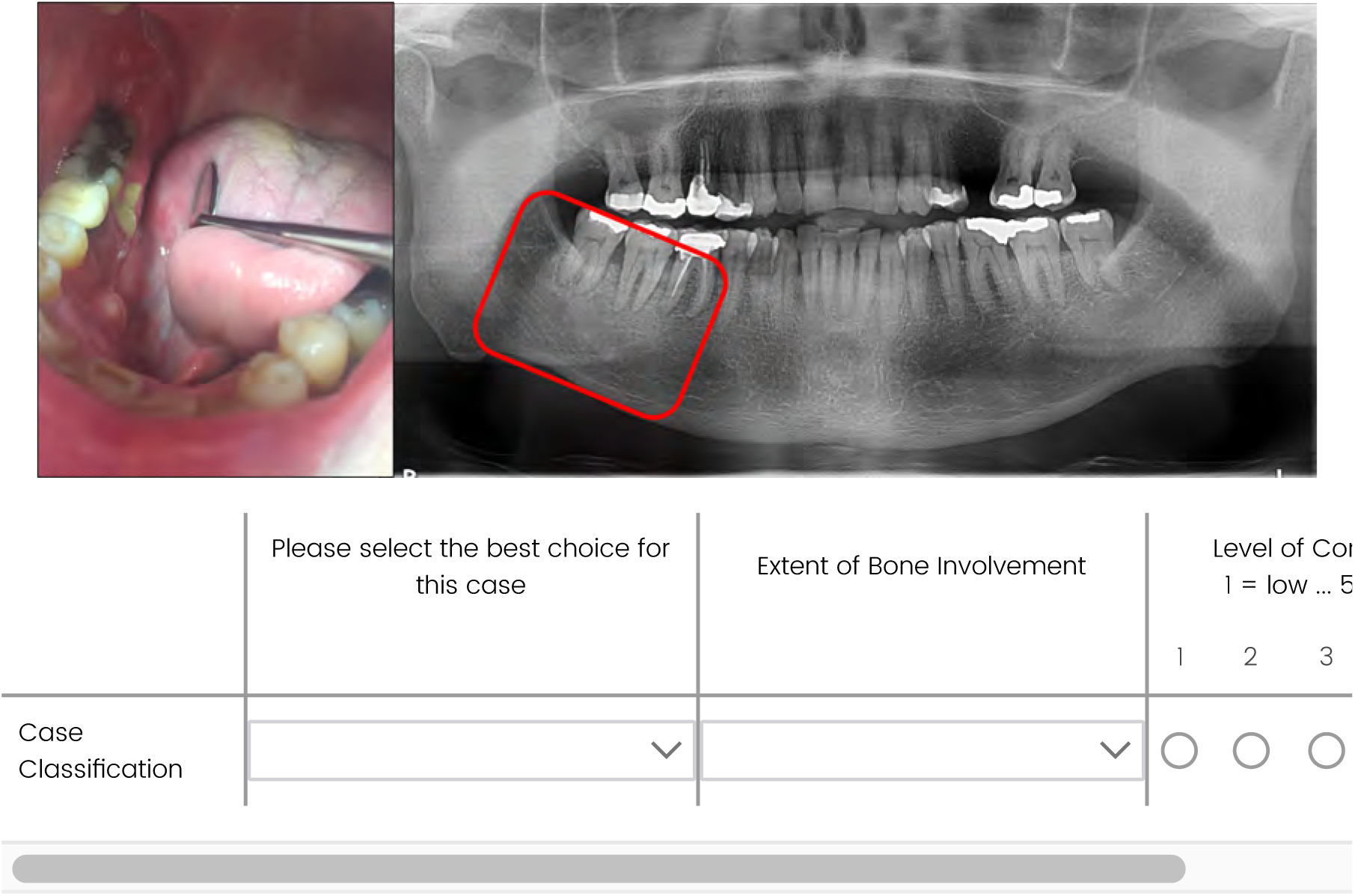

## Case 7

A mucosal ulceration with no bone exposure is noted on clinical exam. MRI-DCE shows ‘extensive loss of fat signal in the left mandible with marrow edema and associated gingival swelling and hyper-enhancement.’ Black bone MRI shows intact cortical bone. CT shows ‘soft tissue swelling and enhancement of the left mandibular gingiva and adjacent buccal space. Tooth #20 has been extracted. No suspicious bony abnormality is otherwise noted in the adjacent mandible despite evidence of bone marrow edema on recent MRI.’

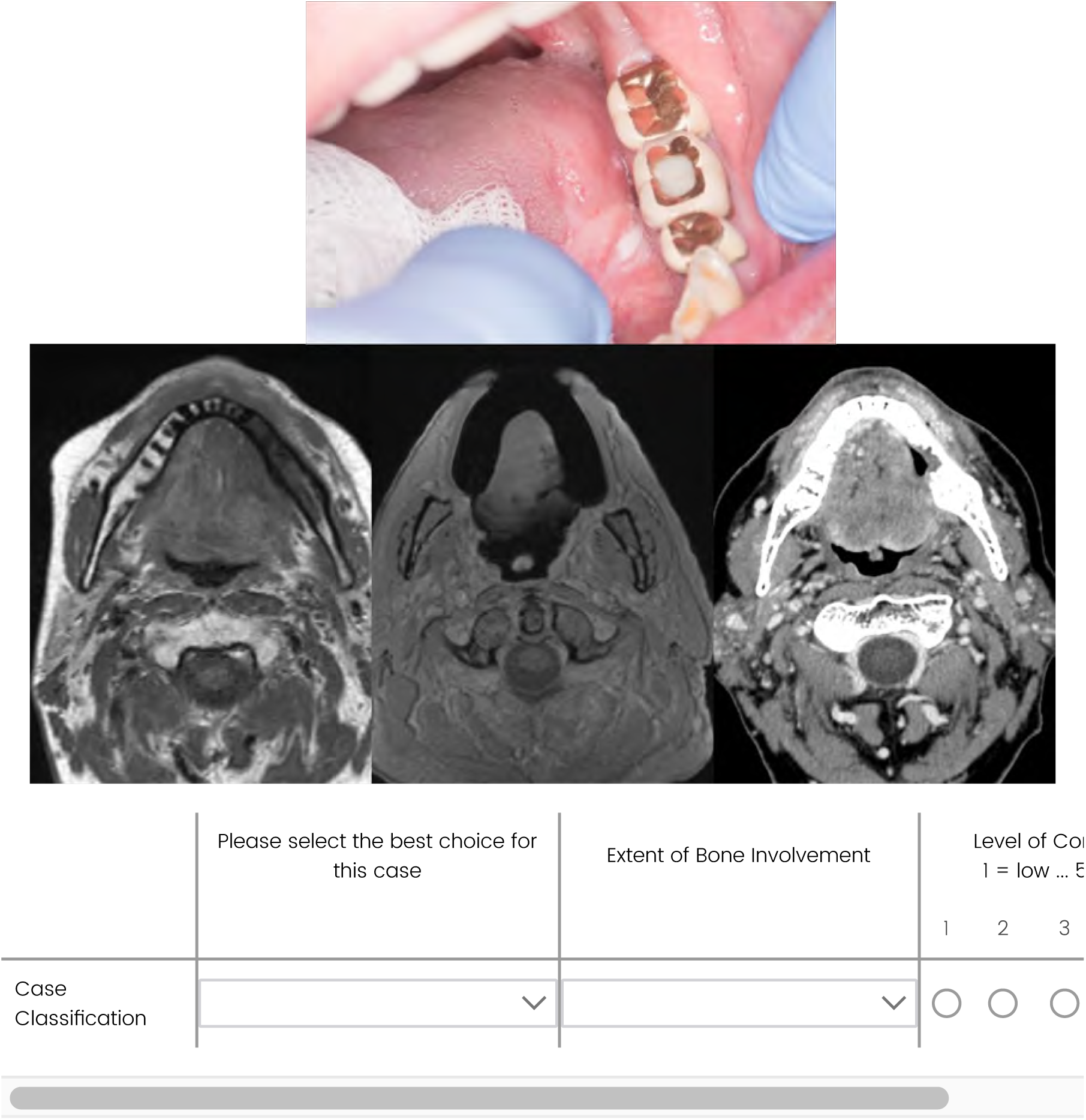

## Case 8

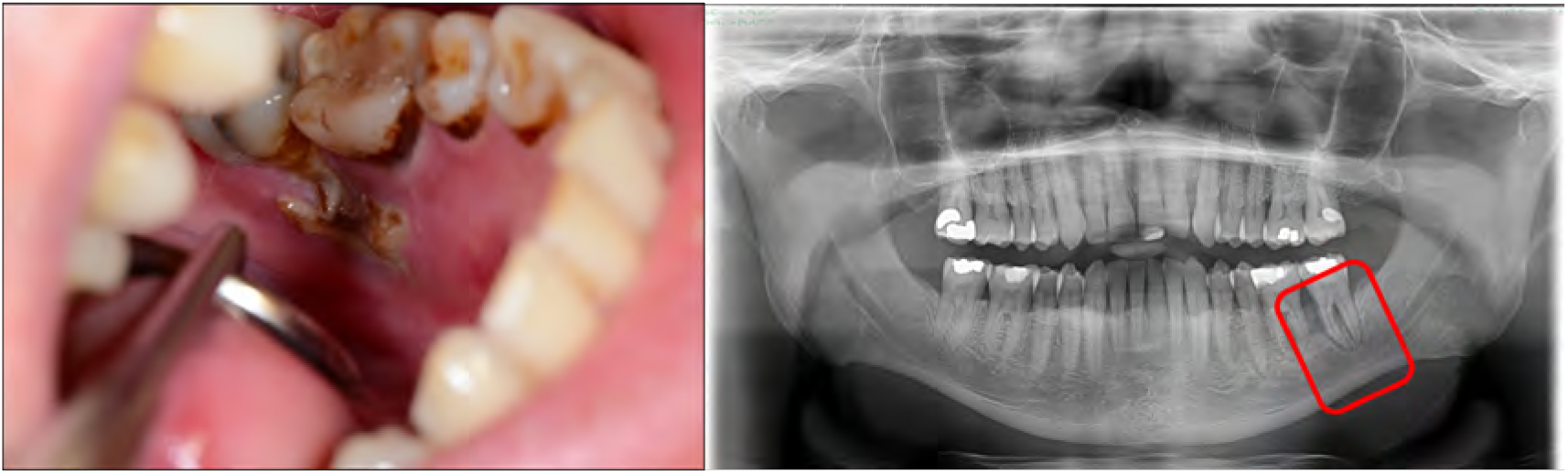
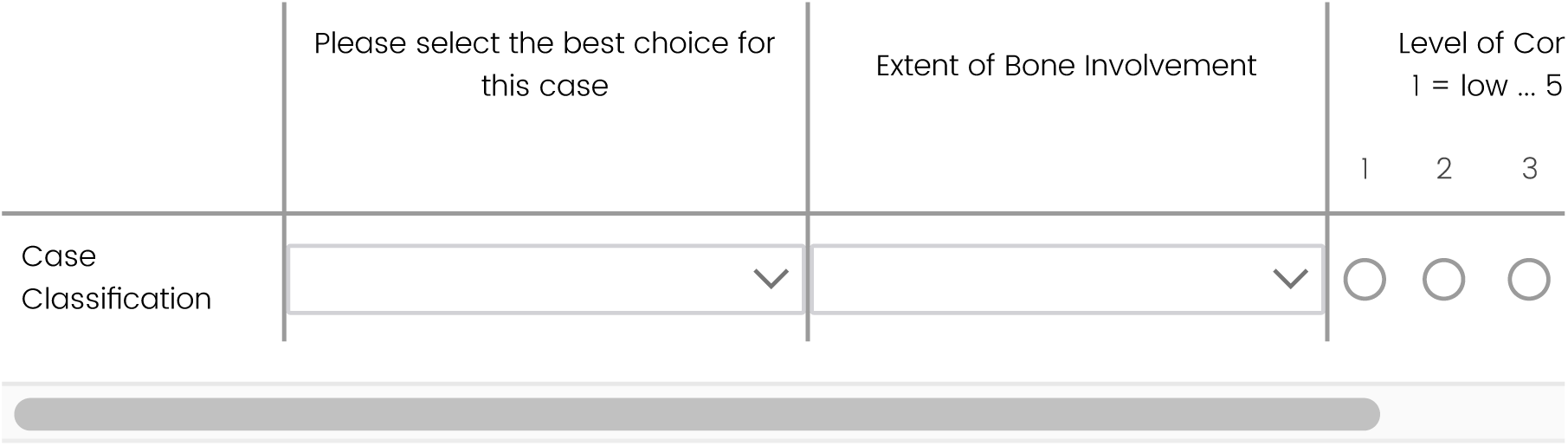

## Case 9

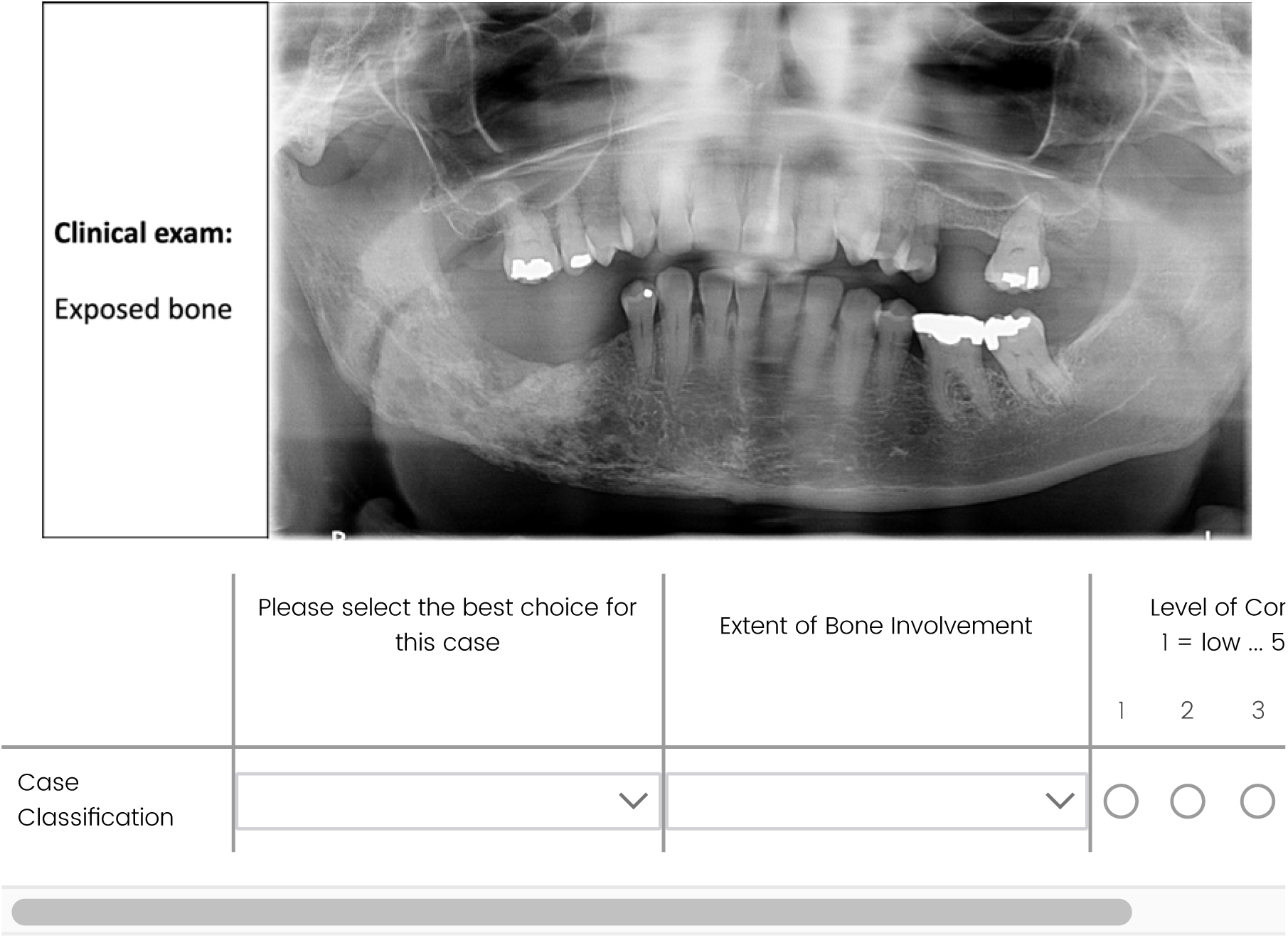

## Case 10

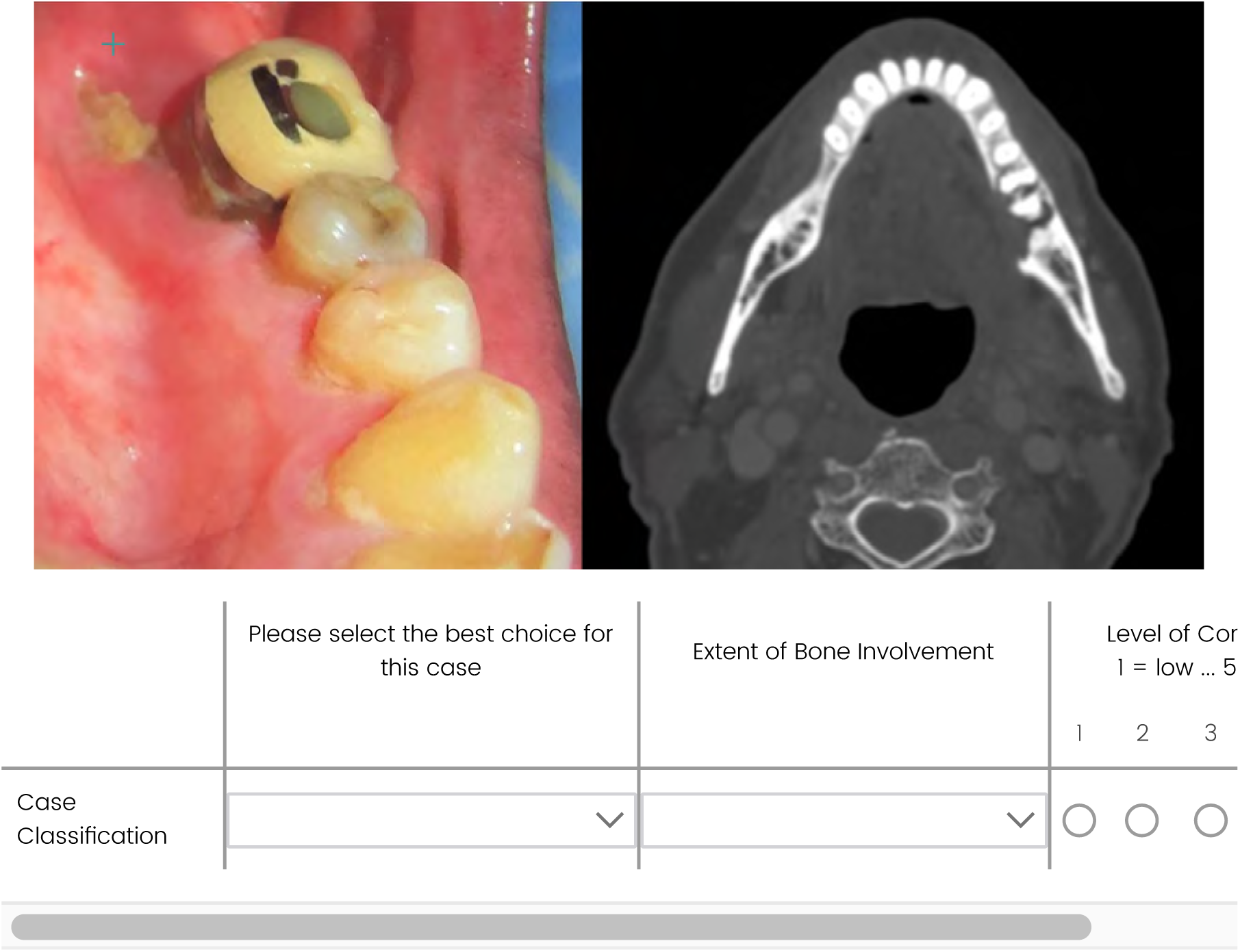

## Case 11

**Case Series**

This is a patient diagnosed with ORN with serial bony changes seen on orthopantomagram (OPG). The photo below is the patient’s teeth <u>before RT</u>.

Baseline OPG before RT:

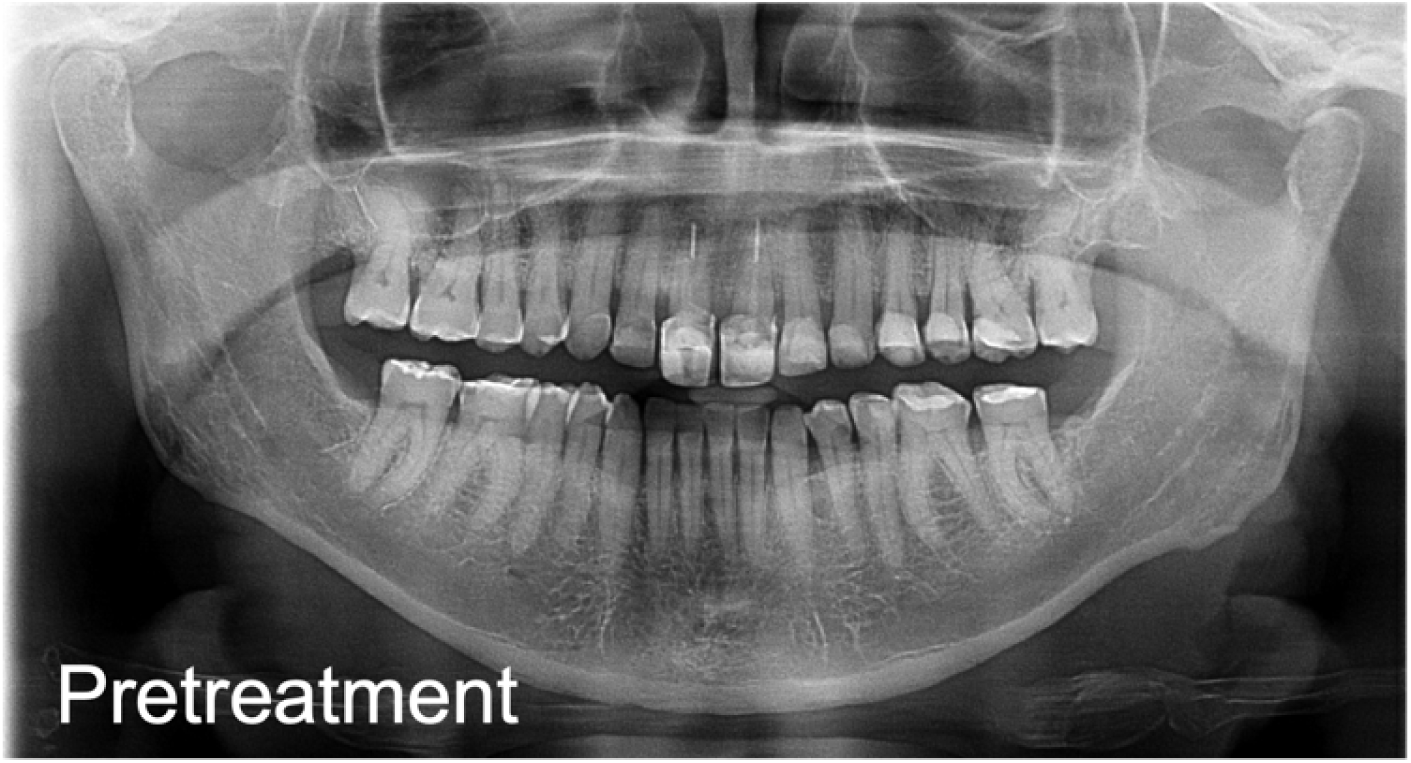

Click on the image links below to open surveillance imaging <u>after RT</u>. Upon review of the images, drag and drop the item names to the best option for stage. If more than one image could be staged the same, place them in the same staging box.

Links to surveillance imaging after RT:

[OPG: 1 year (intact mucosa)](https://mdanderson.co1.qualtrics.com/ControlPanel/Graphic.php?IM=IM_EAeh9q24iUBeUHJ)

[OPG: 3 years (exposed bone)](https://mdanderson.co1.qualtrics.com/ControlPanel/Graphic.php?IM=IM_bORDdbJ2saLhJPz)

[OPG: 4 years (exposed bone)](https://mdanderson.co1.qualtrics.com/ControlPanel/Graphic.php?IM=IM_Wf7WFfJhUoJCCpy)

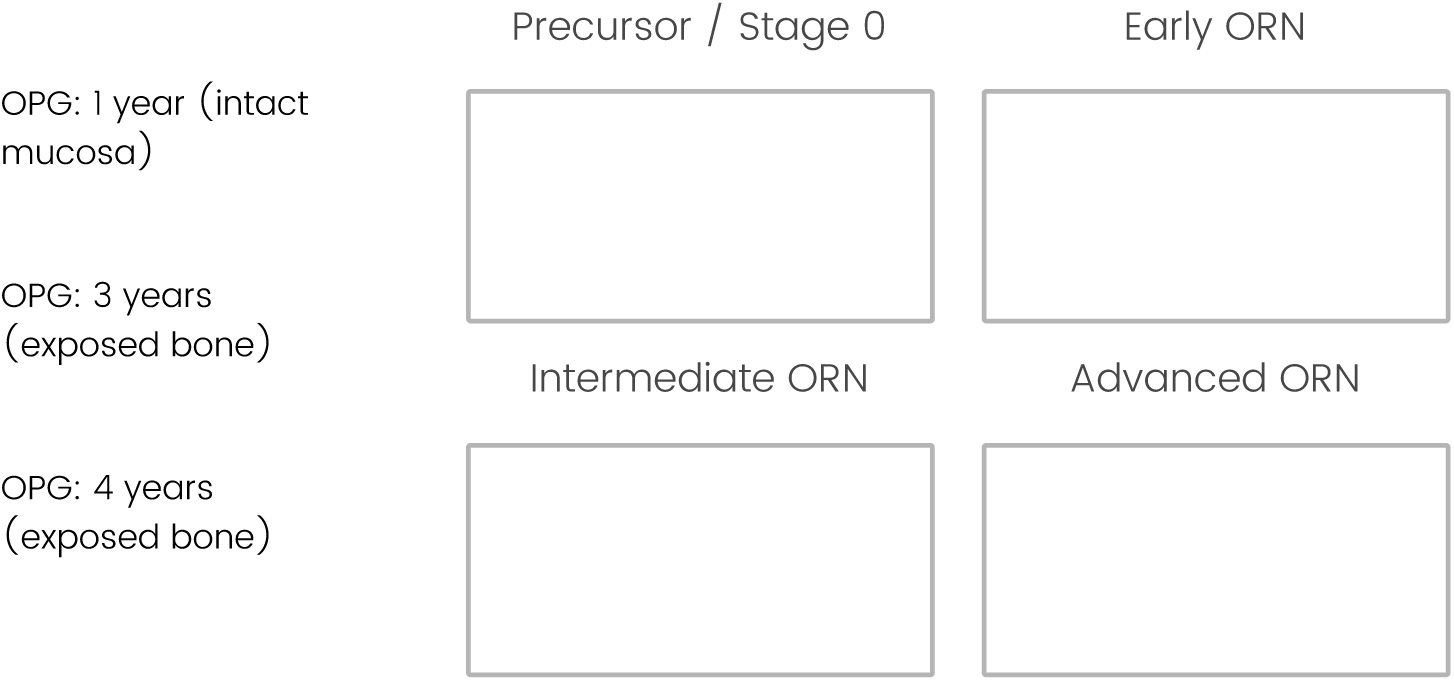

## RADMAP Summary

Regarding customization of a report with radiation dose data over an odontogram:

- 92% (54/59) recommended inclusion of systemic therapy information
- 86% (51/59) recommended inclusion of tobacco / smoking information
- 60% (35/58) recommended inclusion of gender

The top-rated heat map preferences were: single hue color palette with varying saturation (n=33) followed by a diverging color palette (n=22). Heat map colors should be color blind friendly.

This officially closes the RADMAP section of the study with expert input being taken into consideration for future report building and testing. Round 4 will provide final group feedback related to ORN diagnosis and staging.

Any comments for Round 3?

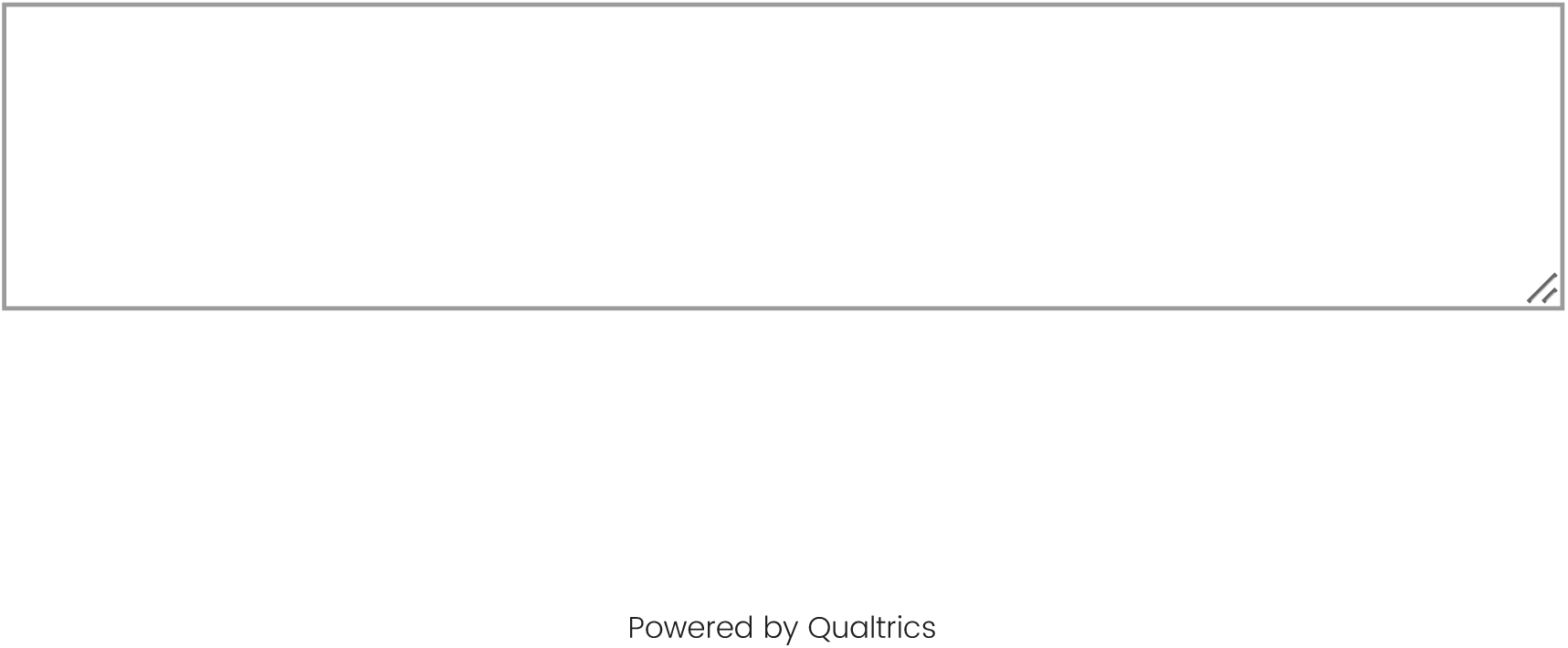

## R4_Intro

### ORAL Consortium Round 4 (Final) Survey

Hello everyone!

This will be the fourth and final round of the ORAL Consortium Delphi Study and will focus on finalizing the Consortium’s definition of ORN and clinical/radiographic classification of ORN based on group feedback from the prior rounds. Beyond this round, any topics that do not meet a new pre-defined consensus threshold of 60% will be qualitatively discussed in a related manuscript about the ORAL Consortium consensus guidelines.

Any topics that have met consensus during Round 3 will be reported in blue font.

To confirm, please select your desired level of involvement in associated manuscript(s).

- I would like to serve as a group co-author. Selecting this option will include you in regular email updates on the manuscript and a request for completion of an ICMJE disclosure form. Your name will be associated with the ORAL Consortium group authorship.
- I would like to only be acknowledged in the manuscript. Please send manuscript updates to me.
- I would like to only be acknowledged in the manuscript. Please do NOT send manuscript updates to me.
- Please do not include my name in the manuscript and do not send me updates.

## R3_Feedback & Int-Adv ORN

### The Definition of ORN

*A total of 56 panelists partially or fully completed Round 3.* In Round 3, the working ORN definition was further revised. Panelists were asked to: A) select which additional statements should be considered for inclusion in the definition from Round 2, and B) choose the best and second-best option out of 4 working definitions of ORN.

A) No additional comments met consensus and therefore will not be added to the final ORN definition. The highest scoring comment was: “it should include the presence of exposed bone” which was selected by 25 experts or 45% of the group. Distribution of selected statements is shown below.

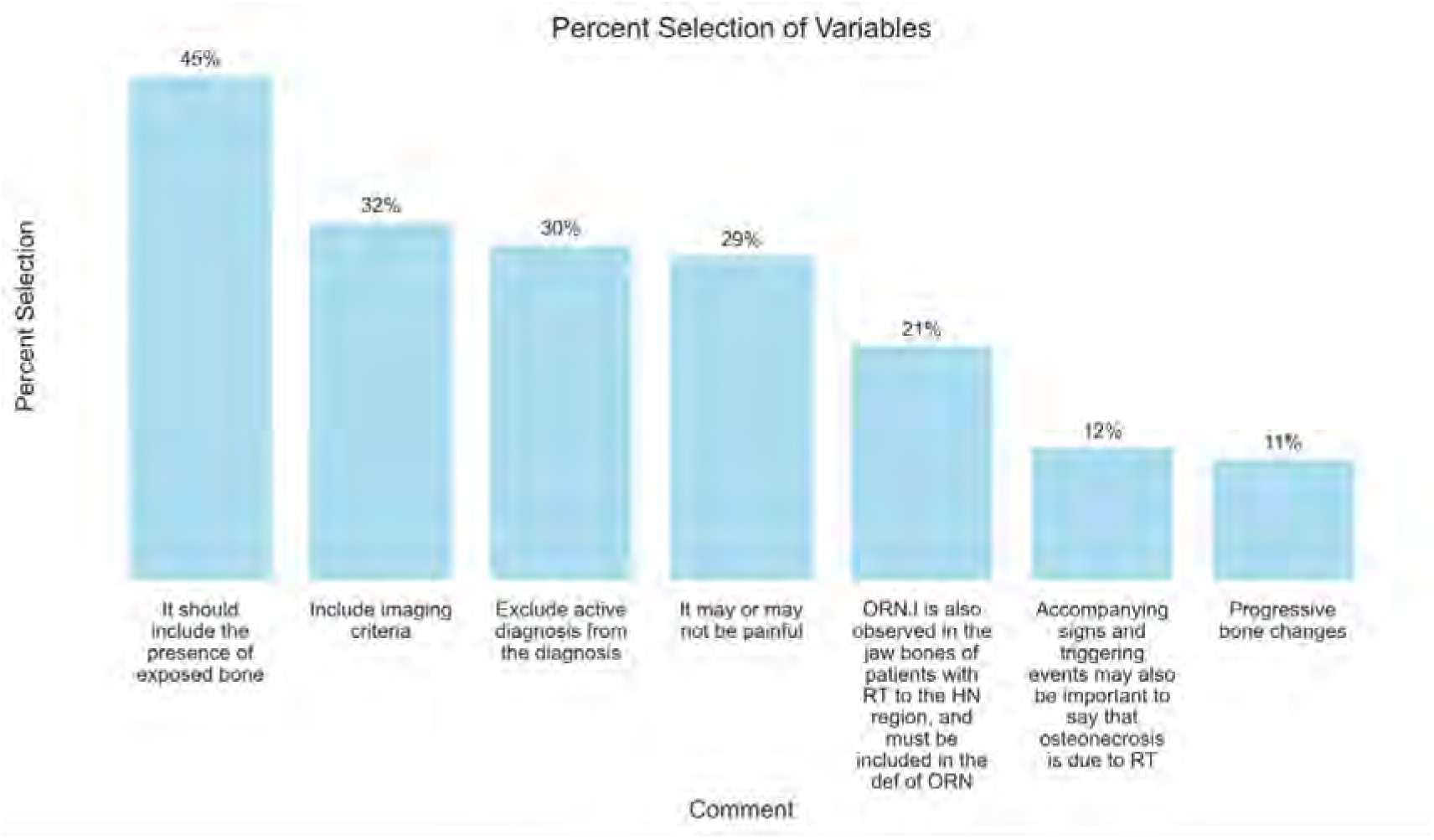

B) Best and Second Best Option. Version 1.2 was most selected for ‘Best option’ by 62% of responders followed by version 1.3 as ‘Second-best option’ selected by 47% of the group.

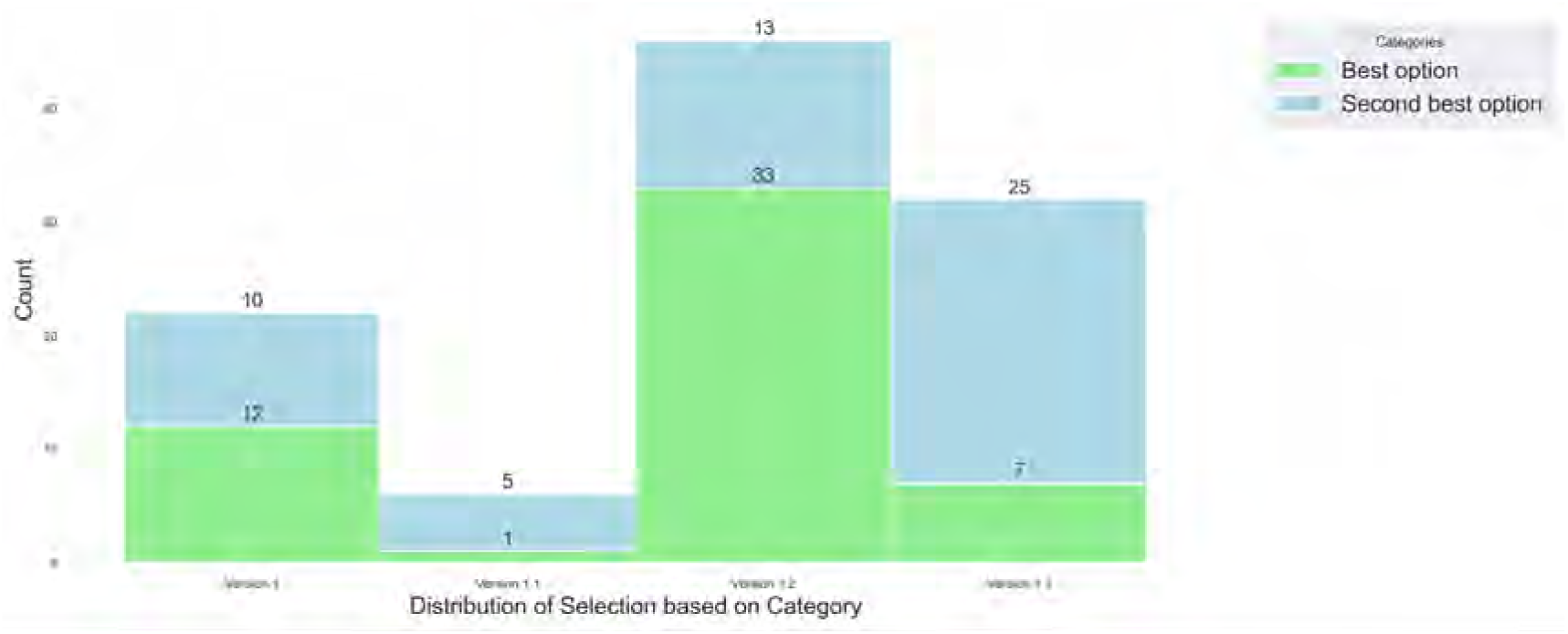

For this final round, please select your most preferred ORN definition derived from versions 1.2 and 1.3. Differences between definitions has been underlined.

- A condition in which there is a loss of blood flow to bone tissue, which causes the bone to die. Findings of bone death may be clinical (i.e., exposed bone) and/or radiographic (i.e., sclerosis, pathologic fracture). It is caused by exposure to ionizing radiation and <u>may occur at some point in time after radiation and in the absence of active disease (i.e., cancer)</u> in the site of bone death.
- A condition in which there is a loss of blood flow to bone tissue, which causes the bone to die. Findings of bone death may be clinical (i.e., exposed bone) and/or radiographic (i.e., sclerosis, pathologic fracture). It is caused by exposure to ionizing radiation and occurs in the site of bone death.

### New Staging System Publication Update

A new risk-based model for ORN classification has been published by Watson et al. which incorporates both clinical and imaging features.

Please right click on the link and open a new tab: [Watson ORN Staging System](https://www.medrxiv.org/content/10.1101/2023.09.12.23295454v1)

After review of this publication, do you find this new system helpful?

- Strongly agree
- Somewhat agree
- Neither agree nor disagree
- Somewhat disagree
- Strongly disagree

### Intermediate vs. Advanced Stage Features

In Round 3, panelists were asked to classify 4 features as intermediate or advanced stage. The following met the consensus threshold to be indicative of advanced ORN: pathologic fracture (49/51, 96%), orocutaneous fistula (47/51, 92%) and oro-antral or oro-nasal fistula (43/51, 86%).

Classification of ‘*exposed necrotic bone with positive imaging findings extending beyond the alveolar bone*’ did not meet consensus (43% intermediate; 57% advanced). A significant association between specialty group and recommended ORN stage was observed for this feature with most <u>dental specialists</u> considering it as intermediate stage (69%) while <u>other specialty groups</u> (72% Rad Onc, 57% Surg Onc) considered it as advanced stage (P = 0.04).

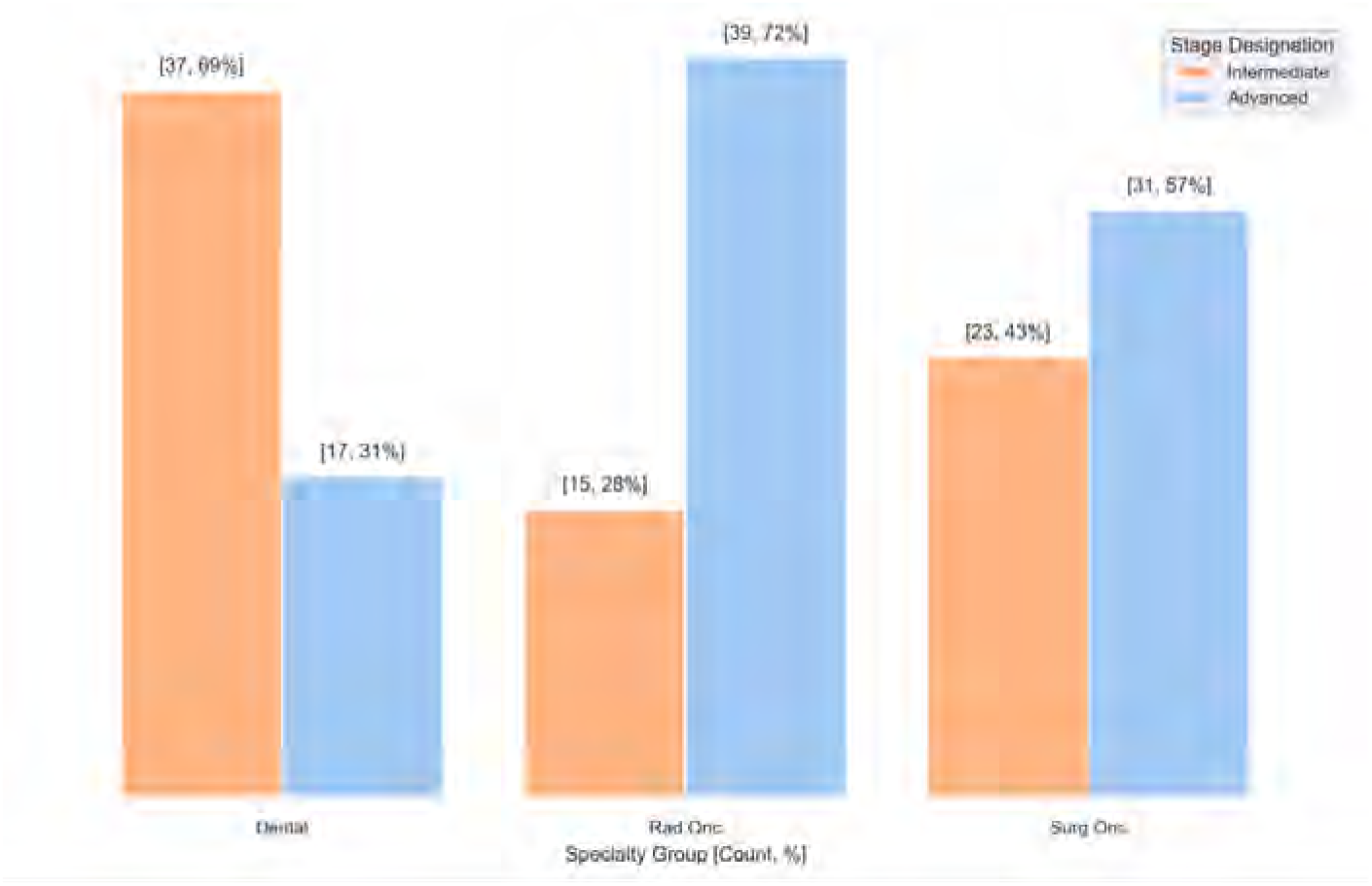

Case 9 represented a similar scenario of clinically exposed bone with radiographic features beyond the alveolar bone in the right mandible.

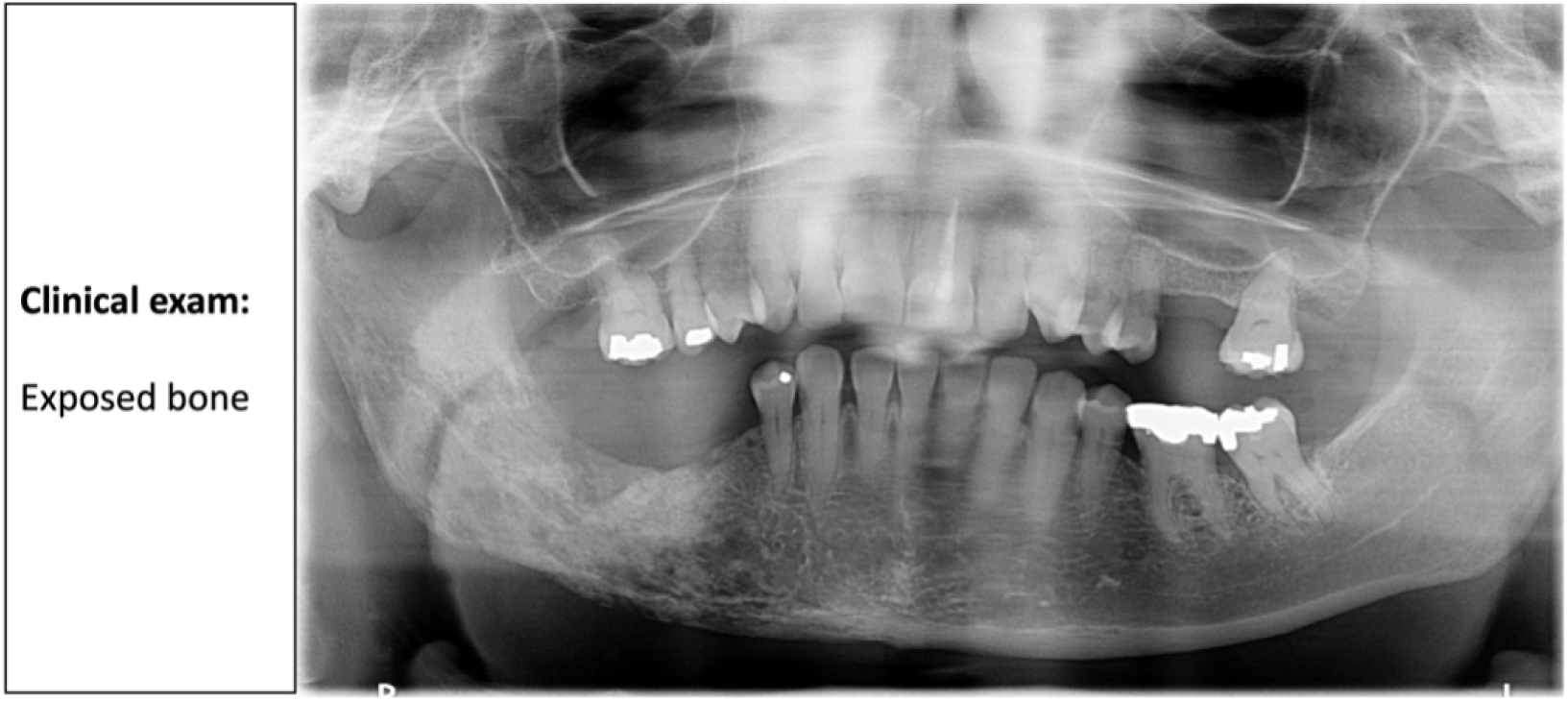

Group feedback for this case is as follows. Note: This case was classified with moderate-to-high level of confidence by 62% panelists in dentistry/oral medicine, 32% RadOnc, and 28% Surgery (56% overall).

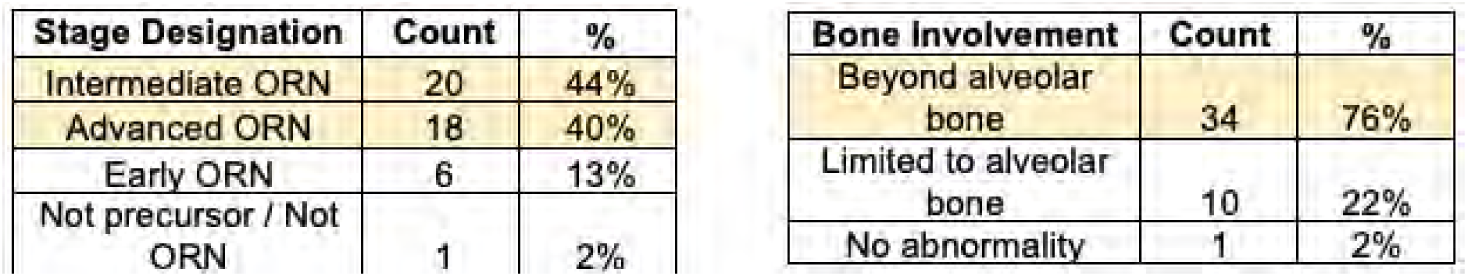

Considering the group feedback and consensus-based features of advanced ORN (pathologic fracture, orocutaneous fistula, oro-antral/oro-nasal fistula), please reclassify the feature:

‘Exposed necrotic bone with positive imaging findings extending beyond the alveolar bone’

- Intermediate ORN
- Advanced ORN

### Precursor vs Early ORN

To differentiate between unrelated, precursor, or early staging features of ORN, panelists were asked to classify 7 items as one of the following:

1. Not a precursor/not related to ORN
2. Precursor/stage 0
3. Early ORN

The figure below shows the distribution of stage designation per feature combination. Only ‘exposed necrotic bone, any image finding in AB’ met consensus to be classified as Early stage ORN (n=49, 100%).

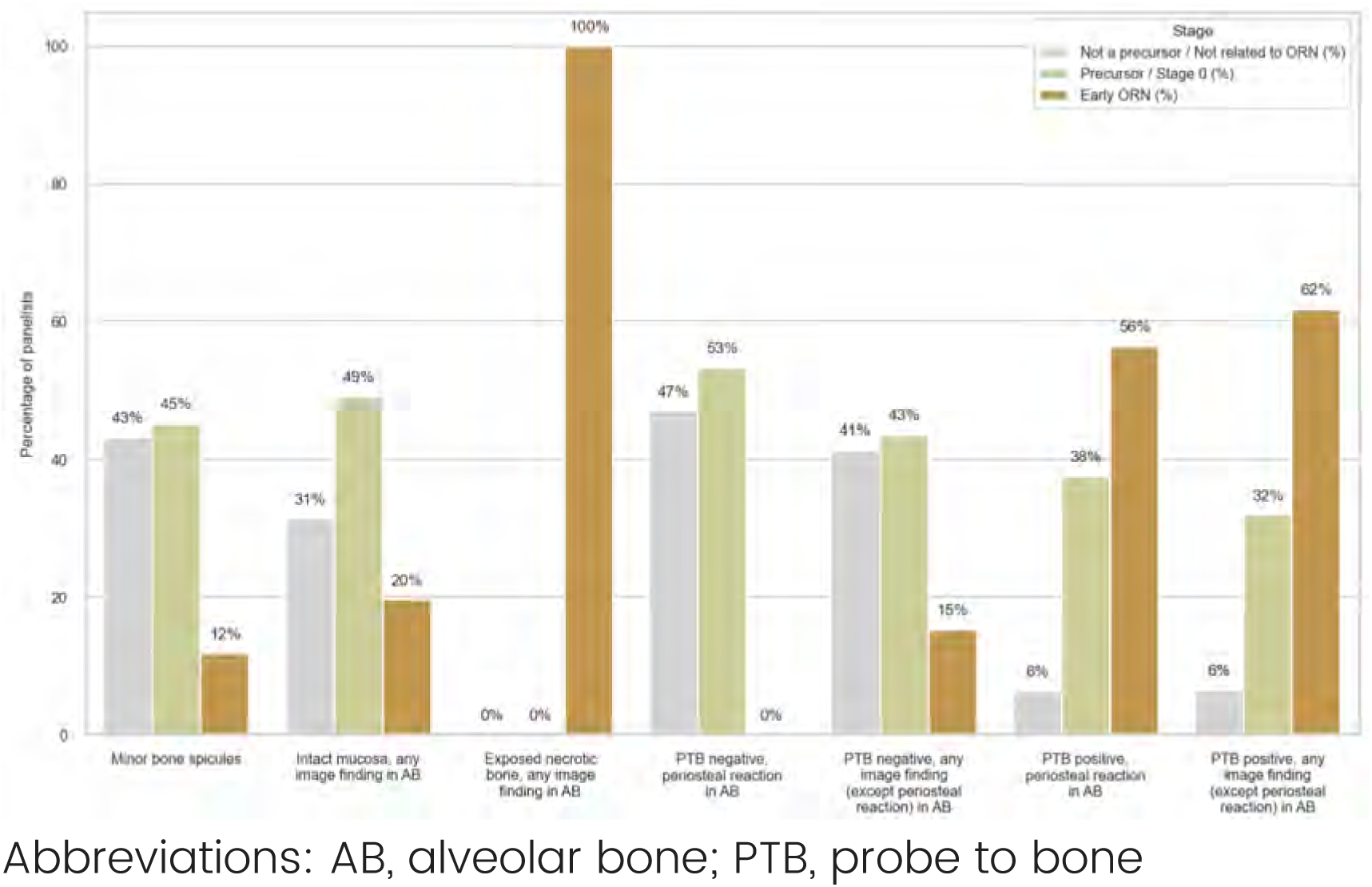

The next series of questions will show group feedback for each feature combination, stratified by specialty. You will then be asked to classify the feature into one of the top 2 categories.

Group feedback for: Minor bone spicules

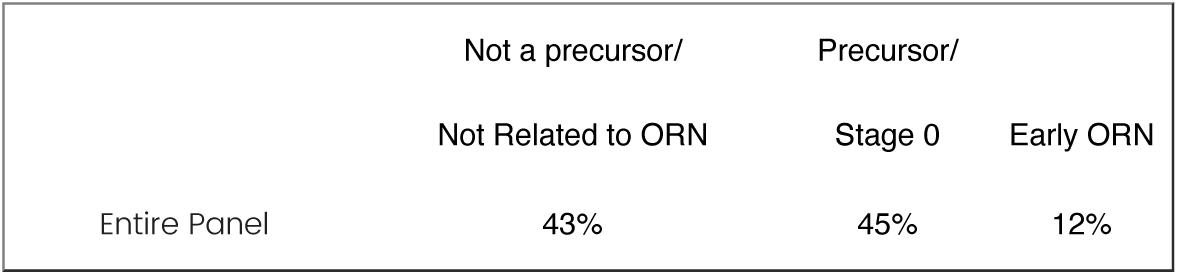

Classification per specialty:

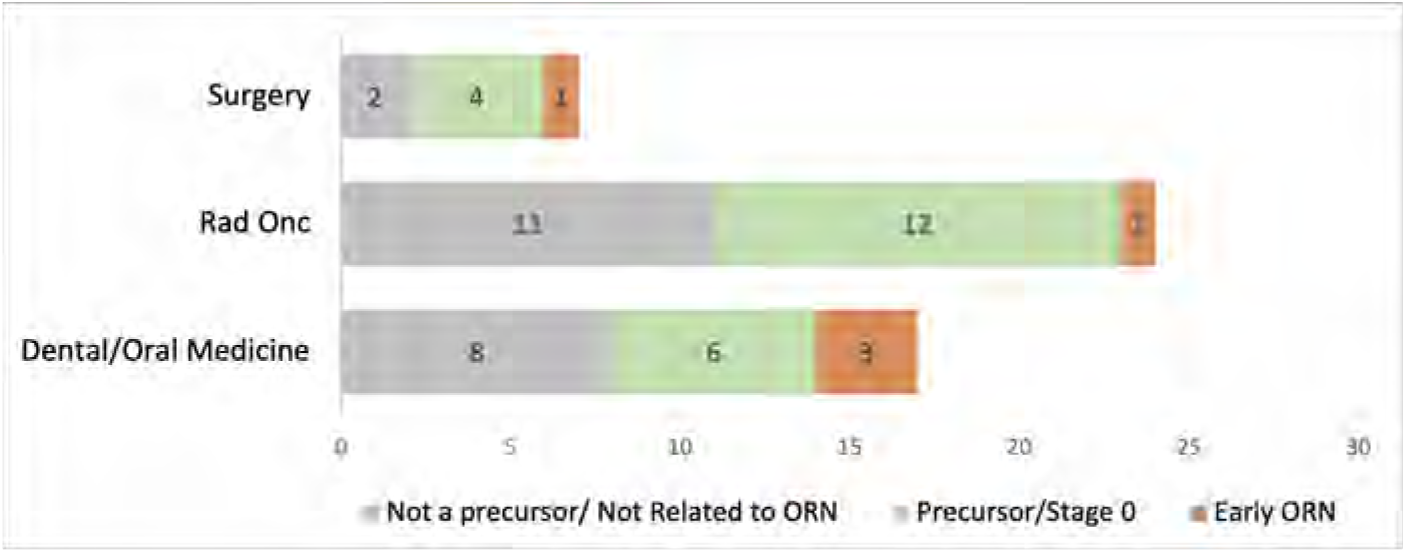

How should this feature be classified?

- Not a precursor / Not related to ORN
- Precursor / Stage 0

Group feedback for: Intact mucosa, any imaging findings in alveolar bone

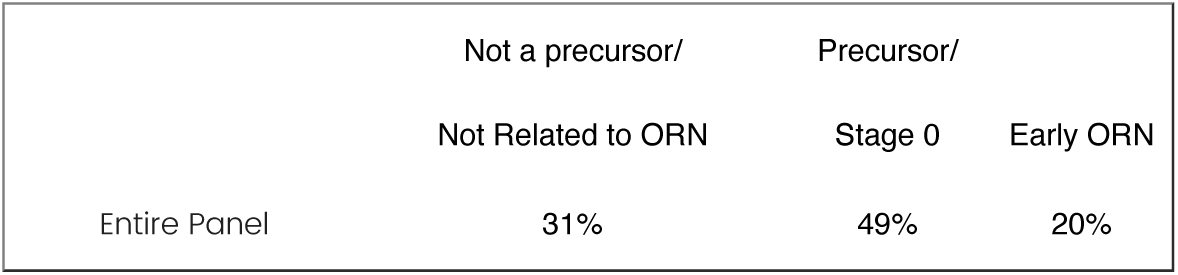

Classification per specialty:

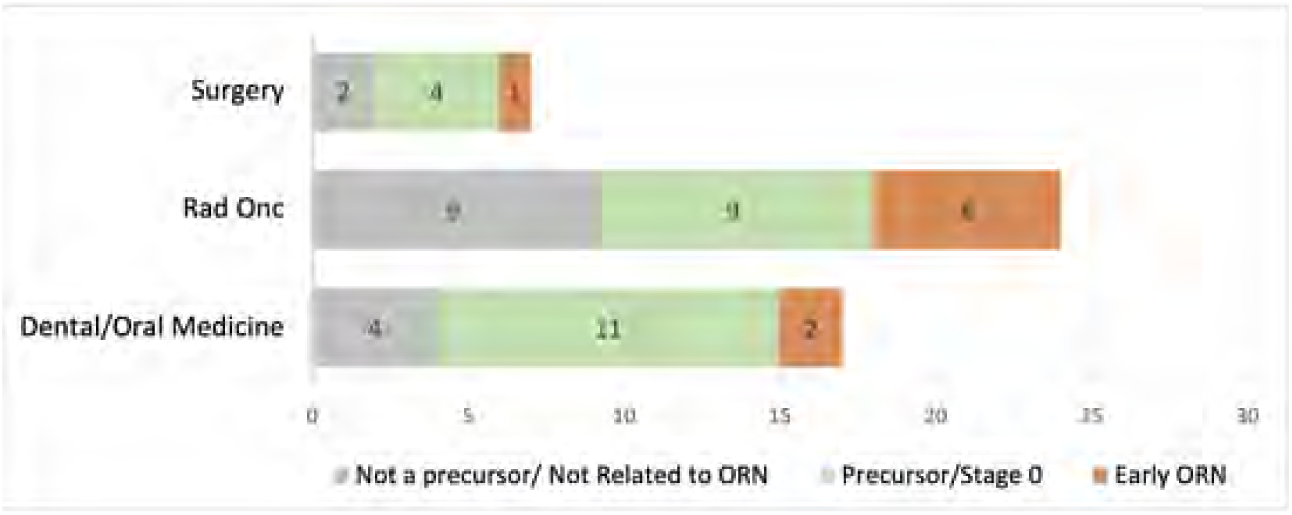

How should this feature be classified?

- Not a precursor / Not related to ORN
- Precursor / Stage 0

Group feedback for: Probe-to-bone negative, periosteal reaction within alveolar bone seen on imaging

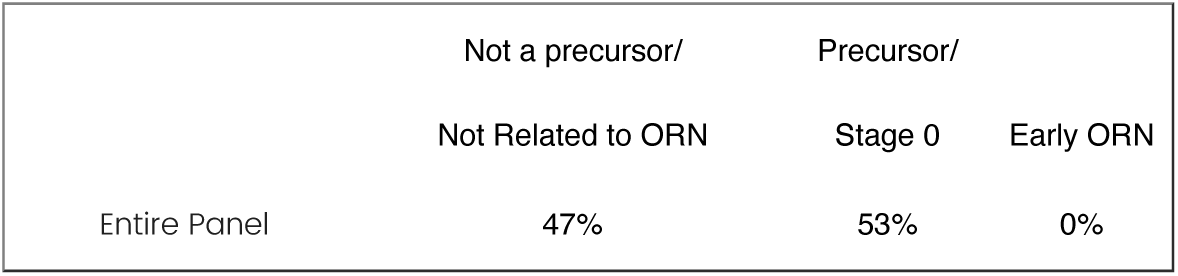

Classification per specialty:

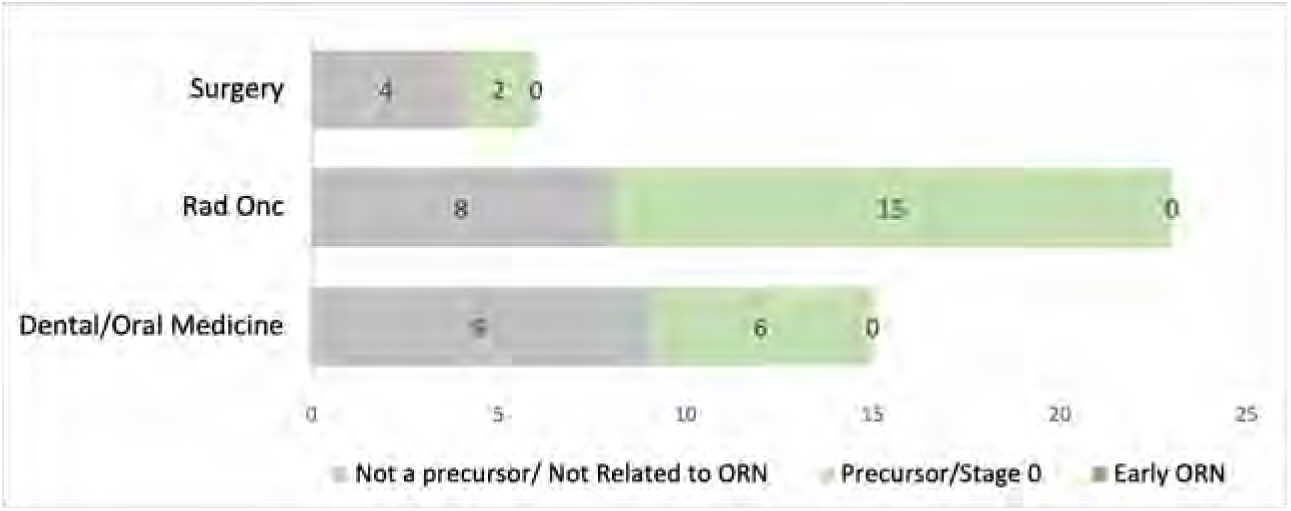

How should this feature be classified?

- Not a precursor / Not related to ORN
- Precursor / Stage 0

Group feedback for: Probe-to-bone negative, any imaging finding (EXCEPT periosteal reaction) within alveolar bone

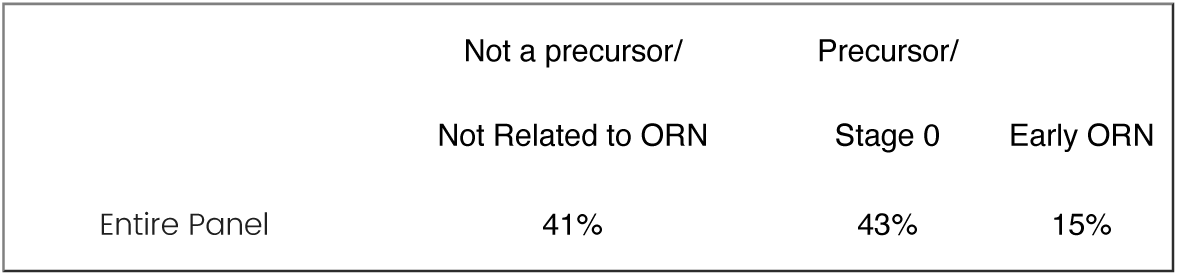

Classification per specialty:

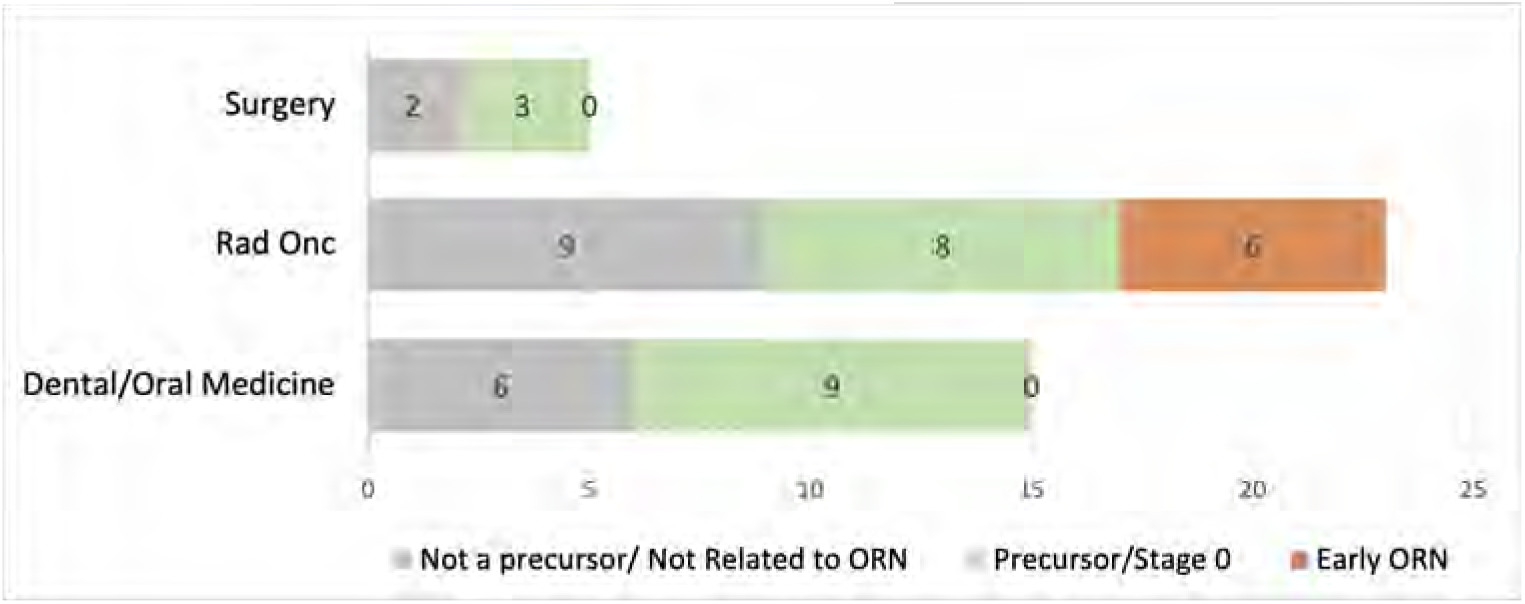

How should this feature be classified?

- Not a precursor / Not related to ORN
- Precursor / Stage 0

Group feedback for: Probe-to-bone POSITIVE, periosteal reaction within alveolar bone seen on imaging

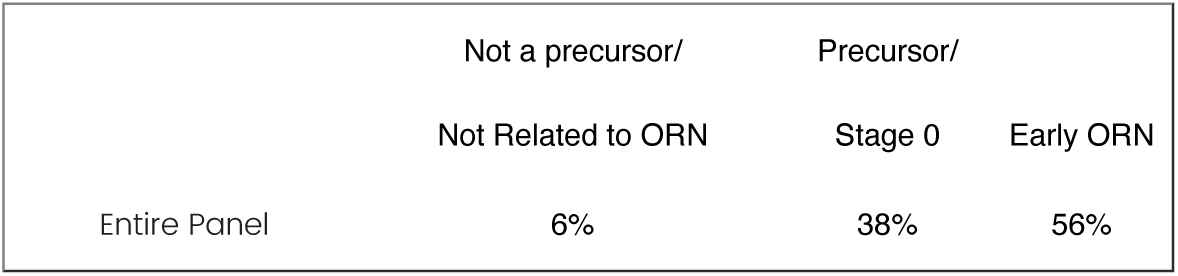

Classification per specialty:

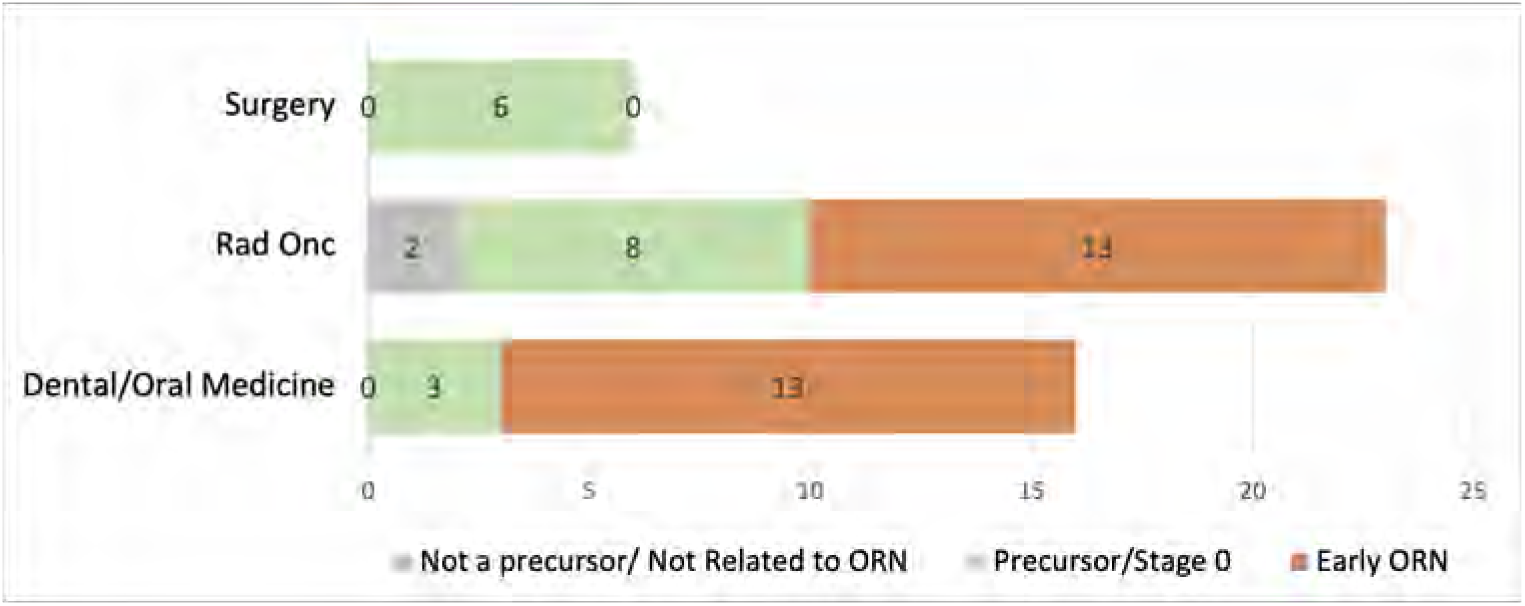

*Specialty specific classifications meeting consensus threshold:*

- *Surgery: 100% favoring precursor / Stage 0*
- *Dental/Oral Medicine: 81% favoring early ORN*

How should this feature be classified?

- Precursor / Stage 0
- Early ORN

Group feedback for: Probe-to-bone POSITIVE, any imaging finding (EXCEPT periosteal reaction) within alveolar bone

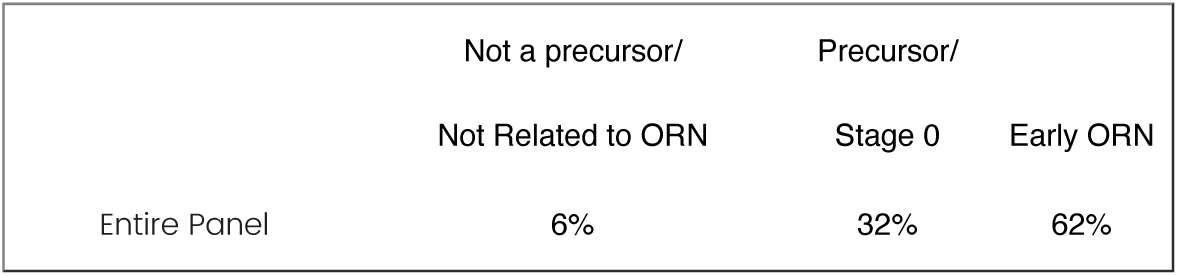

Classification per specialty:

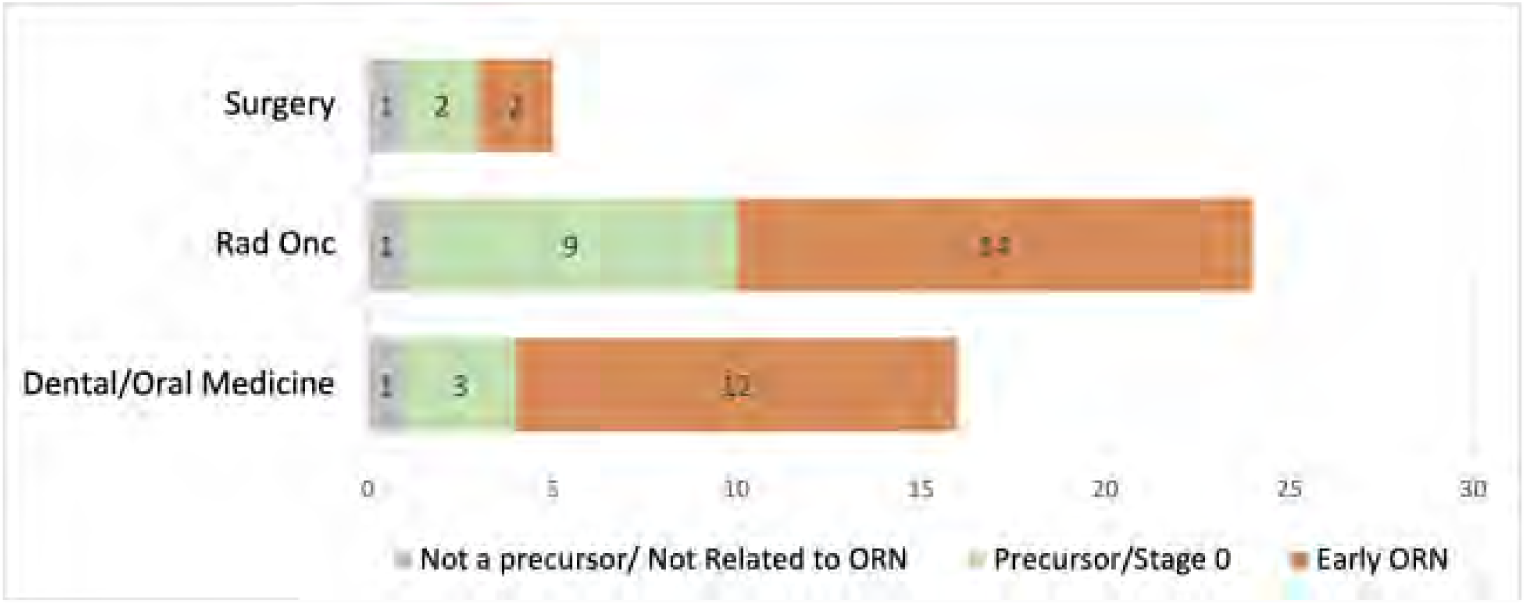

*Specialty specific classifications meeting consensus threshold:*

*- Dental/Oral Medicine: 75% favoring early ORN*

How should this feature be classified?

- Precursor / Stage 0
- Early ORN

#### Vascular Damage on MRI

Panelists were asked to classify two different scenarios whereby vascular damage could be seen on surveillance MRI after RT.

<u>Scenario 1:</u> How should we classify a case with vascular damage in bone seen on MRI without exposed bone and without other imaging findings (i.e., CT shows no bony abnormalities)?

Group feedback: 21 (41%) classified it as *Not a precursor/Not related to ORN*, and 29 (57%) classified it as *Precursor to ORN/Stage 0*.

Please reclassify this scenario

- Not a precursor / Not related to ORN
- Precursor / Stage 0

#### Vascular Damage on MRI

<u>Scenario 2:</u> How should we classify a case with vascular damage in bone seen on MRI without exposed bone and with other imaging findings limited to alveolar bone (i.e., x-ray shows sclerosis limited to alveolar bone)?

Group feedback: 26 (51%) classified it as *Precursor to ORN/Stage 0,* and 19 (37%) classified it as *Early ORN*.

Please reclassify this scenario

- Precursor / Stage 0
- Early ORN

### 10 Case Review

#### Review of 10 Cases with Clinical & Imaging Features

Round 3 ended with the panel reviewing 10 cases which included a clinical description and an image. Experts were asked to 1) stage the case, 2) report extent of bone involvement on imaging, and 3) rate their level of confidence (LOC).

The figure below summarizes the group stage classification (Fig A) and reporting of extent of bone involvement (Fig B) per case. The stacked bars and numbers represent percentages of the panel. Results for Case 2 (clinically exposed bone and pathological fracture on imaging) shows consistency with the panel classifying pathologic fracture as advanced ORN with bony involvement beyond the alveolar bone (AB). However, we can appreciate significant variations in staging assignment for the other cases. This is also true within specialties (figures not shown).

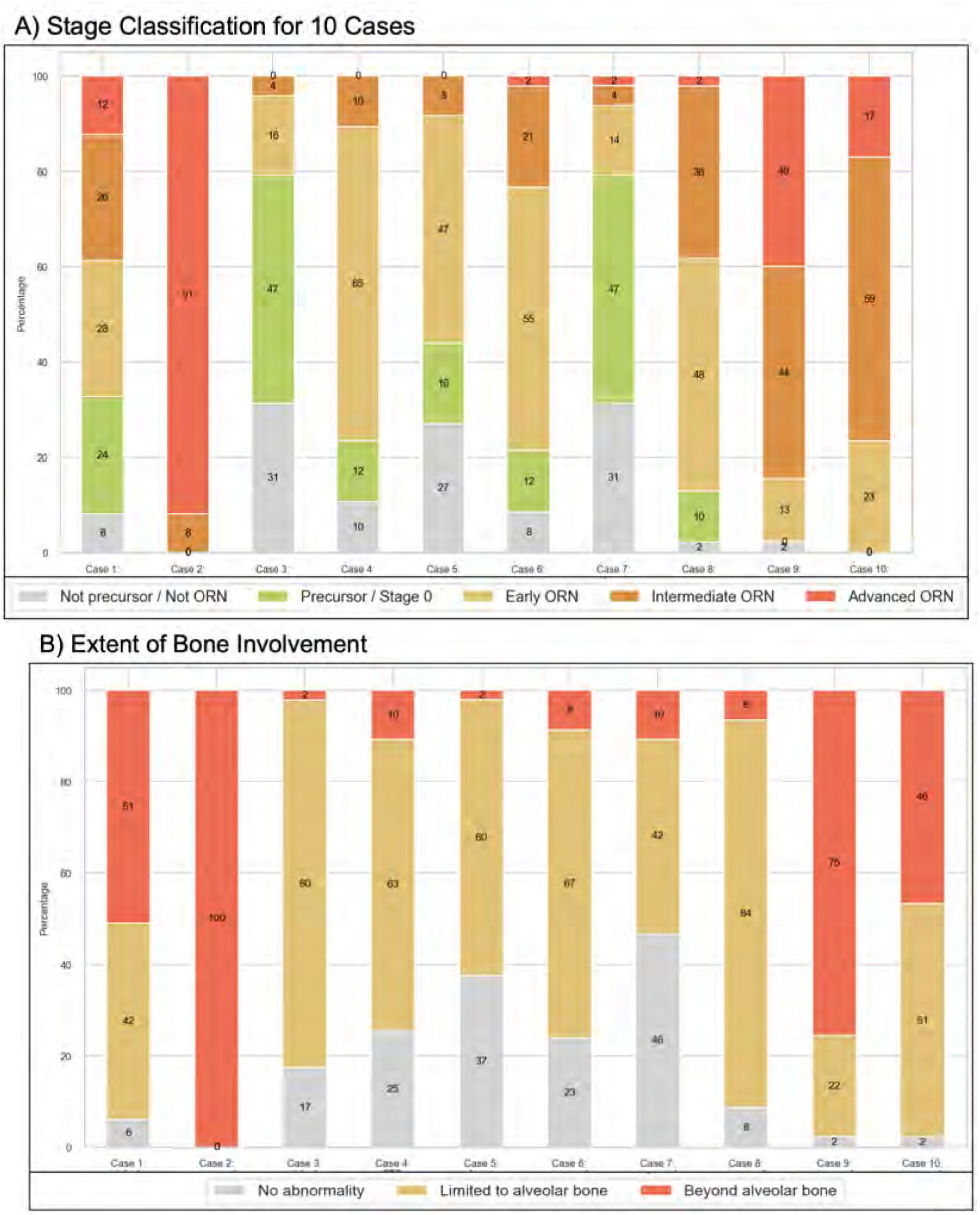

The LOC also varied among cases and specialties with the exception of Case 2. Note, Dental specialists all reported a LOC of 5 for Case 2.

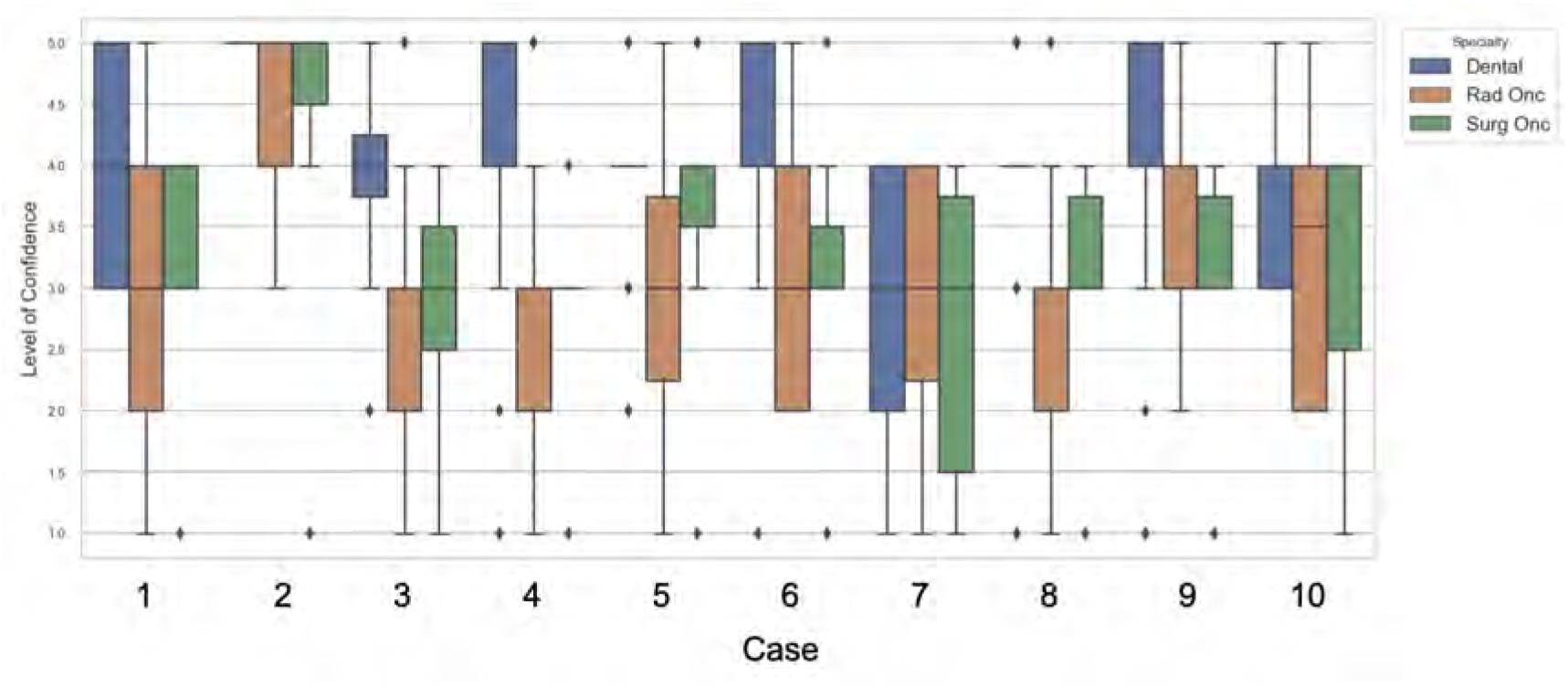

We will now re-review each case (except Case 2) and classify them based on the top 2-3 choices (i.e., minimum to reach 70%) selected during Round 3. Of note, some cases may not ask a question on extent of bone involvement as the panel already met consensus for that question. For cases with images, we will also ask you to document what radiographic features you observe (i.e., periodontal ligament space widening, sclerosis, lytic changes, periosteal reaction, etc). Please answer to your best ability as these details will be important to incorporate in an ORN ontology in the future.

Please right click on the links below to open educational resources on dental/mandibular changes on panoramic imaging after HN RT.

[Mandibular changes on panoramic imaging after HN RT](https://mdanderson.co1.qualtrics.com/CP/File.php?F=F_Ai49b0AMZ3jfyD6)

[Imaging of Radiation and Medication-related Osteonecrosis](https://mdanderson.co1.qualtrics.com/CP/File.php?F=F_izSxrhOO9LddqAn)

*If the pdfs above to not work, please use the following links:*

[Mandibular changes on panoramic imaging after head and neck radiotherapy](https://www.sciencedirect.com/?ref=pdf_download&fr=RR-11&rr=87291df299b57fc8)

[Imaging of Radiation- and Medication-Related Osteonecrosis](https://www.sciencedirect.com/?ref=pdf_download&fr=RR-11&rr=87291e3b5d1a7fc8)

## Case 1

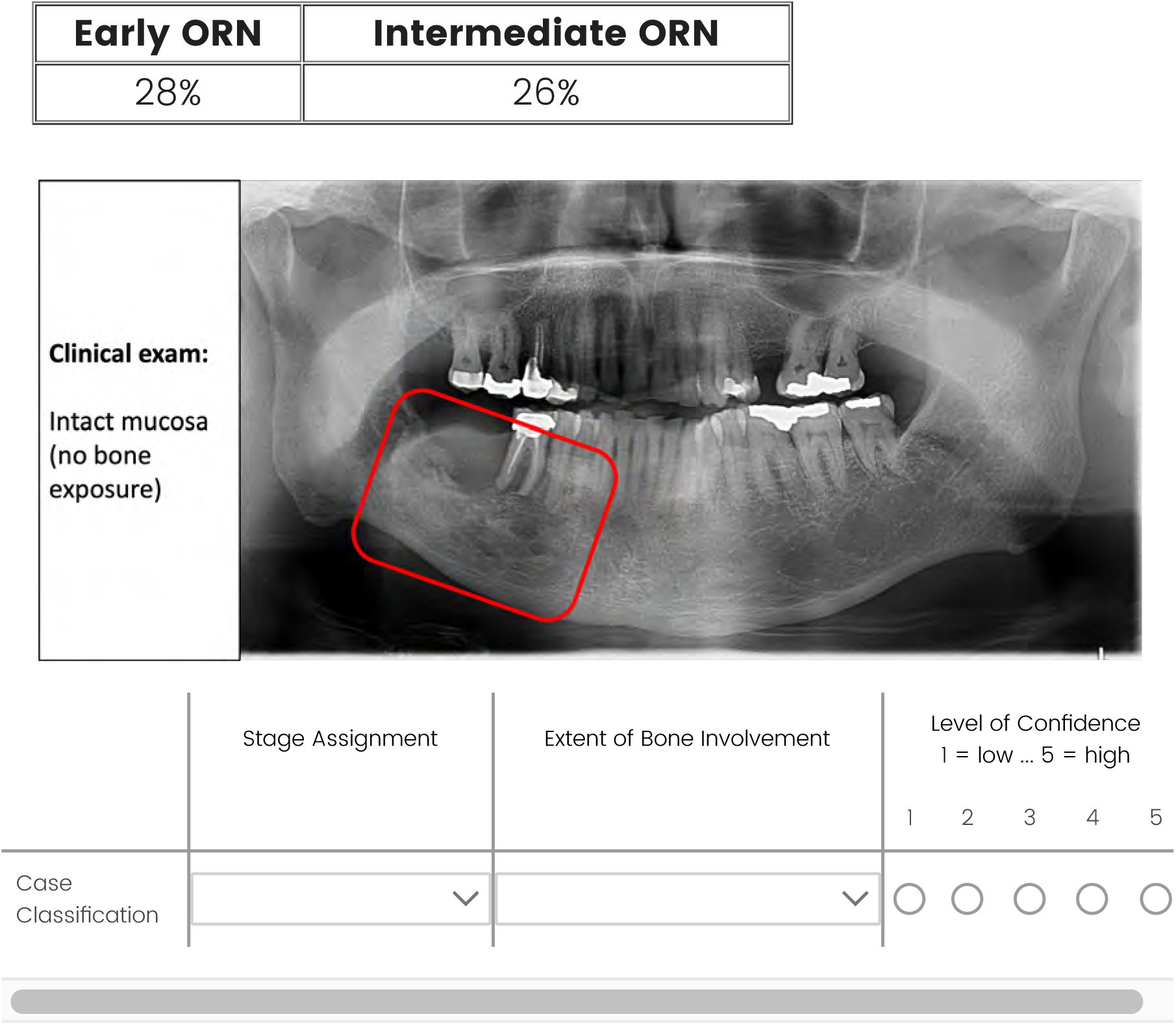

What radiographic features do you observe in Case 1? Type

**NA** if no abnormalities are seen.

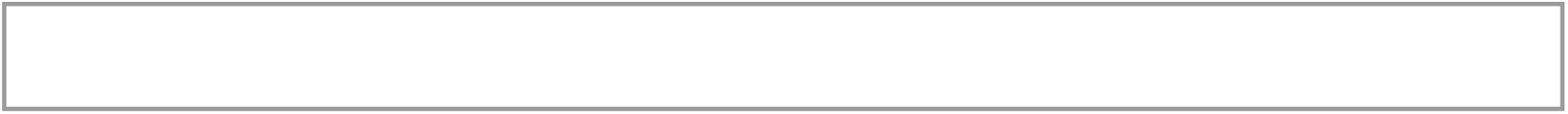

## Case 3 and 11 (1-year follow up)

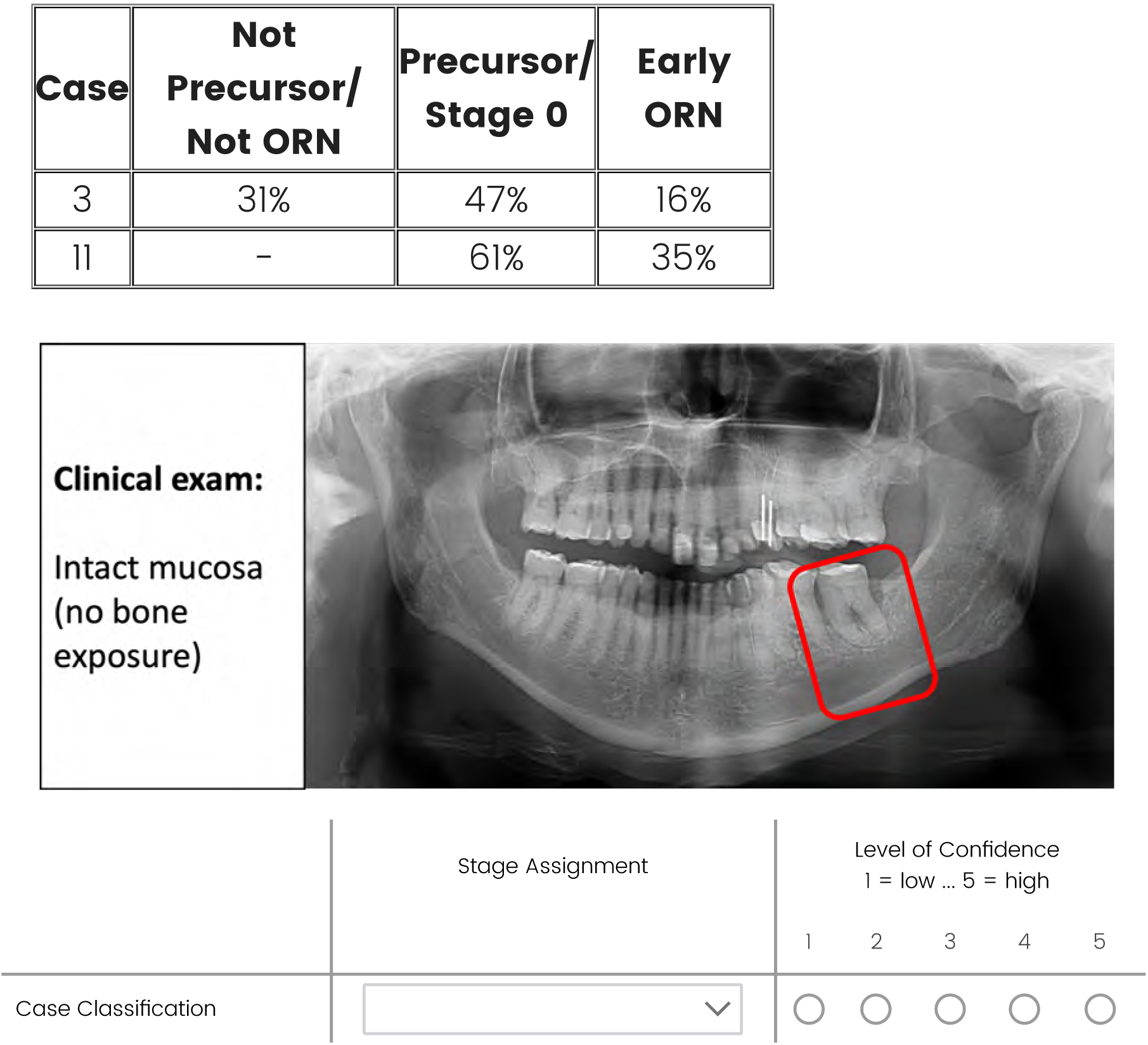

What radiographic features do you observe in Case 3? Type

**NA** if no abnormalities are seen.

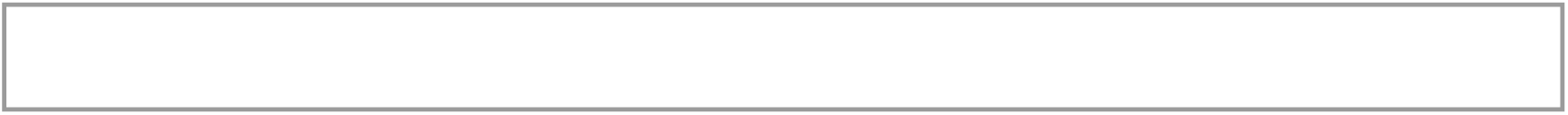

## Case 4

PTB test is positive.

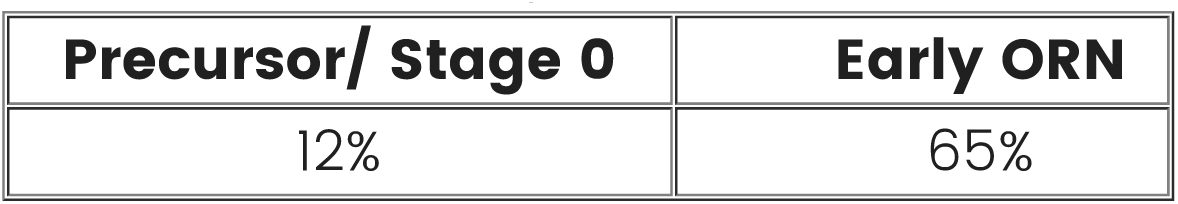
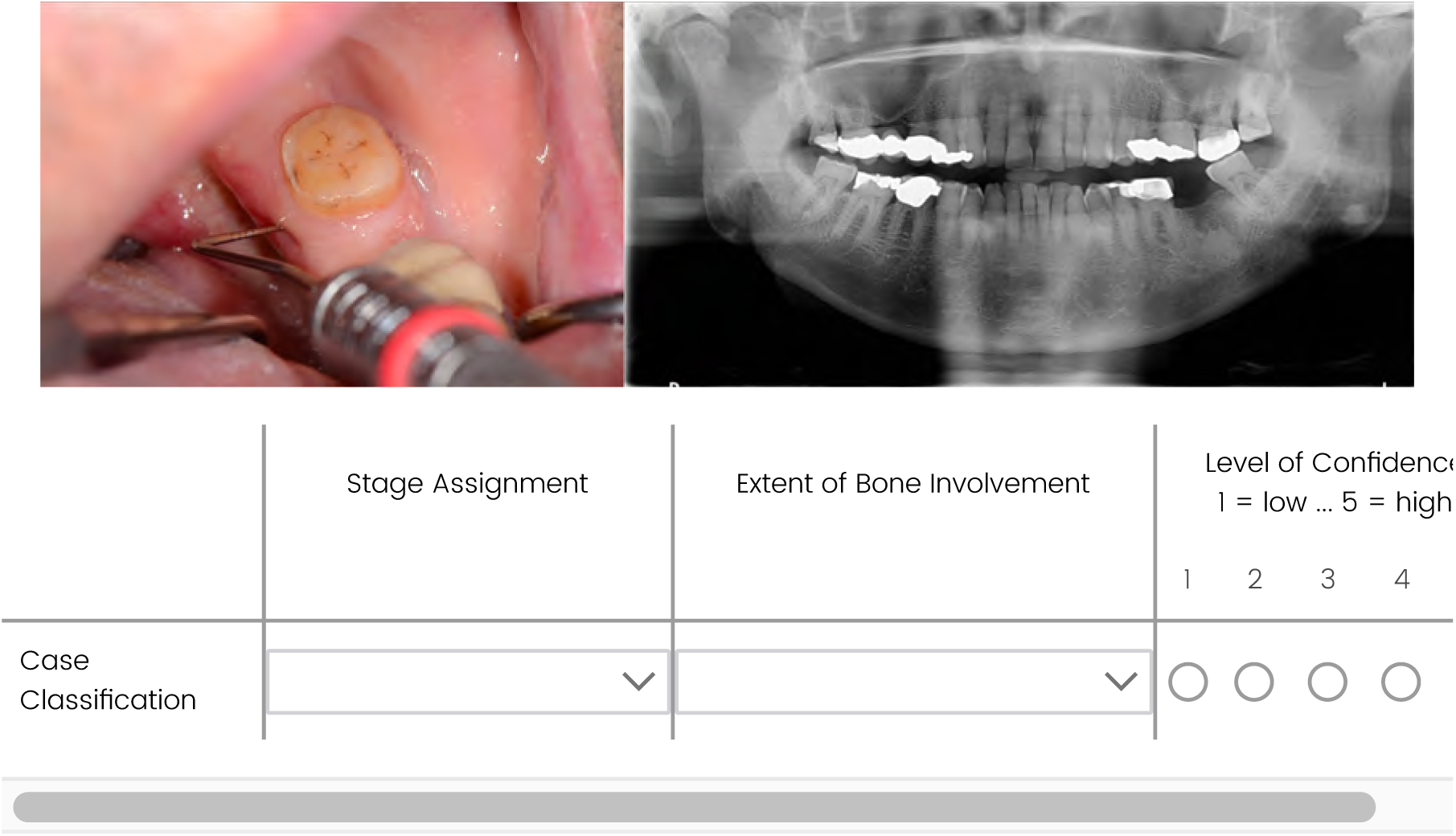

What radiographic features do you observe in Case 4? Type

**NA** if no abnormalities are seen.

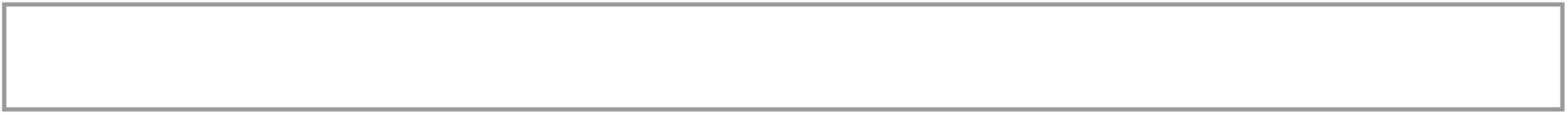

## Case 5

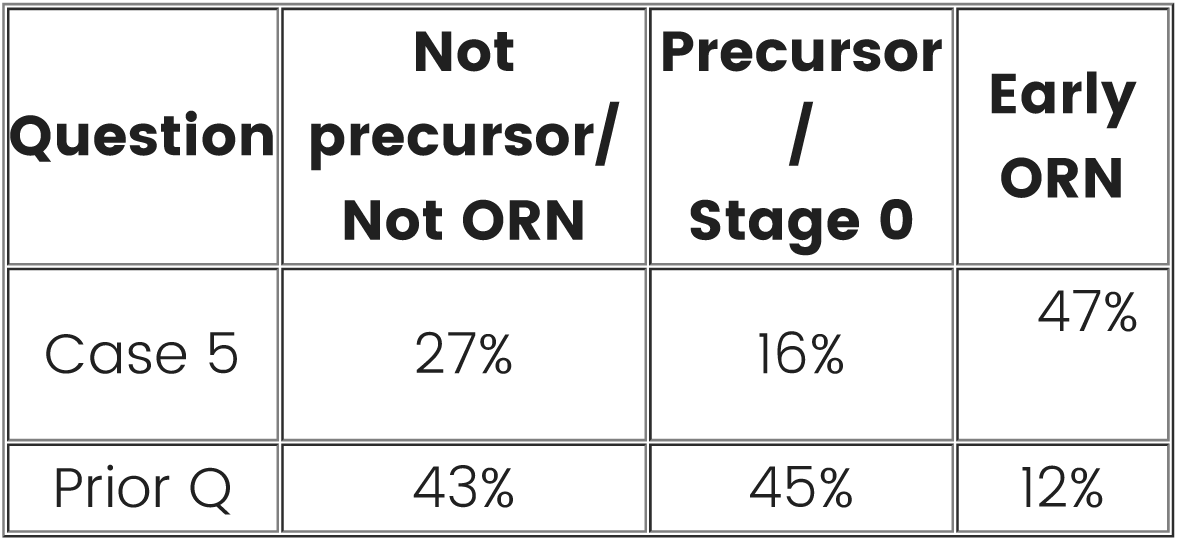
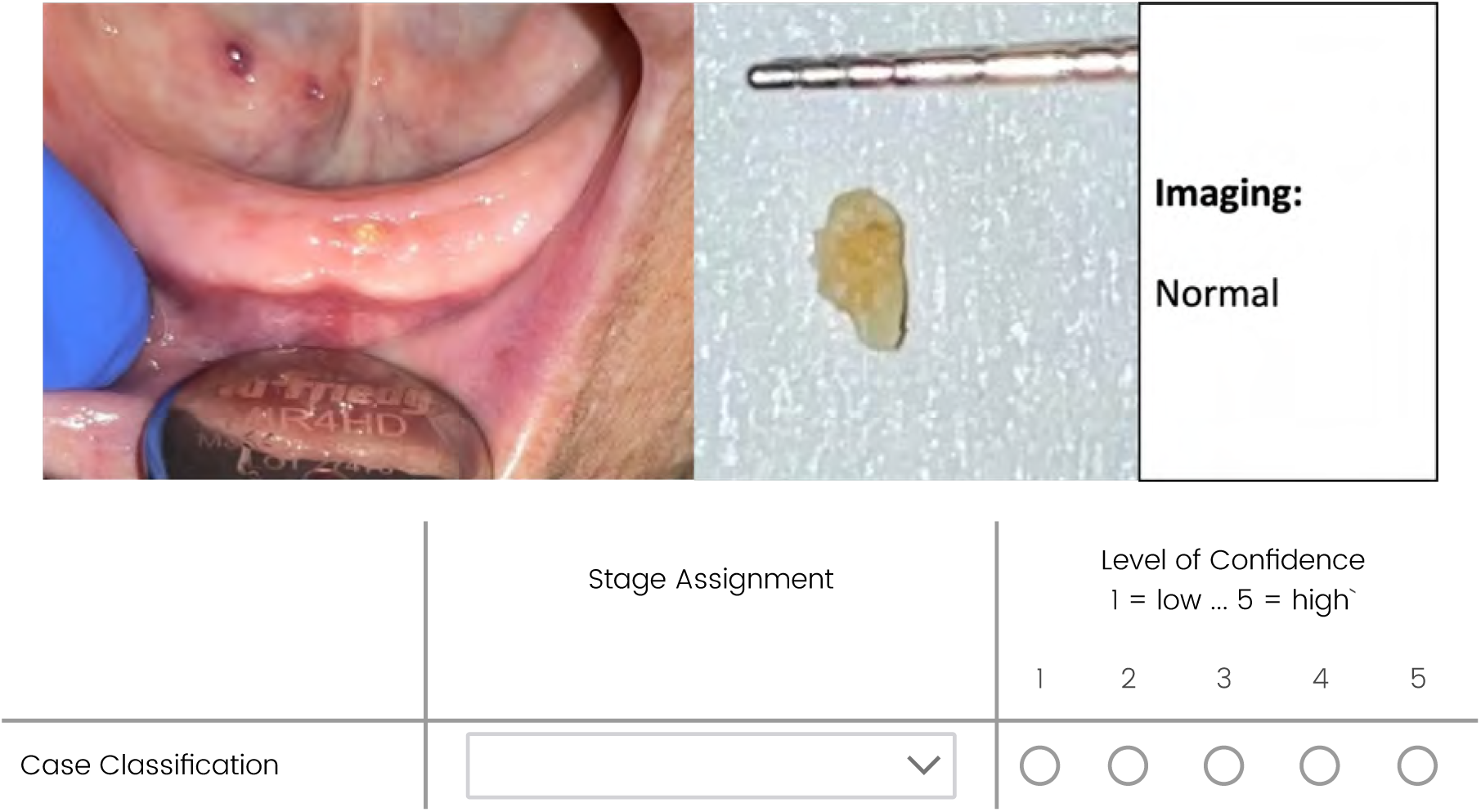

## Case 6

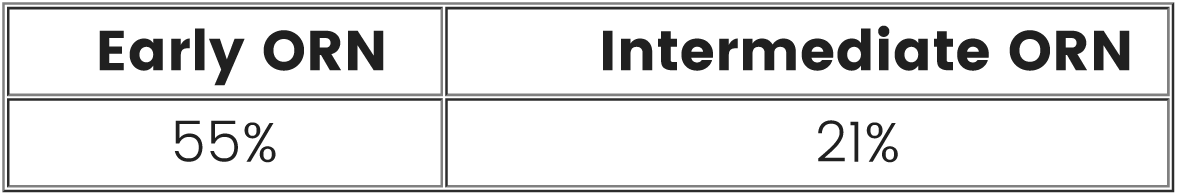

Note, ‘exposed necrotic bone, any image finding in alveolar bone’ was classified as Early stage ORN by 100% of the panel over precursor/Stage 0.

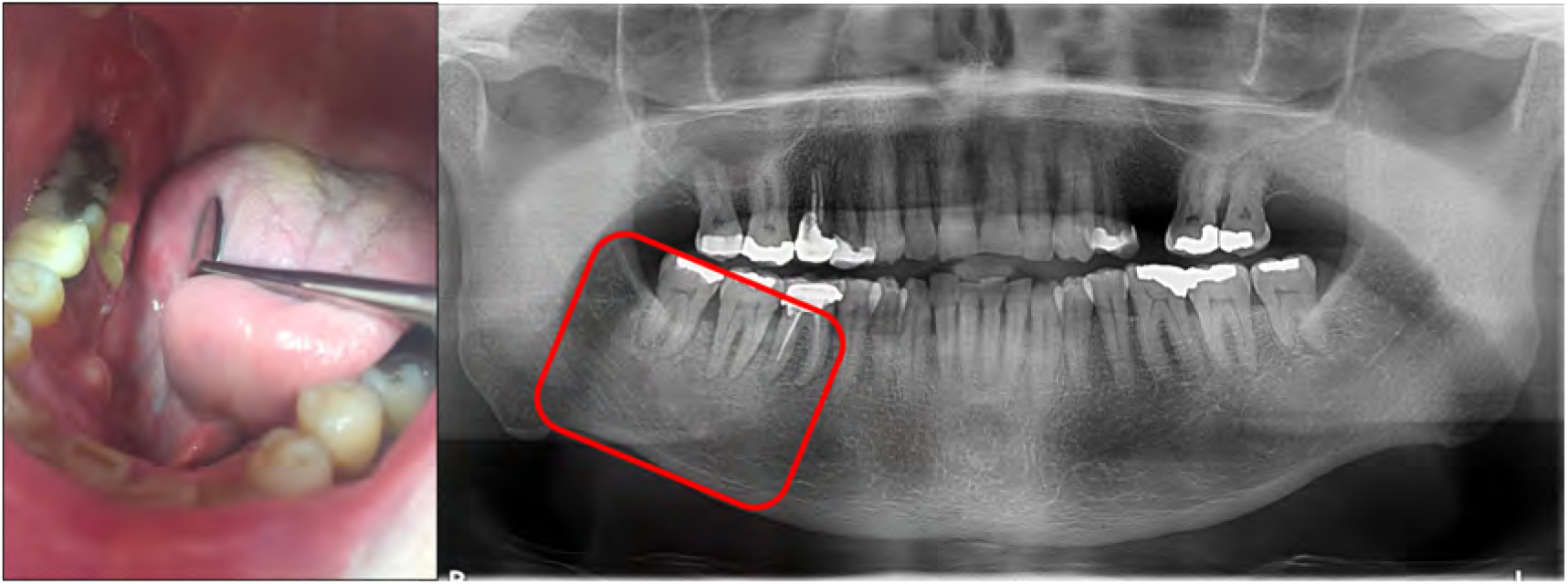
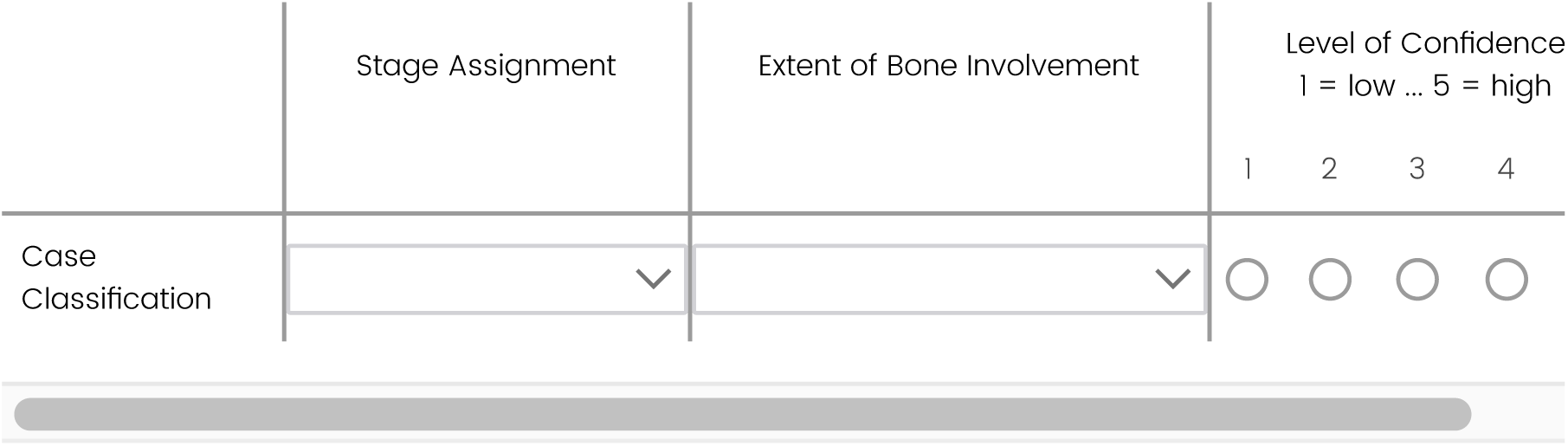

What radiographic features do you observe in Case 6? Type

**NA** if no abnormalities are seen.

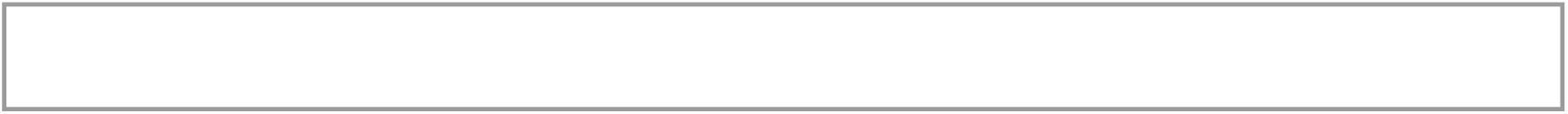

## Case 7

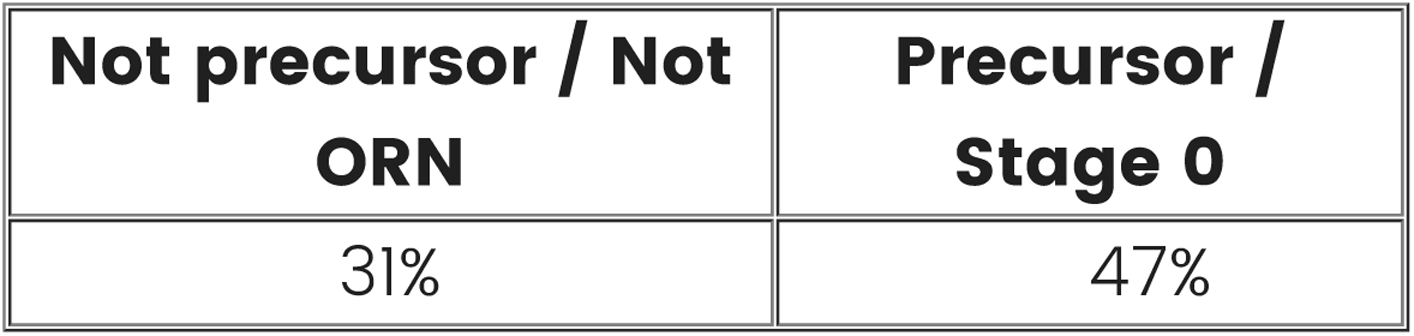
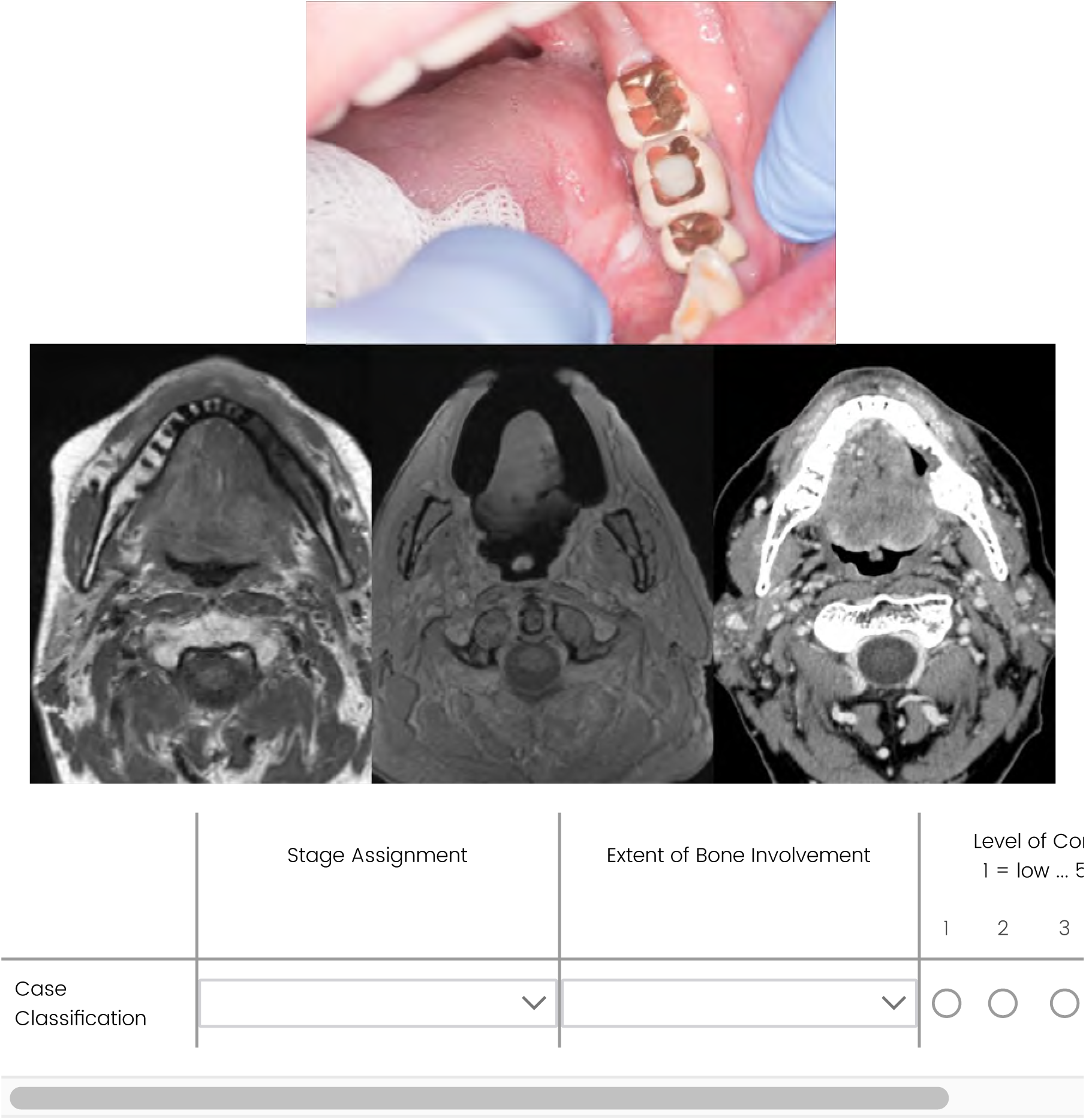

What radiographic features do you observe in Case 7? Type

**NA** if no abnormalities are seen.

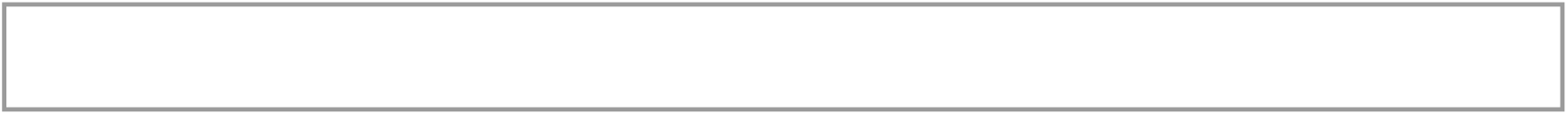

## Case 8

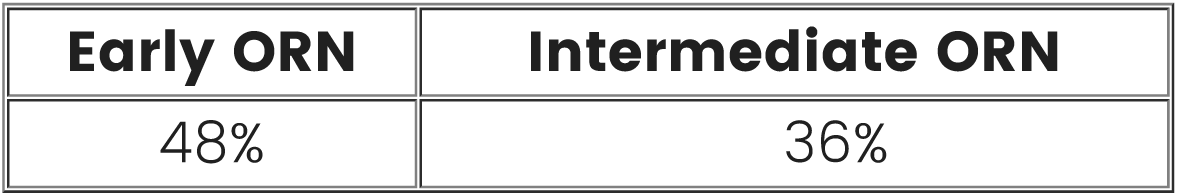
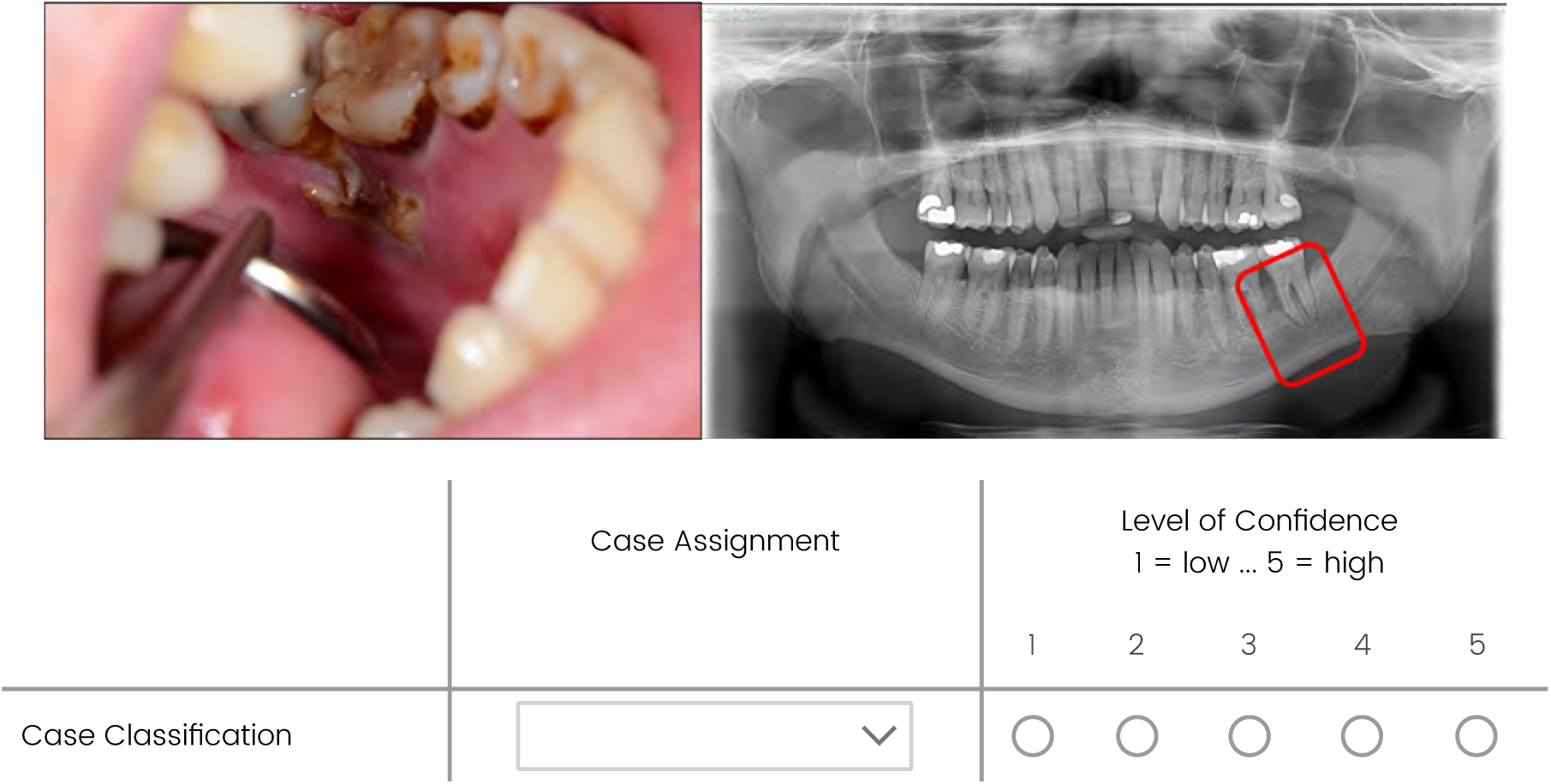

What radiographic features do you observe in Case 8? Type

**NA** if no abnormalities are seen.

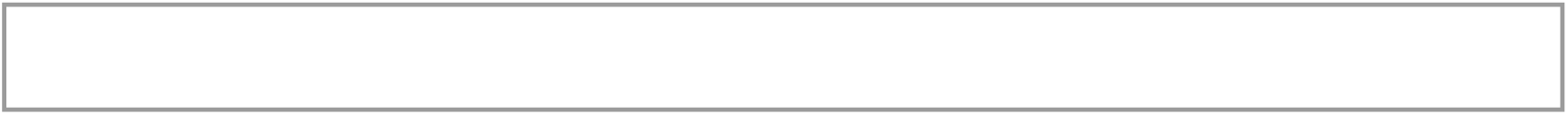

## Case 9

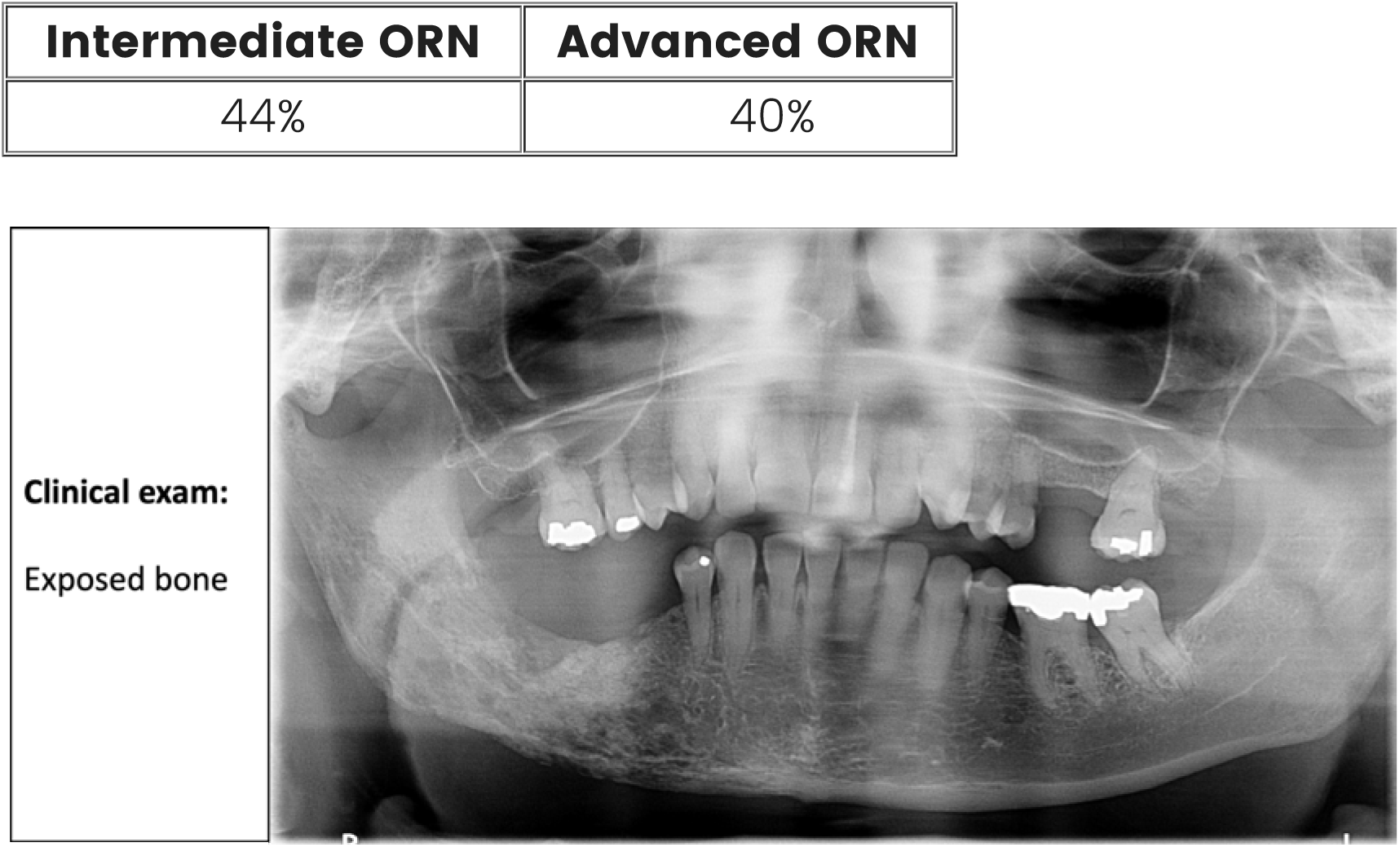
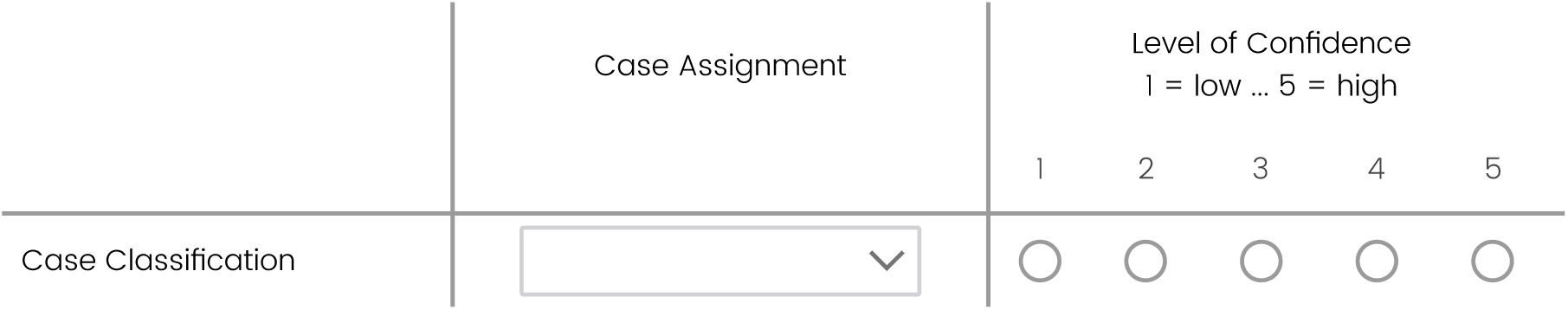

What radiographic features do you observe in Case 9? Type

**NA** if no abnormalities are seen.

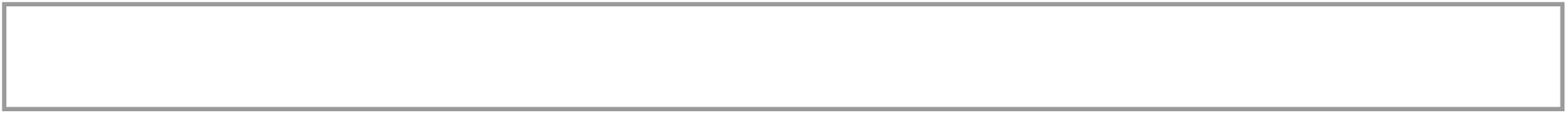

## Case 10

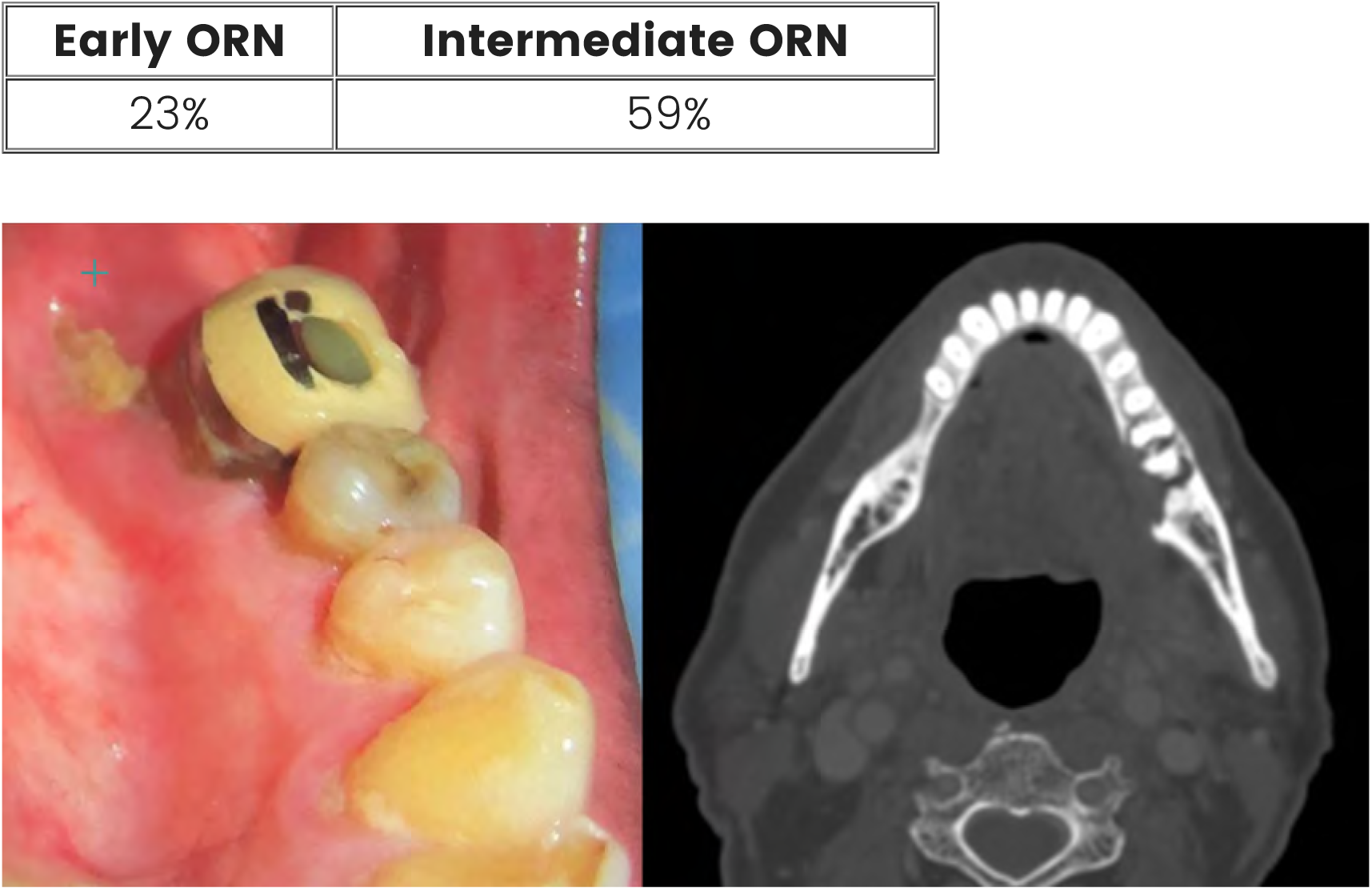
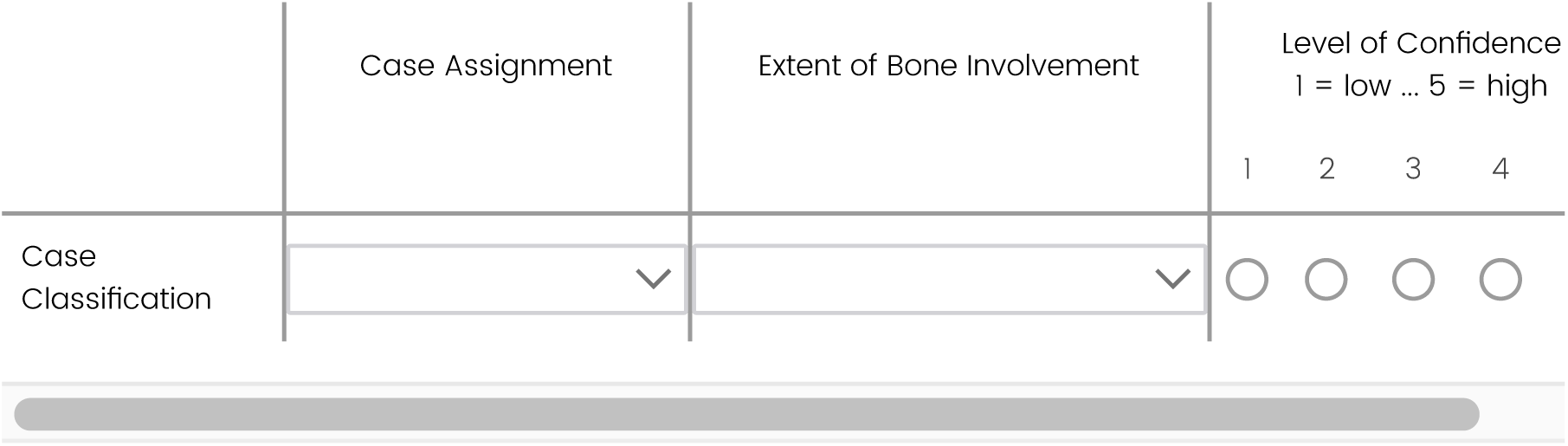

What radiographic features do you observe in Case 10? Type

**NA** if no abnormalities are seen.

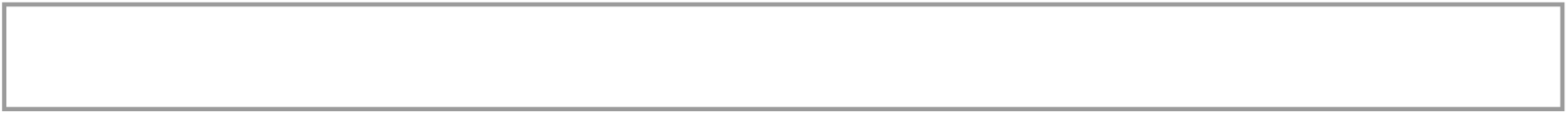

## New Case 12

Imaging: sclerotic changes within the alveolar bone

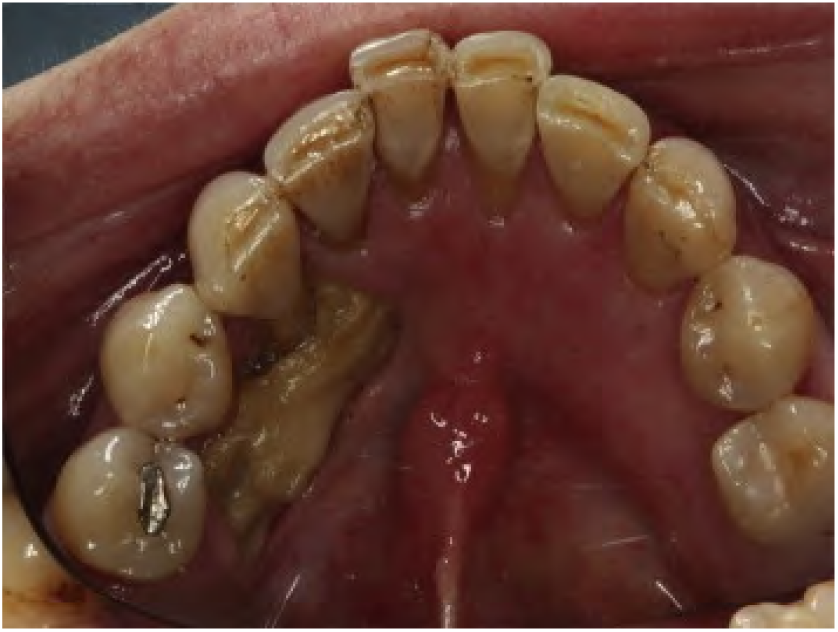

- Early ORN
- Intermediate ORN

## New Case 13

Imaging: Lytic changes beyond the alveolar bone

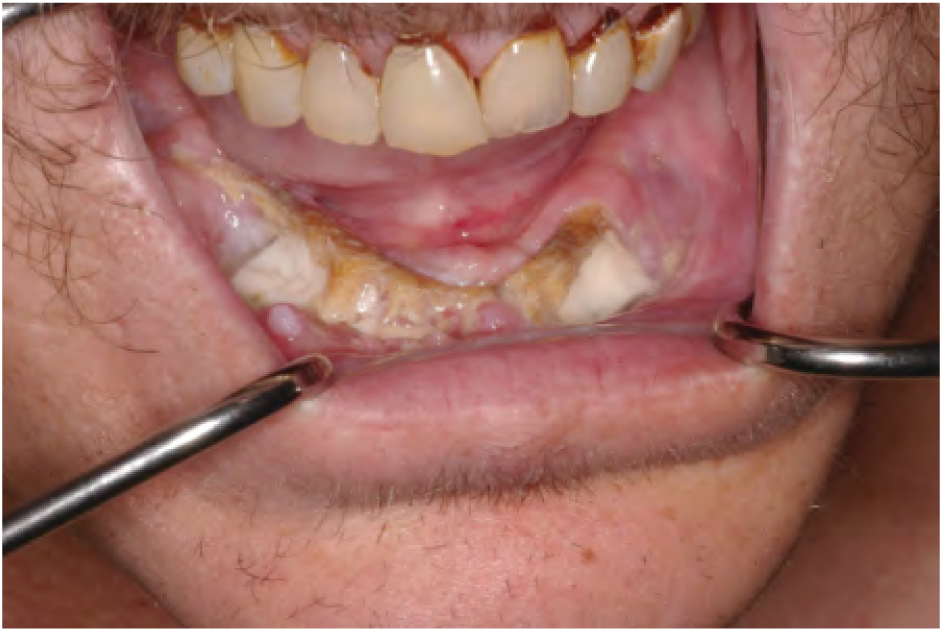

- Intermediate ORN
- Advanced

Did you find the educational resources on panoramic x-ray dental changes informative for your assessment of cases with dental imaging?

- Yes
- No
- I did not open the links

Would you be interested in an educational resource with more detailed multidimensional (i.e., CT, MRI, panoramic xray) imaging features explained to demonstrate the full spectrum of ORN and its precursor stages?

- Yes’
- No

Any additional commentary in preparation of our manuscript?

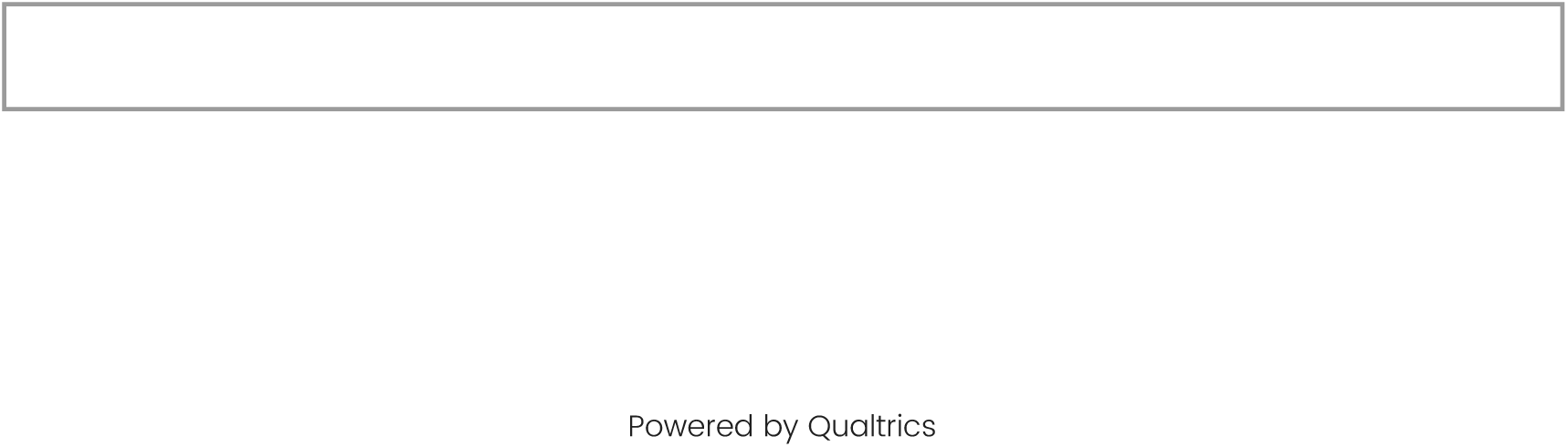

Powered by Qualtrics

## S3_Methods

The process flow chart for this study is shown in Figure 1. A group of international multidisciplinary oncology and dental specialists were sent an electronic invitation to participate in the study (n=75). Members of the ORAL Consortium participated in at least one Delphi survey (n=69). ACM and CDF served as study coordinators and participants in the ORAL Consortium with no extra oversight voting rights throughout the study. No financial incentives were provided to participants. Surveys were developed in REDCap® and Qualtrics (see Supplementary Materials for all questionnaires) and sent to experts using personalized links to preserve annonymity.^1,2^ Survey instruments were piloted by the study coordinator (ACM) to ensure correct formatting and delivery. Every questionnaire was in English and included an introduction, primary objectives for the round, and aggregated group feedback for consensus building. After each round, items meeting consensus were reported in following rounds as “consensus reports” and no further questions were asked on those items. Study preparation began in January 2023 with round 1 launching in March 2023, round 2 in May 2023, round 3 in August-September 2023, round 4 in February 2024, and manuscript review in March 2024 with final approval completed in April 2024. No deviations to study protocol were performed (MDA PA 2020-1096).

A comprehensive literature review was performed as preparatory research. Specifically, a PubMed search was performed in February 2023 for “(osteoradionecrosis) AND (staging OR grading)” which identified 447 publications since 1973 to 2023. Titles were screened for relevance resulting in abstract screening of 193 articles. Further screening was performed on full manuscript of original articles or reviews on one or more staging/grading systems for ORN/ORNJ. This resulted in the formal review and rating of 15 grading/staging systems by the ORAL Consortium. Round 1 included questions on definition preference, data element consideration within a formal ORNJ definition, level of agreement with each system, user preference, and rated effectiveness of systems for classifying ORNJ. Based on features identified in published systems, experts were also asked to classify the following three cases using 14 published staging systems: 1) *a patient with exposed bone (no measurement) not involving lower mandible, unknown duration. Pain present*, 2) a *patient with 1.2cm exposed bone for 4 months, pain present*, and 3) *a patient with 3cm exposed bone with pathologic fracture. Unknown symptoms or duration*. Imaging correlates of ORNJ were also introduced, and experts rated the level of importance of individual data elements (i.e., ulceration, exposed bone, pathologic fracture) on a 3-point ordinal scale matrix (i.e., not important, somewhat important, very important).

The second questionnaire provided anonymized group feedback on existing ORNJ definition rankings and asked experts whether the Consortium’s definition for ORNJ should be in alignment with existing hierarchical international terminologies such as the Medical Dictionary of Regulatory Activities (MedDRA),^3^ the Systematized Nomenclature of Medicine-Clinical Terms (SNOMED-CT),^4^ and the International Classification of Disease (ICD),^5^ the latter which had diagnostic codes for osteonecrosis as a general condition, but not ORNJ at the time of the study (ICD-10). Group feedback was both quantitative (i.e., ranked items) and qualitative (i.e., thematic grouping of commentary). Further clarification on the importance of a time feature for diagnosing ORNJ was asked, and experts reviewed and selected the best of four proposed ORNJ definitions. Open-ended questions were included for experts to provide commentary. Additional scenarios describing explicit combinations of clinical exam and imaging findings were presented for more granular stratification of *not related to ORNJ* versus *early/limited* and *advanced ORNJ* features. Of note, rounds 1 and 2 also included questions related to the parallel development of an information tool for visual representation of received radiation doses onto an odontogram (results not reported in this manuscript).

The third questionnaire presented several consensus statements to the Consortium related to what definitional concepts and criteria should and should not be considered mandatory for diagnosing ORNJ, and how implement the Common Terminology Criteria for Adverse Events (CTCAE) grading system when assessing patients for ORNJ.^6^ Case review summaries were provided, and features rated as *early/limited ORNJ* were refined on a spectrum of *not related to ORNJ*, *precursor/stage 0*, and *early ORNJ* while *advanced ORNJ* features were reclassified as *intermediate ORNJ* or *advanced ORNJ*. Ten new cases with 2-dimensional (i.e., orthopantogram or OPG) or 3D (i.e., computed tomography [CT], magnetic resonance imaging [MRI]) diagnostic imaging snapshots were included for 1) stage assignment, 2) reporting extent of bone involvement as seen on imaging (no abnormality, limited to the alveolar bone [AB], or beyond the AB), and 3) expert-rated level of confidence (LOC) on a 5-point scale (1=low, 5=high). Given recent publications of early MRI-based imaging biomarkers for vascular damage and potential precursor stages of ORNJ,^7^ experts also provided opinion on how to classify vascular damage findings in the presence or absence of additional clinical and imaging findings.

Items on the fourth survey focused on finalizing a consensus-derived definition of ORNJ and reclassifying all cases that did not meet consensus during round 3. Prior to reclassifying cases, the ClinRad model,^8^ a recently published risk-based classification system for ORNJ endorsed by ASCO/IAOO/MASCC expert guidelines,^9^ was presented as well as two educational resources on how to interpret abnormal radiographic imaging findings.^10,11^ Two additional cases with ‘extensive bone exposure’ seen on a clinical photograph and reported imaging findings within (Case 12) or beyond (Case 13) the alveolar bone were also reviewed to investigate the impact of clinical photographs on staging. Lastly, using a yes/no question format, experts were asked if they 1) found the educational resources on dental imaging informative for case review, and 2) whether they would be interested in having an educational resource with more detailed multidimensional (i.e., CT, MRI, panoramic x-ray) imaging features explained to demonstrate the full spectrum of ORNJ and its related precursor stages.

Descriptive statistics were performed to summarize the International ORAL Consortium expert demographics, staging systems review, and case classification as a collective cohort and by grouped specialties. During the second and third rounds, high-yield topics not meeting a pre-defined consensus threshold of 70% were represented in round 4 with a lowered consensus threshold of 60%. Experts were informed of thresholds prior to initiating each survey. The Fleiss’ kappa statistic^12^ was used to assess inter-rater agreement between all experts (n>2) when classifying the ten image-based cases on a categorical scale which included *Not precursor/Not ORNJ, Precursor/Stage 0, Early ORNJ, Intermediate ORNJ, Advanced ORNJ*. Interpretation of the kappa value is as follows: <0, poor agreement; 0.0-0.20, slight agreement; 0.21-0.40, fair agreement; 0.41-0.60, moderate agreement; 0.61-0.80, substantial agreement; and 0.81-1.0, almost perfect agreement.^13^ Only ACM had access to individual panelist response data which was used to summarize group feedback for presentation to the panel on subsequent rounds. All calculations and graphs were performed using R statistical software (Version 2023.06.1+524, R Foundation for Statistical Computing, Vienna, Austria; packages hmisc, table1, irr) and Python/Visual Studio Code.^14,15^

## S4_Results

### Staging Data Element Extraction and Classification

During round 1, a data element tracker flowsheet was developed for reporting all clinical, radiographic, therapy, and treatment response data elements identified in all staging/grading systems (Figure S3). Experts were then asked to rate the importance of each feature (not important, somewhat important, very important). Figure S4 shows the distribution of expert responses with only 3 data elements being considered somewhat/very important by 100% of the group: pathologic fracture, extent of bone involvement, and exposed bone. Other clinical (or radiographic) features achieving >70% rating of importance included orocutaneous fistula (98%), [mucosal] ulceration (85%), sinus formation (82%) and sequestra (81%). All remaining clinical/radiographic features underwent a first pass classification by experts into the following categories-*Not needed for ORNJ staging, Early/Limited ORNJ,* and *Advanced ORNJ* (Figure S5). Five features met high consensus (81-97%) while 2 were equivocal (non-exposed bone/imaging findings beyond the AB; sequestra) and 4 were considered unnecessary for ORNJ staging including bone spicules. All these initially classified features, in addition to dental-based clinical scenarios with probe-to-bone [PTB] assessments (Figure S6), were reclassified in round 3 into tiered groups as follows: Group 1 [*Not a precursor/Not related to ORNJ; Precursor/Stage 0; Early ORNJ*], Group 2 [*Early ORNJ; Intermediate ORNJ*], and Group 3 [*Intermediate ORNJ; Advanced ORNJ*]. Items or cases not meeting the consensus threshold during round 3 were repeated in round 4. Additionally, 10 imaged-based cases (i.e., clinical photographs and/or 2D/3D radiographic images) were reviewed during the last two surveys to reinforce classification of MDE combinations (Figure 4).

#### Not a Precursor / Not related to ORNJ

- *PTB test <u>negative</u> with periosteal reaction <u>within</u> AB seen on imaging:* A comparable division in classifying this feature was noted during round 3 (47% *Not a precursor*) and round 4 (50% *Not a precursor*). RadOncs were the drivers of classifying this feature as a precursor stage compared to surgeons and dentists who favored it as being unrelated. The Consortium recommends this feature combination to be considered as unrelated to ORNJ to differentiate it from consensus-approved precursor features. Of more importance, 2 MDEs are valuable in this description and include **clinical:PTB_test_result** (positive or negative) and **imaging:morphology** (i.e., periosteal reaction, sclerosis, lysis). The PTB test is a commonly utilized dental probing exam that provides diagnostic information on periodontal health;^1,2^ therefore the Consortium recommends standardized documentation of this procedure whenever performed. Moreover, these MDEs are required for other upstaging feature combinations.

#### Precursor / Stage 0

- *Intact mucosa (no clinical bone exposure) with any imaging findings <u>within</u> AB*: This text-only description met high consensus during round 4 with 85% (45/53) of experts favoring a precursor assignment over being unrelated to ORNJ. Final round classification of image-based Case 3 further supported this designation with consensus being met at 67% (33/49). Identified MDEs are **clinical:mucosal_status** (i.e., intact, ulcerated) and the dichotomous **imaging:vertical_ab_abnormality** (i.e., within/above AB, beyond/below AB).
- *Minor bone spicules (MBS)*: This feature posed a significant challenge for classification across all rounds using text-only or image-based (Case 5) descriptions. When asked to classify MBS (text-only) during round 3, there was a slight preference for precursor stage (45%, favored by surgeons) over being unrelated to ORNJ (43%, favored by dentists; RadOncs equivocal) or representing early ORNJ. In round 4 (when *Early ORNJ* was removed as an option), the precursor status was again favored at 55%. However, when presented with clinical photography indicative of MBS, there was consistent upstaging of MBS to *Early ORNJ* in both rounds (49% and 50% *Early ORNJ*) with less than 30% assignment to precursor or unrelated stages. To mitigate classification ambiguity when clinically exposed bone is detected, serial clinical photographs and quantitative measurements of exposed bone (MDE **clinical:exposed_bone_length_in_mm**) are strongly recommended. An additional MDE for **minor_bone_spicules** is proposed to explicitly report MBS which should be accompanied by a quantitative measurement.
- *PTB test <u>negative</u> with any imaging findings (EXCEPT periosteal reaction) <u>within</u> AB seen on imaging*: The Consortium achieved consensus during the final round for this feature combination (76%, 41/54). Associated MDEs are **clinical:PTB_test_result** (negative), **imaging:morphology,** and **imaging:vertical_ab_abnormality** (within AB).
- *Vascular damage in bone seen on MRI <u>without</u> exposed bone and <u>without</u> other imaging findings (i.e., CT shows no bony abnormalities):* This scenario is considered a precursor stage as it met consensus during the final round as a text-only description (60%, 32/53) and in Case 7 which depicted minor mucosal ulceration (i.e., no bone exposure) with reported dynamic contrast-enhanced (DCE) MRI changes (63% agree, 31/49).
- *Vascular damage in bone seen on MRI <u>without</u> exposed bone and <u>with</u> other imaging findings limited to AB (i.e., x-ray shows sclerosis limited to AB):* The Consortium demonstrated consensus convergence with classifying this combination as a precursor stage (78%) over early ORNJ. This is also in alignment with classification of cases of intact mucosa with abnormal imaging findings restricted within the alveolar bone.

#### Early Stage ORNJ

- *PTB test <u>positive</u> with imaging findings <u>within</u> AB seen on imaging:* Compared to intact mucosa classification, a positive PTB test was considered to be an upstaging feature by the Consortium when combined with localized periosteal reactions (91%, 49/54) or other morphological radiographic changes (89%, 48/54) limited to the alveolar bone. This classification was supported by Case 4 which showed a photograph of a positive PTB test and OPG changes within the AB (86%, 43/50, staged it as *Early ORNJ*). Associated MDEs are **clinical:PTB_test_result** (positive), **imaging:morphology,** and **imaging:vertical_ab_abnormality** (within AB).
- *Exposed bone with any imaging findings <u>within</u> AB*: During round 3, this text-only presentation was unanimously classified as an *Early ORNJ* when limited to options ranging within Group 1. When asked to classify Case 6, a photograph with clinical bone exposure spanning the width of one molar and an OPG image showing periodontal ligament space widening, experts converged to consensus in assigning this feature combination as *Early ORNJ* (round 3, 55%; round 4, 74%). While these examples are in alignment with the ClinRad classification model for ORNJ,^3^ it became evident that <u>the extent of visualized bone exposure influences stage designation</u>. For example, when presented with another case with similar imaging findings but extended clinical bone exposure spanning the length of two molars (Case 8), there was reduced consensus at classifying it as *Early ORNJ* (64%) over *Intermediate ORNJ* (36%). Case 12 showing clinical bone exposure beyond two teeth with text-only ‘reported imaging findings within AB’, was presented once at the end of round 4 after review of the ClinRad model. The clinical photograph influenced the upstaging of this case of extensive bone exposure to *Intermediate ORNJ* (76%, 39/51). To reduce mis-classification risks for the same or nearly identical patients, the Consortium again strongly recommends the use of a quantitative MDE (i.e., **clinical:exposed_bone_length_in_mm**) for facilitated data harmonization across disciplines and evolving staging systems. Using predefined cutoffs (i.e., </> 2cm) is discouraged as they limit future analysis of stage subclassification or reclassification as new knowledge on ORNJ emerges. A separate quantitative measurement (clinical:exposed_bone_width_in_mm) can be considered to capture 2-dimensional data on extent of bony exposure as needed for the area classification in the modified Shaw system.^4^ Additional MDEs for this presentation include **clinical:mucosal_status** (absent)**, imaging:morphology, imaging:vertical_ab_abnormality** (beyond AB).

#### Intermediate Stage ORNJ

- *Exposed bone with any imaging findings <u>beyond</u> AB:* This feature combination was challenging to classify. When prompted with a text description in round 3, experts were split (44% intermediate, 40% advanced). A significant association between specialty group and classification was observed with most dental specialists considering it intermediate stage (69%) while others (72% RadOnc, 57% surgery) favored upstaging the feature combination (*P*=0.04). The repeated question in round 4, limited to these two stage options, resulted in convergence towards classification as intermediate stage (63%, 34/54, consensus threshold met). Case 9, a clinical report of exposed bone with OPG images showing full thickness sclerosis of the right mandible, and Case 10, a photograph of clinical bone exposure extending less than a molar width with axial CT image showing cortical bone changes, were presented as variations of this combination, with convergence towards classifying both also as *Intermediate ORNJ* during the final round (Case 9, 58%; Case 10, 86%). However, the upstaging effects of visualizing more extensive clinical bone exposure was reproducible using Case 13 which was staged as *Advanced ORNJ* by 71% (36/51) of experts over *Intermediate ORNJ* (29%). A more accurate summarization of such scenarios is through the use of standardized MDEs such as **clinical:exposed_bone_length_in_mm**, **clinical:mucosal_status**(absent)**, imaging:morphology,** and **imaging:vertical_ab_abnormality** (beyond AB).

#### Advanced Stage ORNJ

The Consortium exhibited high agreement in classifying pathologic fracture (96%, 49/51), orocutaneous fistula (92%, 47/51), and oro-antral or oro-nasal fistula (86%, 43/51) as advanced features of ORNJ. These results are in alignment with advanced features reported in the ClinRad model and other staging systems, and their presence on clinical examination should be explicitly reported in a standardized fashion such as a **disorder_present** MDE with unique identifiers for each disorder (Table 2).

### Specialty-Specific Knowledge Siloes & Inter-Rater Reliability

Within the ORAL Consortium, a significant difference was found in the utilization of OPGs with experts from Oral Medicine/Oncology and OMFS (i.e., Oral/Dental) using them twice as often as oncologists (84% v 43%, *P=0.008*; Table S1). CT scans were commonly used by dental and oncology groups (75% v 90%; *P*=0.166) whereas MRI was used more frequently by oncologists than oral specialists (38% v 21%; *P*=0.334). When questioned on the effectiveness of each modality for diagnosing ORNJ (Table S2), CT scans were rated the most effective by more than 86% of the Consortium while approximately one-third of both specialty-condensed groups were neutral on the effectiveness of OPGs for ORNJ surveillance. While the least adopted modality, MRI had comparable effectiveness ratings to OPG, with experts favoring the following sequences: T1-weighted with contrast (36%), T2-weighted (29%), and DCE (22%).

The potential impact of familiarity with various imaging modalities on level of confidence in diagnosing imaged-based cases is seen in Figure 4. Out of 9 scenarios presented in round 3 with a radiographic image included (excluding case 5-clinical MBS), dental specialists had the highest LOC for all OPG-based cases but exhibited lowered LOC interquartile ranges comparable to RadOnc and surgery for Case 7 (MRI and CT) and Case 10 (CT). Cases with limited OPG-based abnormalities within the alveolar bone (i.e., periodontal ligament space widening; Cases 3, 4, 6, 8) evoked lower LOC among oncologists in diagnostic capabilities whereas radical changes seen on OPG such as a pathologic fracture (Case 2) were consistently classified the same for stage and extent of bone involvement by all specialists with a reported high LOC.

During round 3, the inter-rater reliability (IRR) for staging image-based cases (Table S3) varied per specialty with slight agreement among RadOnc (IRR 0.13, n=22) and fair or better agreement between surgeons (IRR 0.29, n=6) and dental specialists (IRR 0.34, n=14). Based on likelihood of categorizing the same cases per specific category, all experts showed the highest specialty group-level agreement for categorizing cases of *Advanced ORNJ* (RadOnc IRR 0.43; Surgery IRR 0.57; Oral/Dental IRR 0.67) whereas the least agreement, if any, was around what cases should be considered unrelated to ORNJ (RadOnc IRR 0.1; Surgery IRR -0.07; Oral/Dental IRR 0.15), a precursor stage (Oral/Dental IRR 0.27), or an intermediate stage (RadOnc IRR 0.17; Surgery IRR 0.17). After providing group feedback from round 3, an introduction to the ClinRad risk-based model, and educational imaging resources for how to interpret OPG images, the overall specialty-level IRR during the last round improved for all groups (Oral/Dental IRR 0.38, n=18; RadOnc IRR 0.39, n=22; Surgery IRR 0.58, n=5). Moderate to substantial agreement was achieved for classifying cases as *Precursor/ Stage 0, Early ORNJ,* and *Intermediate ORNJ* but at a cost of decreasing agreement on non-fistula and non-pathologic fracture cases of *Advanced ORNJ*.

## S5_Supplemental Tables

**Table S1.** Use of Imaging Modalities per Condensed Specialties.

|  | Oncology<br>(N=40) | Oral/OMFS<br>(N=19) | P-value |
| --- | --- | --- | --- |
| Use of CT for ORNJ eval in past 1 year |  |  |  |
| Yes | 30 (75.0%) | 17 (89.5%) | 0.166 |
| No | 10 (25.0%) | 1 (5.3%) |  |
| Missing | 0 (0%) | 1 (5.3%) |  |
| Use of MRI for ORNJ eval in past 1 year |  |  |  |
| Yes | 15 (37.5%) | 4 (21.1%) | 0.334 |
| No | 25 (62.5%) | 15 (78.9%) |  |
| Use of OPG for ORNJ eval in past 1 year |  |  |  |
| Yes | 17 (42.5%) | 16 (84.2%) | 0.008 |
| No | 22 (55.0%) | 3 (15.8%) |  |
| Missing | 1 (2.5%) | 0 (0%) |  |

CT, Computed Tomography; MRI, Magnetic Resonance Imaging; OMFS, Oral and Maxillofacial Surgery; OPG, Orthopantomogram; ORNJ, Osteoradionecrosis of the Jaw

**Table S2.** Rated Effectiveness of Each Imaging Modality for ORNJ Surveillance.

|  | Oncology<br>(N=40) | Oral/OMFS<br>(N=19) | Overall<br>(N=59) |
| --- | --- | --- | --- |
| **Effectiveness of CT for ORNJ Surveillance** |  |  |  |
| Very/somewhat effective | 35 (87.5%) | 16 (84.2%) | 51 (86.4%) |
| Neutral | 2 (5.0%) | 2 (10.5%) | 4 (6.8%) |
| Somewhat ineffective | 1 (2.5%) | 0 (0%) | 1 (1.7%) |
| Missing | 2 (5.0%) | 1 (5.3%) | 3 (5.1%) |
| **Effectiveness of MRI for ORNJ Surveillance** |  |  |  |
| Very/somewhat effective | 25 (62.5%) | 8 (42.1%) | 33 (55.9%) |
| Neutral | 9 (22.5%) | 7 (36.8%) | 16 (27.1%) |
| Somewhat/very ineffective | 3 (7.0%) | 2 (10.5%) | 5 (8.5%) |
| Missing | 3 (7.5%) | 2 (10.5%) | 5 (8.5%) |
| **Effectiveness of OPG for ORNJ Surveillance** |  |  |  |
| Very/somewhat effective | 21 (62.5%) | 10 (52.7%) | 31 (52.6%) |
| Neutral | 13 (32.5%) | 6 (31.6%) | 19 (32.2%) |
| Somewhat/very ineffective | 3 (7.5%) | 3 (15.8%) | 6 (10.2%) |
| Missing | 3 (7.5%) | 0 (0%) | 3 (5.1%) |

CT, Computed Tomography; MRI, Magnetic Resonance Imaging; OMFS, Oral and Maxillofacial Surgery; OPG, Orthopantomogram; ORNJ, Osteoradionecrosis of the Jaw

**Table S3.** Inter-Rater Reliability for Staging Image-Based Cases.

|  | Round 3 | Round 4* |
| --- | --- | --- |
| **ALL CASES** |  |  |
| Oral/Dental | 0.34 | 0.38 |
| Rad Onc | 0.13 | 0.39 |
| Surgery | 0.29 | **0.58** |
| **PER CATEGORY** |  |  |
| *Not Precursor/ Not ORNJ* |  |  |
| Oral/Dental | 0.15 | 0.2 |
| Rad Onc | 0.01 | 0.22 |
| Surgery | -0.07 | -0.02 |
| *Precursor/ Stage 0* |  |  |
| Oral/Dental | 0.27 | 0.36 |
| Rad Onc | 0.1 | **0.44** |
| Surgery | 0.27 | **0.62** |
| *Early ORNJ* |  |  |
| Oral/Dental | 0.28 | 0.39 |
| Rad Onc | 0.06 | **0.44** |
| Surgery | 0.31 | **0.56** |
| *Intermediate ORNJ* |  |  |
| Oral/Dental | 0.32 | **0.46** |
| Rad Onc | 0.07 | **0.42** |
| Surgery | 0.17 | **0.65** |
| *Advanced ORNJ* |  |  |
| Oral/Dental | **0.67** | 0.38 |
| Rad Onc | **0.43** | 0.4 |
| Surgery | **0.57** | NA** |

\*Round 4 excludes Case 2 – pathologic fracture where there was high agreement during Round 3.
\*\*No surgeon selected *Advanced ORN* as a stage category for all cases.
All values in **bold** reflect moderate or higher agreement (IRR ≥ 0.41).
Abbreviations; ORNJ, Osteoradionecrosis of the Jaw; Rad Onc, Radiation Oncology

## REFERENCES

1. Watson, E. E. et al. Development and Standardization of a Classification System for Osteoradionecrosis: Implementation of a Risk-Based Model. medRxiv 2023.09.12.23295454 (2023) doi:10.1101/2023.09.12.23295454.

2. Wilde, D. C. et al. Early detection of mandible osteoradionecrosis risk in a high comorbidity veteran population. Am J Otolaryngol 44, (2023).

3. Treister, N. S. et al. Exposed bone in patients with head and neck cancer treated with radiation therapy: An analysis of the Observational Study of Dental Outcomes in Head and Neck Cancer Patients (OraRad). Cancer 128, 487–496 (2022).

4. Mohamed, A. S. R. et al. Dose-volume correlates of mandibular osteoradionecrosis in oropharynx cancer patients receiving intensity-modulated radiotherapy: Results from a case-matched comparison. Radiother Oncol 124, 232 (2017).

5. Dijk, L. V. van et al. Normal Tissue Complication Probability (NTCP) prediction model for osteoradionecrosis of the mandible in head and neck cancer patients following radiotherapy: Large-scale observational cohort. medRxiv 2021.03.04.21252505 (2021) doi:10.1101/2021.03.04.21252505.

6. Caparrotti, F. et al. Osteoradionecrosis of the mandible in patients with oropharyngeal carcinoma treated with intensity-modulated radiotherapy. Cancer 123, 3691–3700 (2017).

7. Wong, A. T. T. et al. Symptom burden and dysphagia associated with osteoradionecrosis in long-term oropharynx cancer survivors: A cohort analysis. Oral Oncol 66, 75–80 (2017).

8. De Melo, N. B. et al. Oral health related quality of life and determinant factors in patients with head and neck cancer. Med Oral Patol Oral Cir Bucal 24, e281 (2019).

9. Peterson, D. et al. Prevention and Management of Osteoradionecrosis in Patients with Head and Neck Cancer Treated with Radiation Therapy: ISOO-MASCC-ASCO Guideline. Journal of Clinical Oncology (2024).

10. Patel, V., Ormondroyd, L., Lyons, A. & McGurk, M. The financial burden for the surgical management of osteoradionecrosis. Br Dent J 222, 177–180 (2017).

11. Elting, L. S. & Chang, Y. C. Costs of Oral Complications of Cancer Therapies: Estimates and a Blueprint for Future Study. JNCI Monographs 2019, (2019).

12. ICD-11 for Mortality and Morbidity Statistics. https://icd.who.int/browse/2024-01/mms/en.

13. Coffin, F. The incidence and management of osteoradionecrosis of the jaws following head and neck radiotherapy. Br J Radiol 56, 851–857 (1983).

14. Marx, R. E. A new concept in the treatment of osteoradionecrosis. Journal of Oral and Maxillofacial Surgery 41, 351–357 (1983).

15. Epstein, J. B., Wong, F. L. W. & Stevenson-Moore, P. Osteoradionecrosis: Clinical experience and a proposal for classification. Journal of Oral and Maxillofacial Surgery 45, 104–110 (1987).

16. Glanzmann, C. & Grätz, K. W. Radionecrosis of the mandibula: a retrospective analysis of the incidence and risk factors. Radiotherapy and Oncology 36, 94–100 (1995).

17. Clayman, L. Management of dental extractions in irradiated jaws: A protocol without hyperbaric oxygen therapy. Journal of Oral and Maxillofacial Surgery 55, 275–281 (1997).

18. Støre, G. & Boysen, M. Mandibular osteoradionecrosis: Clinical behaviour and diagnostic aspects. Clin Otolaryngol Allied Sci 25, 378–384 (2000).

19. Schwartz, H. C. & Kagan, A. R. Osteoradionecrosis of the mandible: Scientific basis for clinical staging. American Journal of Clinical Oncology: Cancer Clinical Trials 25, 168–171 (2002).

20. Notani, K. ichi et al. Management of mandibular osteoradionecrosis corresponding to the severity of osteoradionecrosis and the method of radiotherapy. Head Neck 25, 181–186 (2003).

21. Shaw, R., Tesfaye, B., Bickerstaff, M., Silcocks, P. & Butterworth, C. Refining the definition of mandibular osteoradionecrosis in clinical trials: The cancer research UK HOPON trial (Hyperbaric Oxygen for the Prevention of Osteoradionecrosis). Oral Oncol 64, 73–77 (2017).

22. Tsai, C. J. et al. Osteoradionecrosis and Radiation Dose to the Mandible in Patients With Oropharyngeal Cancer. International Journal of Radiation Oncology*Biology*Physics 85, 415–420 (2013).

23. Karagozoglu, K. H. et al. Proposal for a new staging system for osteoradionecrosis of the mandible. Med Oral Patol Oral Cir Bucal 19, e433–e437 (2014).

24. Lyons, A., Osher, J., Warner, E., Kumar, R. & Brennan, P. A. Osteoradionecrosis—A review of current concepts in defining the extent of the disease and a new classification proposal. British Journal of Oral and Maxillofacial Surgery 52, 392–395 (2014).

25. He, Y. et al. Retrospective analysis of osteoradionecrosis of the mandible: proposing a novel clinical classification and staging system. Int J Oral Maxillofac Surg 44, 1547–1557 (2015).

26. CTCAE Files. https://evs.nci.nih.gov/ftp1/CTCAE/About.html.

27. Morton, M. E. & Simpson, W. The management of osteoradionecrosis of the jaws. Br J Oral Maxillofac Surg 24, 332–341 (1986).

28. Delphi Method | RAND. https://www.rand.org/topics/delphi-method.html.

29. Moossdorff, M. et al. Maastricht Delphi consensus on event definitions for classification of recurrence in breast cancer research. J Natl Cancer Inst 106, (2014).

30. Chollette, V., Weaver, S. J., Huang, G., Tsakraklides, S. & Tu, S.-P. Identifying Cancer Care Team Competencies to Improve Care Coordination in Multiteam Systems: A Modified Delphi Study. JCO Oncol Pract 16, e1324–e1331 (2020).

31. Gattrell, W. T., et al. ACCORD (ACcurate COnsensus Reporting Document): A reporting guideline for consensus methods in biomedicine developed via a modified Delphi. PLoS Med 21, e1004326 (2024).

32. REDCap. https://www.project-redcap.org/.

33. Qualtrics XM: The Leading Experience Management Software. https://www.qualtrics.com/.

34. NCI Thesaurus. https://thesaurus.cancer.gov/ncitbrowser/pages/concept_details.jsf?dictionary=NCI_Thesaurus&version=21.02d&code=C63924&ns=ncit&type=all&key=null&b=1&n=0&vse=null.

35. Beumer, J., Curtis, T. & Harrison, R. E. Radiation therapy of the oral cavity: Sequelae and management, part 1. Head Neck Surg 1, 301–312 (1979).

36. Widmark, G., Sagne, S. & Heikel, P. Osteoradionecrosis of the jaws. Int J Oral Maxillofac Surg 18, 302–306 (1989).

37. Harris, M. The conservative management of osteoradionecrosis of the mandible with ultrasound therapy. Br J Oral Maxillofac Surg 30, 313–318 (1992).

38. LENT-SOMA Tables. https://www.ada.org/-/media/project/ada-organization/ada/ada-org/files/publications/cdt/ada_utds_value_set_v1_2022_aug.pdf.

39. Common toxicity Criteria (CTC). https://www.eortc.be/services/doc/ctc/.

40. Ruggiero, S. L. et al. American Association of Oral and Maxillofacial Surgeons’ Position Paper on Medication-Related Osteonecrosis of the Jaws-2022 Update. (2022) doi:10.1016/j.joms.2022.02.008.

41. Singer, S. et al. Validation of the EORTC QLQ-C30 and EORTC QLQ-H&N35 in patients with laryngeal cancer after surgery. Head Neck 31, 64–76 (2009).

42. Rosenthal, D. I. et al. Patterns of symptom burden during radiotherapy or concurrent chemoradiotherapy for head and neck cancer: A prospective analysis using the University of Texas MD Anderson Cancer Center Symptom Inventory-Head and Neck Module. Cancer 120, 1975–1984 (2014).

43. Rosenthal, D. I. et al. Measuring head and neck cancer symptom burden: The development and validation of the M. D. Anderson symptom inventory, head and neck module. Head Neck 29, 923–931 (2007).

44. Ho, J.-H. University of Washington Quality of Life Questionnaire. Encyclopedia of Quality of Life and Well-Being Research 6821–6824 (2014) doi:10.1007/978-94-007-0753-5_3115.

45. Mohamed, A. S. R. et al. Quantitative Dynamic Contrast-Enhanced MRI Identifies Radiation-Induced Vascular Damage in Patients With Advanced Osteoradionecrosis: Results of a Prospective Study. Int J Radiat Oncol Biol Phys 108, 1319–1328 (2020).

46. McHugh, M. L. Interrater reliability: the kappa statistic. Biochem Med (Zagreb) 22, 276 (2012).

47. Beiderbeck, D., Frevel, N., Gracht, H. A. von der, Schmidt, S. L. & Schweitzer, V. M. Preparing, conducting, and analyzing Delphi surveys: Cross-disciplinary practices, new directions, and advancements. MethodsX 8, 101401 (2021).

48. Shah, H. A. & Kalaian, S. A. Which Is the Best Parametric Statistical Method For Analyzing Delphi Data? Journal of Modern Applied Statistical Methods 8, 5–6 (2009).

49. Listgarten, M. A. Periodontal probing: what does it mean? J Clin Periodontol 7, 165–176 (1980).

50. Hefti, A. F. Periodontal probing. Crit Rev Oral Biol Med 8, 336–356 (1997).

## References

1. REDCap. https://www.project-redcap.org/.

2. Qualtrics XM: The Leading Experience Management Software. https://www.qualtrics.com/.

3. MSSO Updates | MedDRA. https://www.meddra.org/.

4. SNOMED - Home | SNOMED International. https://www.snomed.org/.

5. New ICD-10-CM & ICD-10-PCS Codes | NC Medicaid. https://medicaid.ncdhhs.gov/blog/2023/01/31/new-icd-10-cm-icd-10-pcs-codes.

6. CTCAE Files. https://evs.nci.nih.gov/ftp1/CTCAE/About.html.

7. Mohamed, A. S. R. et al. Quantitative Dynamic Contrast-Enhanced MRI Identifies Radiation-Induced Vascular Damage in Patients With Advanced Osteoradionecrosis: Results of a Prospective Study. Int J Radiat Oncol Biol Phys 108, 1319–1328 (2020).

8. Watson, E. E. et al. Development and Standardization of a Classification System for Osteoradionecrosis: Implementation of a Risk-Based Model. medRxiv 2023.09.12.23295454 (2023) doi:10.1101/2023.09.12.23295454.

10. Mallya, S. M. & Tetradis, S. Imaging of Radiation- and Medication-Related Osteonecrosis. Radiol Clin North Am 56, 77–89 (2018).

11. Chan, K. C. et al. Mandibular changes on panoramic imaging after head and neck radiotherapy. Oral Surg Oral Med Oral Pathol Oral Radiol 121, 666–672 (2016).

12. McHugh, M. L. Interrater reliability: the kappa statistic. Biochem Med (Zagreb) 22, 276 (2012).

13. Table 2, Interpretation of Fleiss’ kappa (κ) (from Landis and Koch 1977). (2012).

14. R: The R Project for Statistical Computing. https://www.r-project.org/.

15. Download Python | Python.org. https://www.python.org/downloads/.

## REFERENCES

1. Listgarten, M. A. Periodontal probing: what does it mean? J Clin Periodontol 7, 165–176 (1980).

2. Hefti, A. F. Periodontal probing. Crit Rev Oral Biol Med 8, 336–356 (1997).

3. Watson, E. E. et al. Development and Standardization of a Classification System for Osteoradionecrosis: Implementation of a Risk-Based Model. medRxiv 2023.09.12.23295454 (2023) doi:10.1101/2023.09.12.23295454.

4. Shaw, R., Tesfaye, B., Bickerstaff, M., Silcocks, P. & Butterworth, C. Refining the definition of mandibular osteoradionecrosis in clinical trials: The cancer research UK HOPON trial (Hyperbaric Oxygen for the Prevention of Osteoradionecrosis). Oral Oncol 64, 73–77 (2017).

